# Guidelines for preventing respiratory illness in older adults aged 60 years and above living in long-term care: A rapid review of clinical practice guidelines

**DOI:** 10.1101/2020.03.19.20039180

**Authors:** Patricia Rios, Amruta Radhakrishnan, Sonia M. Thomas, Nazia Darvesh, Sharon E. Straus, Andrea C. Tricco

**Affiliations:** Li Ka Shing Knowledge Institute, St. Michael’s Hospital, Unity Health Toronto; Li Ka Shing Knowledge Institute, St. Michael’s Hospital, Unity Health Toronto and Department of Medicine, University of Toronto; Li Ka Shing Knowledge Institute, St. Michael’s Hospital, Unity Health Toronto and Dalla Lana School of Public Health & Institute of Health Policy, Management, and Evaluation, University of Toronto

## Abstract

**Background:** The overall objective of this rapid review was to identify infection protection and control recommendations from published clinical practice guidelines (CPGs) for adults aged 60 years and older in long-term care settings

**Methods:** Comprehensive searches in MEDLINE, EMBASE, the Cochrane Library, and relevant CPG publishers/repositories were carried out in early March 2020. Title/abstract and full-text screening, data abstraction, and quality appraisal (AGREE-II) were carried out by single reviewers.

**Results:** A total of 17 relevant CPGs were identified, published in the USA (n=8), Canada (n=6), Australia (n=2), and the United Kingdom (n=1). All of the CPGs dealt with infection control in long-term care facilities (LTCF) and addressed various types of viral respiratory infections (e.g., influenza, COVID-19, severe acute respiratory syndrome). Ten or more CPGs recommended the following infection control measures in LTCF: hand hygiene (n=13), wearing personal protective equipment (n=13), social distancing or isolation (n=13), disinfecting surfaces (n=12), droplet precautions (n=12), surveillance and evaluation (n=11), and using diagnostic testing to confirm illness (n=10). While only two or more CPGs recommended these infection control measures: policies and procedures for visitors, staff and/or residents (n=9), respiratory hygiene/cough etiquette (n=9), providing supplies (n=9), staff and/or residents education (n=8), increasing communication (n=6), consulting or notifying health professionals (n=6), appropriate ventilation practices (n=2), and cohorting equipment (n=2). Ten CPGs also addressed management of viral respiratory infections in LTCF and recommended antiviral chemoprophylaxis (n=10) and one CPG recommended early mobilization of residents.

**Conclusion:** The recommendations from current guidelines overall seem to support environmental measures for infection prevention and antiviral chemoprophylaxis for infection management as the most appropriate first-line response to viral respiratory illness in long-term care.

## INTRODUCTION

### Purpose and Research Questions

The Infection Prevention & Control of the World Health Organization (WHO) Health Emergency Programme presented a query on preventing and managing COVID-19 in older adults aged 60 years and above living in long-term care facilities including privately paid for and publicly paid for settings with a 5-business day timeline. According to the WHO, “long-term care covers those activities undertaken by others to ensure that people with, or at risk of, a significant ongoing loss of intrinsic capacity can maintain a level of functional ability consistent with their basic rights, fundamental freedoms and human dignity” (https://www.who.int/ageing/long-term-care/WHO-LTC-series-subsaharan-africa.pdf?ua=1). Examples of long-term care include nursing homes, charitable homes, municipal homes, long-term care hospitals, long-term care facilities, skilled nursing facilities, convalescent homes, and assisted living facilities (https://www.canada.ca/en/health-canada/services/home-continuing-care/long-term-facilities-based-care.html).

The overall objective of this rapid review of clinical practice guidelines was to identify infection protection and control measures for adults aged 60 years and older in long-term care settings. In order to focus the research question to increase feasibility, we proposed the following key research questions:

1. What are the infection prevention and control practices/measures for preventing or reducing respiratory viruses (including coronavirus and influenza) in older adults aged 60 years and above living in long-term care?
2. How do infection prevention and control practices differ for adults aged 60 years and above living in long-term care with respiratory illness and severe comorbidities or frailty differ than those without such severe comorbidities/frailty?
3. How do infection prevention and control practices differ for adults aged 60 years and above living in long-term care with respiratory illness from low- and middle-income economy countries (LMIC) differ than those living in high-income economy countries and do differences exist across different cultural contexts?

## METHODS

The rapid review conduct was guided by the Cochrane Handbook for Systematic Reviews of Interventions^1^ along with the Rapid Review Guide for Health Policy and Systems Research^2^. The team used an integrated knowledge translation approach, with consultation from the knowledge users from the WHO at the following stages: question development, interpretation of results, and draft report. We used the Preferred Reporting Items for Systematic Reviews and Meta-Analyses (PRISMA) Statement^3^ to guide the reporting of our rapid review results; a PRISMA extension for rapid reviews is currently under development. This rapid review of guidelines was completed in conjunction with a rapid overview of systematic reviews published in a separate report titled: *Preventing respiratory illness in older adults aged 60 years and above living in long-term care: A rapid overview of reviews*”.

### Protocol

We prepared a brief protocol for this query that is available in Appendix 1. If publication in a peer-reviewed journal is planned in the future, we will register this rapid review with the Open Science Framework (https://osf.io/).

### Literature search

Comprehensive literature searches addressing all research questions were developed by an experienced librarian for MEDLINE, EMBASE, the Cochrane Library, as well as online guideline repositories (e.g., guidelines.ecri.org, joulecma.ca/cpg, www.guidelinecentral.com). The full MEDLINE search strategy can be found in Appendix 2 and the full list of CPG sites searched can be found in Appendix 3. Due to the rapid timelines for this review, a peer review of the literature search was not conducted.

### Eligibility criteria

The Eligibility criteria followed the PICOST framework and consisted of:

<u>Population:</u> Individuals aged 60 years and above residing in long-term care facilities. The age cut-off for an older adult might be 50 years and above in different LMIC and/or cultural settings. As such, we included these in level 1 screening of titles and abstracts and presented potentially relevant studies in an appendix.
<u>Intervention:</u> Any form of infection control and prevention, such as hand hygiene, respiratory hygiene/etiquette, personal protective equipment (for patients and health care providers), triage (on arrival), source control, isolation, daily monitoring/surveillance for signs and symptoms of respiratory illness (e.g., COVID-19) in residence, environmental cleaning, cleaning of linen and medical equipment used by patients, restrictions on resident movement and transportation, restrictions on visitors, restrictions on travel for health care providers and other long-term care facility staff, waste management, dead body management. Only those measures used to prevent and control respiratory illnesses, including influenza and coronavirus (e.g., COVID-19, MERS, SARS) were included. Interventions focused on preventing bacterial respiratory outbreaks (e.g., strep, pneumonia, klebsiella) or aspiration pneumonia were excluded. Interventions specifically focused on vaccination were excluded, as a vaccine for the coronavirus currently does not exist.
<u>Comparator:</u> One of the interventions listed above or no intervention
<u>Outcomes:</u> Lab-confirmed respiratory illness due to the virus (e.g., SARS, MERS, COVID-19, influenza) [primary outcome], secondary bacterial infection, symptoms, secondary transmission (e.g., other patients, healthcare workers, visitors), goal concordant care, hospitalization, intensive-care unit (ICU) admission, and mortality.
<u>Study designs:</u> We limited inclusion to clinical practice guidelines using the Institute of Medicines definition of a guideline (http://www.iom.edu/Reports/2011/Clinical-Practice-Guidelines-We-Can-Trust.aspx).
<u>Time periods:</u> All periods of time and duration of follow-up were eligible.
<u>Other:</u> No other restrictions were imposed.

### Study selection

For both level 1 (title/abstract) and level 2 (full-text) screening, a screening form was prepared based on the eligibility criteria and pilot-tested by the review team using 25 citations prior to level 1 screening and 5 full text articles prior to level 2 screening. Agreement between reviewers was sufficiently high (>75%) in both cases so no further pilot-testing was required. Full screening was completed by a single reviewer for both level 1 and level 2 using Synthesi.SR, the team’s proprietary online software (https://breakthroughkt.ca/login.php).

### Data items and data abstraction

Items for data abstraction included guideline scope, target audience, guideline development methods, recommendations, and level of evidence for reach recommendation. A standardized data abstraction form was developed. Prior to data abstraction, a calibration exercise was completed by 2 team members independently. Following successful completion of the calibration exercise, included guidelines were abstracted by single reviewers.

### Quality appraisal

Quality appraisal was carried out using the AGREE-2 tool (https://www.agreetrust.org/resource-centre/agree-reporting-checklist/) for clinical practice guidelines. The AGREE-2 tool includes 23 items that are categorized according to the following six domains: scope and purpose, stakeholder involvement, rigour of development, clarity of presentation, applicability, and editorial independence.

### Synthesis

Included guidelines were synthesized descriptively including summary and detailed tables of guideline characteristics and recommendations. Where available, information on relevant subgroups were highlighted.

## RESULTS

### Literature Search

The database search returned a total of 3,267 citations, while the grey literature searches returned 32 citations for level 1 screening. A total of 3,183 citations were excluded after level 1 screening. Of the 116 full-text articles screened at level 2, 17 clinical practice guidelines were included (Figure 1).

**Figure 1:**
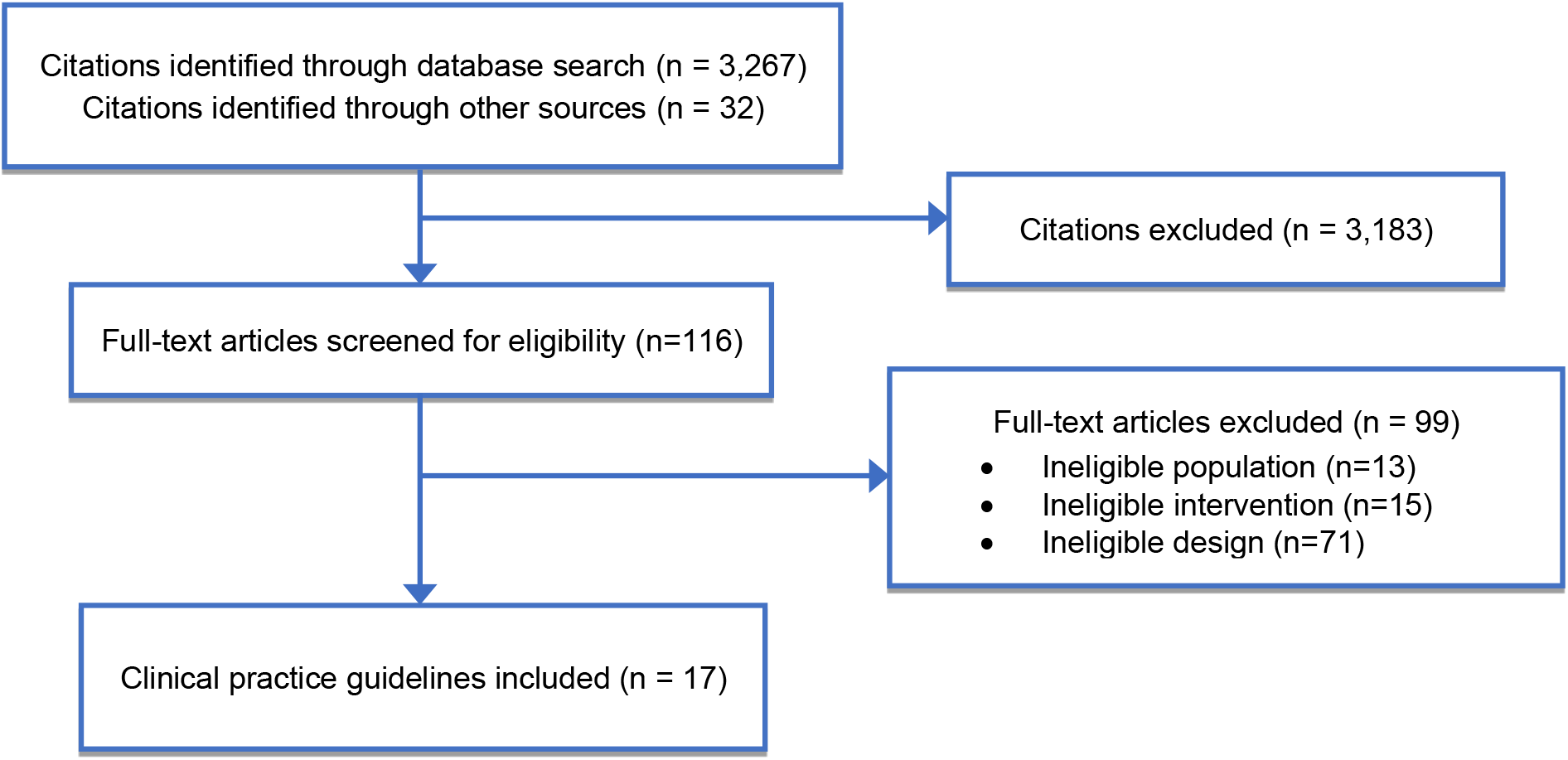
Study flow.

### Characteristics of included clinical practice guidelines

The 17 clinical practice guidelines were produced by organizations in the United States (n=8), Canada (n=6), Australia (n=2), and the United Kingdom (n=1). The topics covered by the guidelines included influenza (n=6), infection control in long-term care facilities in general (n=2), respiratory illness outbreaks (n=5), pneumonia (n=1), severe acute respiratory syndrome (n=1), and COVID-19 (n=2). Detailed characteristics of the included guidelines can be found in Appendix 4.

### Quality Appraisal Results

The 17 clinical practice guidelines were of very low quality (Table 1). For the scope and purpose domain, the clinical practice guidelines reported 3 to 10 of the relevant details out of 12 in total. For the stakeholder domain, the guidelines reported 0 to 6 of the relevant details out of 11 in total. For the rigour of development domain, the guidelines reported 0 to 14 of the relevant details out of 35 in total. For the clarity of presentation domain, the guidelines reported 1 to 8 of the relevant details out of 8 in total. For the applicability domain, the guidelines reported 0 to 3 of the relevant details out of 13 in total. For the editorial independence domain, the guidelines reported 0 to 4 of the relevant details out of 6 in total. The clinical practice guideline that fulfilled most of the criteria was Uyeki 2019 (33 of the 85 potential points), whereas the guidelines fulfilling the least of the criteria were the AMA 2008 and ECRI 2020 (2 of the 85 potential points). The full appraisal results for each included clinical practice guideline can be found in Appendix 5.

**Table 1:** Summary AGREE-II scores for the clinical practice guidelines.

|  | Tablan 2004 | AMA, 2008 | Smith, 2008 | PHAC, 2010 | Woodhead, 2011 | Rutimann, 2013 | Aoki, 2015 | Victoria State, 2015 | Burdsall, 2017 | Buynder, 2017 | MOHLTC, 2018 | CDPH, 2019 | Manitoba Health, 2019 | Uyeki, 2019 | CDC, 2020 | ECRI, 2020 | MOH, 2020 |
| --- | --- | --- | --- | --- | --- | --- | --- | --- | --- | --- | --- | --- | --- | --- | --- | --- | --- |
| **DOMAIN 1: Scope and Purpose (12 points in total)** |  |  |  |  |  |  |  |  |  |  |  |  |  |  |  |  |  |
| Item 1 | 2 | 1 | 3 | 2 | 2 | 2 | 1 | 2 | 2 | 2 | 2 | 2 | 1 | 3 | 1 | 1 | 2 |
| Item 2 | 1 | 1 | 2 | 1 | 4 | 1 | 1 | 1 | 1 | 1 | 2 | 1 | 2 | 4 | 1 | 1 | 1 |
| Item 3 | 3 | 2 | 2 | 2 | 2 | 2 | 1 | 1 | 2 | 1 | 2 | 1 | 1 | 3 | 1 | 1 | 1 |
| Totals | 6 | 4 | 7 | 5 | 8 | 5 | 3 | 4 | 5 | 4 | 6 | 4 | 4 | 10 | 3 | 3 | 4 |
| **DOMAIN 2: Stakeholder Involvement (11 points in total)** |  |  |  |  |  |  |  |  |  |  |  |  |  |  |  |  |  |
| Item 4 | 2 | 0 | 4 | 0 | 4 | 2 | 2 | 0 | 1 | 3 | 0 | 0 | 0 | 4 | 0 | 0 | 0 |
| Item 5 | 0 | 0 | 0 | 0 | 0 | 0 | 0 | 0 | 0 | 0 | 0 | 0 | 0 | 0 | 0 | 0 | 0 |
| Item 6 | 1 | 0 | 0 | 1 | 2 | 0 | 2 | 1 | 1 | 2 | 1 | 1 | 2 | 2 | 1 | 0 | 1 |
| Totals | 3 | 0 | 4 | 1 | 6 | 2 | 4 | 1 | 2 | 5 | 1 | 1 | 2 | 6 | 1 | 0 | 1 |
| **DOMAIN 3: Rigour of Development (35 points in total)** |  |  |  |  |  |  |  |  |  |  |  |  |  |  |  |  |  |
| Item 7 | 0 | 0 | 0 | 0 | 2 | 1 | 0 | 0 | 0 | 0 | 0 | 0 | 0 | 3 | 0 | 0 | 0 |
| Item 8 | 0 | 0 | 0 | 0 | 0 | 0 | 0 | 0 | 0 | 1 | 0 | 0 | 0 | 1 | 0 | 0 | 0 |
| Item 9 | 0 | 0 | 0 | 0 | 4 | 1 | 0 | 0 | 0 | 0 | 0 | 0 | 0 | 0 | 0 | 0 | 0 |
| Item 10 | 0 | 0 | 0 | 0 | 0 | 1 | 0 | 0 | 0 | 0 | 0 | 0 | 0 | 3 | 0 | 0 | 0 |
| Item 11 | 0 | 0 | 1 | 0 | 1 | 0 | 0 | 0 | 0 | 0 | 0 | 0 | 0 | 1 | 0 | 0 | 0 |
| Item 12 | 1 | 0 | 1 | 0 | 2 | 1 | 0 | 1 | 0 | 0 | 0 | 0 | 0 | 2 | 0 | 0 | 0 |
| Item 13 | 0 | 0 | 0 | 0 | 0 | 0 | 0 | 0 | 0 | 0 | 0 | 0 | 0 | 2 | 0 | 0 | 0 |
| Item 14 | 0 | 0 | 0 | 0 | 0 | 0 | 0 | 0 | 0 | 0 | 0 | 0 | 0 | 2 | 0 | 0 | 0 |
| Totals | 1 | 0 | 2 | 0 | 9 | 4 | 0 | 1 | 0 | 1 | 0 | 0 | 0 | 14 | 0 | 0 | 0 |
| **DOMAIN 4: Clarity of Presentation (8 points in total)** |  |  |  |  |  |  |  |  |  |  |  |  |  |  |  |  |  |
| Item 15 | 0 | 0 | 1 | 1 | 3 | 0 | 0 | 0 | 1 | 2 | 1 | 1 | 2 | 4 | 0 | 0 | 1 |
| Item 16 | 0 | 0 | 1 | 1 | 2 | 0 | 0 | 0 | 1 | 1 | 2 | 0 | 1 | 2 | 0 | 0 | 1 |
| Item 17 | 1 | 1 | 1 | 1 | 1 | 1 | 1 | 0 | 1 | 1 | 0 | 1 | 1 | 2 | 1 | 1 | 1 |
| Totals | 1 | 1 | 3 | 3 | 6 | 1 | 1 | 0 | 3 | 4 | 3 | 2 | 4 | 8 | 1 | 1 | 3 |
| **DOMAIN 5: Applicability (13 points in total)** |  |  |  |  |  |  |  |  |  |  |  |  |  |  |  |  |  |
| Item 18 | 0 | 0 | 0 | 0 | 0 | 0 | 0 | 0 | 0 | 0 | 0 | 0 | 0 | 0 | 0 | 0 | 0 |
| Item 19 | 0 | 0 | 1 | 1 | 0 | 0 | 0 | 1 | 1 | 1 | 1 | 1 | 1 | 1 | 0 | 0 | 1 |
| Item 20 | 0 | 0 | 0 | 0 | 0 | 0 | 0 | 0 | 0 | 0 | 0 | 0 | 0 | 0 | 0 | 0 | 0 |
| Item 21 | 0 | 0 | 0 | 0 | 0 | 0 | 0 | 0 | 0 | 2 | 0 | 0 | 0 | 0 | 0 | 0 | 0 |
| Totals | 0 | 0 | 1 | 1 | 0 | 0 | 0 | 1 | 1 | 3 | 1 | 1 | 1 | 1 | 0 | 0 | 1 |
| **DOMAIN 6: Editorial Independence (6 points in total)** |  |  |  |  |  |  |  |  |  |  |  |  |  |  |  |  |  |
| Item 22 | 1 | 1 | 0 | 1 | 1 | 1 | 1 | 1 | 1 | 0 | 1 | 0 | 0 | 1 | 1 | 1 | 1 |
| **Item 23** | 0 | 0 | 0 | 0 | 0 | 1 | 1 | 0 | 0 | 0 | 0 | 0 | 0 | 3 | 0 | 0 | 0 |
| **Totals** | 1 | 1 | 0 | 1 | 1 | 2 | 2 | 1 | 1 | 0 | 1 | 0 | 0 | 4 | 1 | 1 | 1 |

### Guideline Recommendations

*Preventing respiratory illness in long-term care facilities:* Two or more clinical practice guidelines recommended the following: hand hygiene (n=13), wearing personal protective equipment (n=13), social distancing/isolation (n=13), disinfecting surfaces (n=12), droplet precautions (n=12), surveillance and evaluation (n=11), conducting diagnostic testing to confirm suspected respiratory illness (n=10), policies and procedures for visitors (n=9), respiratory hygiene/cough etiquette (n=9), policies and procedures for staff and/or residents (n=9), providing supplies (n=9), education of staff and/or residents (n=8), increasing communication (n=6), consulting or notifying health professionals (n=6), appropriate ventilation practices (n=2), and cohorting equipment (n=2) (Table 2, Appendix 6). One clinical practice guideline recommended appropriate air ventilation or smoking cessation.

*Managing respiratory illness in long-term care facilities:* Ten clinical practice guidelines recommended the use of antivirals for prophylaxis of staff and/or residents and one recommended early mobilization of residents (Table 2, Appendix 6).

**Table 2:** Summary of recommendations from included clinical practice guidelines.

|  | Tablan, 2004 | AMA 2008 | Smith, 2008 | PHAC, 2010 | Woodhead, 2011 | Ruttimann, 2013 | Aoki, 2015 | Victoria State, 2015 | Burdsall, 2017 | Buynder, 2017 | CDPH, 2018 | MOHLTC, 2018 | Manitoba Health, 2019 | CDC, 2020 | ECRI, 2020 | Uyeki, 2019 | MOH, 2020 |
| --- | --- | --- | --- | --- | --- | --- | --- | --- | --- | --- | --- | --- | --- | --- | --- | --- | --- |
| **PREVENTION** |  |  |  |  |  |  |  |  |  |  |  |  |  |  |  |  |  |
| Appropriate ventilation |  |  | X |  |  |  |  |  | X |  |  |  |  |  |  |  |  |
| Cohorting equipment |  |  |  |  |  |  |  | X |  |  |  |  |  |  | X |  |  |
| Communication |  |  |  |  |  |  | X | X | X | X | X | X |  |  |  |  |  |
| Consulting/notifying health professionals |  |  |  |  |  |  |  | X |  |  | X | X |  | X | X |  | X |
| Diagnostic testing | X | X |  | X |  | X | X |  | X | X | X | X |  |  |  |  | X |
| Disinfecting surfaces | X |  | X | X |  | X |  | X | X | X | X |  | X | X | X |  | X |
| Droplet precautions | X |  | X | X |  | X |  | X | X | X | X | X | X | X |  |  | X |
| Education | X |  | X | X |  |  |  |  | X | X | X |  |  | X |  |  | X |
| Hand hygiene |  | X | X | X |  | X |  | X | X | X | X | X | X | X | X |  | X |
| Personal protective equipment | X |  | X | X |  |  | X | X | X | X | X | X | X | X | X |  | X |
| Policies for visitors | X |  |  | X |  |  |  | X | X | X | X | X | X | X |  |  | X |
| Policies for staff/residents |  | X | X | X |  |  | X |  | X | X | X | X |  |  |  |  | X |
| Provide supplies |  |  | X | X |  |  |  | X | X | X | X | X |  | X |  |  | X |
| Respiratory hygiene/ cough etiquette |  |  |  | X |  | X | X | X | X | X | X | X |  | X |  |  | X |
| Smoking cessation |  | X |  |  |  |  |  |  |  |  |  |  |  |  |  |  |  |
| Social distancing/ isolation/cohorting | X |  | X | X |  | X | X | X | X | X | X | X | X | X | X |  |  |
| Surveillance/monitoring/ evaluation | X |  | X | X |  |  | X | X |  | X | X | X |  | X |  | X | X |
| **MANAGEMENT** |  |  |  |  |  |  |  |  |  |  |  |  |  |  |  |  |  |
| Antivirals for prophylaxis for staff/residents | X |  |  | X | X | X | X | X |  | X | X | X |  |  |  | X |  |
| Early mobilization |  |  |  |  | X |  |  |  |  |  |  |  |  |  |  |  |  |

## DISCUSSION

The WHO commissioned a rapid review to address the urgent question of the infection prevention and control practices/measures for respiratory viruses in long-term care facilities that could be applied to COVID-19. A comprehensive literature search of both electronic databases and grey literature sources resulted 17 included clinical practice guidelines. None of the clinical practice guidelines specifically focused on issues related to residents with respiratory illness and severe comorbidities or frailty. Furthermore, none focused on issues in LMIC or different cultural contexts.

The most commonly recommended prevention strategies across the clinical practice guidelines were hand hygiene, wearing personal protective equipment, social distancing/isolation, disinfecting surfaces, droplet precautions, surveillance and evaluation, conducting diagnostic testing to confirm suspected respiratory illness, policies and procedures for visitors, policies and procedures for staff, and respiratory hygiene/cough etiquette. For managing respiratory illness in long-term care facilities, the majority of the clinical practice guidelines recommended antivirals for prophylaxis of staff and/or residents. However, most of the clinical practice guidelines failed to address multiple AGREE-II items, suggesting that they are most likely based on expert opinion.

There are several limitations to the review methods employed here, single screening and abstraction for example, however they were selected to thoughtfully tailor our methods according to our knowledge user needs and the urgent nature of the request to provide timely results. There is also a chance that our literature search missed guidance documents from various state and provincial authorities. However, we were unable to perform an exhaustive grey literature search of websites, due to the timelines imposed on this review.

## CONCLUSIONS

The recommendations from current guidelines overall seem to support environmental measures for infection prevention and antiviral chemoprophylaxis for infection management as the most appropriate first-line response to viral respiratory illness in long-term care. However, these recommendations should be viewed with caution as it is unclear how many of these guidelines are based on the best available evidence due to their poor overall quality.

## Data Availability

All datasets supporting the conclusions of this article are included within the article

## Acknowledgements

Jessie McGowan (literature search), Krystle Amog (report preparation), Chantal Williams (report preparation), Naveeta Ramkissoon (report preparation)

## Copyright claims/Disclaimers

The intellectual property rights in data and results generated from the work reported in this document are held in joint ownership between the Knowledge Translation Program team and the named Contributors.

Users are permitted to disseminate the data and results presented in this report provided that the dissemination (i) does not misrepresent the data, results, analyses or conclusions, and (ii) is consistent with academic practice, the rights of any third party publisher, and applicable laws. Any dissemination of the data and results from this document shall properly acknowledge the Knowledge Translation Program team and named Contributors.

## Funding Statement

This systematic review was commissioned and paid for by the World Health Organization and conducted through the Strategy for Patient Oriented-Research (SPOR) Evidence Alliance. The authors alone are responsible for the views expressed in this article and they do not necessarily represent the decisions, policy or views of the World Health Organization.

For questions about this report, please contact:

## APPENDIX 1 – PROTOCOL

**Team name:** Knowledge Translation Program of St. Michael’s Hospital, Unity Health Toronto (Drs. Tricco and Straus, contact)

**Query:** Preventing the transmission of coronavirus (COVID-19) in older adults aged 60 years and above living in long-term care

**Query Submitter:** Infection Prevention & Control, World Health Organization (WHO) Health Emergency (WHE) Programme

### Objective and research questions

The Infection Prevention & Control of the WHO WHE Programme has presented a query on the transmission of COVID-19 in older adults aged 60 years and above living in long-term care including privately paid for and publicly paid for settings with a 5-business day timeline. According to the World Health Organization, “long-term care covers those activities undertaken by others to ensure that people with, or at risk of, a significant ongoing loss of intrinsic capacity can maintain a level of functional ability consistent with their basic rights, fundamental freedoms and human dignity” (https://www.who.int/ageing/long-term-care/WHO-LTC-series-subsaharan-africa.pdf?ua=1) Examples of long-term care will include nursing homes, charitable homes, municipal homes, long-term care hospitals, long-term care facilities, skilled nursing facilities, convalescent homes, and assisted living facilities (https://www.canada.ca/en/health-canada/services/home-continuing-care/long-term-facilities-based-care.html).

The proposed research questions are:

### Research approach

The research question will be addressed through a rapid review informed by the methods proposed by the WHO Guide to rapid reviews (https://www.who.int/alliance-hpsr/resources/publications/rapid-review-guide/en/).

#### Protocol

Due to the urgent nature of the request and limited time frame to complete the work, this document will serve as the protocol for this query. If publication in a peer-reviewed journal is planned in the future, we will register this rapid review with the Open Science Framework (https://osf.io/).

#### Literature search

Comprehensive literature searches will be developed by an experienced librarian for MEDLINE, EMBASE, the Cochrane Library, biorxiv.org/medrxiv.org, and GIDEON (Global Infectious Diseases and Epidemiology Network). Grey (i.e., difficult to locate or unpublished) literature will be searched via clinicaltrials.gov. Due to the rapid timelines for this review a peer review of the literature search will not be conducted.

#### Eligibility criteria

The Eligibility criteria will follow the PICOST framework and will consist of:

<u>Population:</u> Individuals aged 60 years and above residing in long-term care facilities. The age cut-off for an older adult might be 50 years and above in some LMIC and/or cultural settings. As such, we will include these in level 1 screening of titles and abstracts and include anything deemed relevant at level 2 screening of full-text articles in an appendix.
<u>Interventions:</u> Any form of infection control and prevention, such as hand hygiene, respiratory hygiene/etiquette, personal protective equipment (for patients and health care providers), triage (on arrival), source control, isolation, daily monitoring/surveillance for signs and symptoms of respiratory illness (e.g., COVID-19) in residence, environmental cleaning, cleaning of linen and medical equipment used by patients, restrictions on resident movement and transportation, restrictions on visitors, restrictions on travel for health care providers and other long-term care facility staff, waste management, dead body management. Only those measures used to prevent and control respiratory illnesses, including influenza and coronavirus (e.g., COVID-19, MERS, SARS) will be included. Interventions focused on preventing bacterial respiratory outbreaks (e.g., strep, pneumonia, klebsiella) will be excluded.
<u>Comparator:</u> One of the interventions listed above or no intervention
<u>Outcomes:</u> Lab-confirmed respiratory illness due to the virus (e.g., SARS, MERS, COVID-19, influenza) [primary outcome], secondary bacterial infection, symptoms, secondary transmission (e.g., other patients, healthcare workers, visitors), goal concordant care, hospitalization, intensive-care unit (ICU) admission, mortality
<u>Study designs:</u> Due to the rapid nature of this request, we will limit inclusion to clinical practice guidelines and systematic reviews, using the Cochrane definition of a systematic review. If there is scant evidence from these study designs, we will expand inclusion to include the following study designs:
  - Randomized controlled trials (RCTs)
  - NRCTs (e.g., such as quasi-RCTs, non-randomized trials, interrupted time series, controlled before after), o Observational studies (e.g., cohort, case control, cross-sectional)
  - Case studies, case reports, and case series, including outbreak reports
<u>Time periods:</u> All periods of time and duration of follow-up will be included.
<u>Other limitations:</u> No other limitations will be imposed. If possible, we will translate studies written in languages other than English (e.g., Mandarin, Cantonese) that are deemed relevant.

#### Study selection process

In order to meet the requested timeline of 5 working days a streamlined approach to study selection will be employed. A screening form based on the eligibility criteria will be prepared and a brief calibration exercise will be conducted prior to citation and full-text screening. Screening will be completed by single reviewers using Synthesi.SR, the team’s proprietary online software (https://breakthroughkt.ca/login.php).

#### Data items and data abstraction process

Items for data abstraction will include study characteristics (e.g., duration of follow-up, study design, country of conduct, multi-center vs. single site, long-term care setting characteristics, such as availability of medical support, characteristics of care staff, family/community engagement, accommodation characteristics, collective practices), patient characteristics (e.g., mean age, age range, co-morbidities), intervention details (e.g., type of intervention, duration and frequency of intervention, timing of intervention), comparator details (e.g., comparator intervention, duration and frequency of intervention, timing of intervention), and outcome results (e.g., lab-confirmed viral respiratory infection, symptoms, secondary transmission, hospitalization, ICU admission, mortality) at the longest duration of follow-up. For the clinical practice guidelines, we will abstract the recommendations and level of evidence for reach recommendation.

Prior to data abstraction, we will complete a calibration exercise of the form amongst all reviewers using a random sample of 2 included articles. Following calibration, included studies will be abstracted by single reviewers.

#### Risk of bias appraisal

Risk of bias appraisal will be carried out by single reviewers using the AMSTAR-2 tool (https://amstar.ca/Amstar-2.php) for systematic reviews and the AGREE-2 tool (https://www.agreetrust.org/resource-centre/agree-reporting-checklist/) for clinical practice guidelines.

#### Synthesis

The synthesis will involve providing a descriptive summary of included studies with summary tables and detailed tables of study results. Tables of study results will be organized according to interventions of interest and reported outcomes and where available, information on relevant subgroups will be presented separately.

### Preliminary knowledge translation plan

The summary of results will be sent to the WHO and other relevant policy-makers as a brief summary report (1-5 pages) and 1-page policy brief (see http://www.cihr-irsc.gc.ca/e/documents/dsen-abstract-en.pdf for an example). We will work with the WHO team to consider submitting this paper to an open-access, peer-reviewed journal for publication (e.g. British Medical Journal).

### Timeline (from the point of official approval)

Five business days (March 16, 2020)

### Updates provided to the WHO

Daily emails will be sent to the WHO

### Funding

Funding will be obtained from the WHO and the Canadian Institutes of Health Research Strategy for Patient Oriented Research Evidence Alliance (https://sporevidencealliance.ca/).

## APPENDIX 2 – MEDLINE SEARCH STRATEGY

1. respiratory tract infections/ or exp bronchitis/ or exp common cold/ or exp influenza, human/ or laryngitis/ or exp pharyngitis/ or exp pleurisy/ or exp pneumonia/ or exp rhinitis/ or exp rhinoscleroma/ or exp severe acute respiratory syndrome/ or exp sinusitis/ or exp supraglottitis/ or exp tracheitis/ or exp whooping cough/
2. coronaviridae infections/ or coronavirus infections/ or SARS Virus/
3. (coronavirus* or “corona virus*” or mers or “middle east respiratory syndrome*” or “Severe Acute Respiratory Syndrome*” or SARS or CoV or SARS-CoV or MERS-CoV or 2019-nCoV or COVID-19 or “2019 novel coronavirus disease” or “2019 ncov disease” or “2019 ncov infection” or “coronavirus disease 19” or “severe acute respiratory syndrome coronavirus 2” or “severe acute respiratory syndrome coronavirus 2” or “wuhan” or “sars cov 2”).tw,kf.
4. (flu or influenza or “respiratory tract infection*” or “respiratory infection*” or bronchitis or “common cold” or laryngitis or pharyngitis or pneumonia or rhinitis or rhinoscleroma or sinusitis or supraglottitis or tracheitis or “whooping cough”).tw,kf.
5. or/1-4
6. pc.fs.
7. exp Infection Control/ or secondary prevention/
8. exp hand hygiene/ or hygiene/
9. (prevent* or “respiratory hygiene” or “respiratory etiquette “ or “cough etiquette” or “Hand Hygiene” or “hand wash*” or “handwash*” or “patient isolation” or “quarant*” or “infection control” or “blood safety” or steril* or disinfect* or “contract tracing” or “disease notification” or fumigat* or “personal protective equipment” or triage or “source control” or isolation or “daily monitoring” or surveillance or “waste management” or cadaver or body or corpse or “face mask*” or facemask* or “social distanc*” or housekeeping). tw,kw.
10. (clean* adj3 (linen or equipment or environment)).tw,kf.
11. (restrict* adj3 (resident* or patient* or visit* or family or travel* or staff or provider* or employee*)).tw,kf.
12. personal protective equipment/
13. Housekeeping, Hospital/
14. Waste Management/
15. patient isolation/
16. triage/
17. Cadaver/
18. or/6-17
19. 5 and 18
20. Long-Term Care/ or exp Nursing Homes/ or Homes for the Aged/ or Assisted Living Facilities/
21. (“long-term care” or “long term care” or “senior* home*” or “senior* residence*” or “nursing home*” or “old age home*” or “old age residence*” or “home* for the aged”).tw,kf. (48416)
22. 20 or 21
23. 19 and 22

## APPENDIX 3 – CLINICAL PRACTICE GUIDELINE RESOURCES

### Canadian Guidelines

- Alberta Medical Association; Toward Optimized Practice (TOP)
  - http://www.topalbertadoctors.org/cpgs.php?sid=1
- British Columbia Ministry of Health
  - http://www.2.gov.bc.ca/gov/content/health/practitioner-professional-resources/bc-guidelines
- Canadian Medical Association (CMA); CMA Infobase: Clinical Practice Guidelines
  - https://www.cma.ca/En/Pages/clinical-practice-guidelines.aspx
- Canadian Standards Association (CSA). Occupational Health and Safety
  - http://shop.csa.ca/en/canada/products/occupational-health-and-safety/
- The College of Physicians and Surgeons of Ontario (CPSO)
  - http://www.cpso.on.ca/Policies-Publications/CPGs-Other-Guidelines
- Ontario Association of Medical Laboratories (OAML)
  - https://oaml.com/guidelines/
- Registered Nurses’ Association of Ontario (RNAO); Nursing Best Practice Guidelines
  - http://rnao.ca/bpg
- University of Ottawa; School of Rehabilitation Science: Evidence-based Practice
  - http://www.health.uottawa.ca/rehabguidelines/en/search.php
- Winnipeg Regional Health Authority (WRHA); Evidence Informed Practice Tools
  - http://www.wrha.mb.ca/professionals/ebpt/

### International Guidelines

- Academy of Medicine of Malaysia. Clinical Practice Guidelines
  - http://www.acadmed.org.my/index.cfm?&menuid=67
- Aetna, Inc. Clinical Policy Bulletins
  - http://www.aetna.com/healthcare-professionals/policies-guidelines/medical_clinical_policy_bulletins.html (medical)
  - http://www.aetna.com/healthcare-professionals/policies-guidelines/pcpb_menu.html (pharmaceutical)
- American Association for Clinical Chemistry (AACC). Practice Guidelines
  - https://www.aacc.org/science-and-research/practice-guidelines#
- Best Practice Advocacy Centre New Zealand (bpacNZ)
  - http://www.bpac.org.nz/Default.aspx
- Centers for Disease Control and Prevention (CDC); Public Health Genomics Knowledge Base
  - https://phgkb.cdc.gov/PHGKB/evidencerStartPage.action
- The Regulation and Quality Improvement Authority (RQIA). Guidelines
  - https://rqia.org.uk/what-we-do/rqia-clinical-audit-programme/guidelines/
- Haute Autorite de sante/ French National Authority for Health (HAS). Practice Guidelines
  - http://www.has-sante.fr/portail/jcms/c_6056/en/recherche-avancee?portlet=c_39085&search_antidot=&lang=en&typesf=guidelines
- Institute for Clinical Systems Improvement (ICSI). Guidelines
  - https://www.icsi.org/guidelinesc__more/
- ECRI Institute. ECRI Guidelines Trust (ECRI)
  - https://guidelines.ecri.org/
- National Health and Medical Research Council (NHMRC); Australia’s Clinical Practice Guidelines Portal
  - http://www.clinicalguidelines.gov.au/
- National Institute for Health and Care Excellence (NICE)
  - http://www.nice.org.uk/guidance
- Scottish Intercollegiate Guidelines Network (SIGN)
  - http://www.sign.ac.uk/our-guidelines.html

### Guideline Repositories

- CPG Infobase: Clinical Practice Guidelines; Joule, a CMA Company
  - https://joulecma.ca/cpg/homepage
- Clinical Practice Guidelines & Medical Summaries Clearinghouse & App
  - https://www.guidelinecentral.com/summaries/
- U of T Library Research guides: Medicine: Clinical Practice Guidelines (CPGs)
  - https://guides.library.utoronto.ca/c.php?g=250555&p=1671101
- Clinical practice guidelines - Trip Database
  - https://www.tripdatabase.com/search?criteria=clinical+practice+guidelines

## APPENDIX 4 – CLINICAL PRACTICE GUIDELINE CHARACTERISTICS

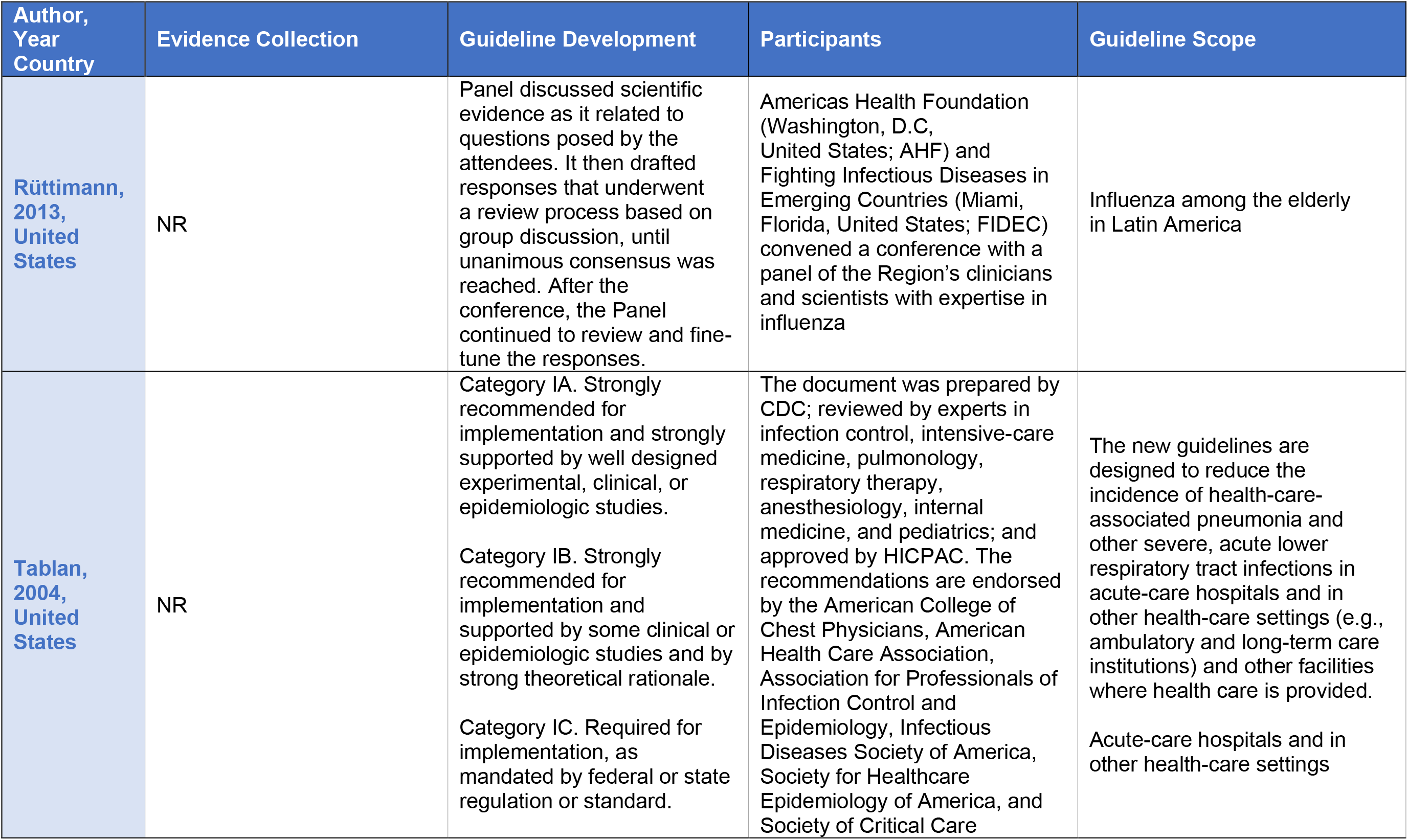

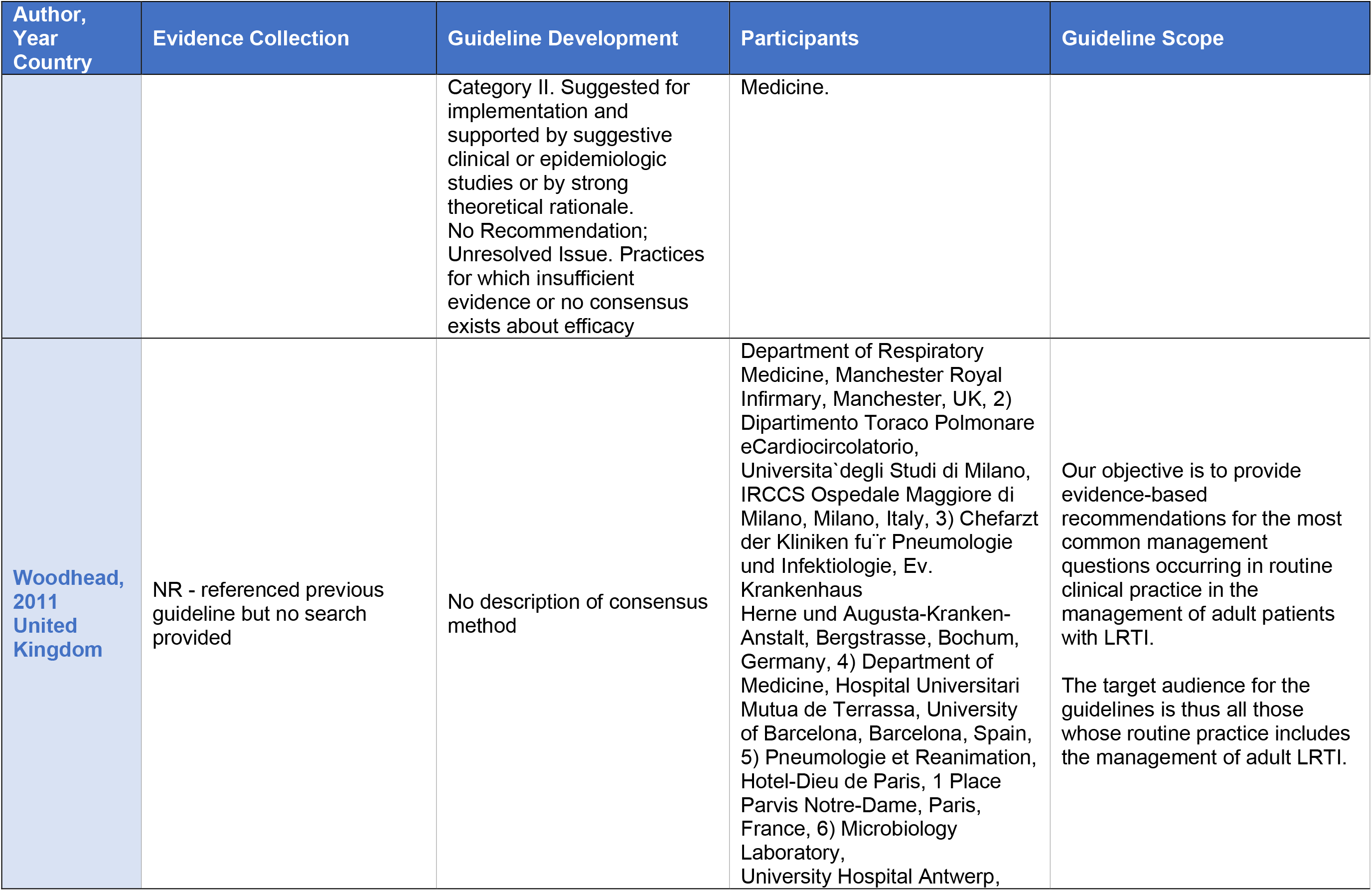

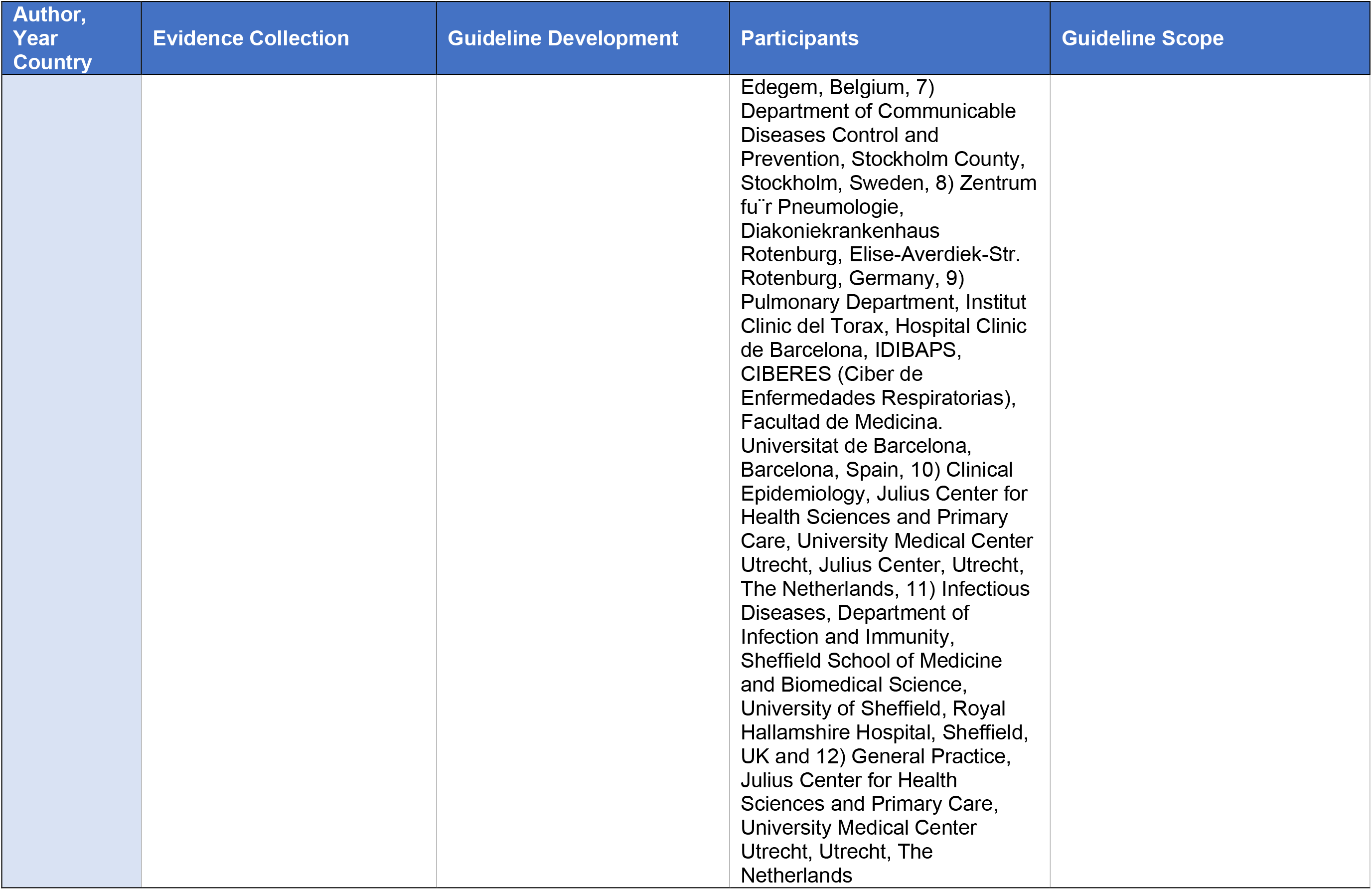

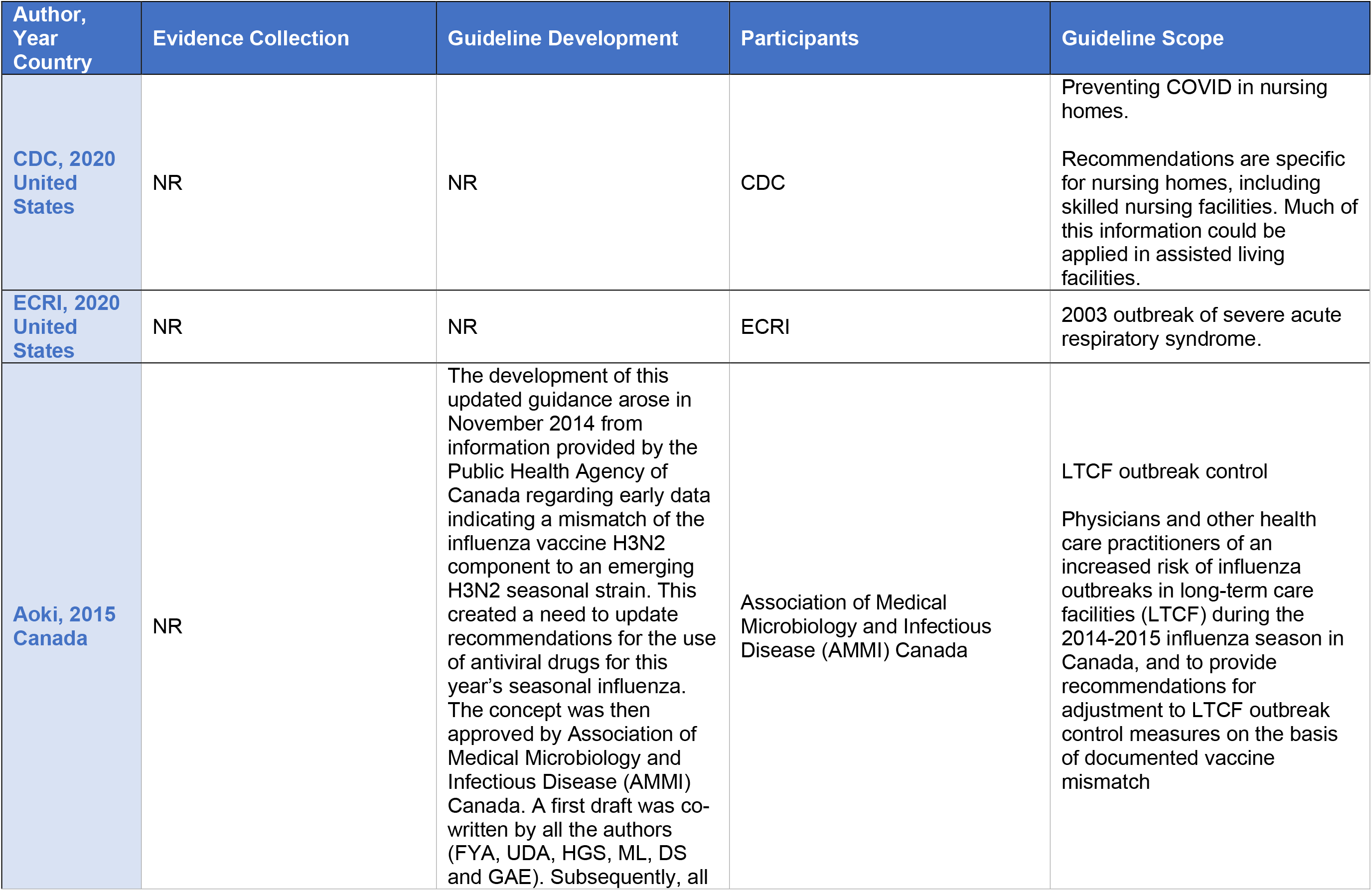

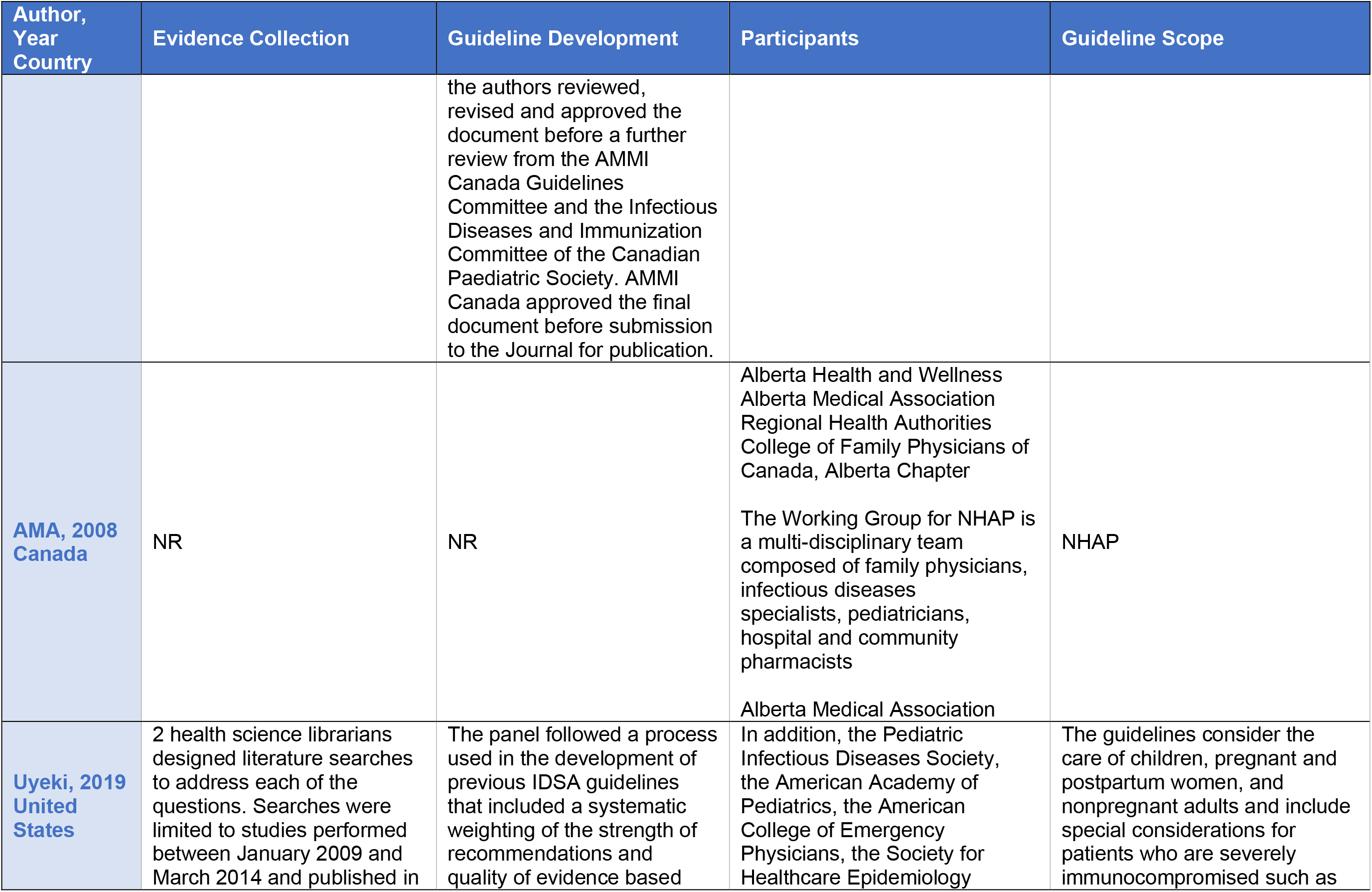

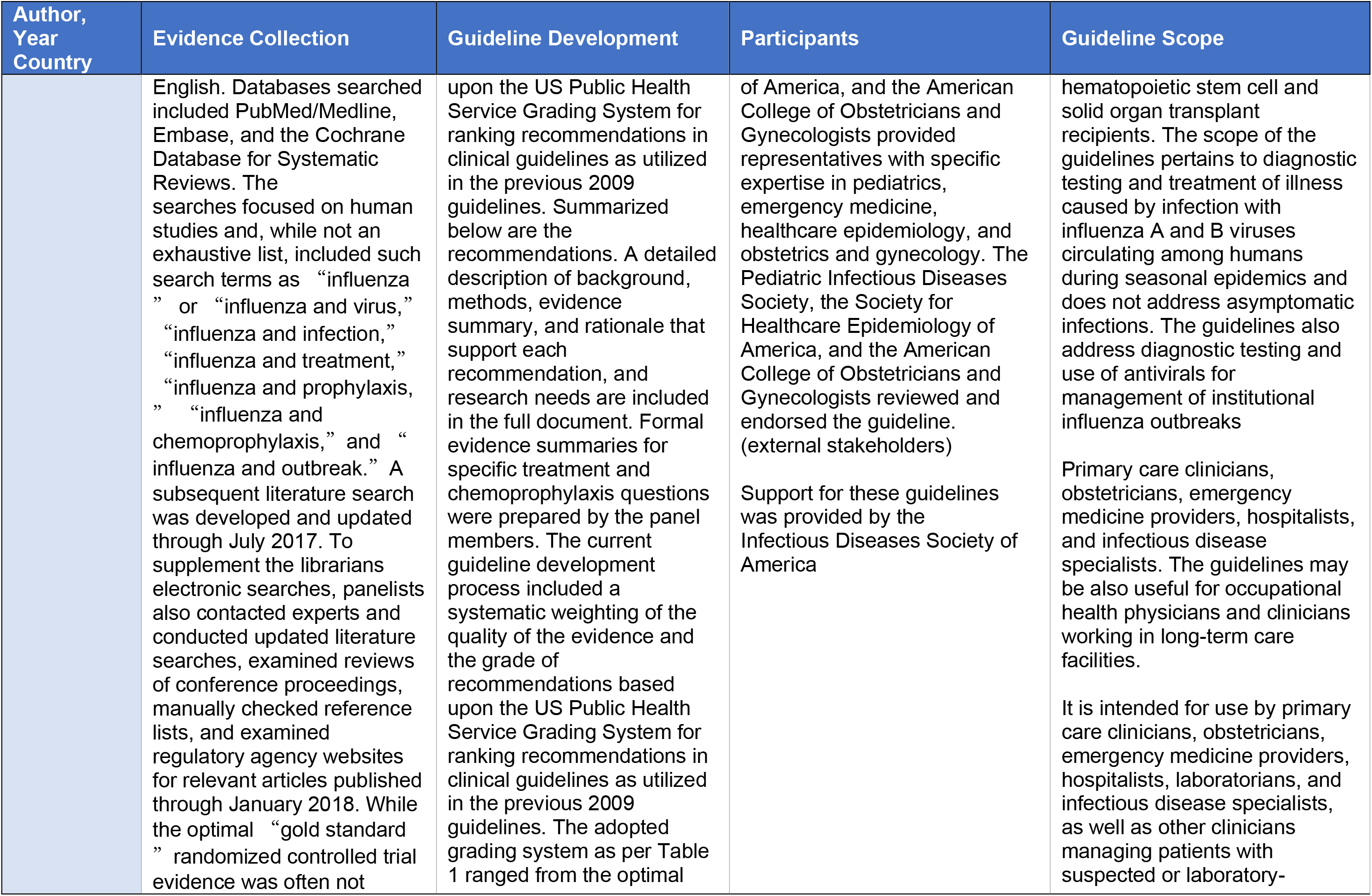

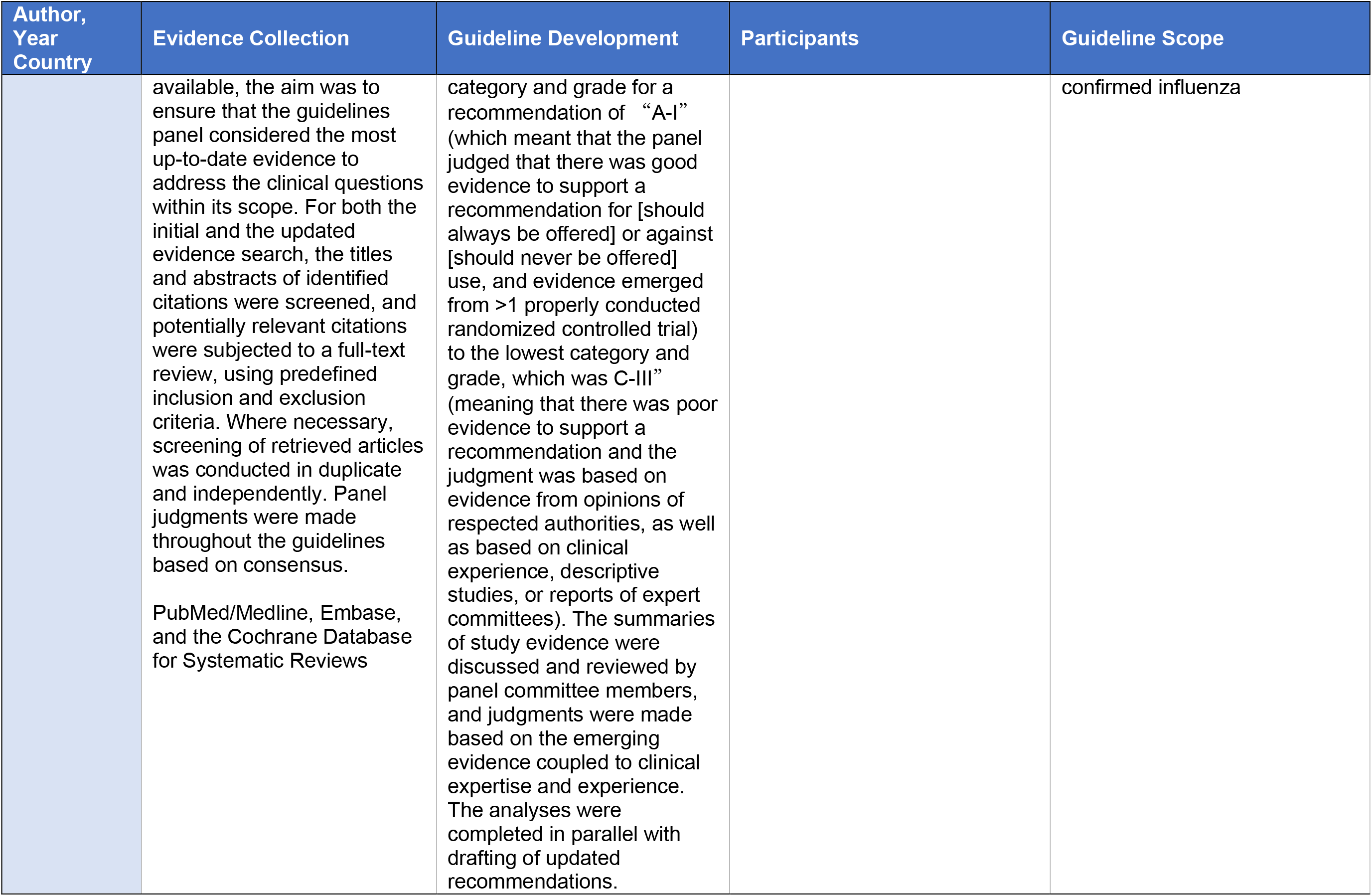

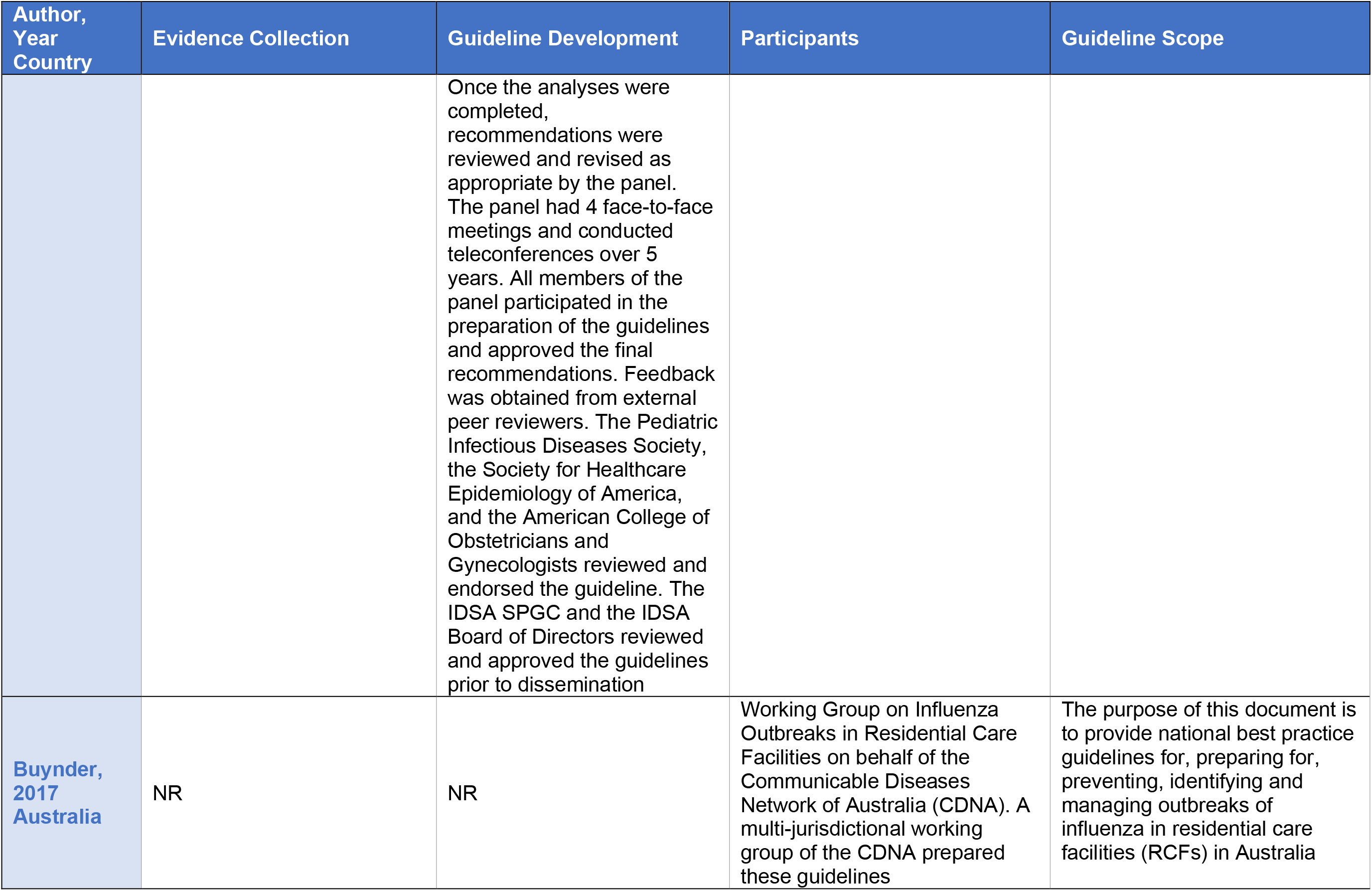

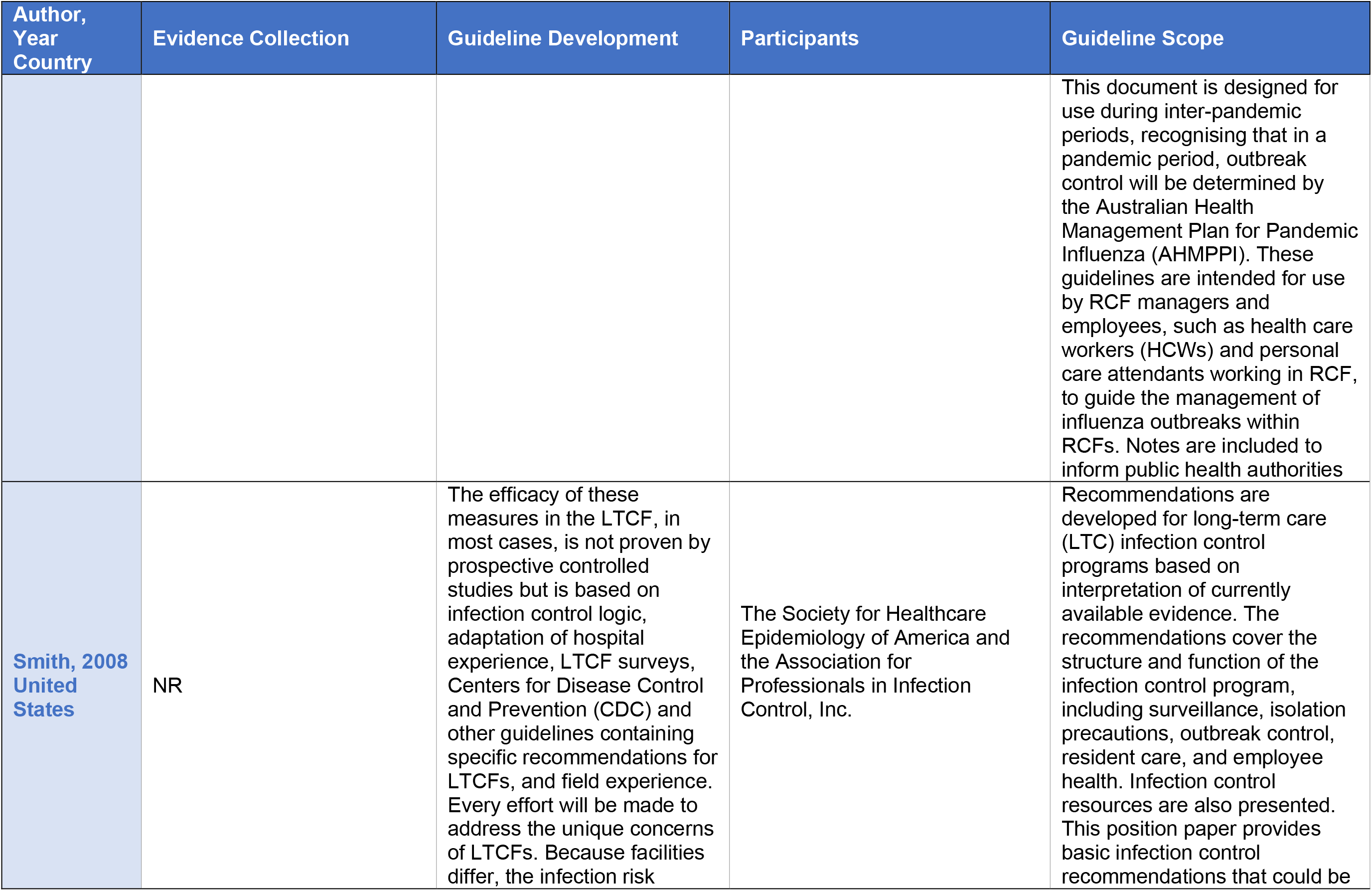

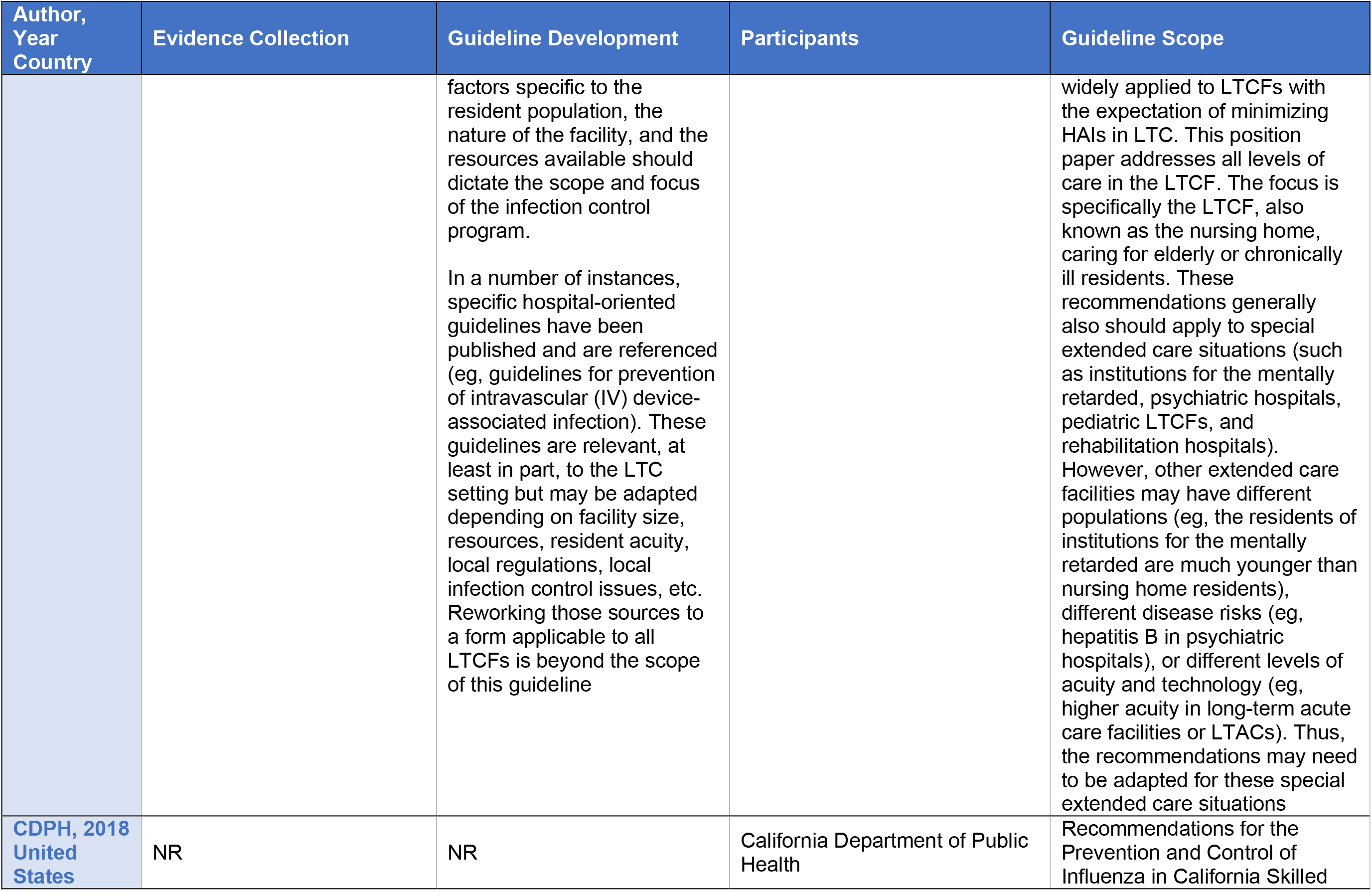

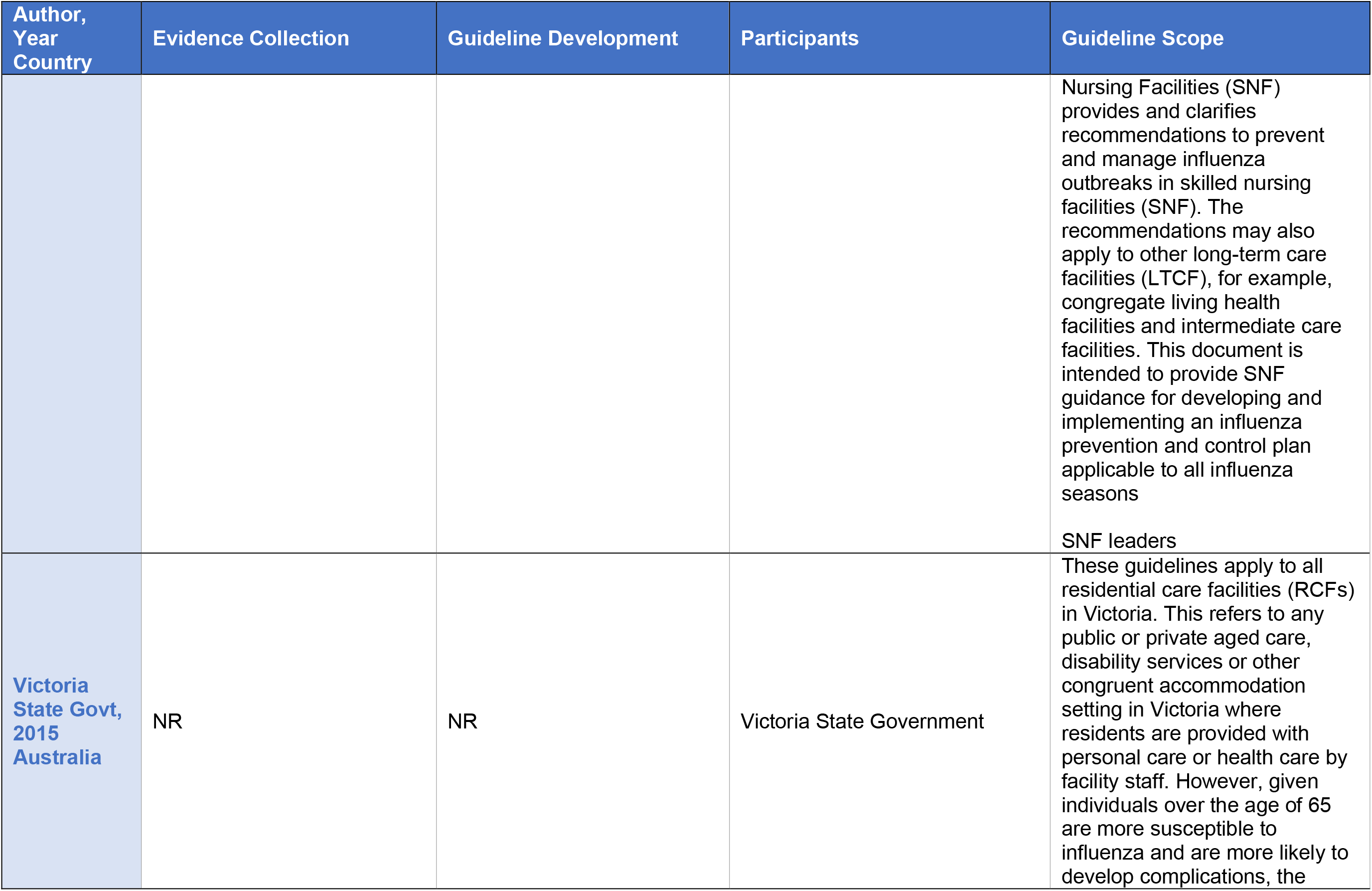

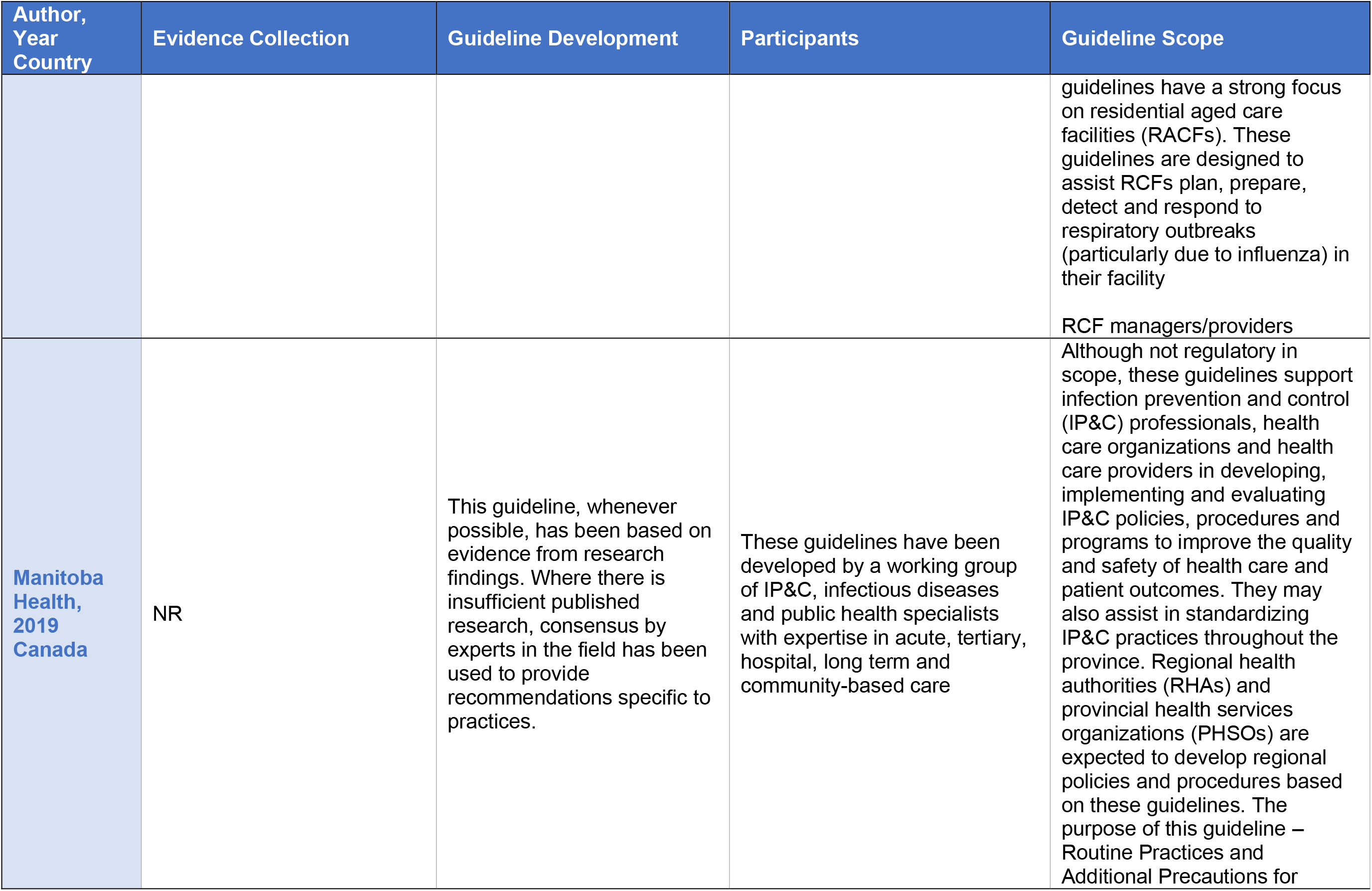

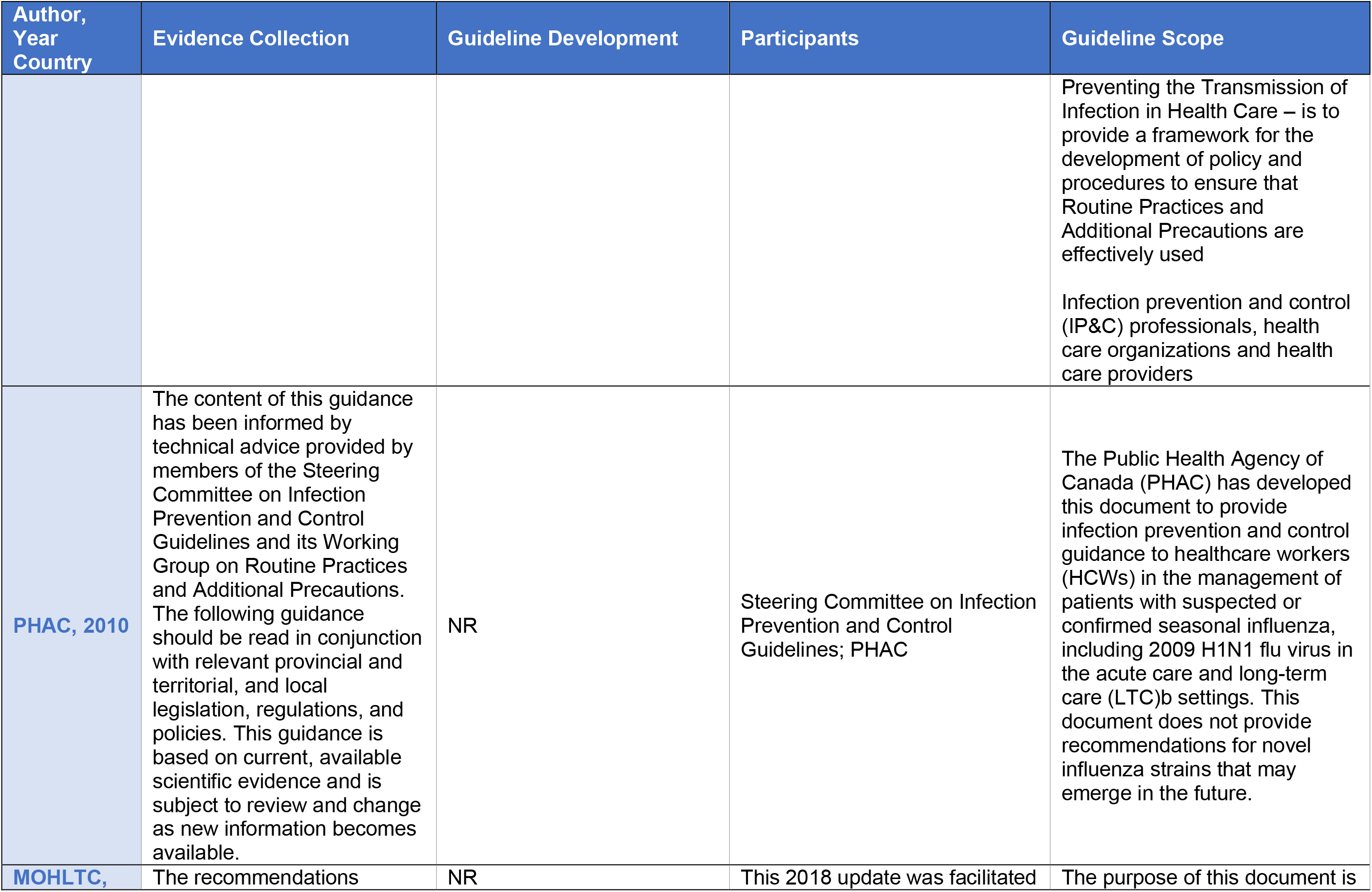

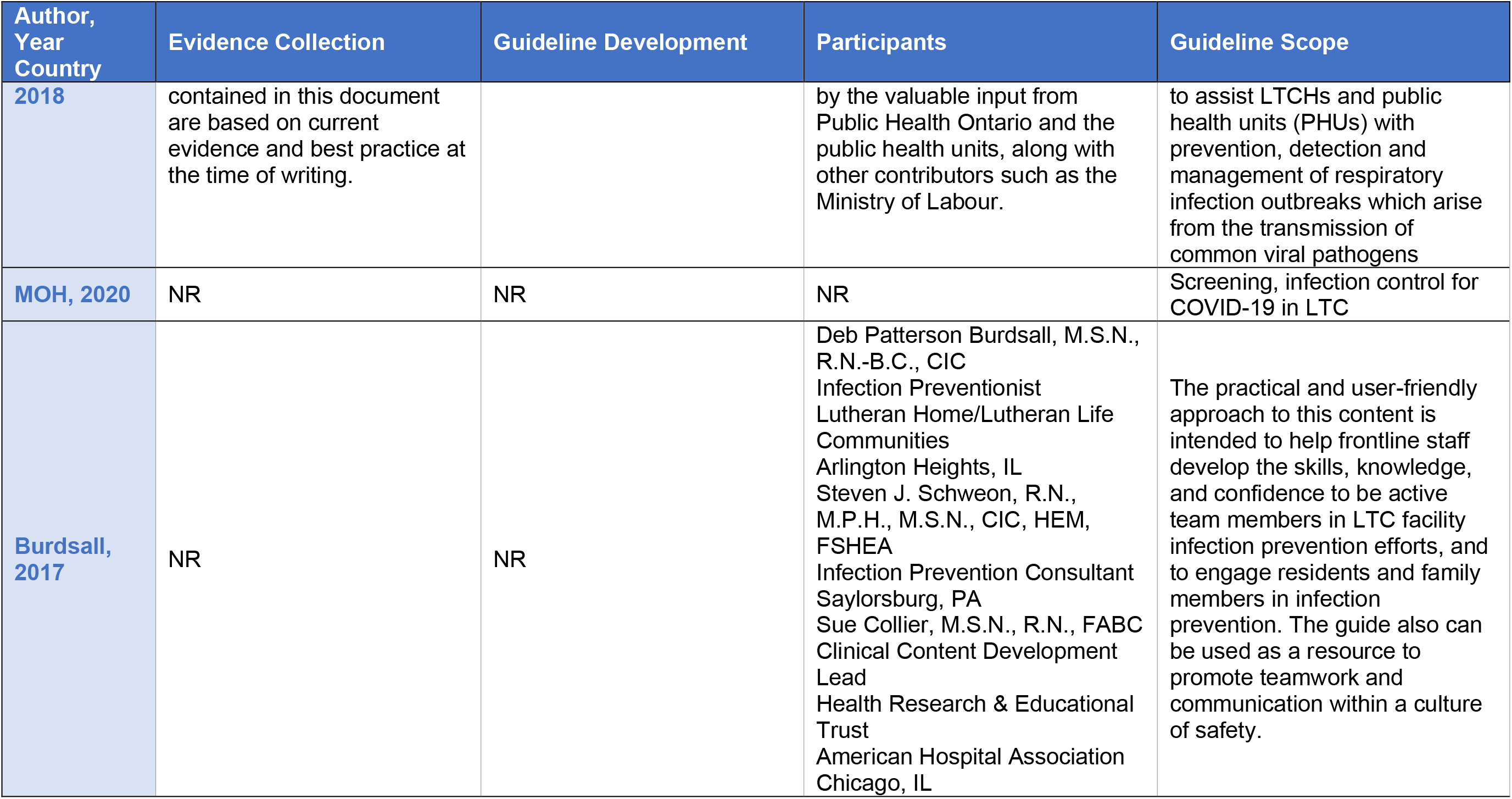

| Author,<br>Year<br>Country | Evidence Collection | Guideline Development | Participants | Guideline Scope |
| --- | --- | --- | --- | --- |
| Rüttimann,<br>2013,<br>United<br>States | NR | Panel discussed scientific evidence as it related to questions posed by the attendees. It then drafted responses that underwent a review process based on group discussion, until unanimous consensus was reached. After the conference, the Panel continued to review and fine-tune the responses. | Americas Health Foundation (Washington, D.C, United States; AHF) and Fighting Infectious Diseases in Emerging Countries (Miami, Florida, United States; FIDEC) convened a conference with a panel of the Region's clinicians and scientists with expertise in influenza | Influenza among the elderly in Latin America |
| Tablan,<br>2004,<br>United<br>States | NR | Category IA. Strongly recommended for implementation and strongly supported by well designed experimental, clinical, or epidemiologic studies.<br>Category IB. Strongly recommended for implementation and supported by some clinical or epidemiologic studies and by strong theoretical rationale.<br>Category IC. Required for implementation, as mandated by federal or state regulation or standard. | The document was prepared by CDC; reviewed by experts in infection control, intensive-care medicine, pulmonology, respiratory therapy, anesthesiology, internal medicine, and pediatrics; and approved by HICPAC. The recommendations are endorsed by the American College of Chest Physicians, American Health Care Association, Association for Professionals of Infection Control and Epidemiology, Infectious Diseases Society of America, Society for Healthcare Epidemiology of America, and Society of Critical Care | The new guidelines are designed to reduce the incidence of health-care-associated pneumonia and other severe, acute lower respiratory tract infections in acute-care hospitals and in other health-care settings (e.g., ambulatory and long-term care institutions) and other facilities where health care is provided.<br>Acute-care hospitals and in other health-care settings |
|  |  | Category II. Suggested for implementation and supported by suggestive clinical or epidemiologic studies or by strong theoretical rationale. No Recommendation; Unresolved Issue. Practices for which insufficient evidence or no consensus exists about efficacy | Medicine. |  |
| Woodhead,<br>2011<br>United Kingdom | NR - referenced previous guideline but no search provided | No description of consensus method | Department of Respiratory Medicine, Manchester Royal Infirmary, Manchester, UK, 2) Dipartimento Toraco Polmonare eCardiocircolatorio, Università degli Studi di Milano, IRCCS Ospedale Maggiore di Milano, Milano, Italy, 3) Chefarzt der Kliniken für Pneumologie und Infektiologie, Ev. Krankenhaus Herne und Augusta-Kranken-Anstalt, Bergstrasse, Bochum, Germany, 4) Department of Medicine, Hospital Universitari Mutua de Terrassa, University of Barcelona, Barcelona, Spain, 5) Pneumologie et Réanimation, Hôtel-Dieu de Paris, 1 Place Parvis Notre-Dame, Paris, France, 6) Microbiology Laboratory, University Hospital Antwerp, | Our objective is to provide evidence-based recommendations for the most common management questions occurring in routine clinical practice in the management of adult patients with LRTI.<br>The target audience for the guidelines is thus all those whose routine practice includes the management of adult LRTI. |
|  |  |  | Edegem, Belgium, 7) Department of Communicable Diseases Control and Prevention, Stockholm County, Stockholm, Sweden, 8) Zentrum für Pneumologie, Diakoniekrankenhaus Rotenburg, Elise-Averdiek-Str. Rotenburg, Germany, 9) Pulmonary Department, Institut Clinic del Torax, Hospital Clinic de Barcelona, IDIBAPS, CIBERES (Ciber de Enfermedades Respiratorias), Facultad de Medicina. Universitat de Barcelona, Barcelona, Spain, 10) Clinical Epidemiology, Julius Center for Health Sciences and Primary Care, University Medical Center Utrecht, Julius Center, Utrecht, The Netherlands, 11) Infectious Diseases, Department of Infection and Immunity, Sheffield School of Medicine and Biomedical Science, University of Sheffield, Royal Hallamshire Hospital, Sheffield, UK and 12) General Practice, Julius Center for Health Sciences and Primary Care, University Medical Center Utrecht, Utrecht, The Netherlands |  |

| Author, Year Country | Evidence Collection | Guideline Development | Participants | Guideline Scope |
| --- | --- | --- | --- | --- |
| **CDC, 2020**<br>**United States** | NR | NR | CDC | Preventing COVID in nursing homes.<br>Recommendations are specific for nursing homes, including skilled nursing facilities. Much of this information could be applied in assisted living facilities. |
| **ECRI, 2020**<br>**United States** | NR | NR | ECRI | 2003 outbreak of severe acute respiratory syndrome. |
| **Aoki, 2015**<br>**Canada** | NR | The development of this updated guidance arose in November 2014 from information provided by the Public Health Agency of Canada regarding early data indicating a mismatch of the influenza vaccine H3N2 component to an emerging H3N2 seasonal strain. This created a need to update recommendations for the use of antiviral drugs for this year's seasonal influenza. The concept was then approved by Association of Medical Microbiology and Infectious Disease (AMMI) Canada. A first draft was co-written by all the authors (FYA, UDA, HGS, ML, DS and GAE). Subsequently, all the authors reviewed, revised and approved the document before a further review from the AMMI Canada Guidelines Committee and the Infectious Diseases and Immunization Committee of the Canadian Paediatric Society. AMMI Canada approved the final document before submission to the Journal for publication. | Association of Medical Microbiology and Infectious Disease (AMMI) Canada | LTCF outbreak control<br>Physicians and other health care practitioners of an increased risk of influenza outbreaks in long-term care facilities (LTCF) during the 2014-2015 influenza season in Canada, and to provide recommendations for adjustment to LTCF outbreak control measures on the basis of documented vaccine mismatch |
| **AMA, 2008**<br>**Canada** | NR | NR | Alberta Health and Wellness<br>Alberta Medical Association<br>Regional Health Authorities<br>College of Family Physicians of Canada, Alberta Chapter<br>The Working Group for NHAP is a multi-disciplinary team composed of family physicians, infectious diseases specialists, pediatricians, hospital and community pharmacists<br>Alberta Medical Association | NHAP |
| **Uyeki, 2019**<br>**United States** | 2 health science librarians designed literature searches to address each of the questions. Searches were limited to studies performed between January 2009 and March 2014 and published in | The panel followed a process used in the development of previous IDSA guidelines that included a systematic weighting of the strength of recommendations and quality of evidence based | In addition, the Pediatric Infectious Diseases Society, the American Academy of Pediatrics, the American College of Emergency Physicians, the Society for Healthcare Epidemiology | The guidelines consider the care of children, pregnant and postpartum women, and nonpregnant adults and include special considerations for patients who are severely immunocompromised such as |
|  | English. Databases searched included PubMed/Medline, Embase, and the Cochrane Database for Systematic Reviews. The searches focused on human studies and, while not an exhaustive list, included such search terms as “influenza” or “influenza and virus,” “influenza and infection,” “influenza and treatment,” “influenza and prophylaxis,” “influenza and chemoprophylaxis,” and “influenza and outbreak.” A subsequent literature search was developed and updated through July 2017. To supplement the librarians electronic searches, panelists also contacted experts and conducted updated literature searches, examined reviews of conference proceedings, manually checked reference lists, and examined regulatory agency websites for relevant articles published through January 2018. While the optimal “gold standard” randomized controlled trial evidence was often not | upon the US Public Health Service Grading System for ranking recommendations in clinical guidelines as utilized in the previous 2009 guidelines. Summarized below are the recommendations. A detailed description of background, methods, evidence summary, and rationale that support each recommendation, and research needs are included in the full document. Formal evidence summaries for specific treatment and chemoprophylaxis questions were prepared by the panel members. The current guideline development process included a systematic weighting of the quality of the evidence and the grade of recommendations based upon the US Public Health Service Grading System for ranking recommendations in clinical guidelines as utilized in the previous 2009 guidelines. The adopted grading system as per Table 1 ranged from the optimal | of America, and the American College of Obstetricians and Gynecologists provided representatives with specific expertise in pediatrics, emergency medicine, healthcare epidemiology, and obstetrics and gynecology. The Pediatric Infectious Diseases Society, the Society for Healthcare Epidemiology of America, and the American College of Obstetricians and Gynecologists reviewed and endorsed the guideline. (external stakeholders)<br>Support for these guidelines was provided by the Infectious Diseases Society of America | hematopoietic stem cell and solid organ transplant recipients. The scope of the guidelines pertains to diagnostic testing and treatment of illness caused by infection with influenza A and B viruses circulating among humans during seasonal epidemics and does not address asymptomatic infections. The guidelines also address diagnostic testing and use of antivirals for management of institutional influenza outbreaks<br>Primary care clinicians, obstetricians, emergency medicine providers, hospitalists, and infectious disease specialists. The guidelines may be also useful for occupational health physicians and clinicians working in long-term care facilities.<br>It is intended for use by primary care clinicians, obstetricians, emergency medicine providers, hospitalists, laboratorians, and infectious disease specialists, as well as other clinicians managing patients with suspected or laboratory- |
|  | available, the aim was to ensure that the guidelines panel considered the most up-to-date evidence to address the clinical questions within its scope. For both the initial and the updated evidence search, the titles and abstracts of identified citations were screened, and potentially relevant citations were subjected to a full-text review, using predefined inclusion and exclusion criteria. Where necessary, screening of retrieved articles was conducted in duplicate and independently. Panel judgments were made throughout the guidelines based on consensus.<br>PubMed/Medline, Embase, and the Cochrane Database for Systematic Reviews | category and grade for a recommendation of “A-I” (which meant that the panel judged that there was good evidence to support a recommendation for [should always be offered] or against [should never be offered] use, and evidence emerged from >1 properly conducted randomized controlled trial) to the lowest category and grade, which was C-III” (meaning that there was poor evidence to support a recommendation and the judgment was based on evidence from opinions of respected authorities, as well as based on clinical experience, descriptive studies, or reports of expert committees). The summaries of study evidence were discussed and reviewed by panel committee members, and judgments were made based on the emerging evidence coupled to clinical expertise and experience. The analyses were completed in parallel with drafting of updated recommendations. |  | confirmed influenza |
|  |  | Once the analyses were completed, recommendations were reviewed and revised as appropriate by the panel. The panel had 4 face-to-face meetings and conducted teleconferences over 5 years. All members of the panel participated in the preparation of the guidelines and approved the final recommendations. Feedback was obtained from external peer reviewers. The Pediatric Infectious Diseases Society, the Society for Healthcare Epidemiology of America, and the American College of Obstetricians and Gynecologists reviewed and endorsed the guideline. The IDSA SPGC and the IDSA Board of Directors reviewed and approved the guidelines prior to dissemination |  |  |
| **Buynder,**<br>**2017**<br>**Australia** | NR | NR | Working Group on Influenza Outbreaks in Residential Care Facilities on behalf of the Communicable Diseases Network of Australia (CDNA). A multi-jurisdictional working group of the CDNA prepared these guidelines | The purpose of this document is to provide national best practice guidelines for, preparing for, preventing, identifying and managing outbreaks of influenza in residential care facilities (RCFs) in Australia |
|  |  |  |  | This document is designed for use during inter-pandemic periods, recognising that in a pandemic period, outbreak control will be determined by the Australian Health Management Plan for Pandemic Influenza (AHMPPI). These guidelines are intended for use by RCF managers and employees, such as health care workers (HCWs) and personal care attendants working in RCF, to guide the management of influenza outbreaks within RCFs. Notes are included to inform public health authorities |
| **Smith, 2008**<br>**United States** | NR | The efficacy of these measures in the LTCF, in most cases, is not proven by prospective controlled studies but is based on infection control logic, adaptation of hospital experience, LTCF surveys, Centers for Disease Control and Prevention (CDC) and other guidelines containing specific recommendations for LTCFs, and field experience. Every effort will be made to address the unique concerns of LTCFs. Because facilities differ, the infection risk | The Society for Healthcare Epidemiology of America and the Association for Professionals in Infection Control, Inc. | Recommendations are developed for long-term care (LTC) infection control programs based on interpretation of currently available evidence. The recommendations cover the structure and function of the infection control program, including surveillance, isolation precautions, outbreak control, resident care, and employee health. Infection control resources are also presented. This position paper provides basic infection control recommendations that could be |
|  |  | factors specific to the resident population, the nature of the facility, and the resources available should dictate the scope and focus of the infection control program.<br>In a number of instances, specific hospital-oriented guidelines have been published and are referenced (eg, guidelines for prevention of intravascular (IV) device-associated infection). These guidelines are relevant, at least in part, to the LTC setting but may be adapted depending on facility size, resources, resident acuity, local regulations, local infection control issues, etc. Reworking those sources to a form applicable to all LTCFs is beyond the scope of this guideline |  | widely applied to LTCFs with the expectation of minimizing HAIs in LTC. This position paper addresses all levels of care in the LTCF. The focus is specifically the LTCF, also known as the nursing home, caring for elderly or chronically ill residents. These recommendations generally also should apply to special extended care situations (such as institutions for the mentally retarded, psychiatric hospitals, pediatric LTCFs, and rehabilitation hospitals). However, other extended care facilities may have different populations (eg, the residents of institutions for the mentally retarded are much younger than nursing home residents), different disease risks (eg, hepatitis B in psychiatric hospitals), or different levels of acuity and technology (eg, higher acuity in long-term acute care facilities or LTACs). Thus, the recommendations may need to be adapted for these special extended care situations |
| **CDPH, 2018**<br>**United**<br>**States** | NR | NR | California Department of Public Health | Recommendations for the Prevention and Control of Influenza in California Skilled |
|  |  |  |  | Nursing Facilities (SNF) provides and clarifies recommendations to prevent and manage influenza outbreaks in skilled nursing facilities (SNF). The recommendations may also apply to other long-term care facilities (LTCF), for example, congregate living health facilities and intermediate care facilities. This document is intended to provide SNF guidance for developing and implementing an influenza prevention and control plan applicable to all influenza seasons<br>SNF leaders |
| Victoria<br>State Govt,<br>2015<br>Australia | NR | NR | Victoria State Government | These guidelines apply to all residential care facilities (RCFs) in Victoria. This refers to any public or private aged care, disability services or other congruent accommodation setting in Victoria where residents are provided with personal care or health care by facility staff. However, given individuals over the age of 65 are more susceptible to influenza and are more likely to develop complications, the<br>guidelines have a strong focus on residential aged care facilities (RACFs). These guidelines are designed to assist RCFs plan, prepare, detect and respond to respiratory outbreaks (particularly due to influenza) in their facility<br>RCF managers/providers |
| Manitoba Health, 2019 Canada | NR | This guideline, whenever possible, has been based on evidence from research findings. Where there is insufficient published research, consensus by experts in the field has been used to provide recommendations specific to practices. | These guidelines have been developed by a working group of IP&C, infectious diseases and public health specialists with expertise in acute, tertiary, hospital, long term and community-based care | Although not regulatory in scope, these guidelines support infection prevention and control (IP&C) professionals, health care organizations and health care providers in developing, implementing and evaluating IP&C policies, procedures and programs to improve the quality and safety of health care and patient outcomes. They may also assist in standardizing IP&C practices throughout the province. Regional health authorities (RHAs) and provincial health services organizations (PHSOs) are expected to develop regional policies and procedures based on these guidelines. The purpose of this guideline – Routine Practices and Additional Precautions for |
|  |  |  |  | Preventing the Transmission of Infection in Health Care – is to provide a framework for the development of policy and procedures to ensure that Routine Practices and Additional Precautions are effectively used<br>Infection prevention and control (IP&C) professionals, health care organizations and health care providers |
| PHAC, 2010 | The content of this guidance has been informed by technical advice provided by members of the Steering Committee on Infection Prevention and Control Guidelines and its Working Group on Routine Practices and Additional Precautions. The following guidance should be read in conjunction with relevant provincial and territorial, and local legislation, regulations, and policies. This guidance is based on current, available scientific evidence and is subject to review and change as new information becomes available. | NR | Steering Committee on Infection Prevention and Control Guidelines; PHAC | The Public Health Agency of Canada (PHAC) has developed this document to provide infection prevention and control guidance to healthcare workers (HCWs) in the management of patients with suspected or confirmed seasonal influenza, including 2009 H1N1 flu virus in the acute care and long-term care (LTC)b settings. This document does not provide recommendations for novel influenza strains that may emerge in the future. |
| MOHLTC, | The recommendations | NR | This 2018 update was facilitated | The purpose of this document is |

| Author, Year, Country | Evidence Collection | Guideline Development | Participants | Guideline Scope |
| --- | --- | --- | --- | --- |
| **2018** | contained in this document are based on current evidence and best practice at the time of writing. |  | by the valuable input from Public Health Ontario and the public health units, along with other contributors such as the Ministry of Labour. | to assist LTCHs and public health units (PHUs) with prevention, detection and management of respiratory infection outbreaks which arise from the transmission of common viral pathogens |
| **MOH, 2020** | NR | NR | NR | Screening, infection control for COVID-19 in LTC |
| **Burdsall, 2017** | NR | NR | Deb Patterson Burdsall, M.S.N., R.N.-B.C., CIC<br>Infection Preventionist<br>Lutheran Home/Lutheran Life Communities<br>Arlington Heights, IL<br>Steven J. Schweon, R.N., M.P.H., M.S.N., CIC, HEM, FSHEA<br>Infection Prevention Consultant<br>Saylorsburg, PA<br>Sue Collier, M.S.N., R.N., FABC<br>Clinical Content Development Lead<br>Health Research & Educational Trust<br>American Hospital Association<br>Chicago, IL | The practical and user-friendly approach to this content is intended to help frontline staff develop the skills, knowledge, and confidence to be active team members in LTC facility infection prevention efforts, and to engage residents and family members in infection prevention. The guide also can be used as a resource to promote teamwork and communication within a culture of safety. |

## APPENDIX 5 – QUALITY APPRAISAL RESULTS FOR CLINICAL PRACTICE GUIDELINES

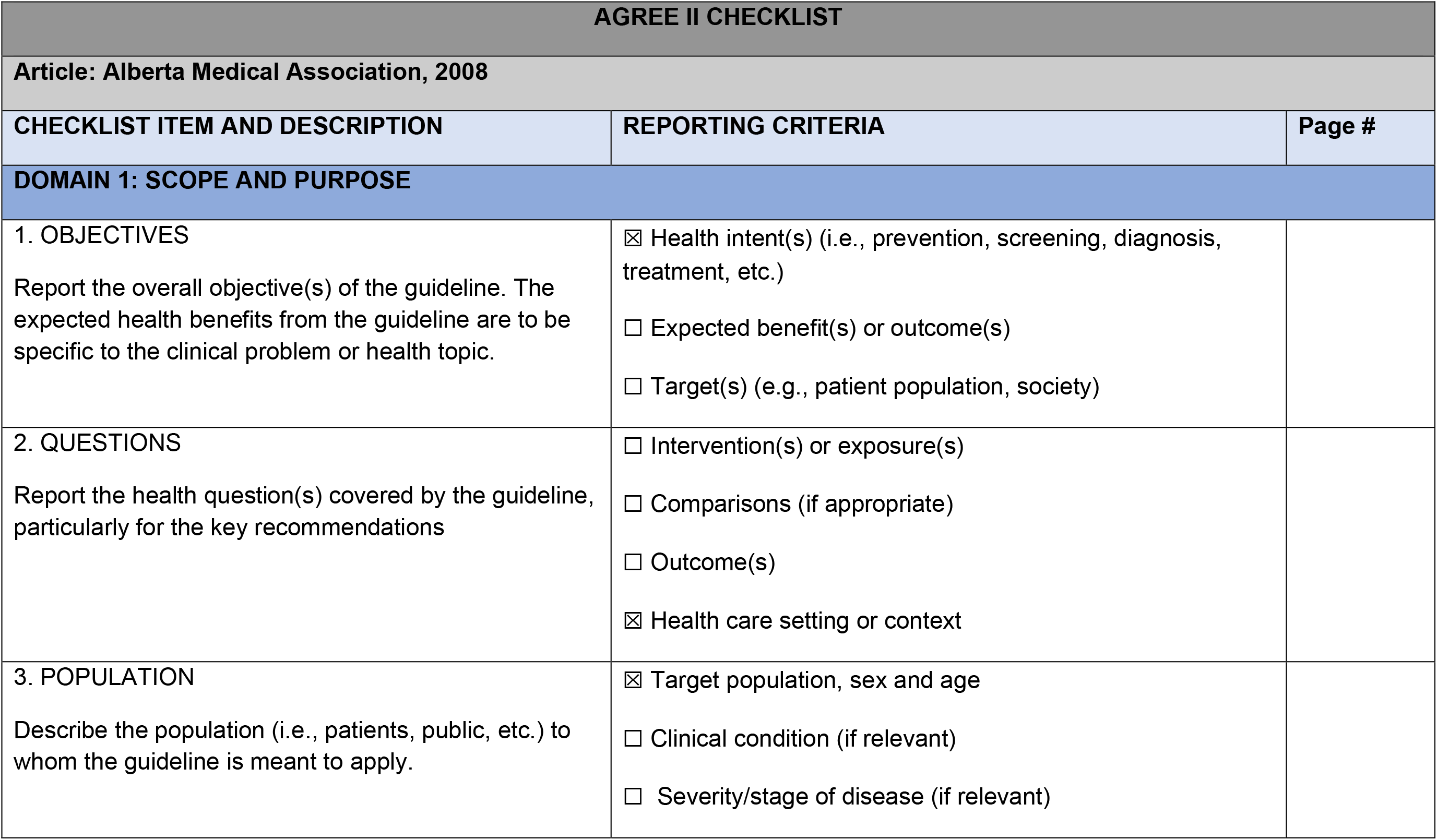

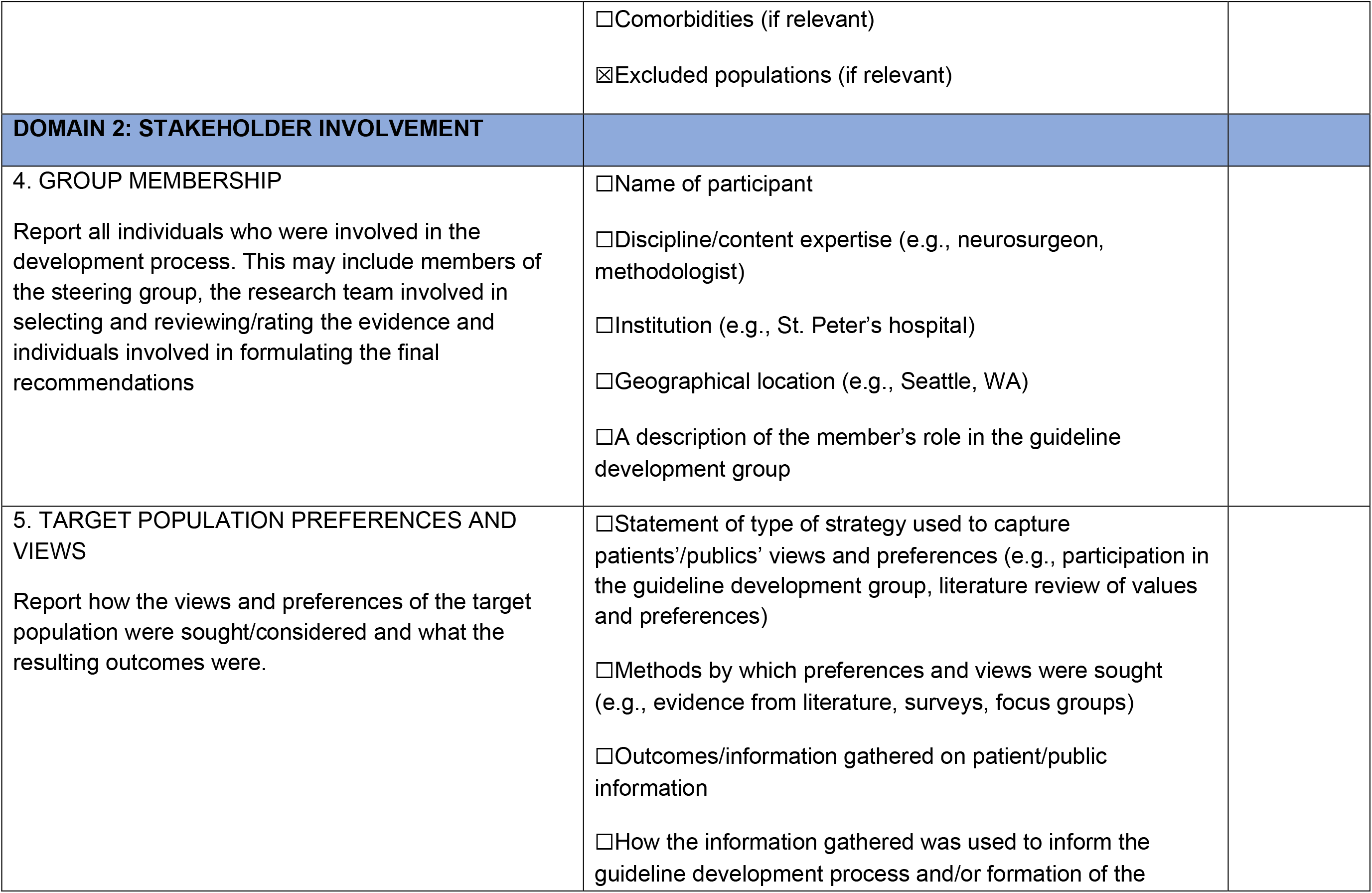

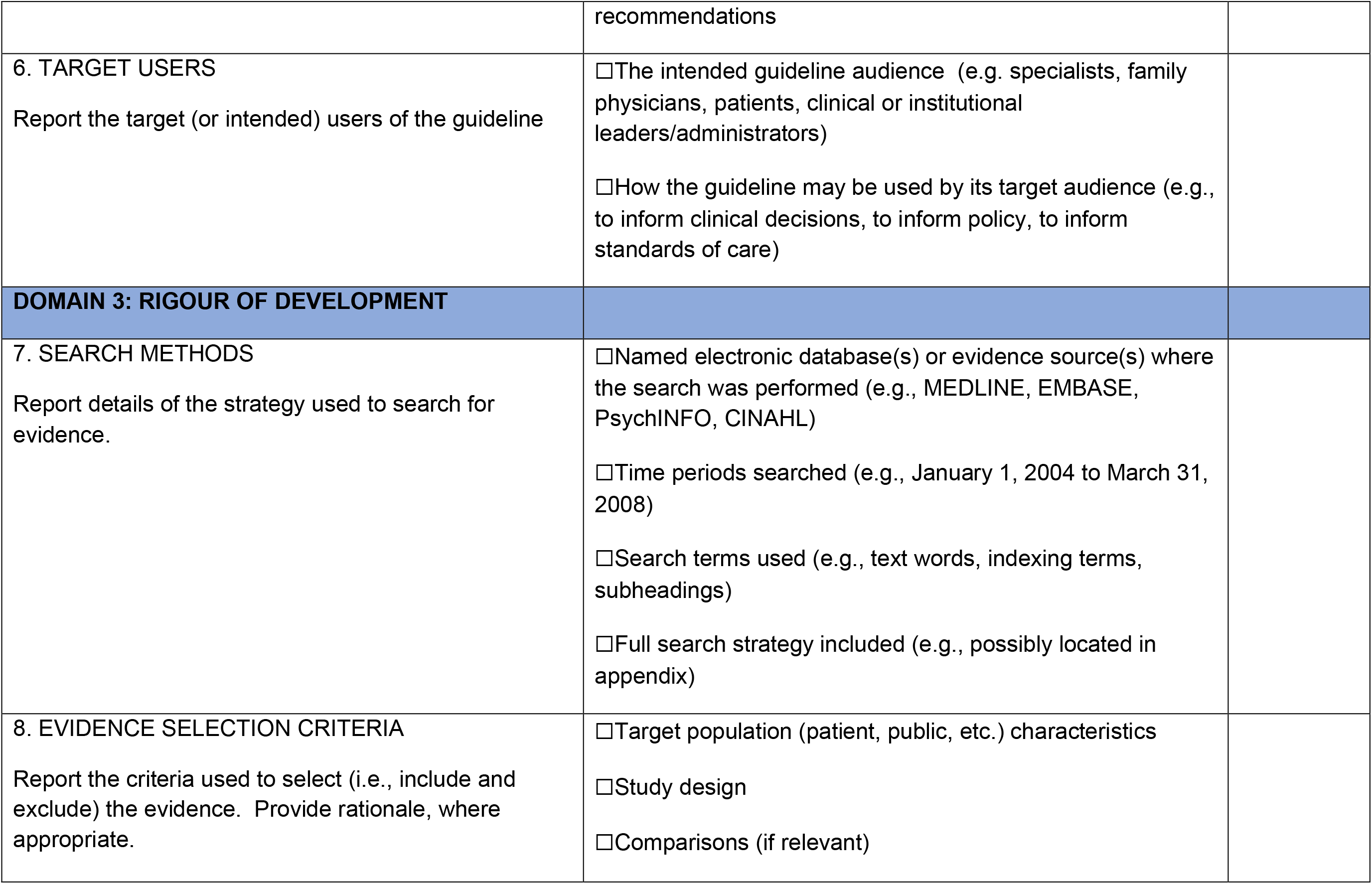

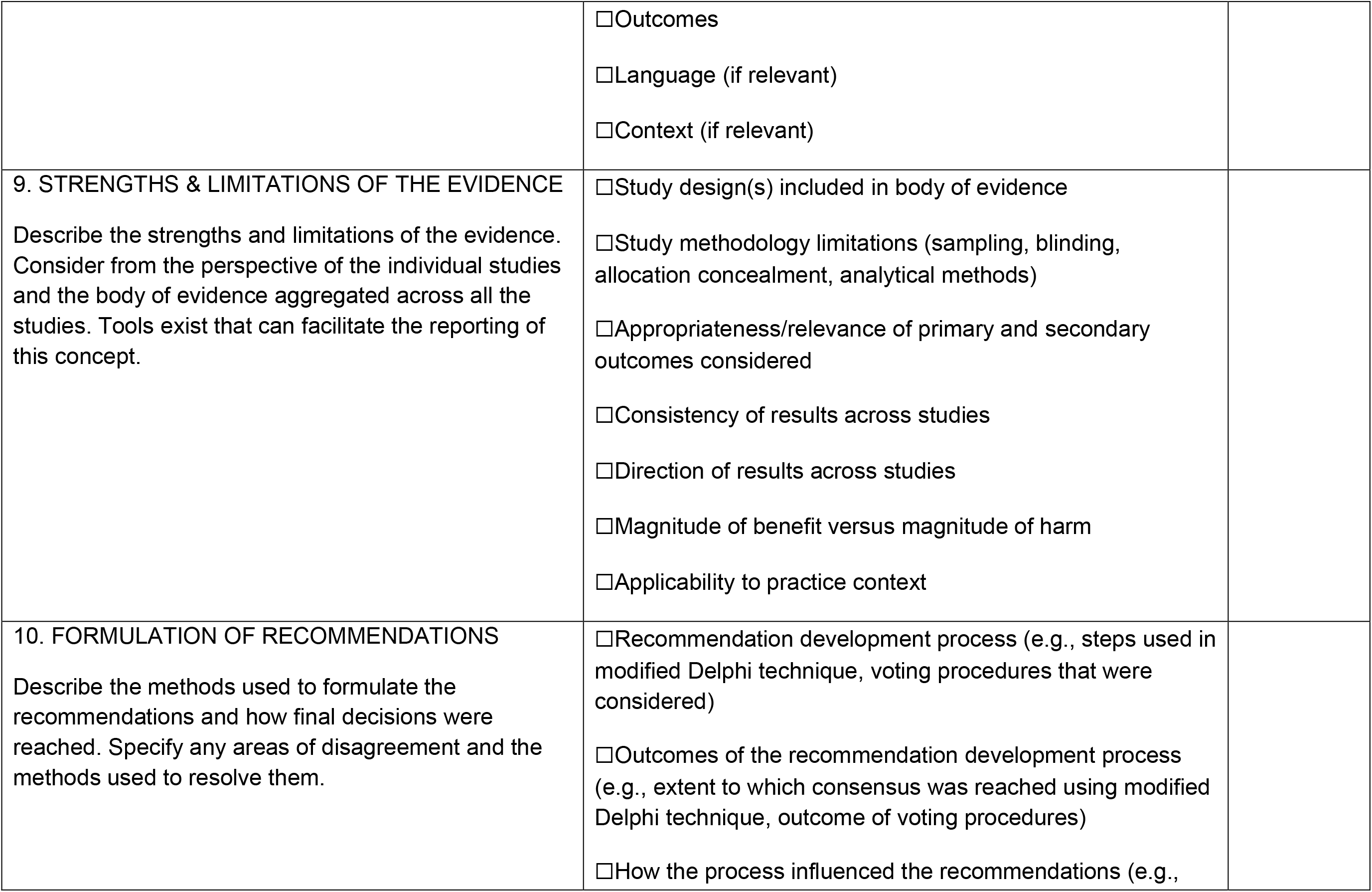

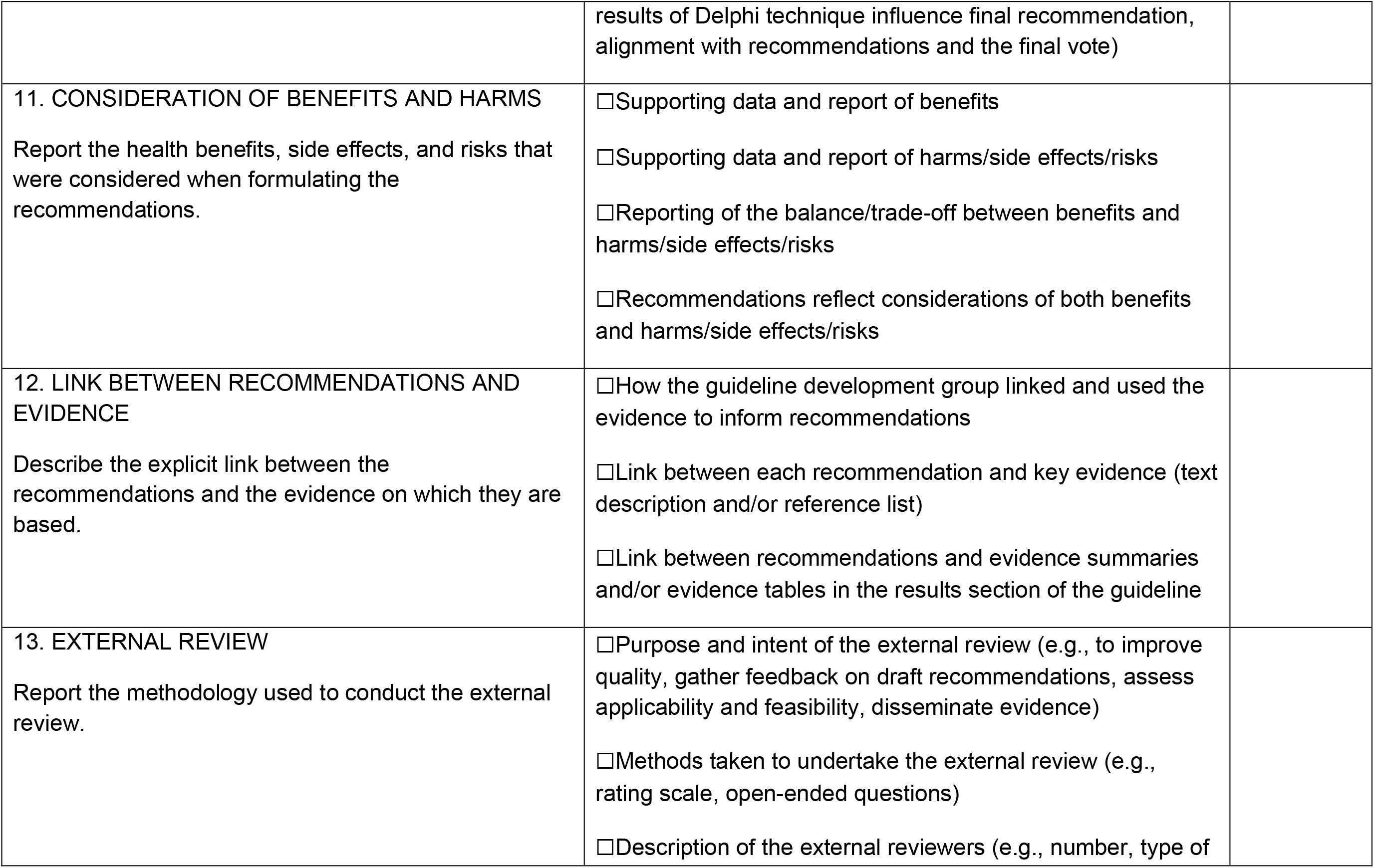

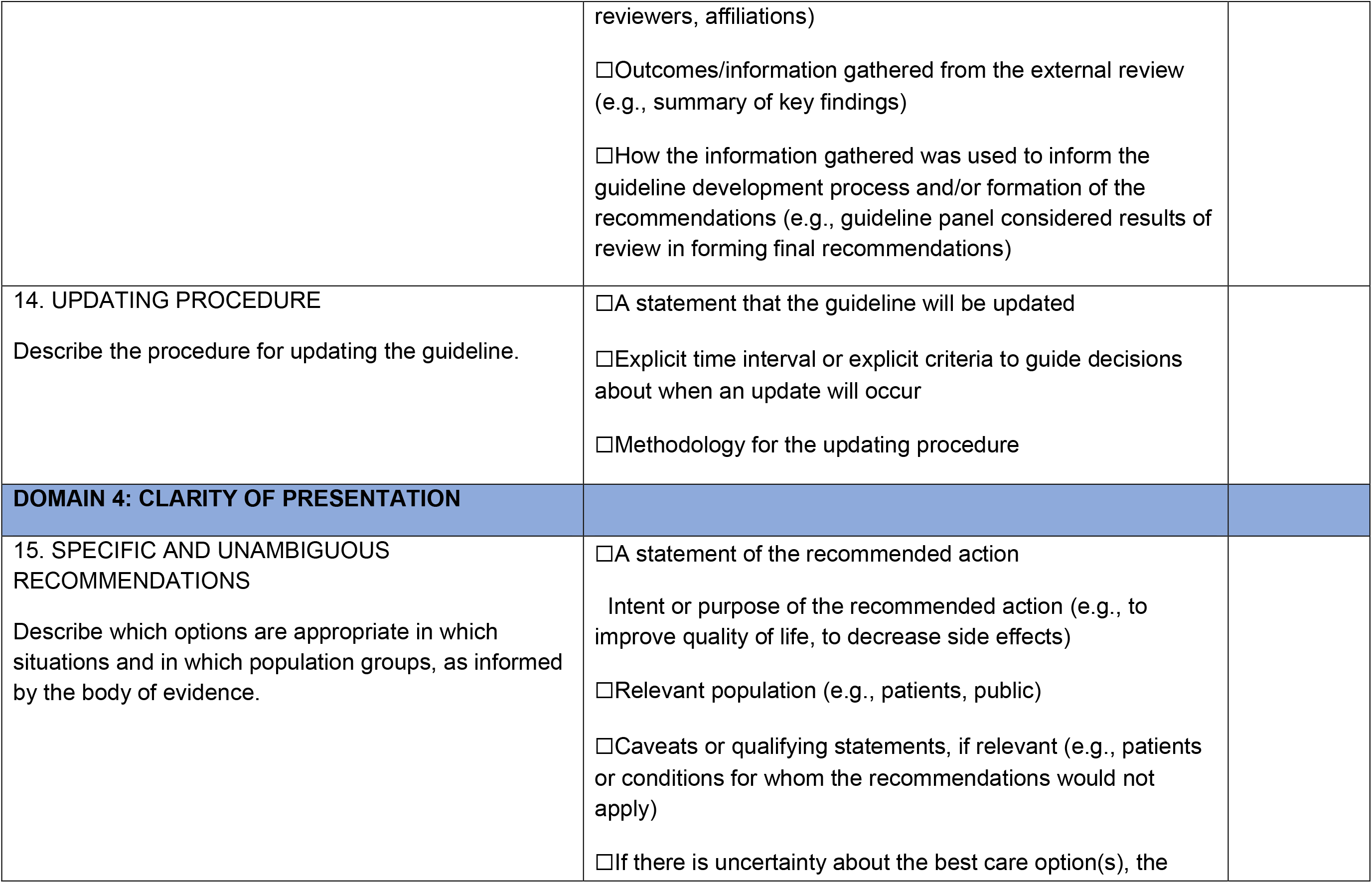

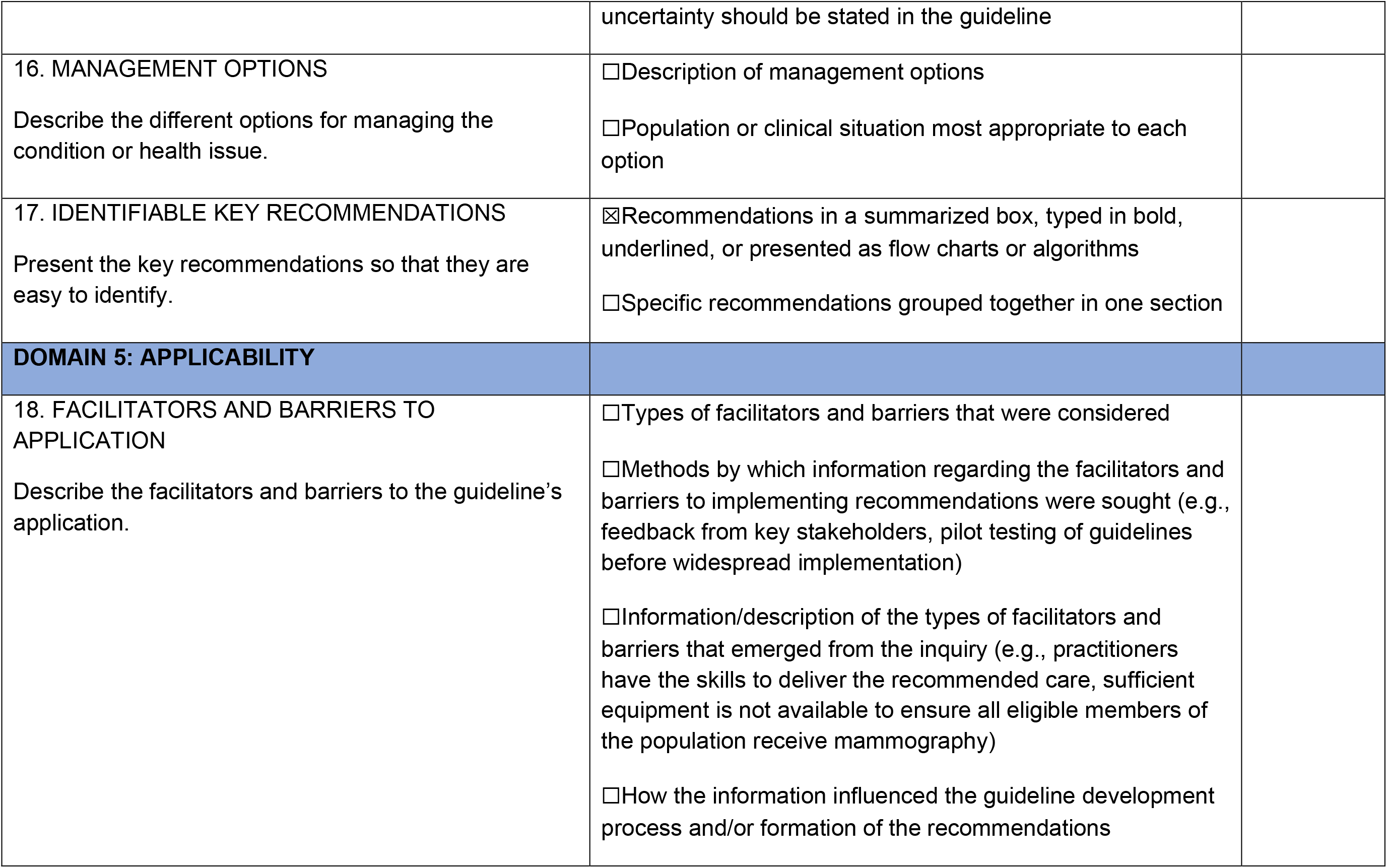

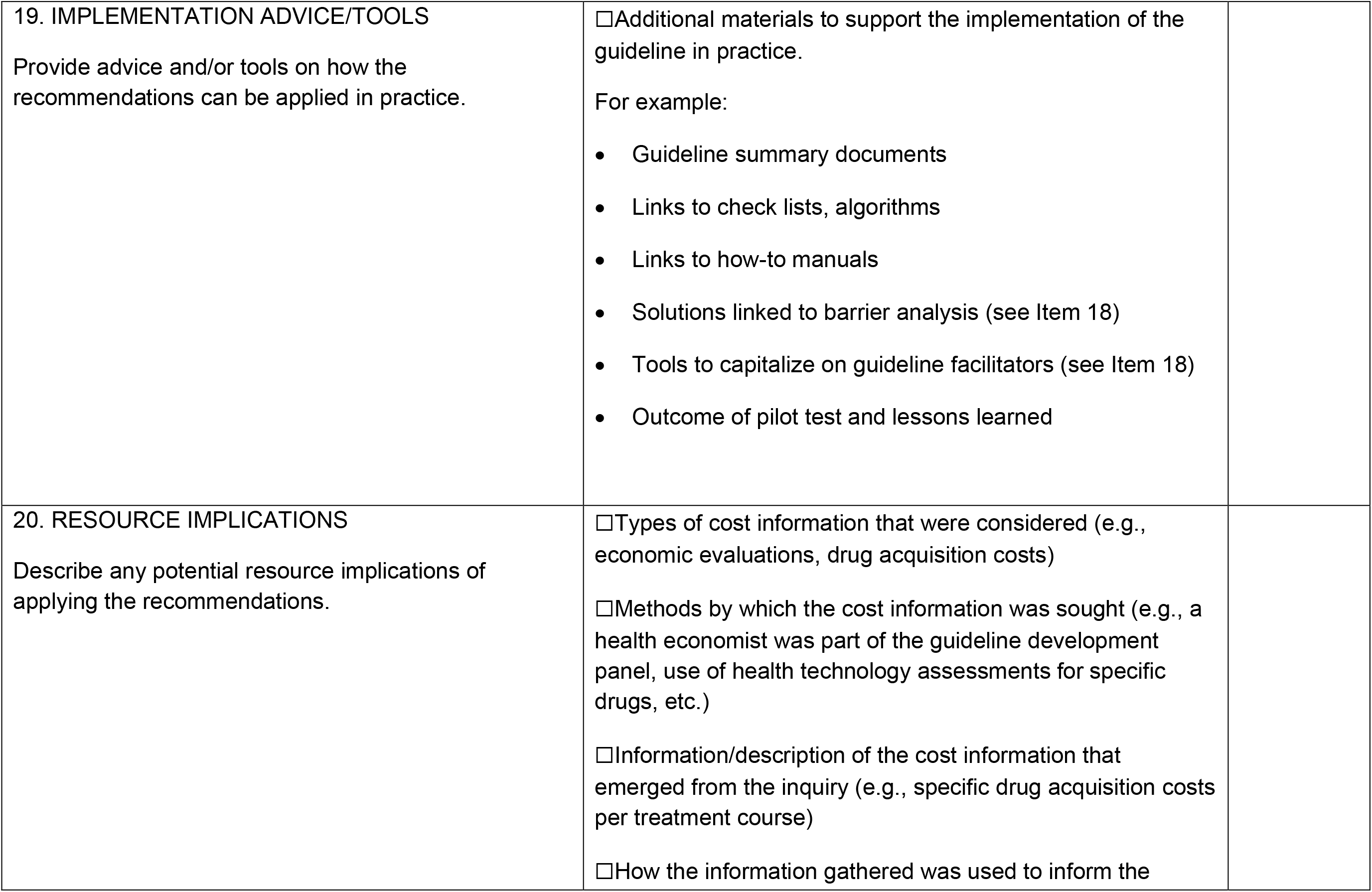

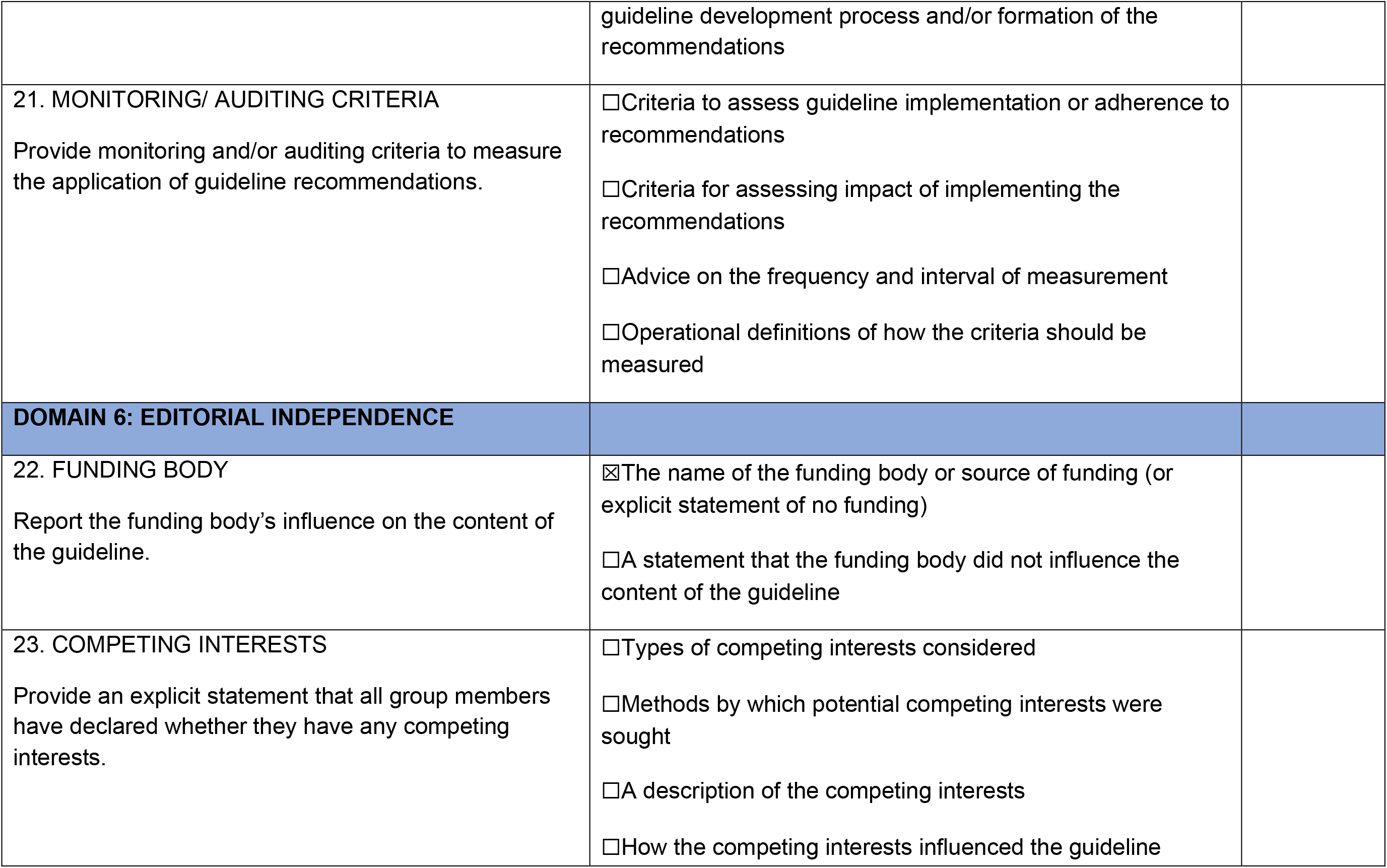

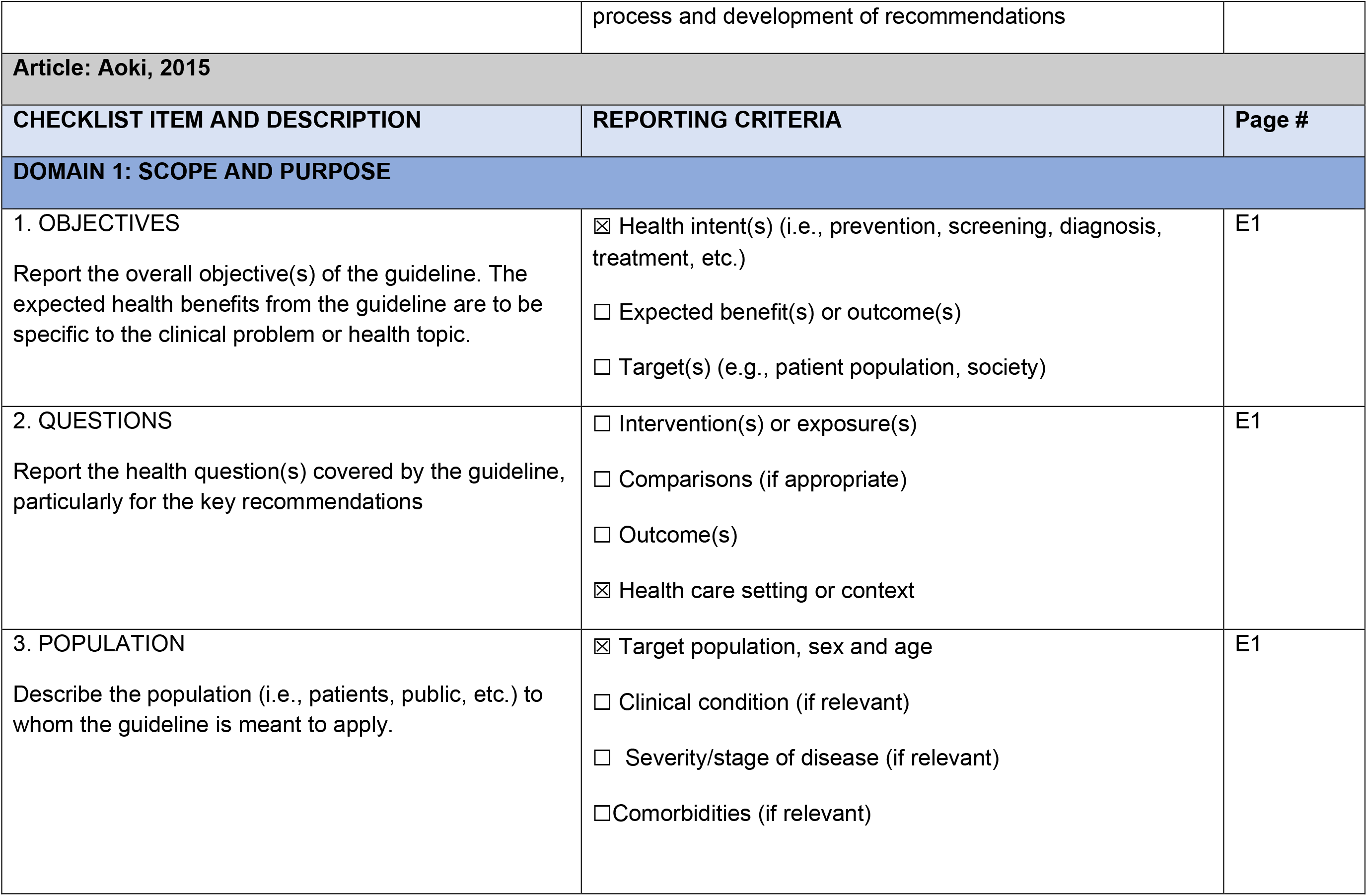

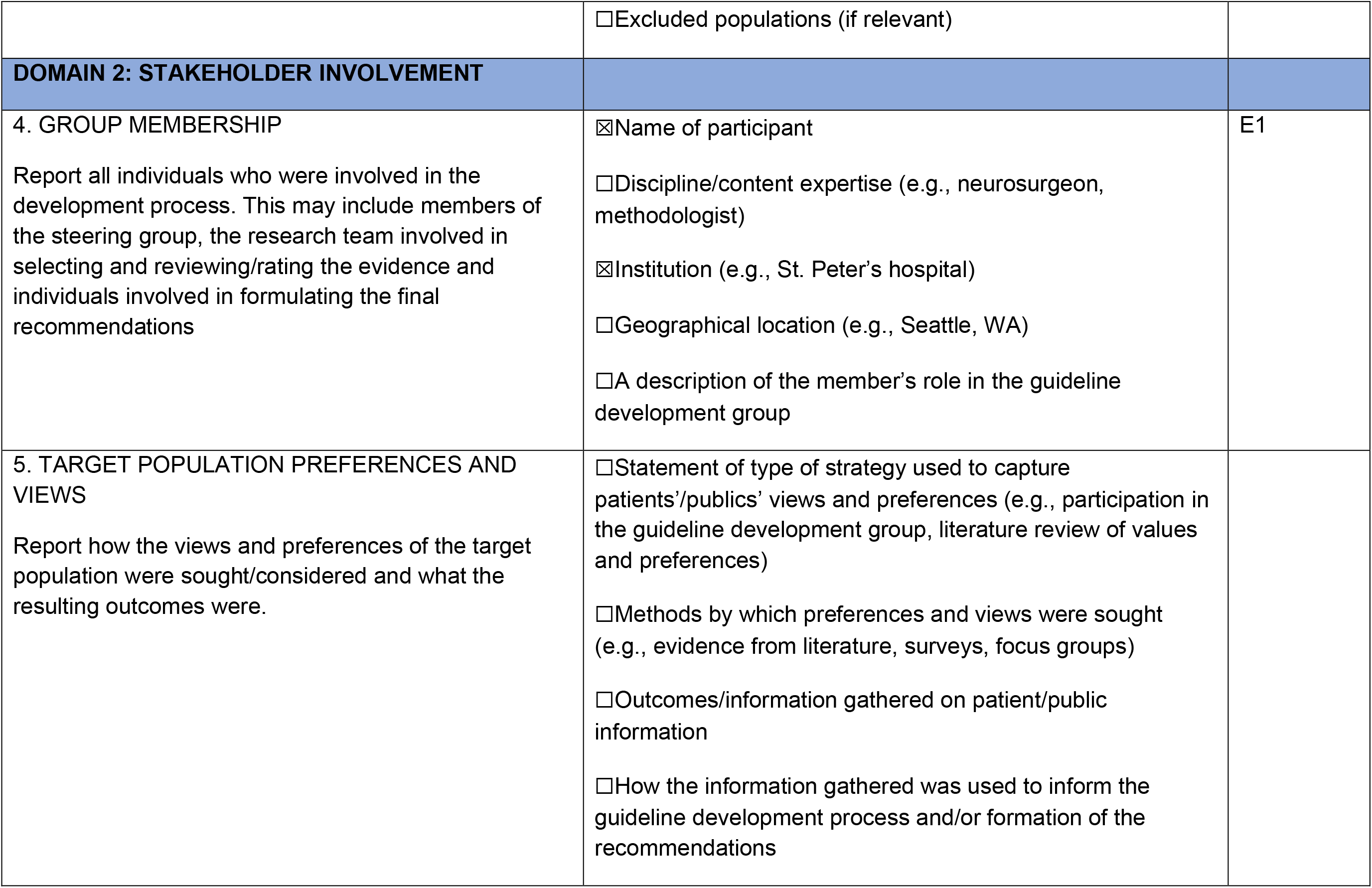

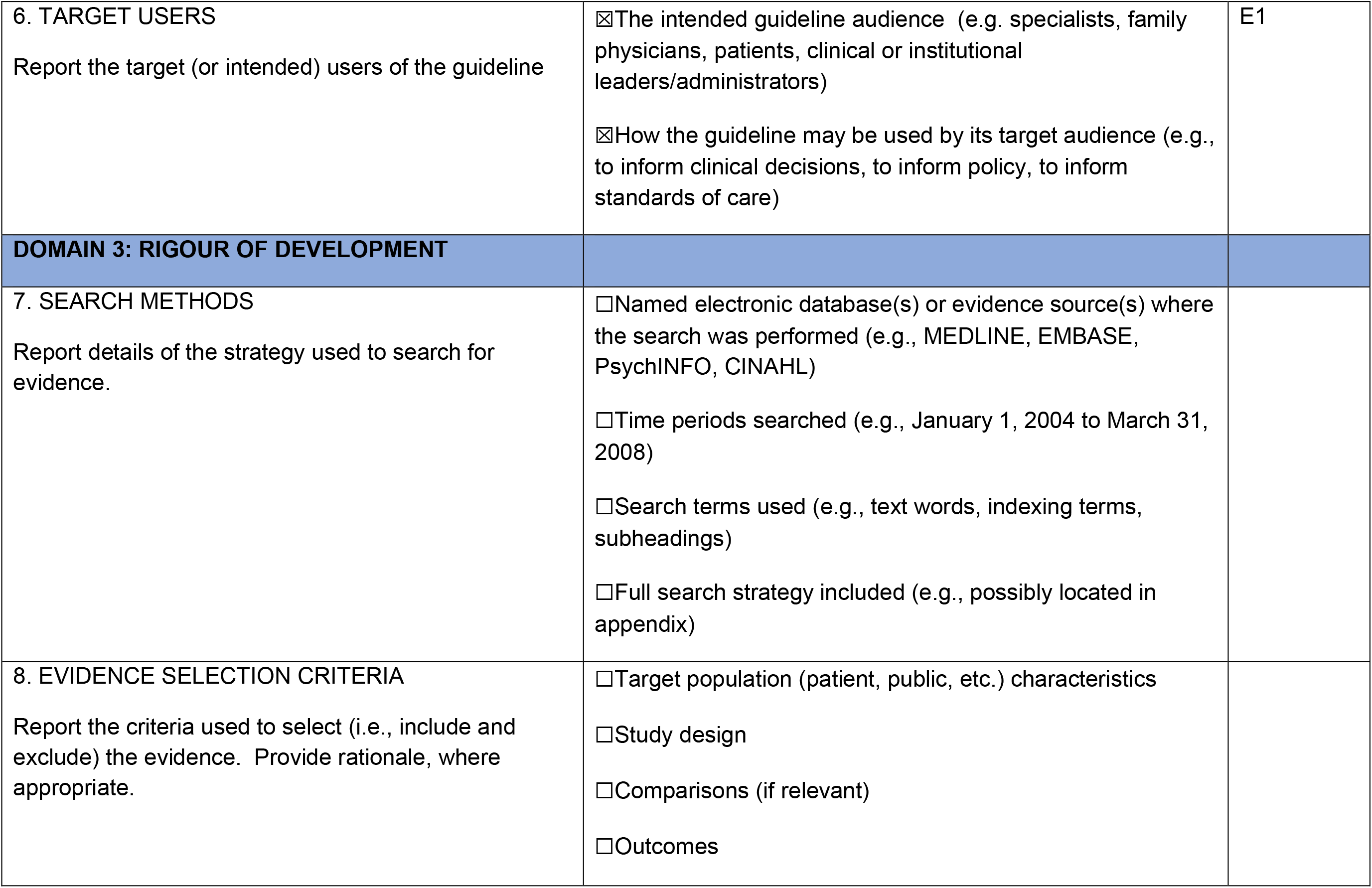

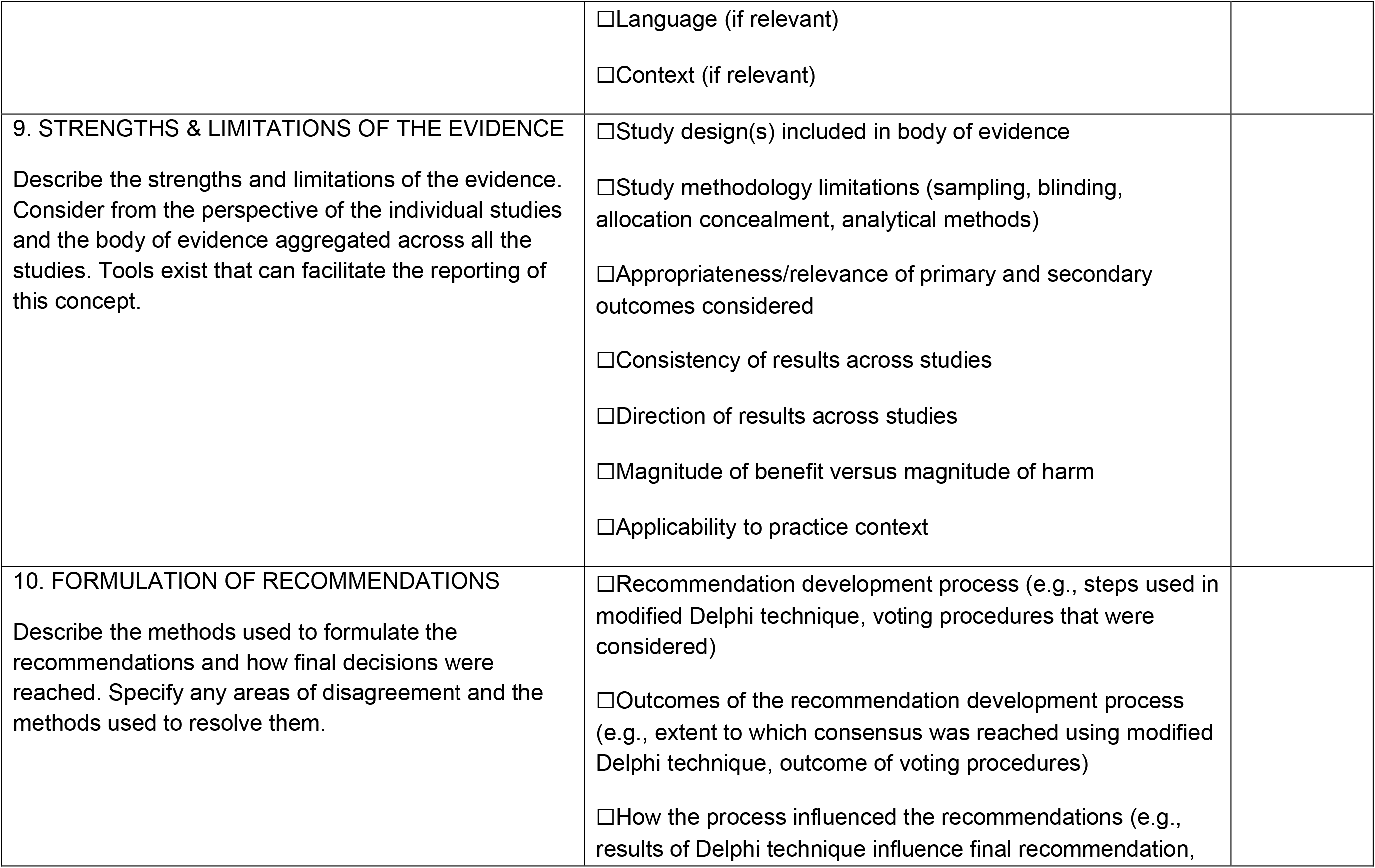

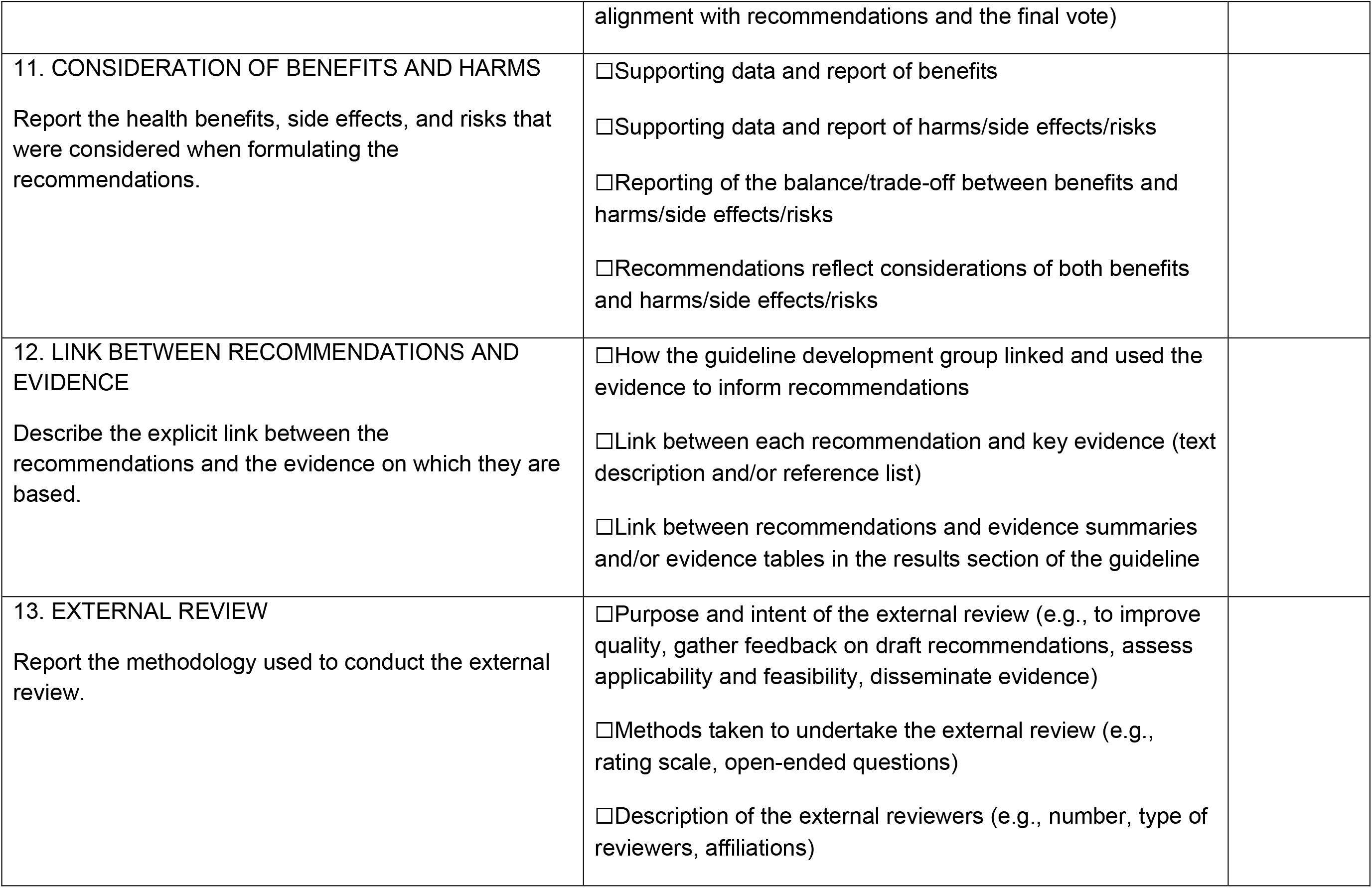

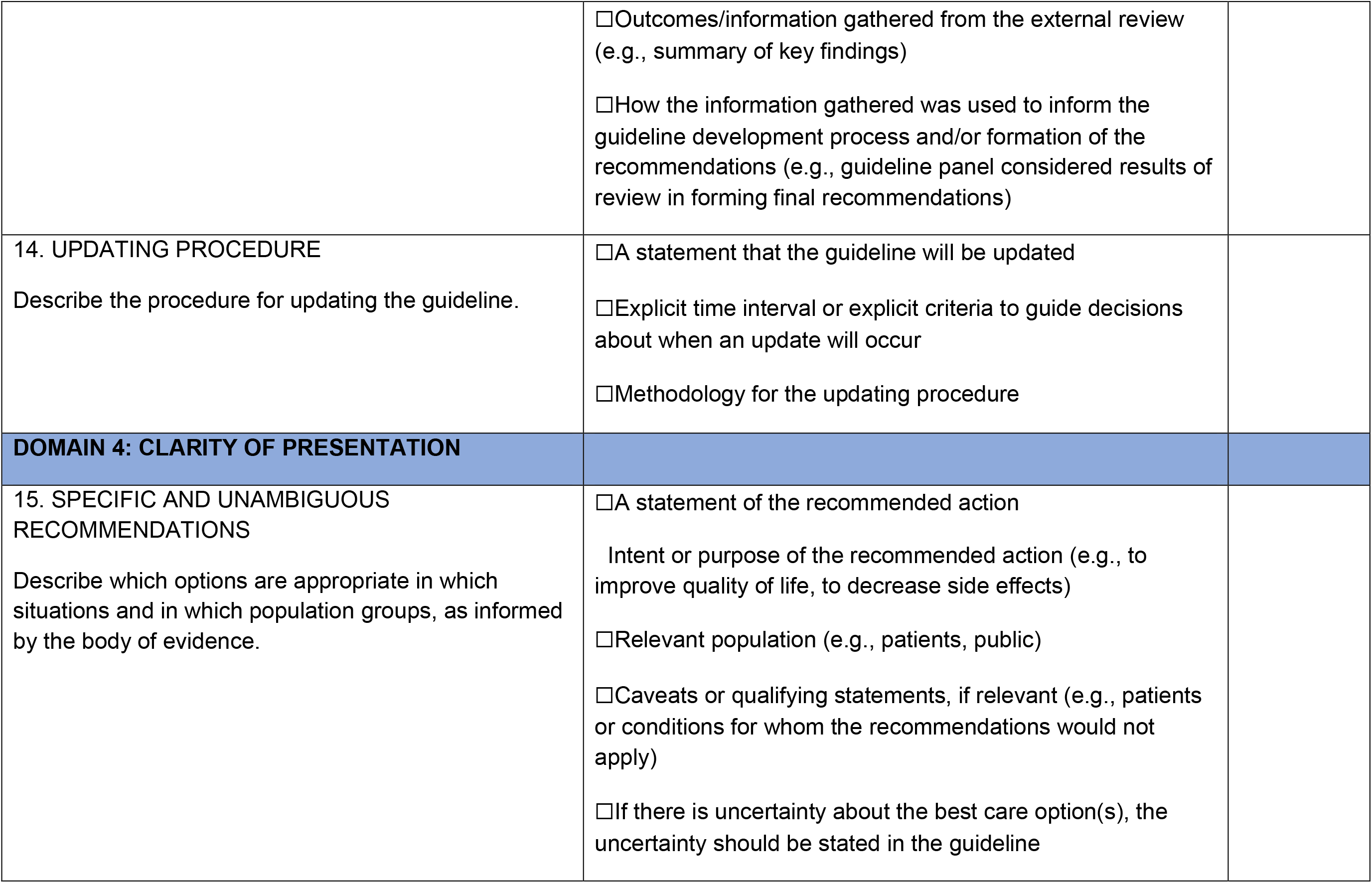

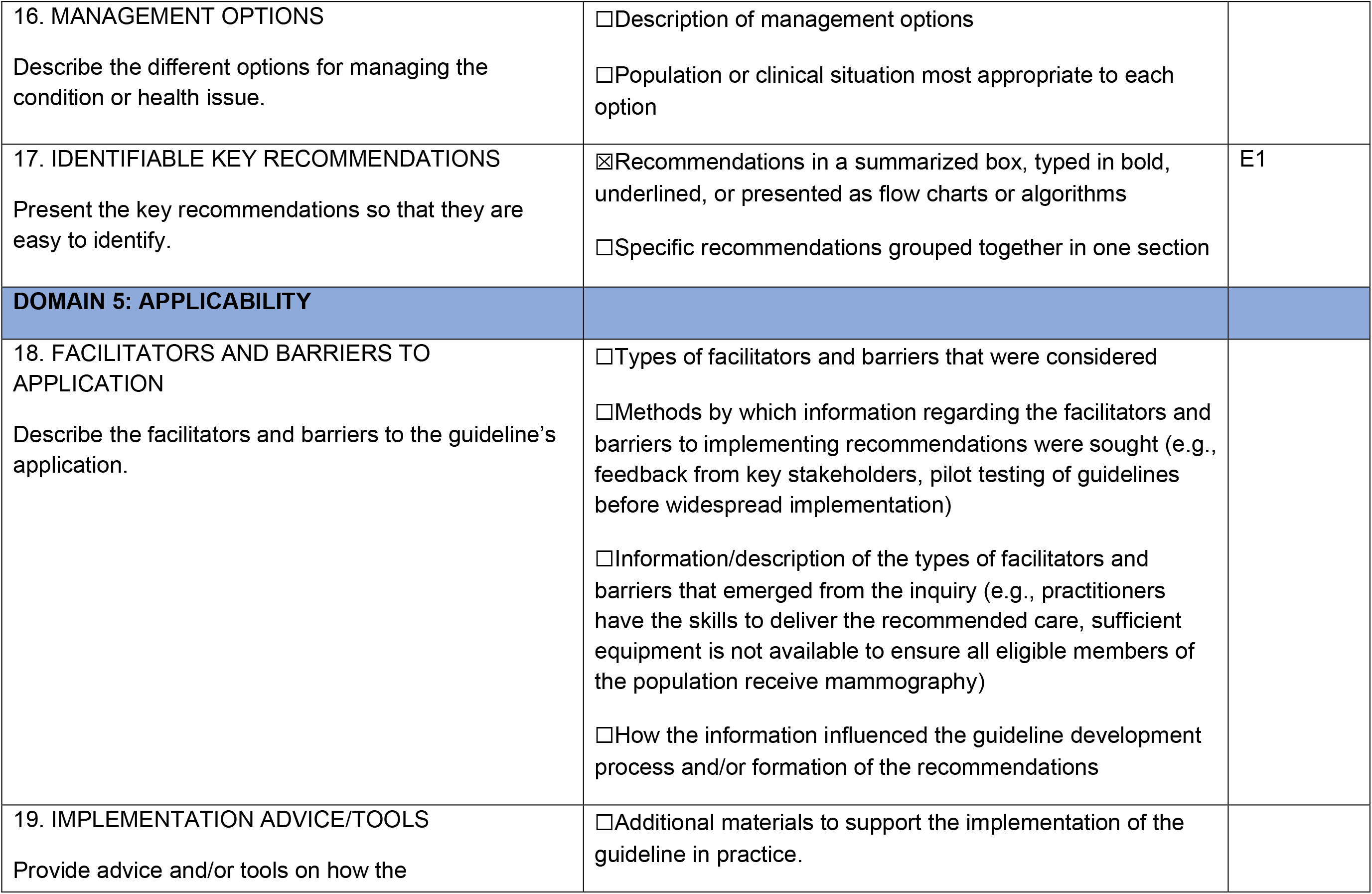

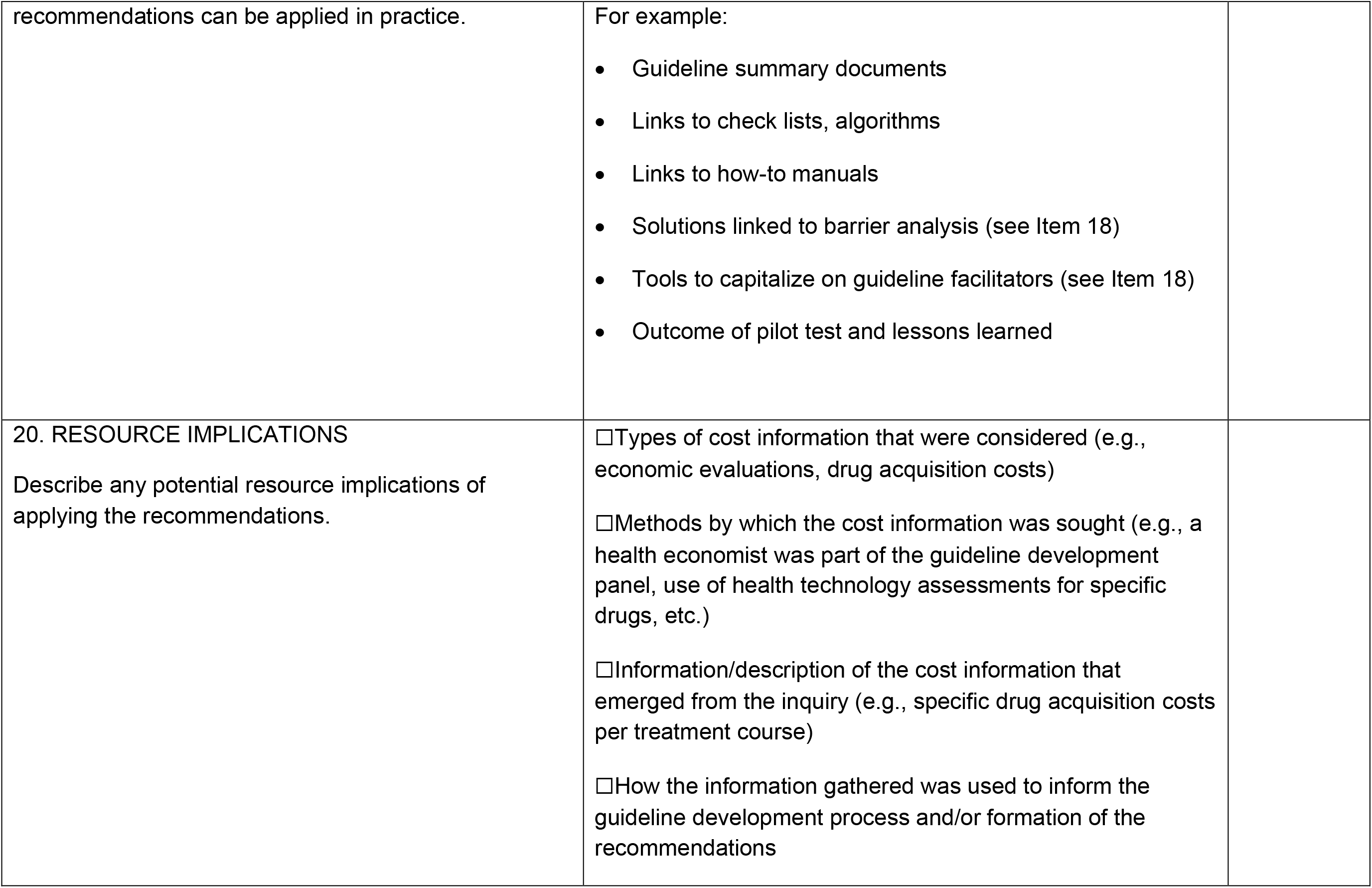

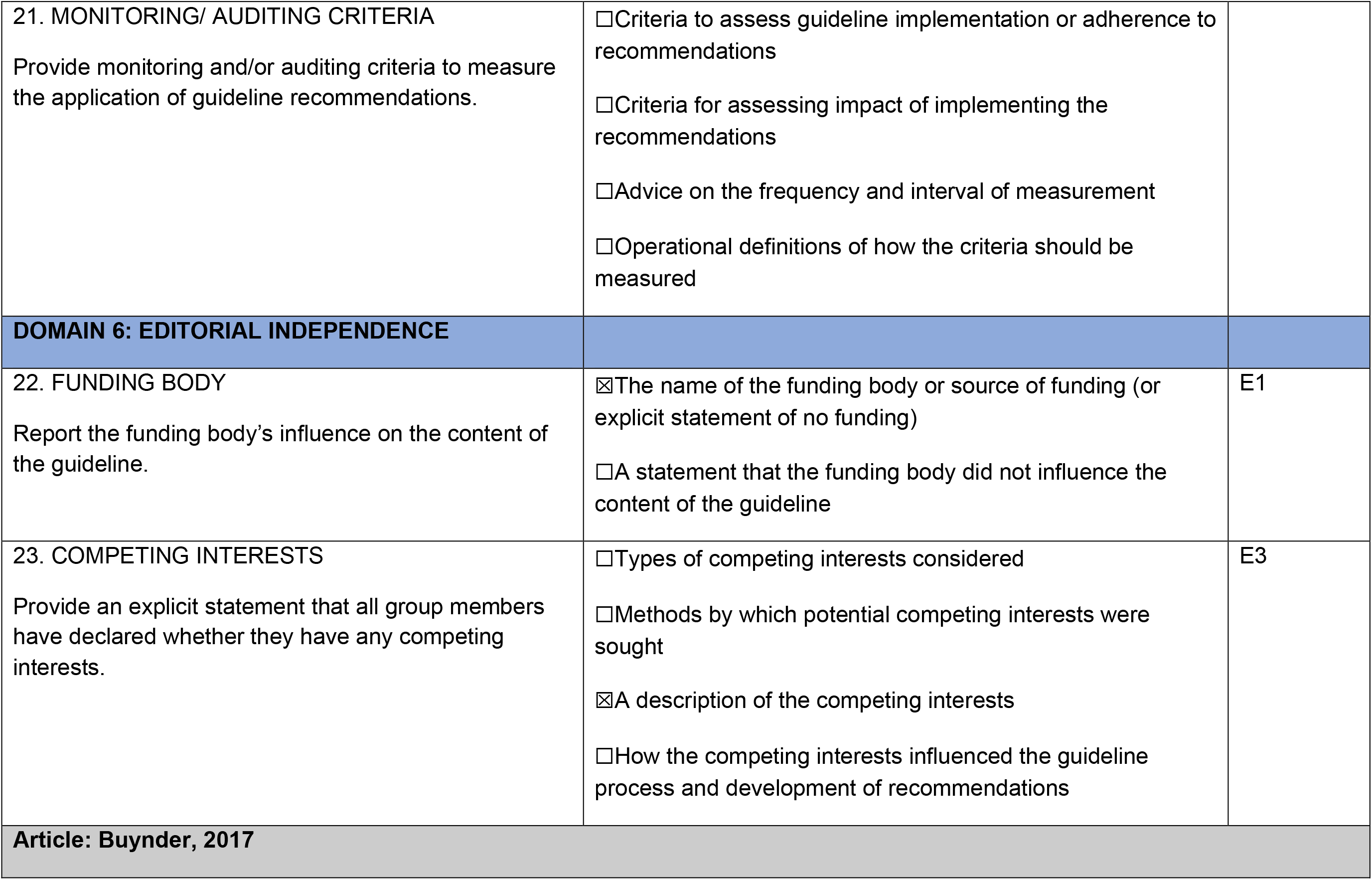

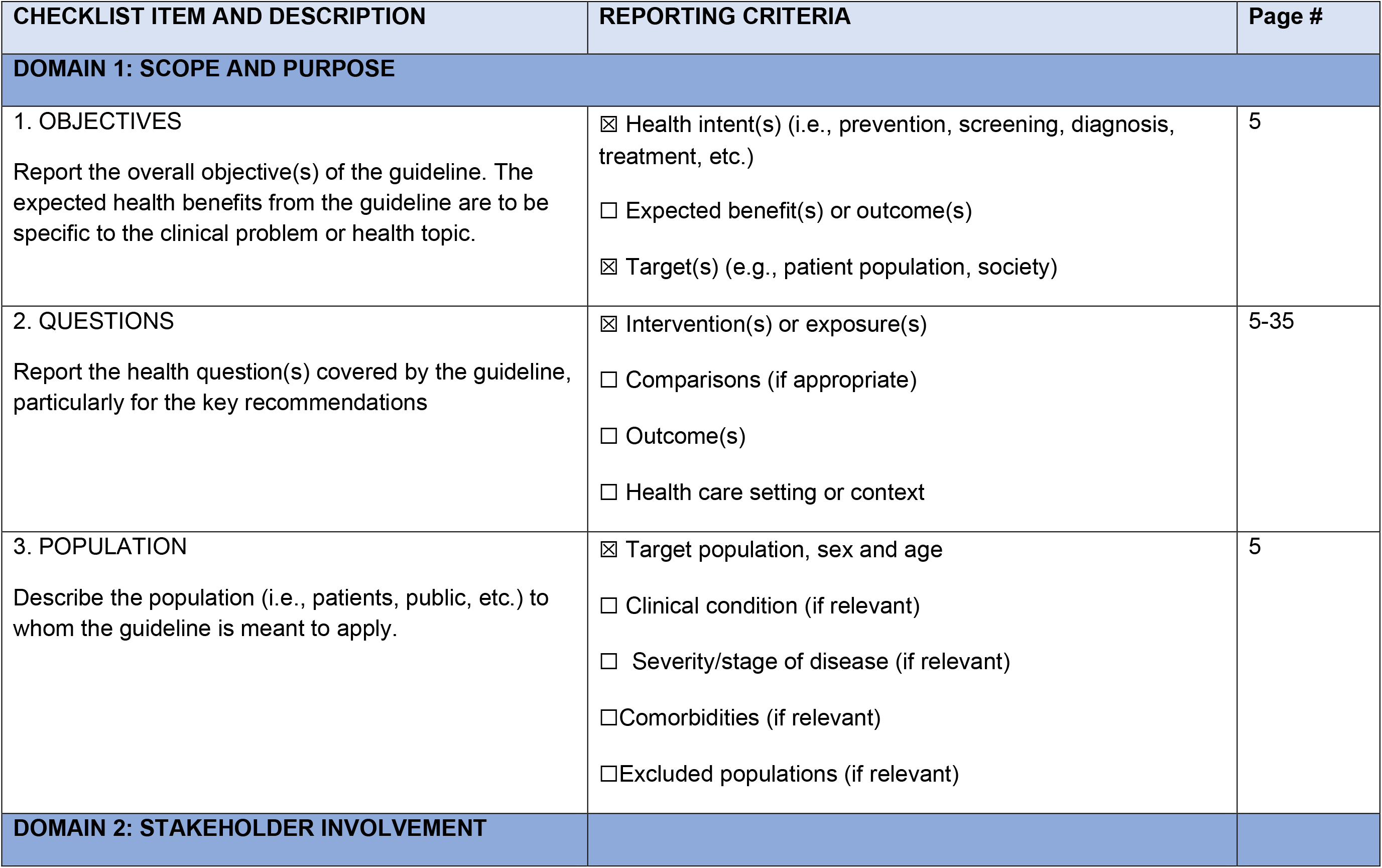

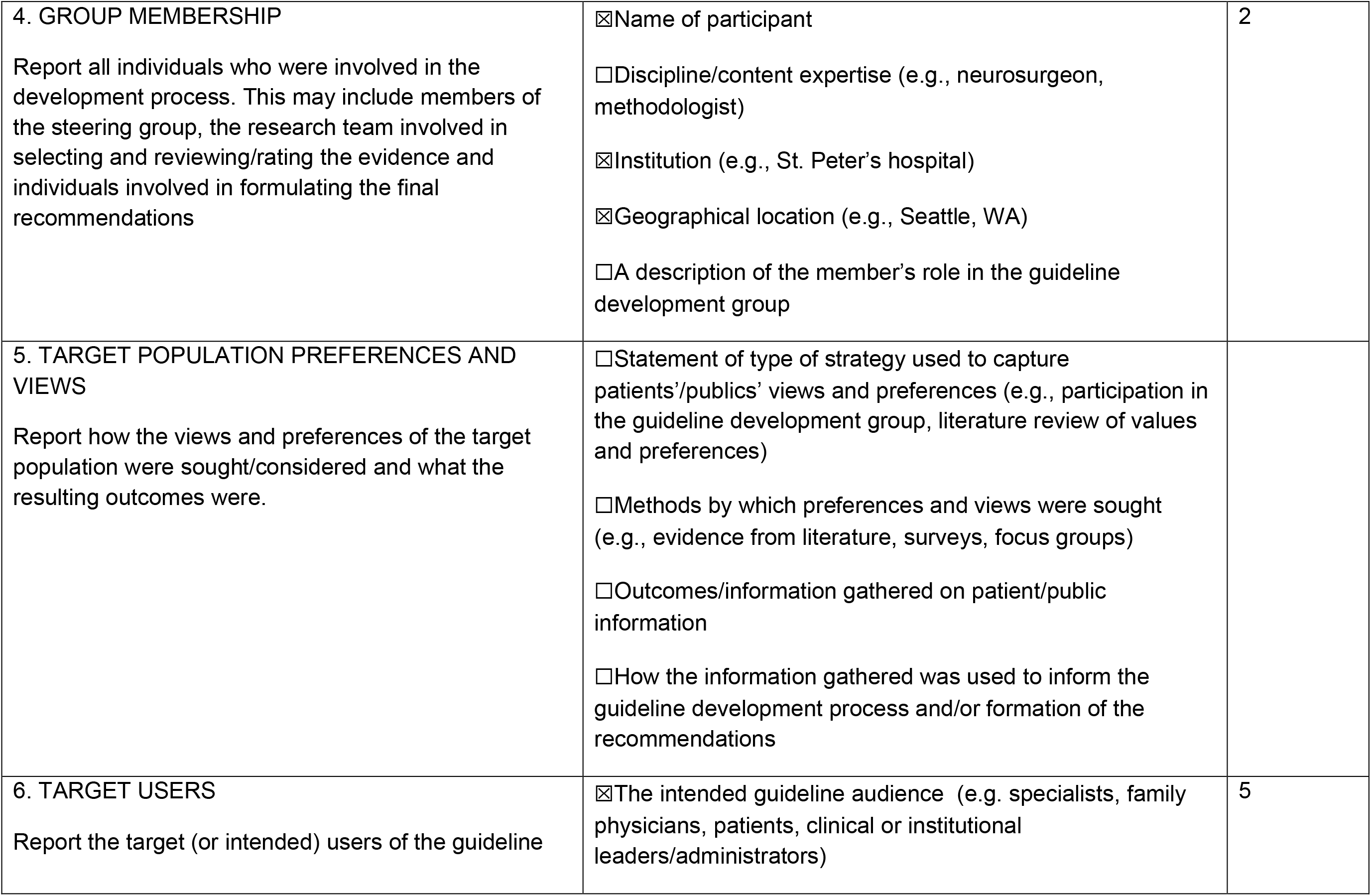

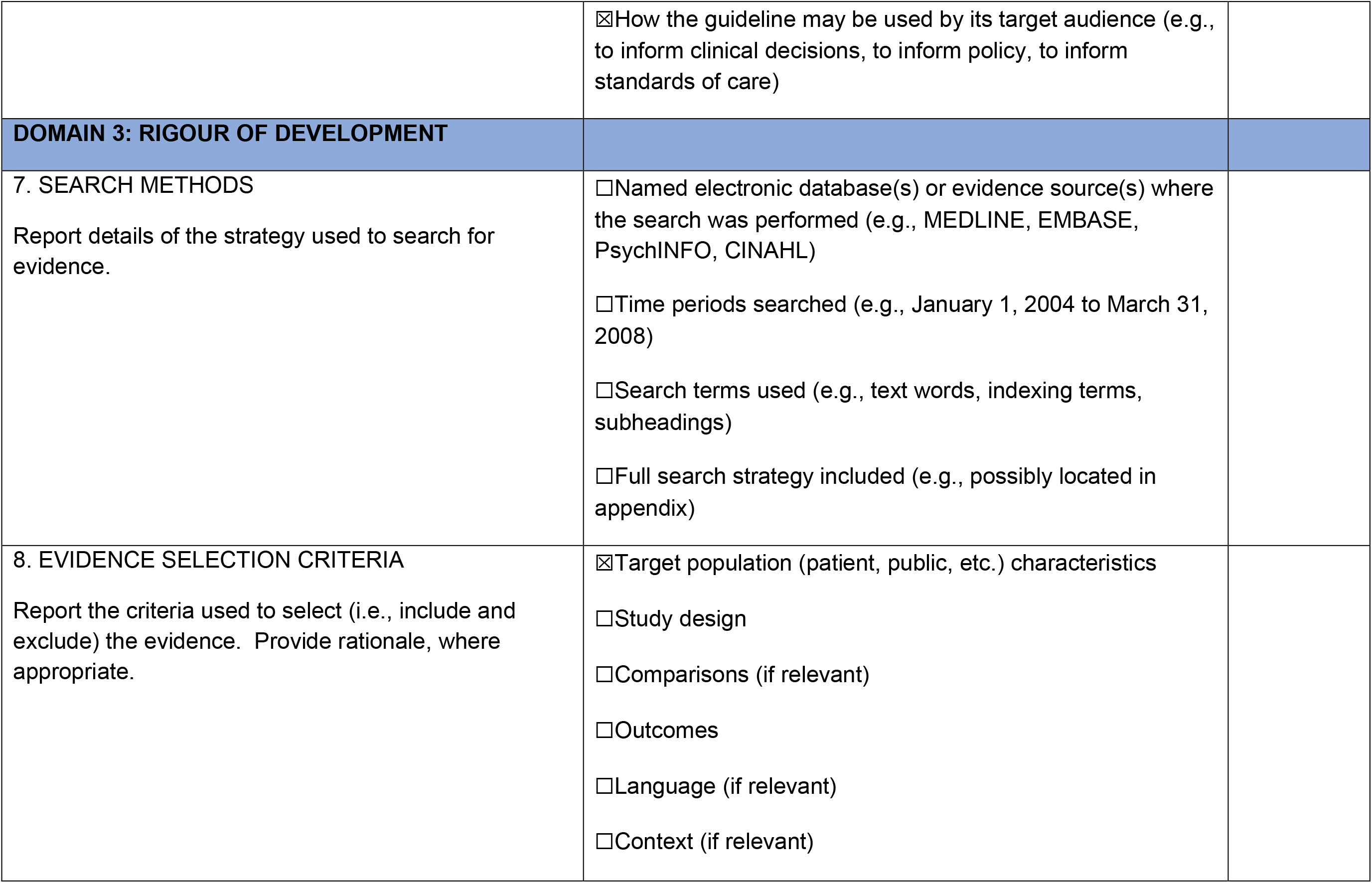

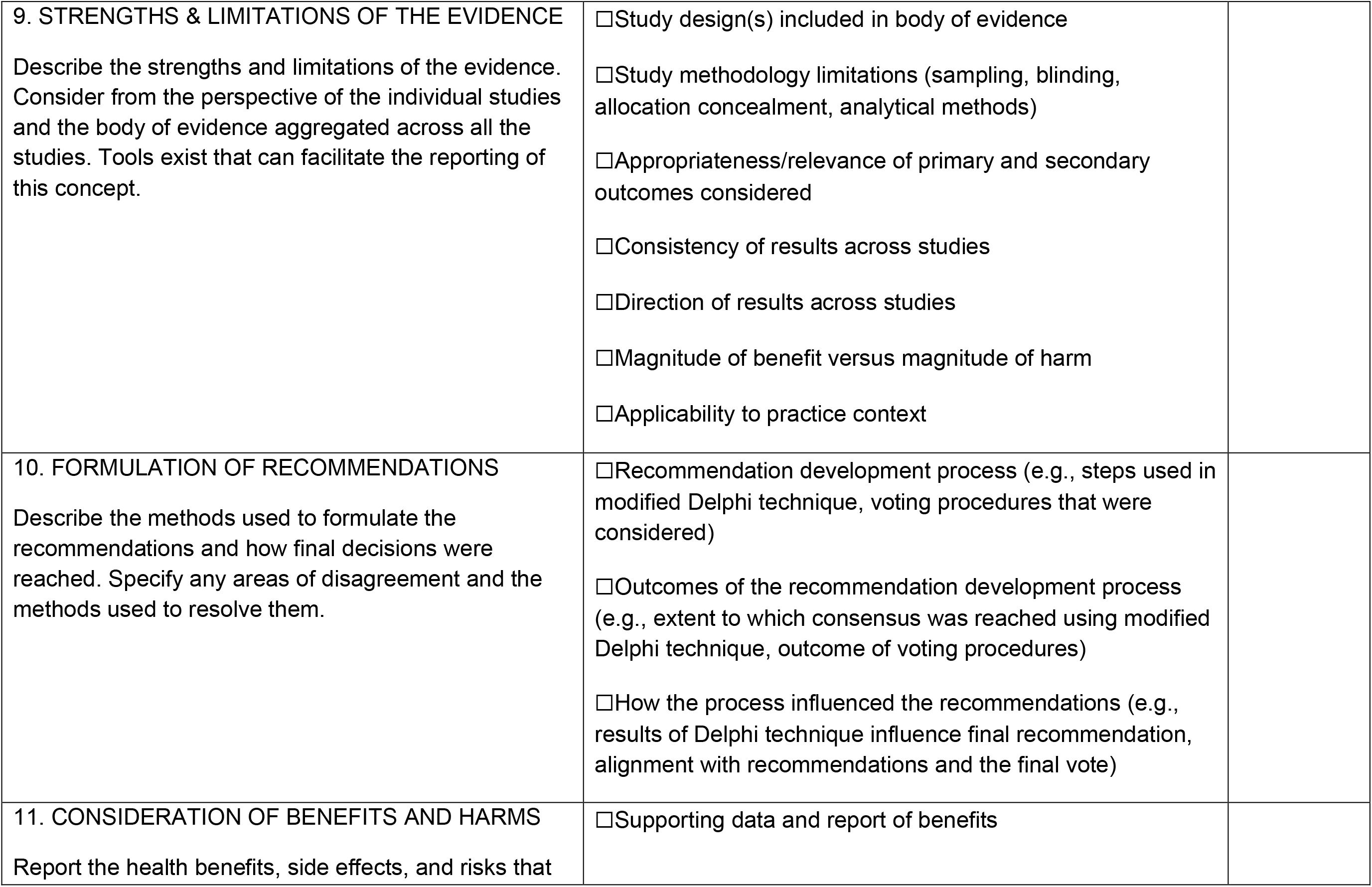

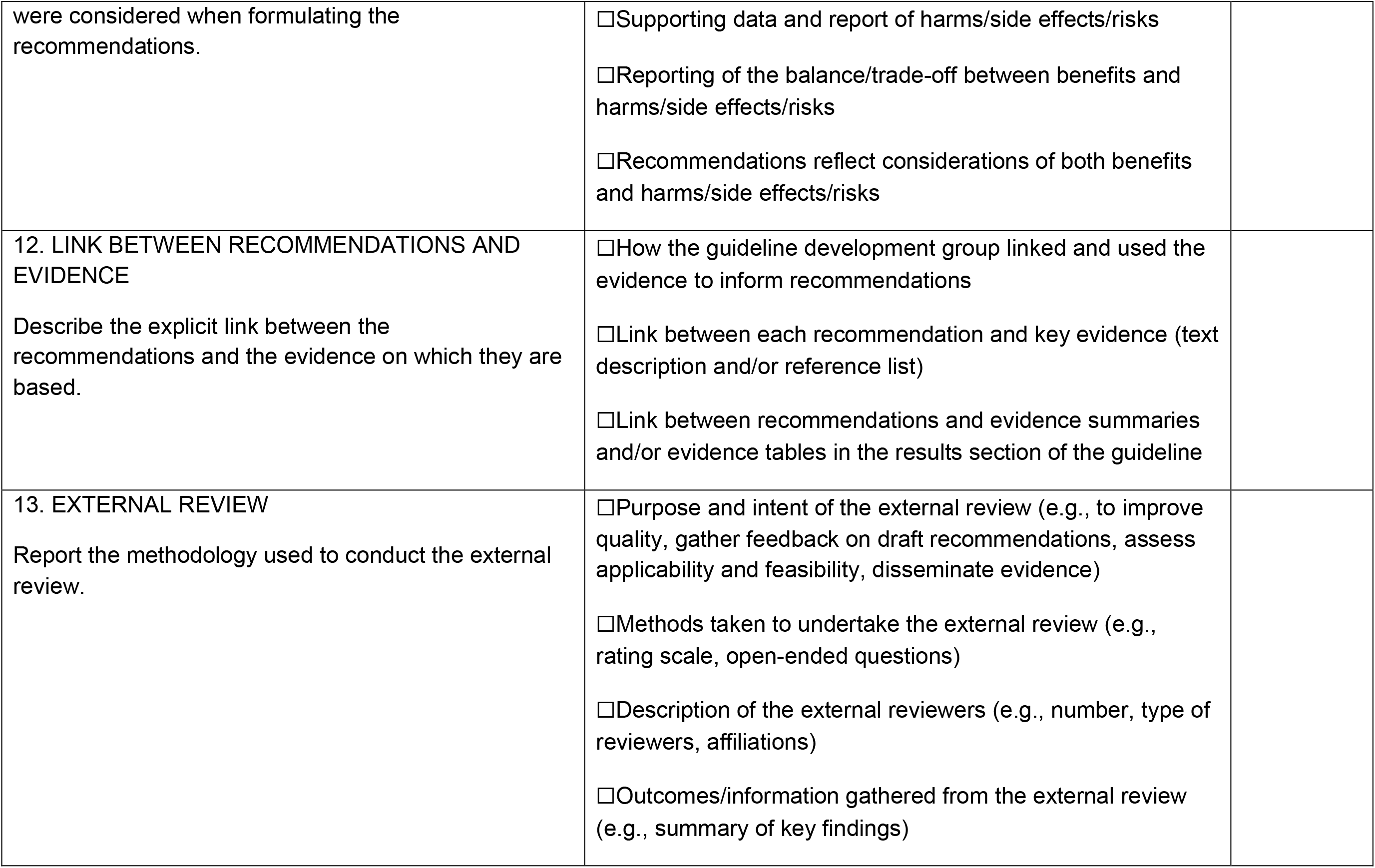

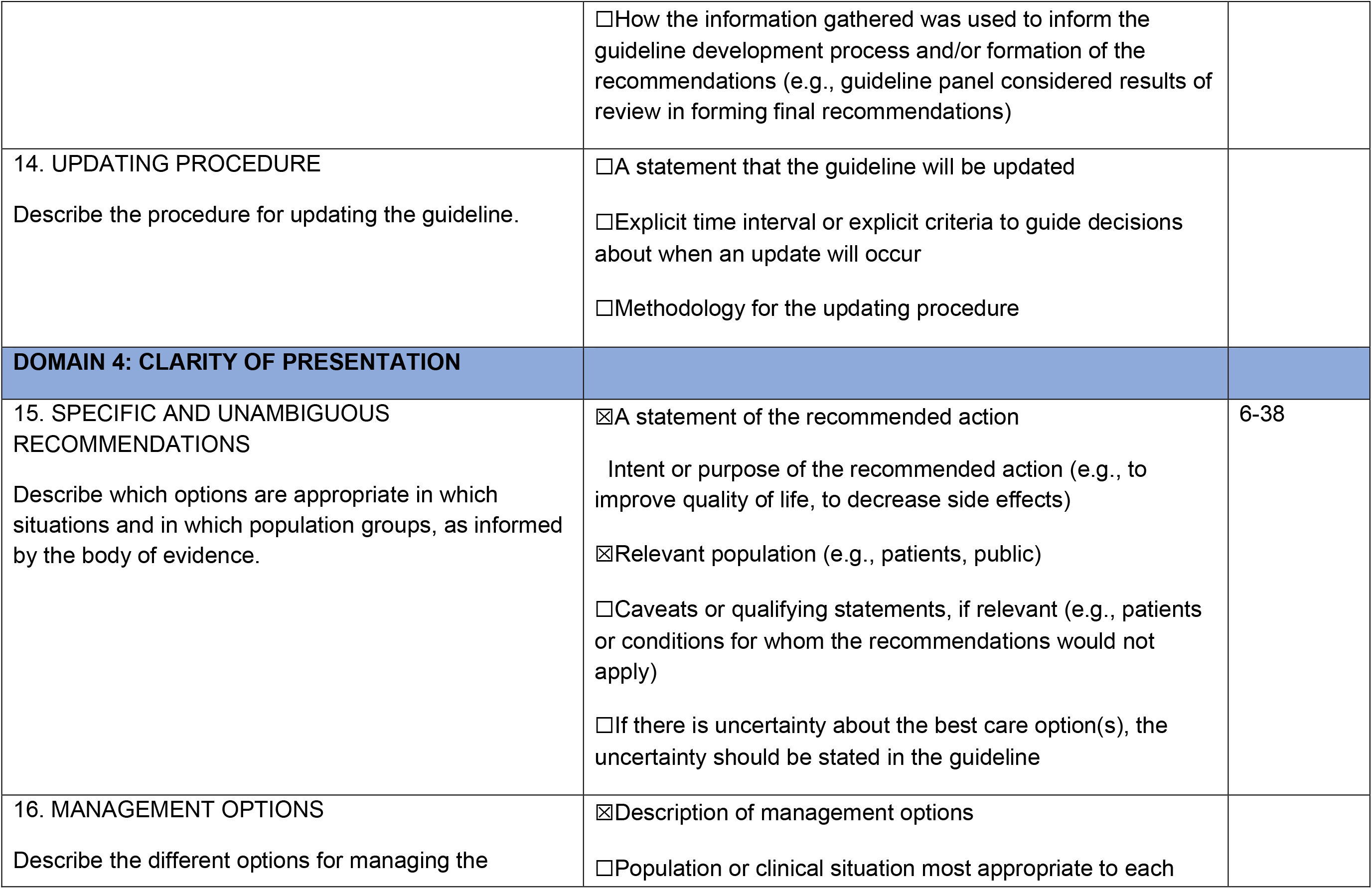

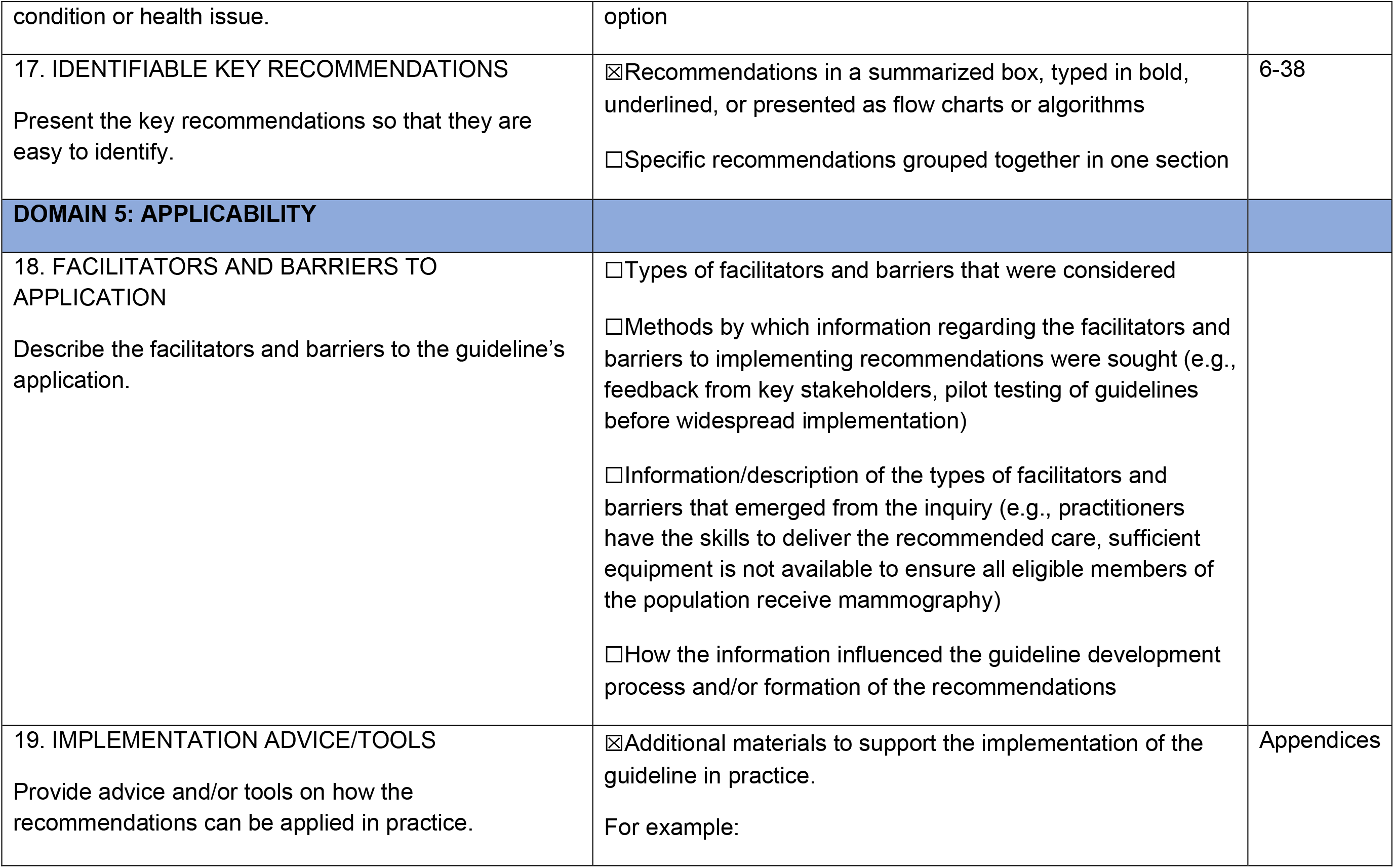

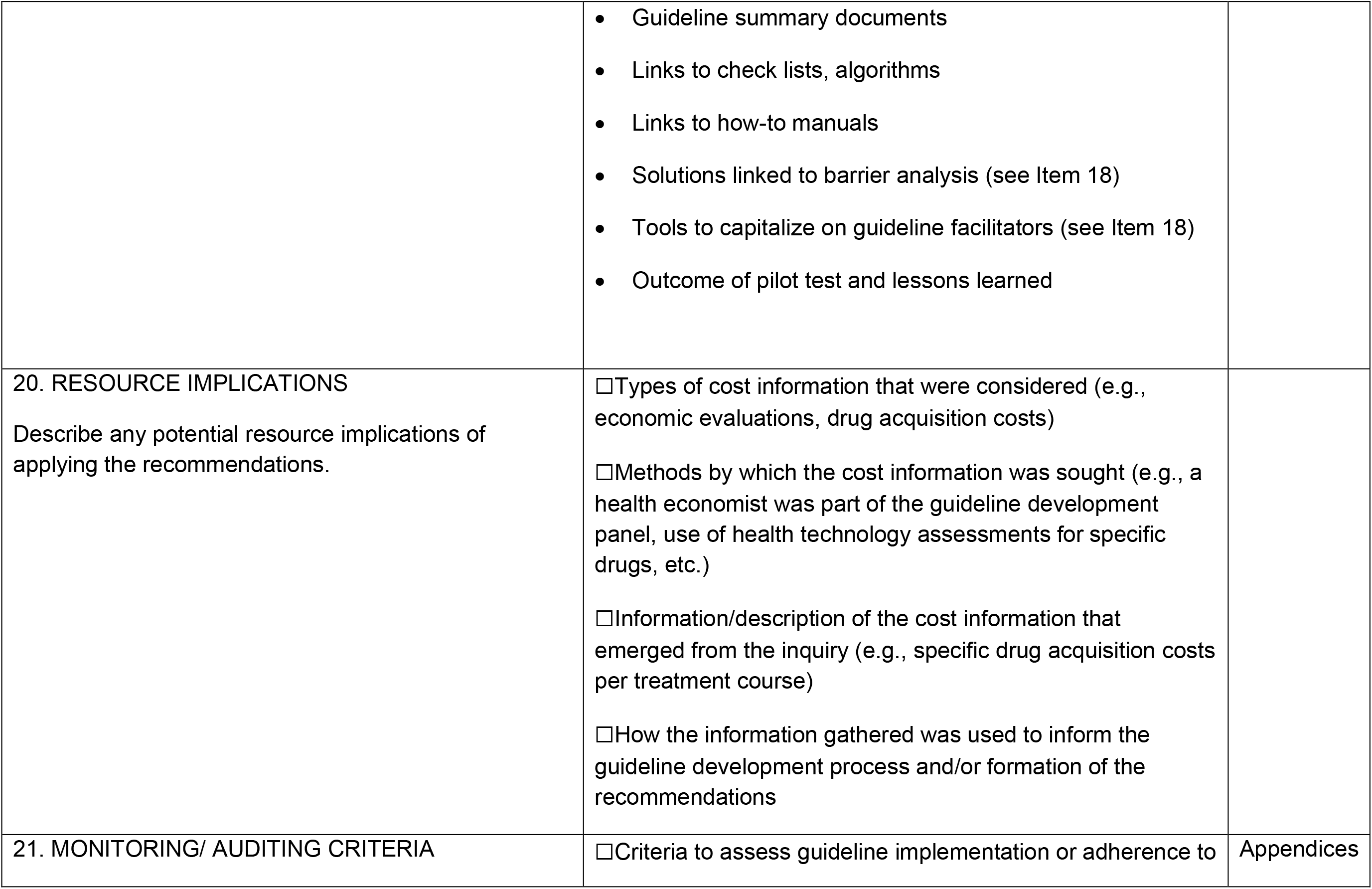

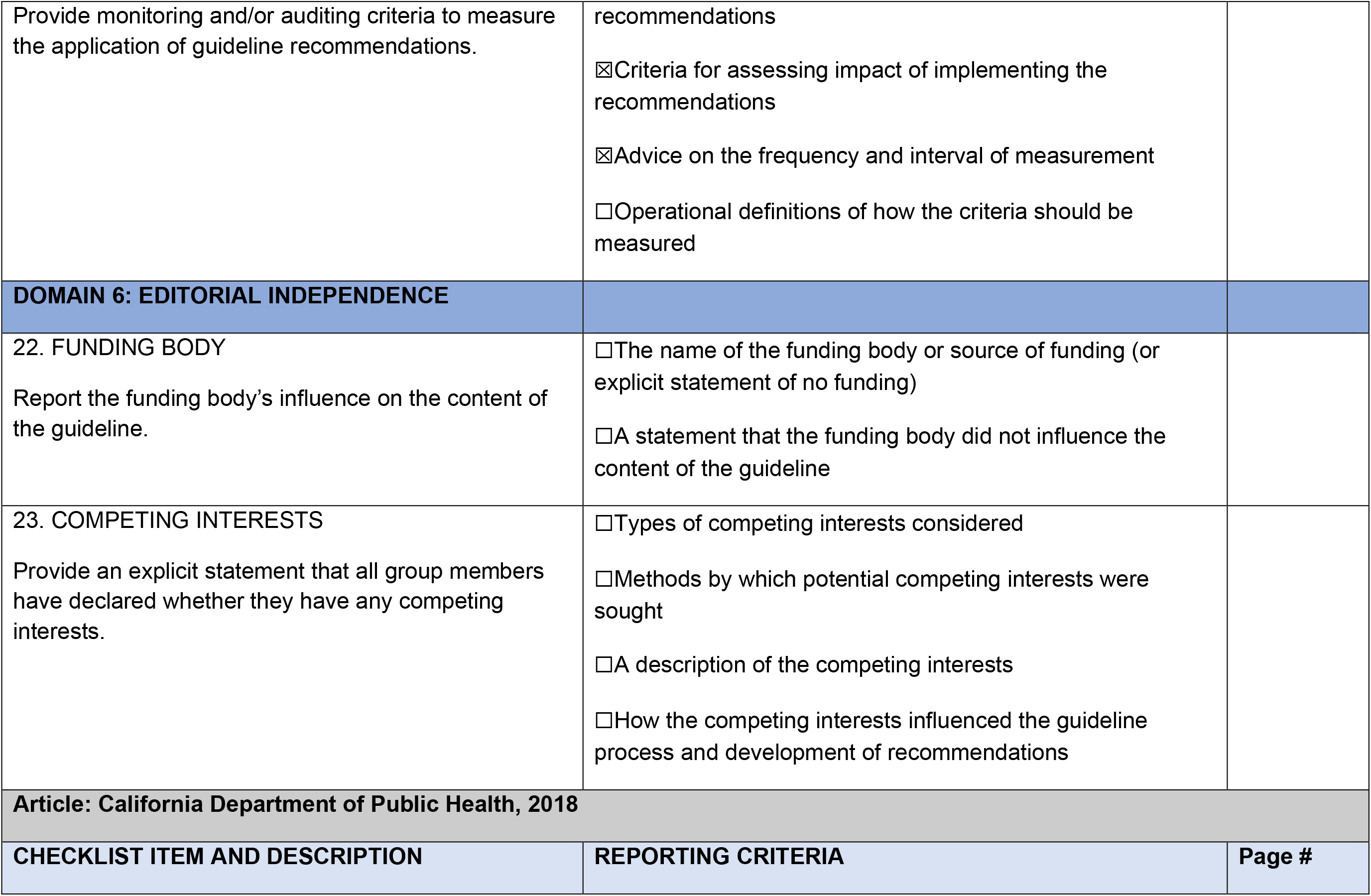

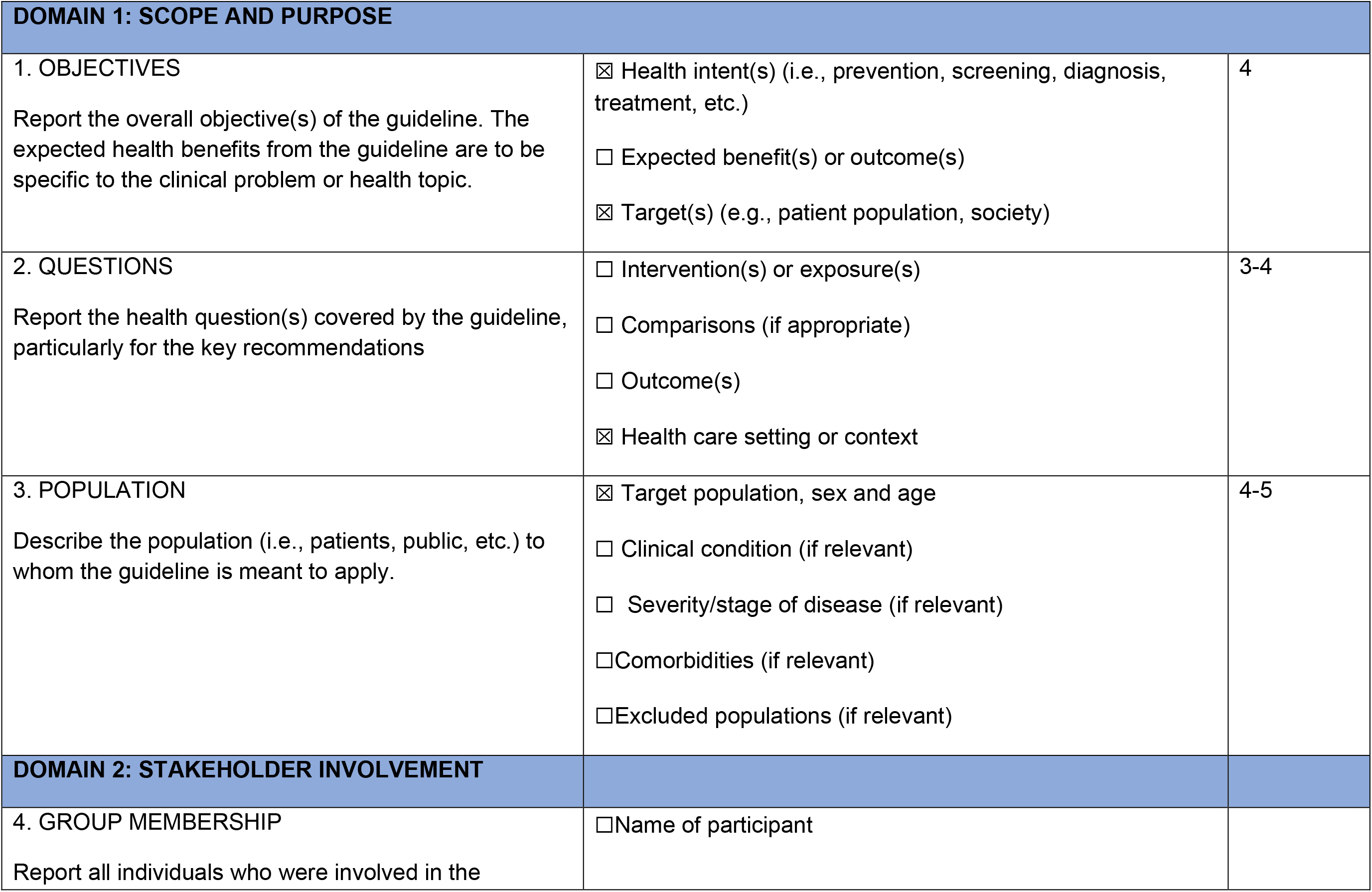

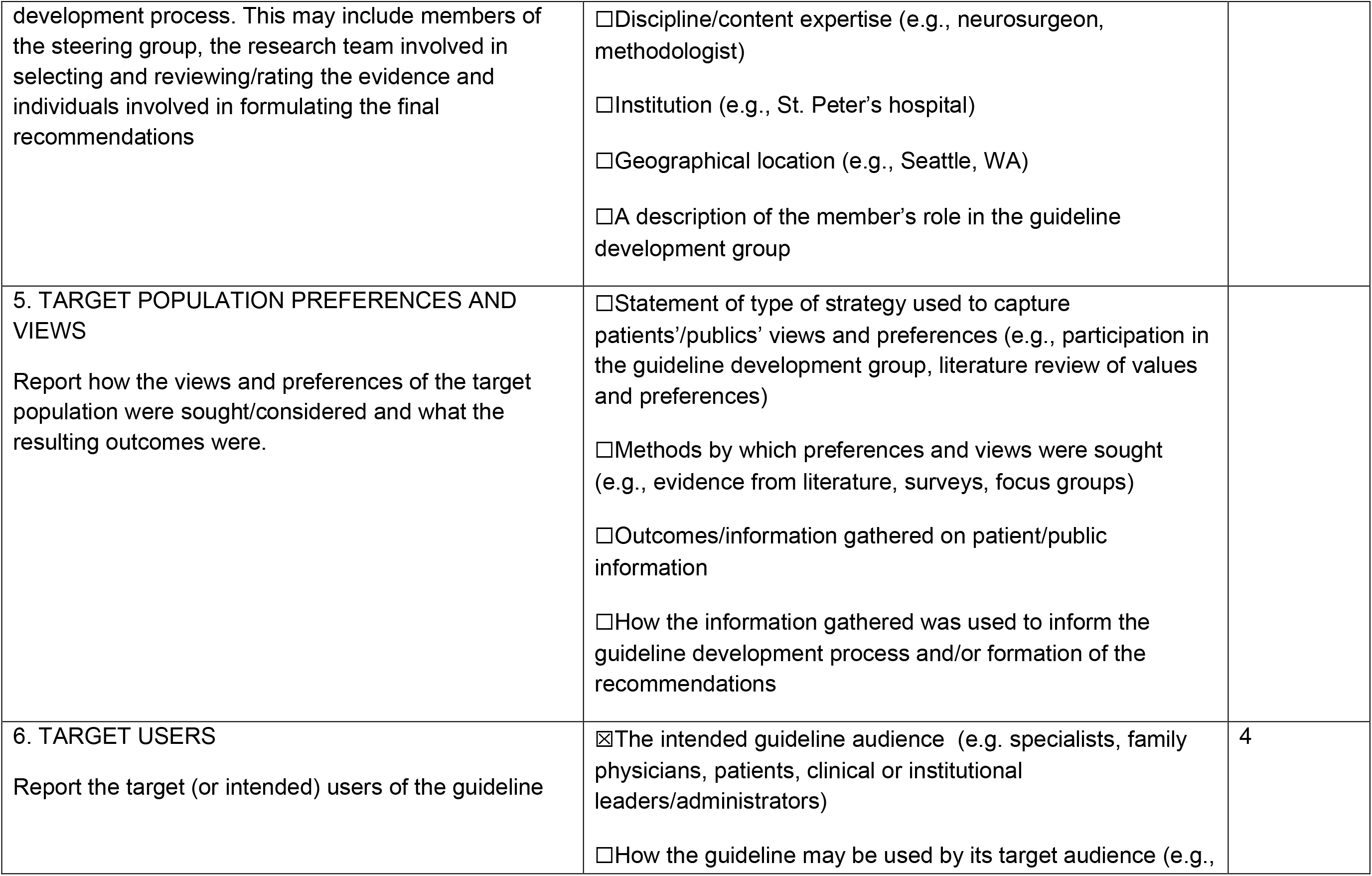

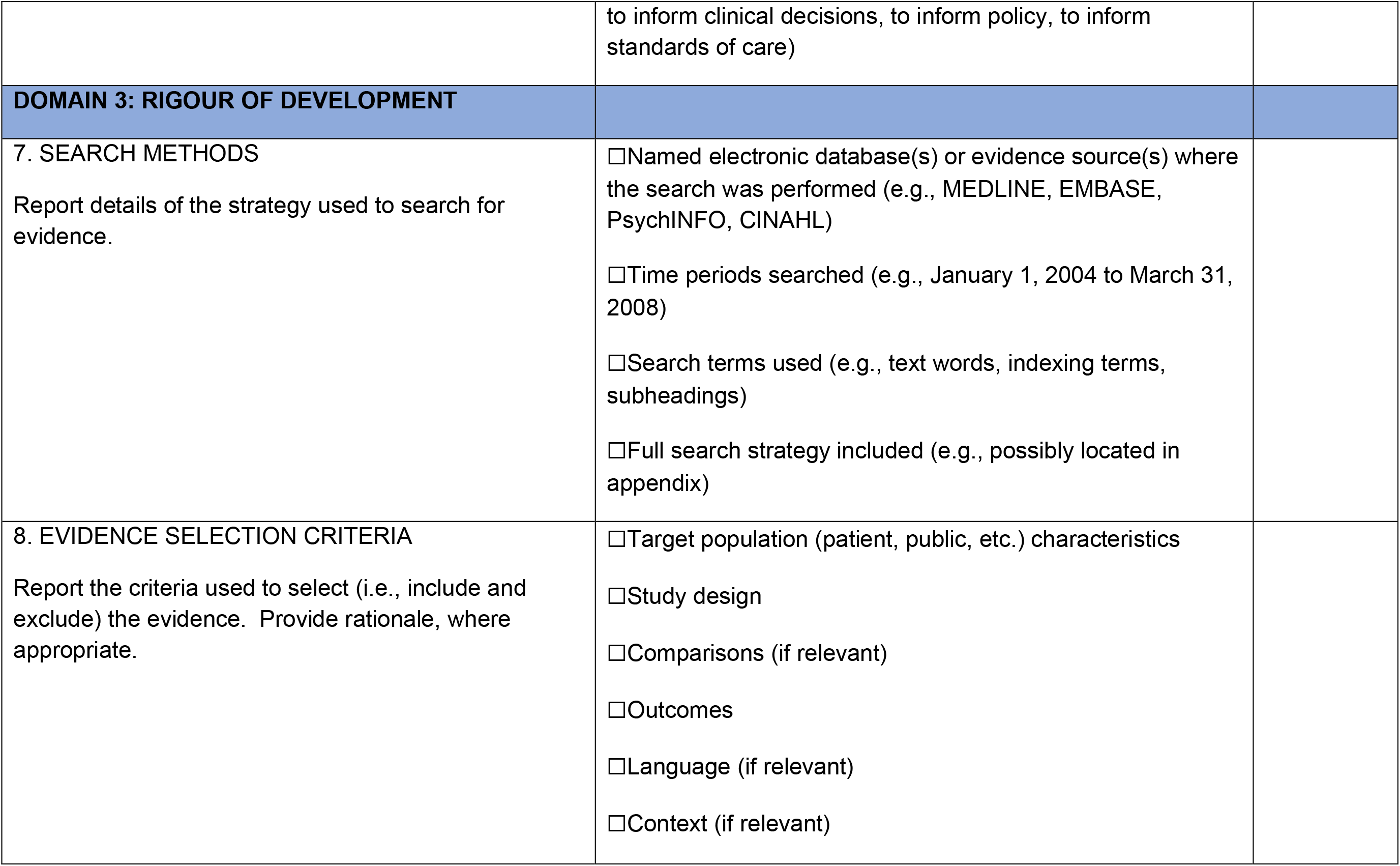

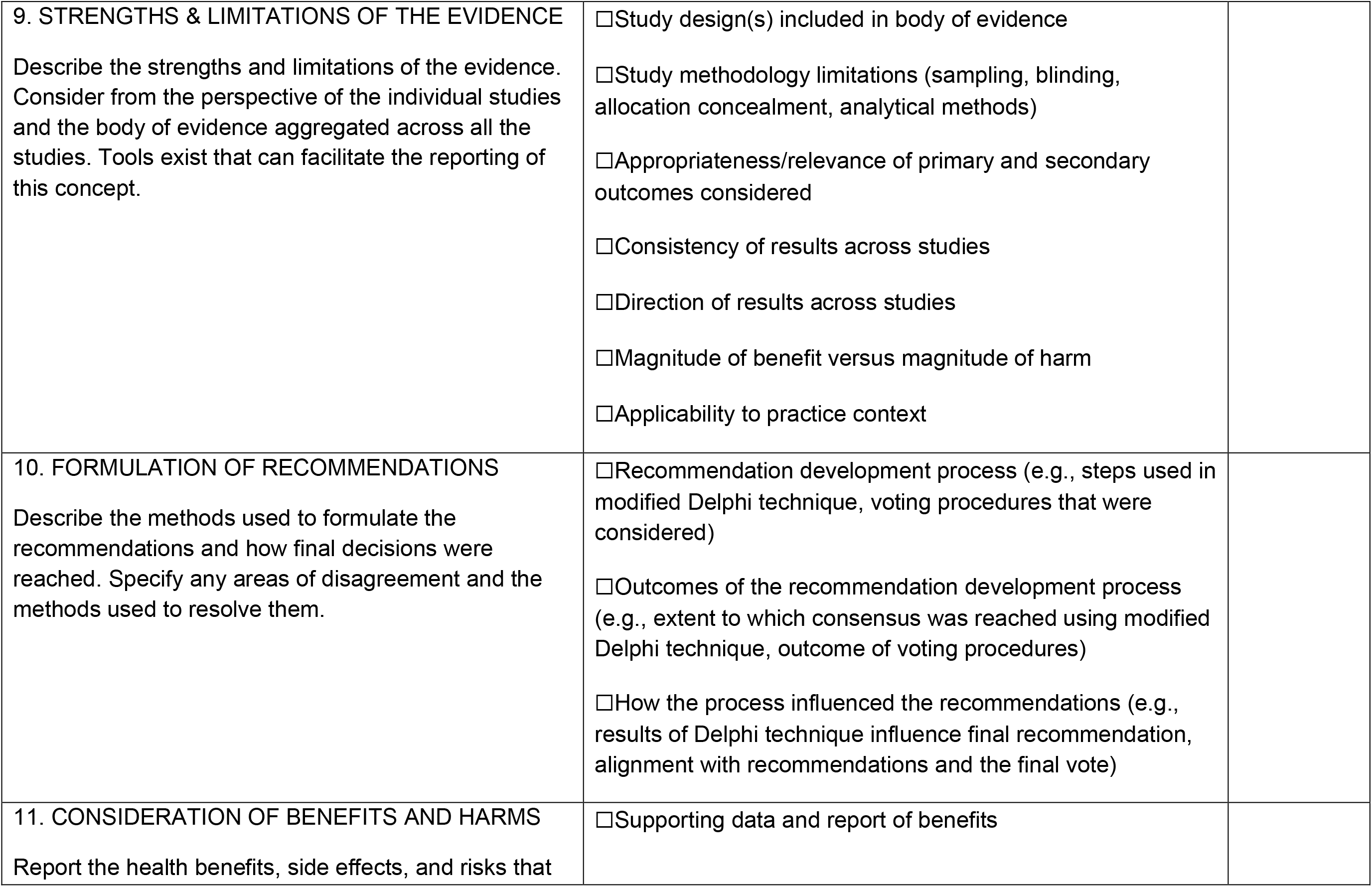

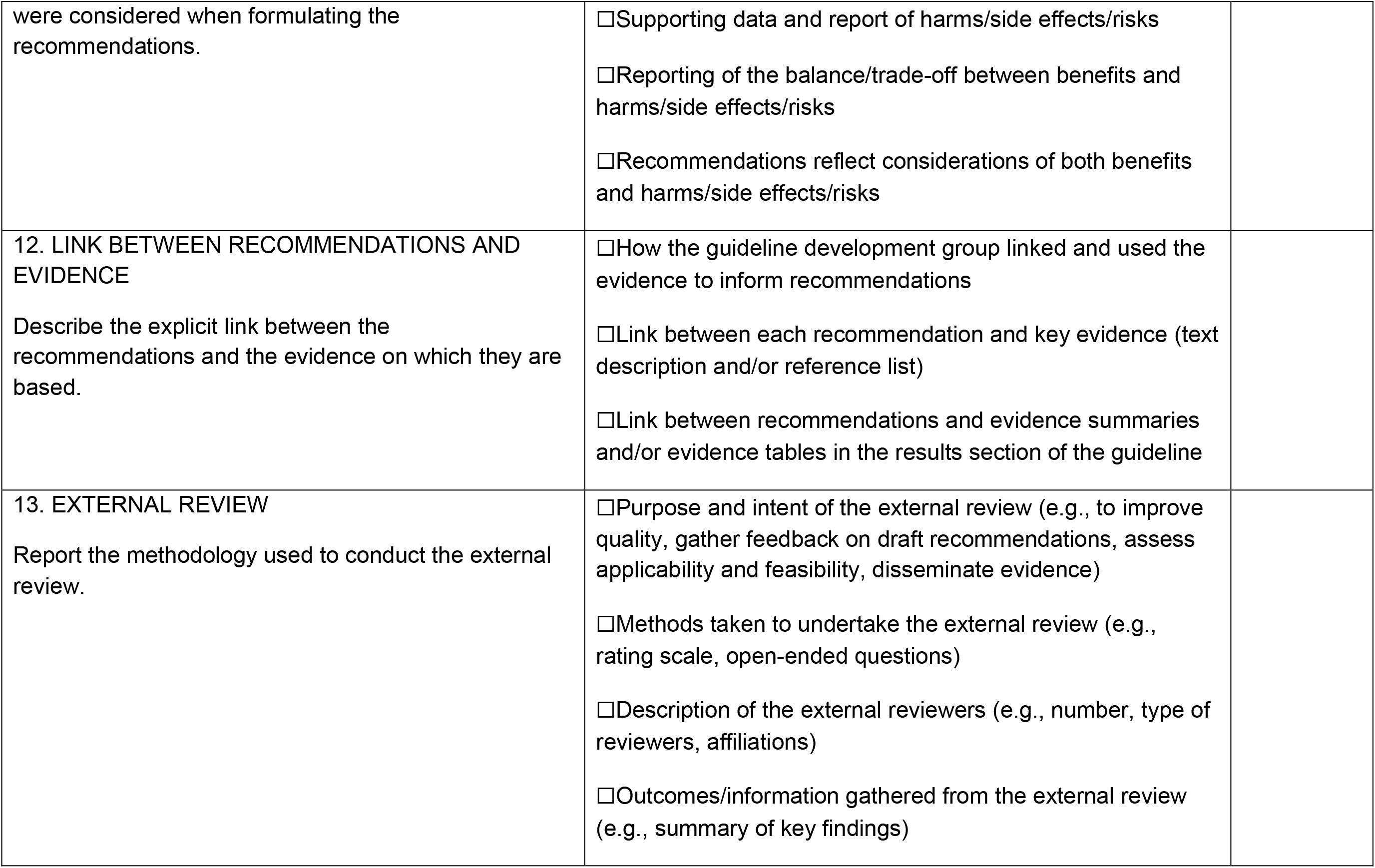

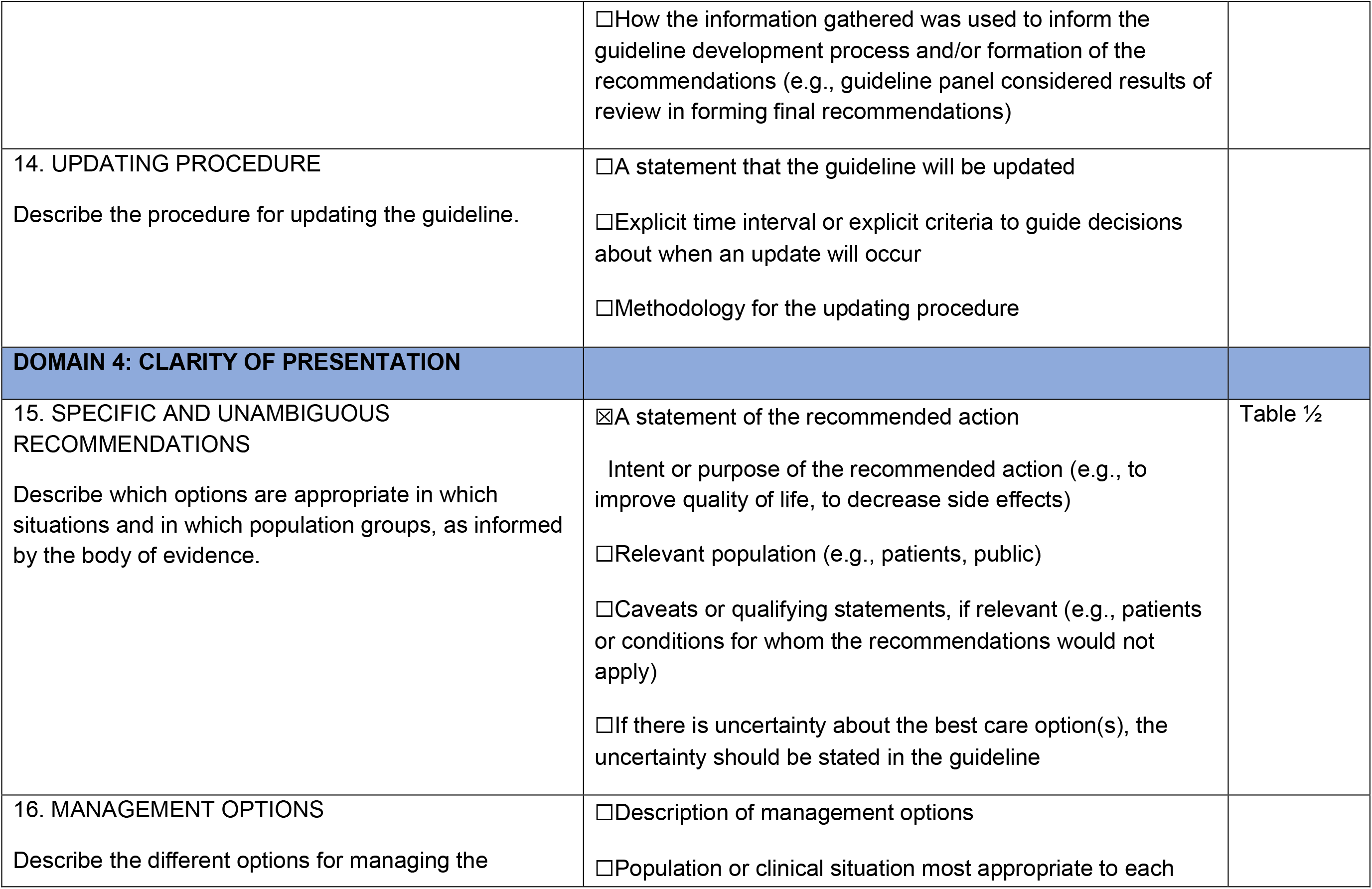

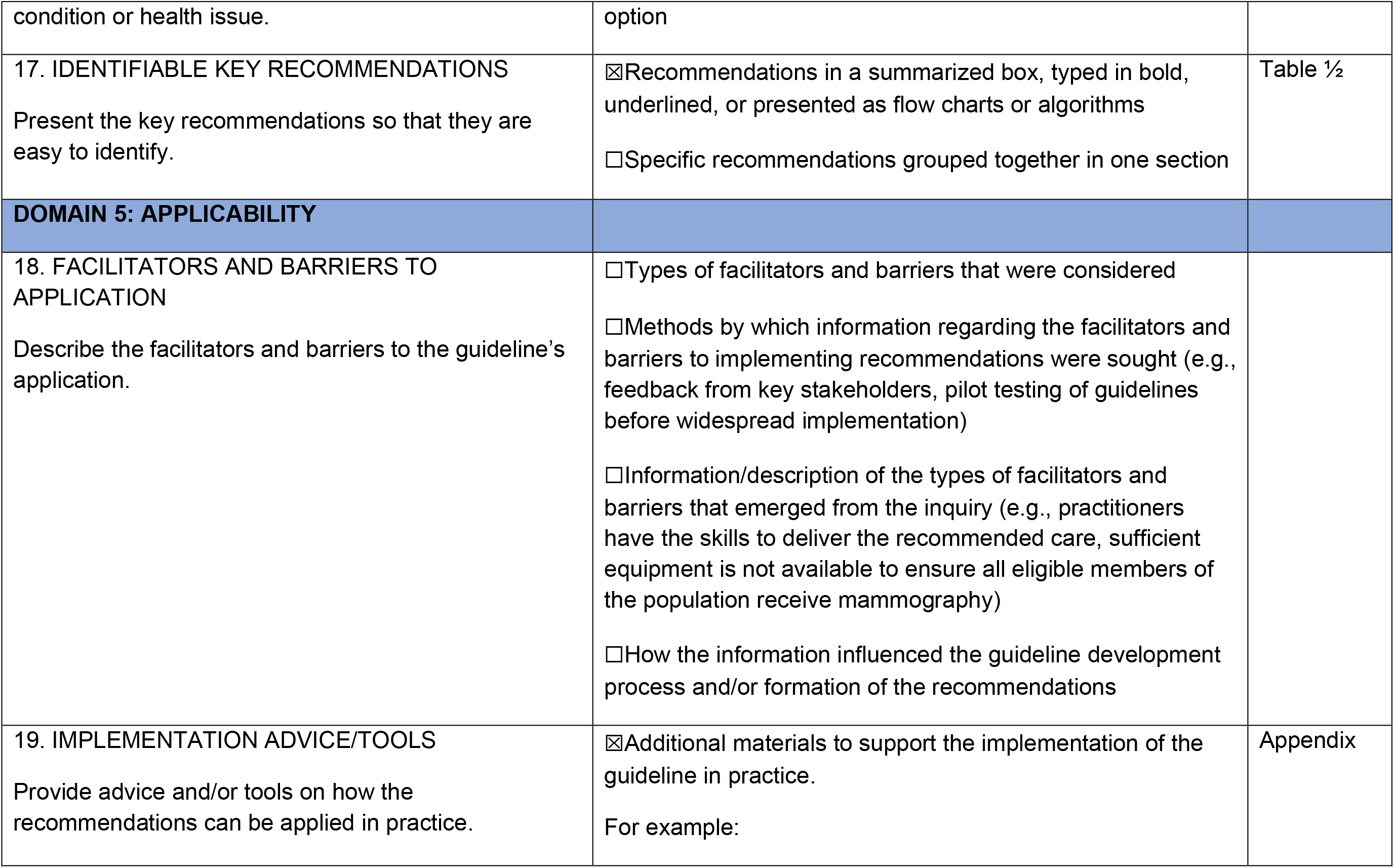

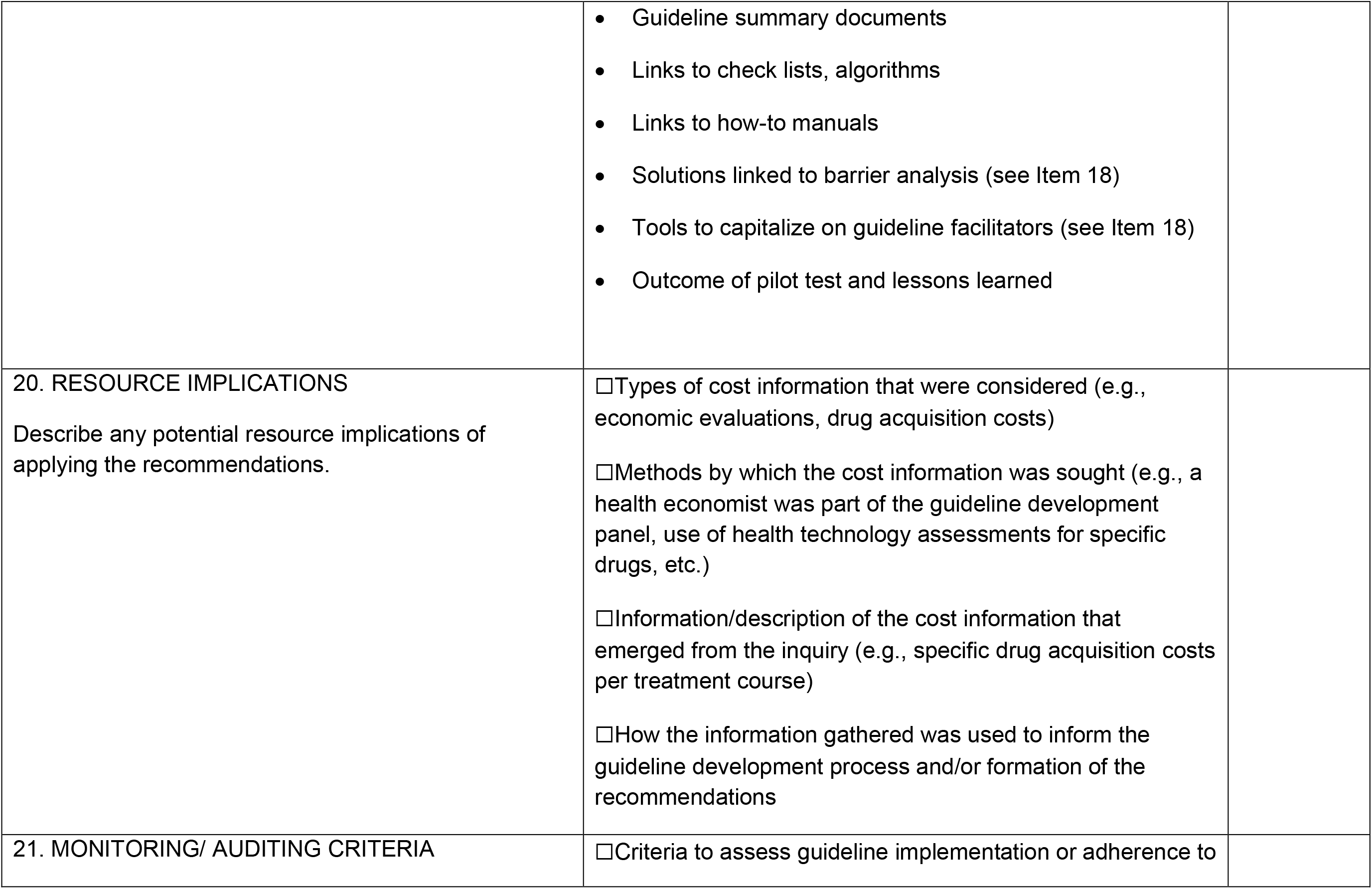

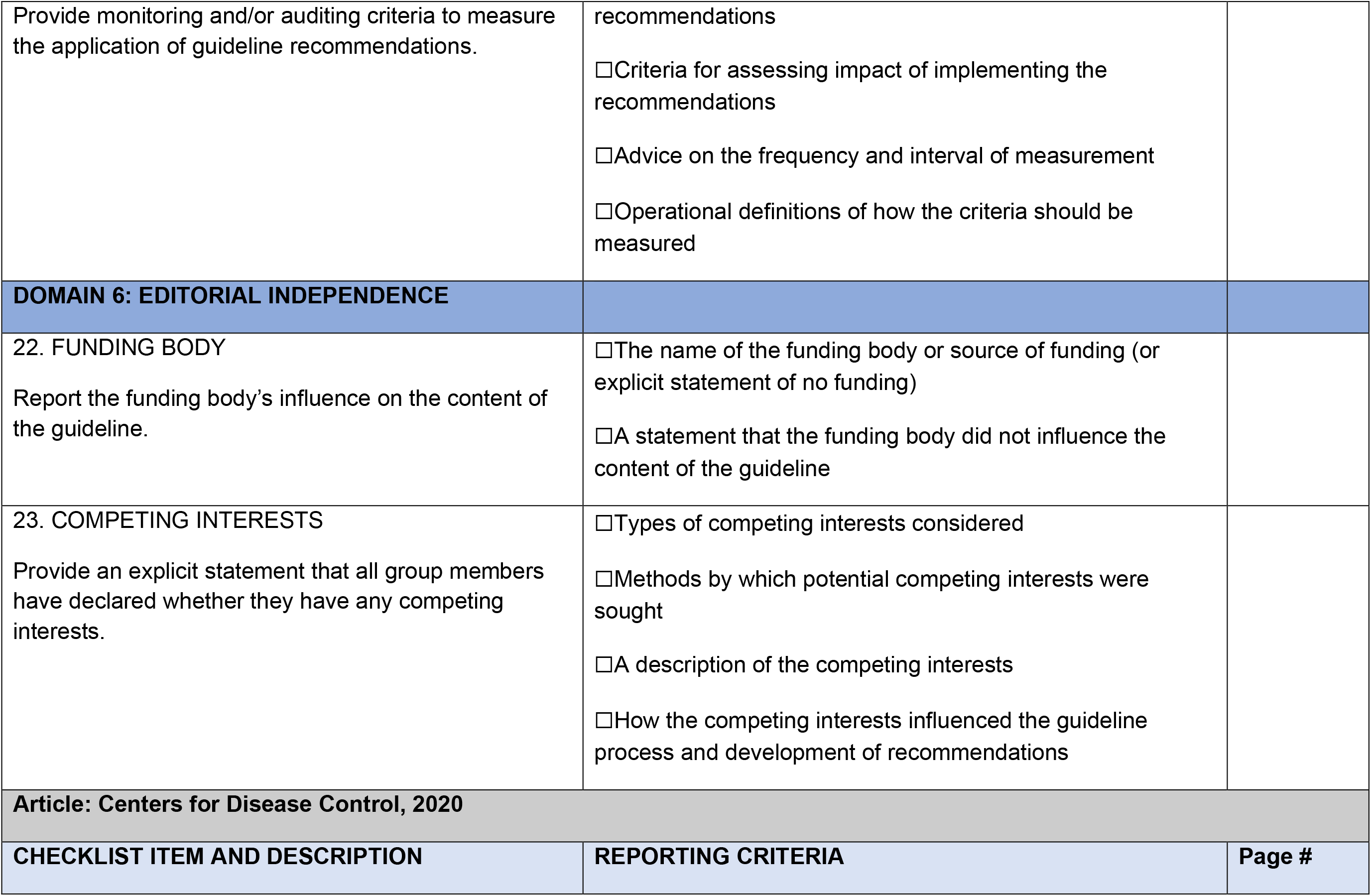

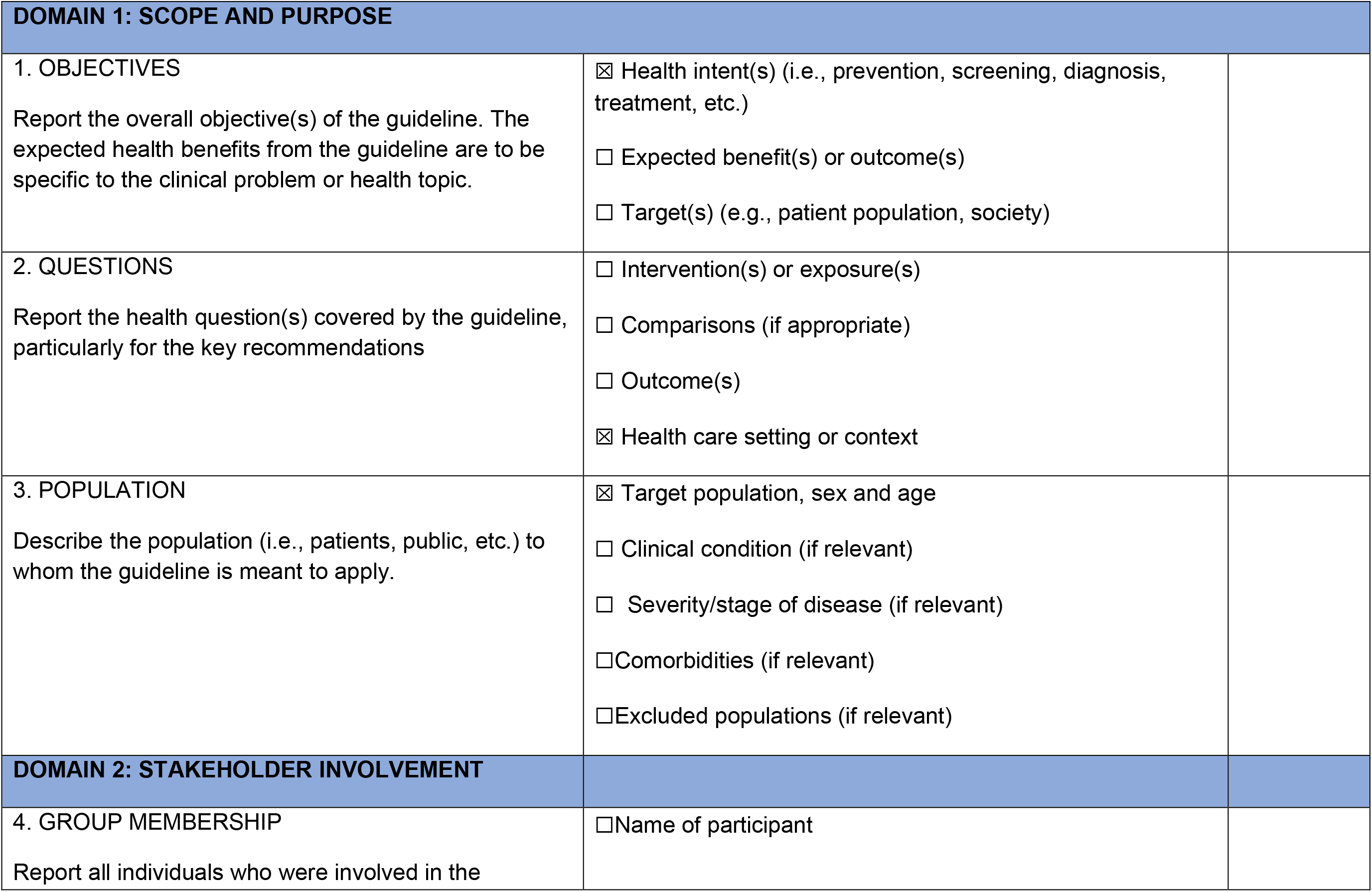

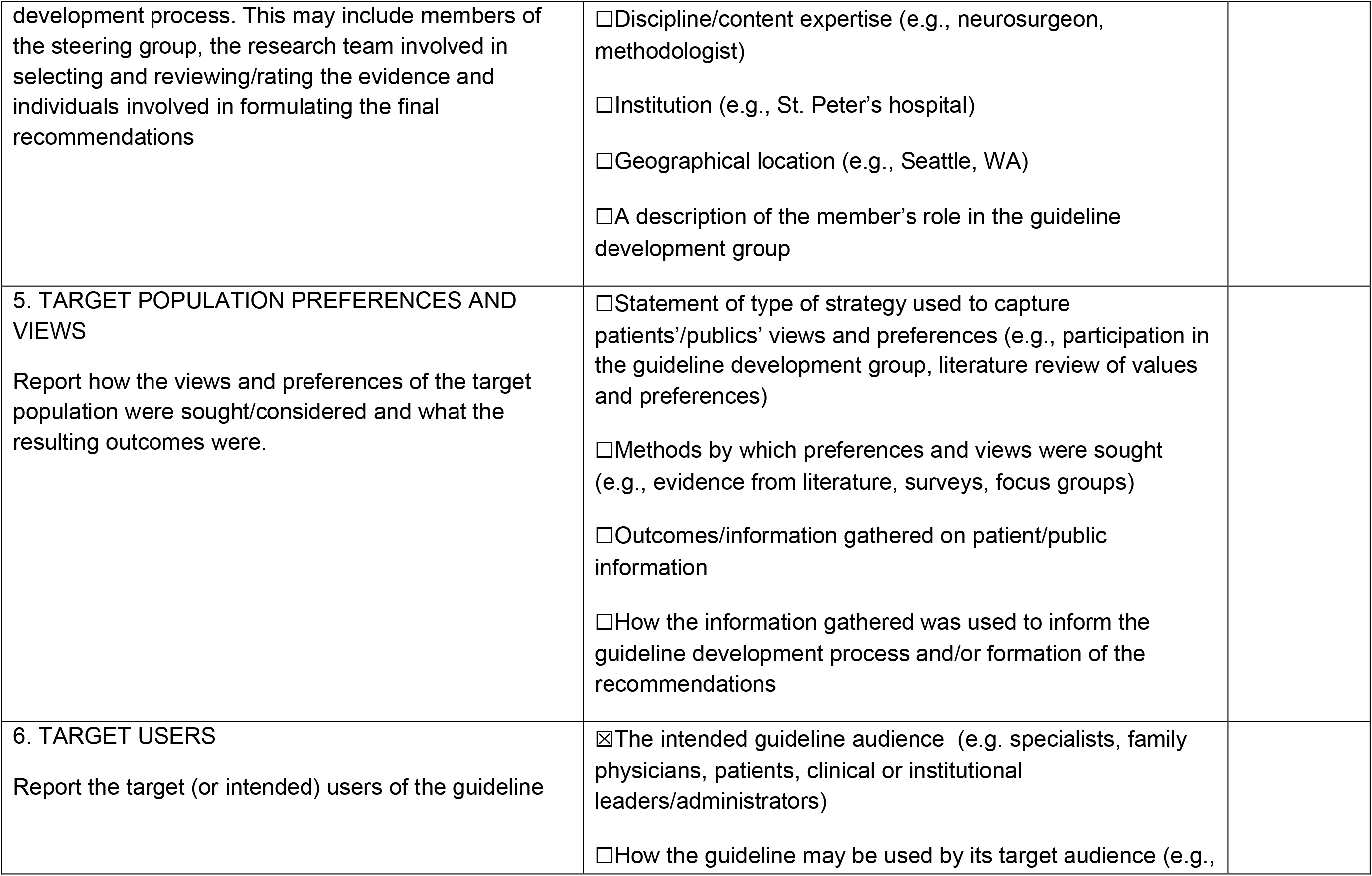

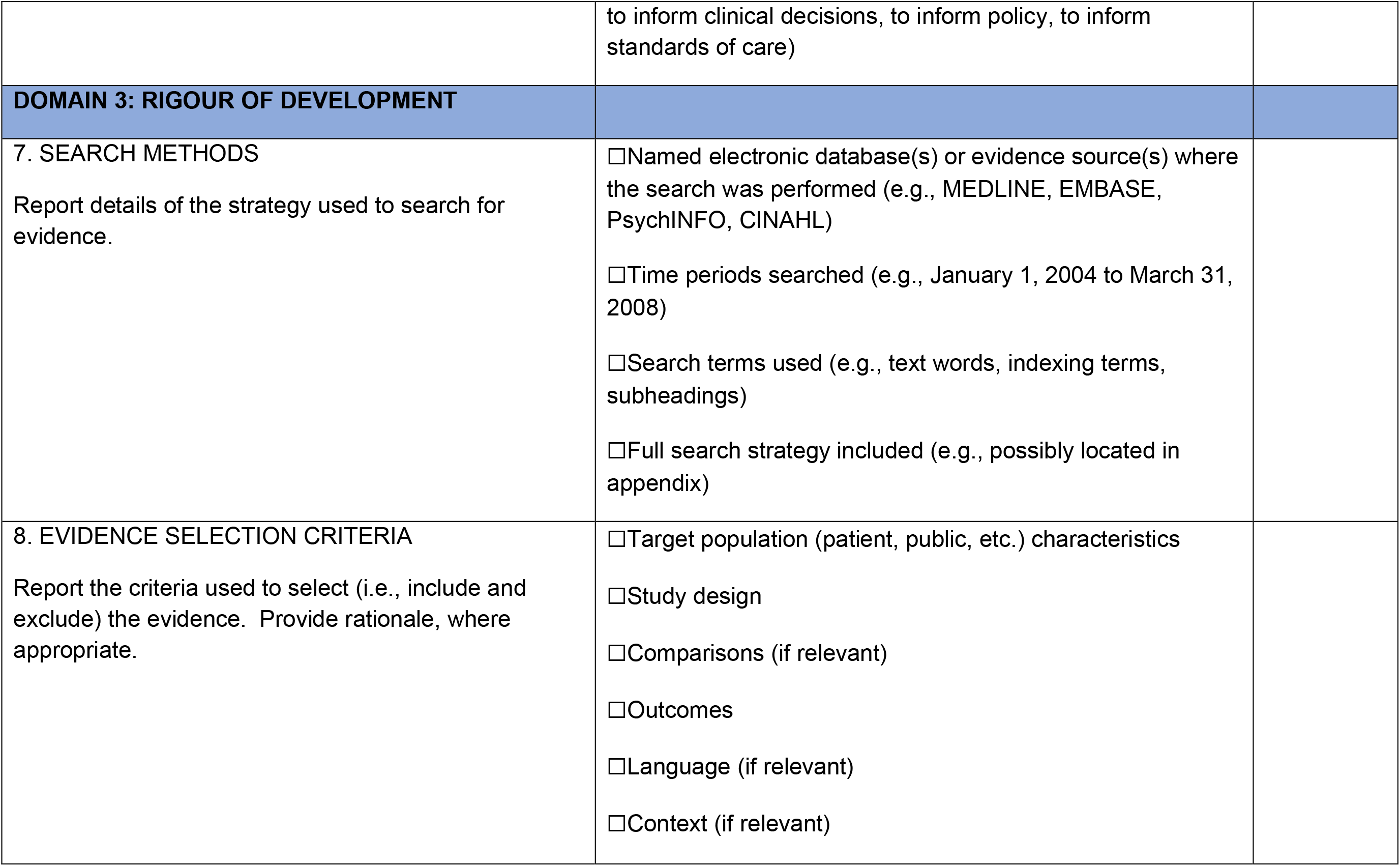

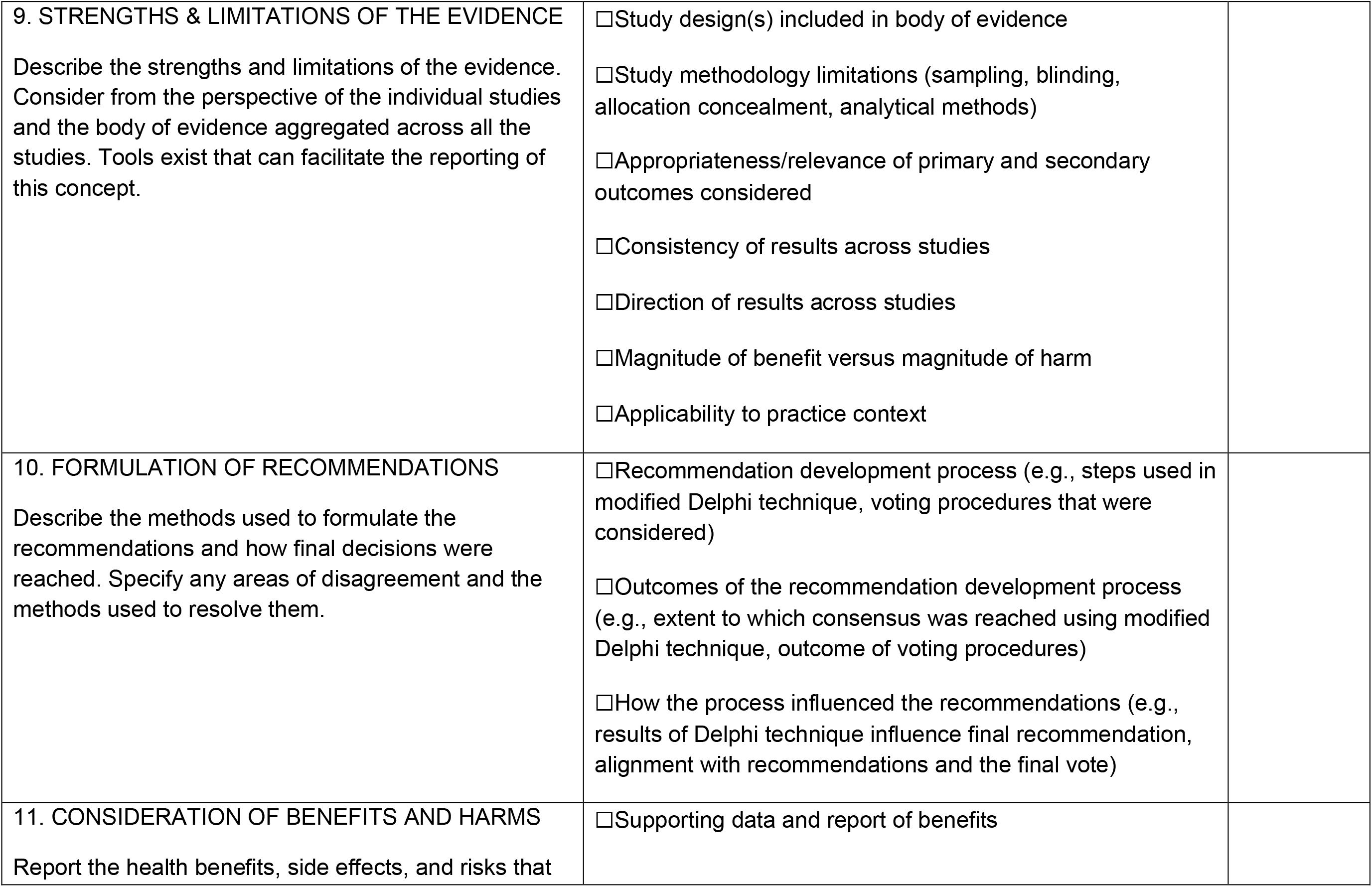

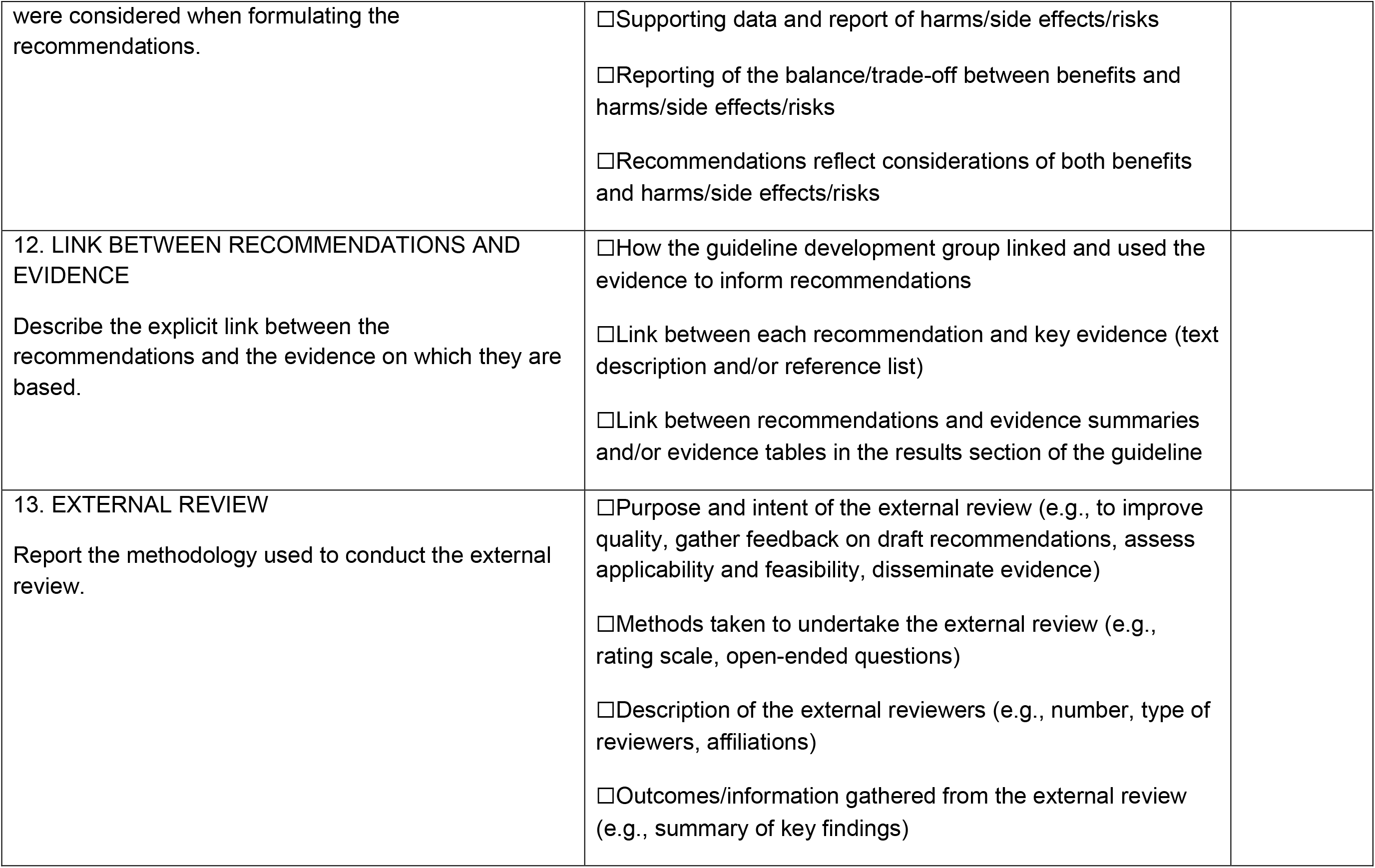

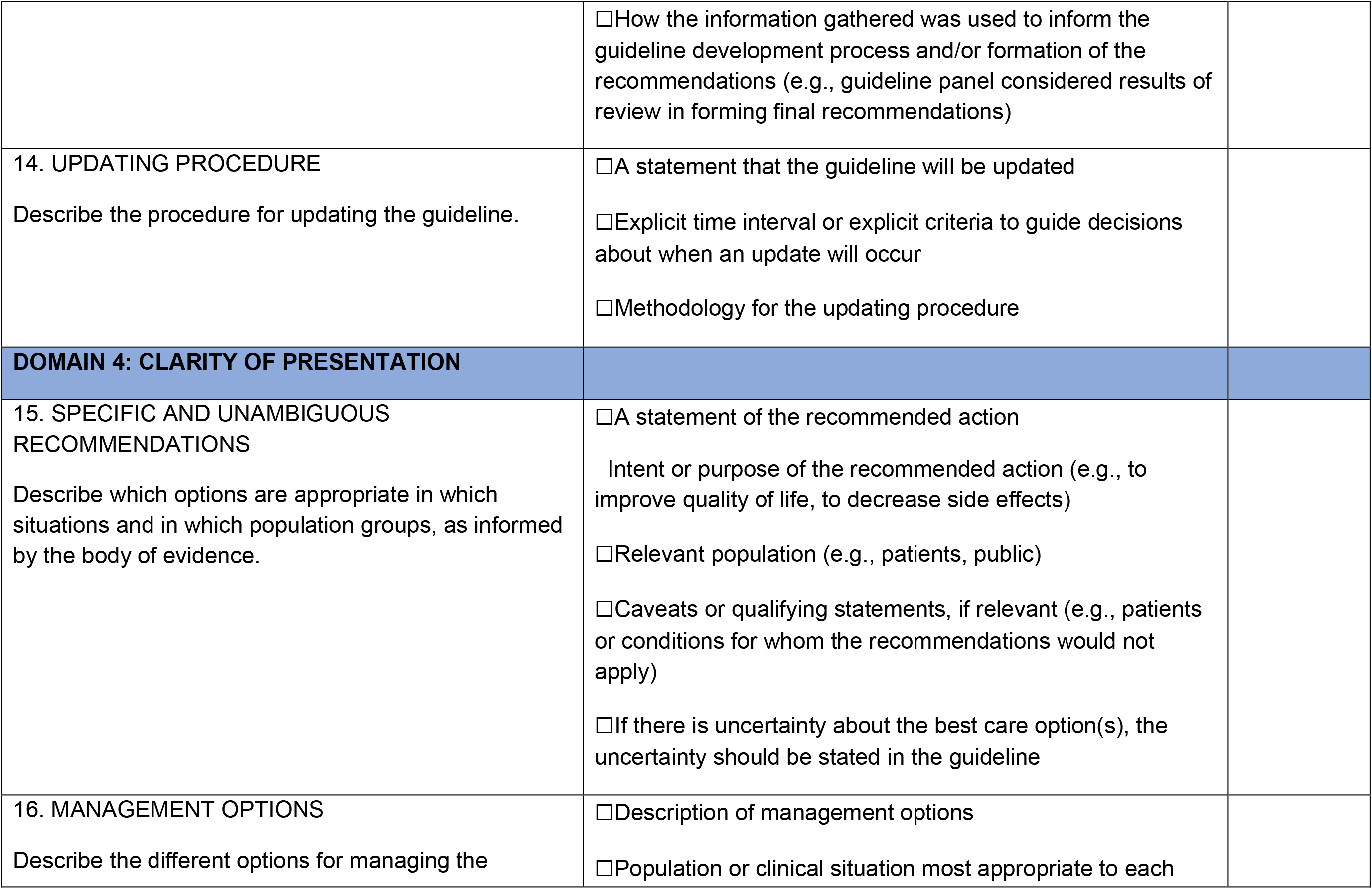

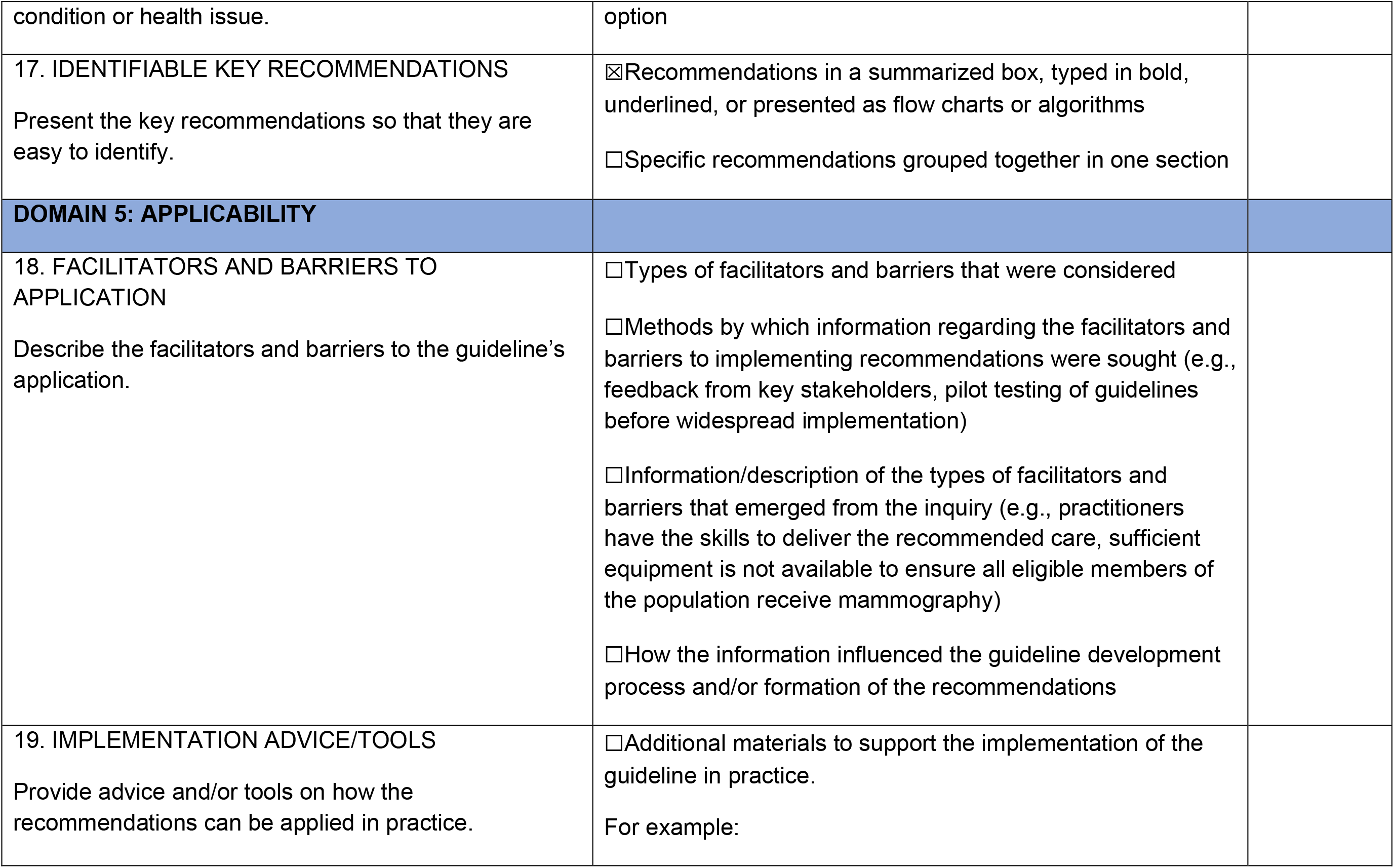

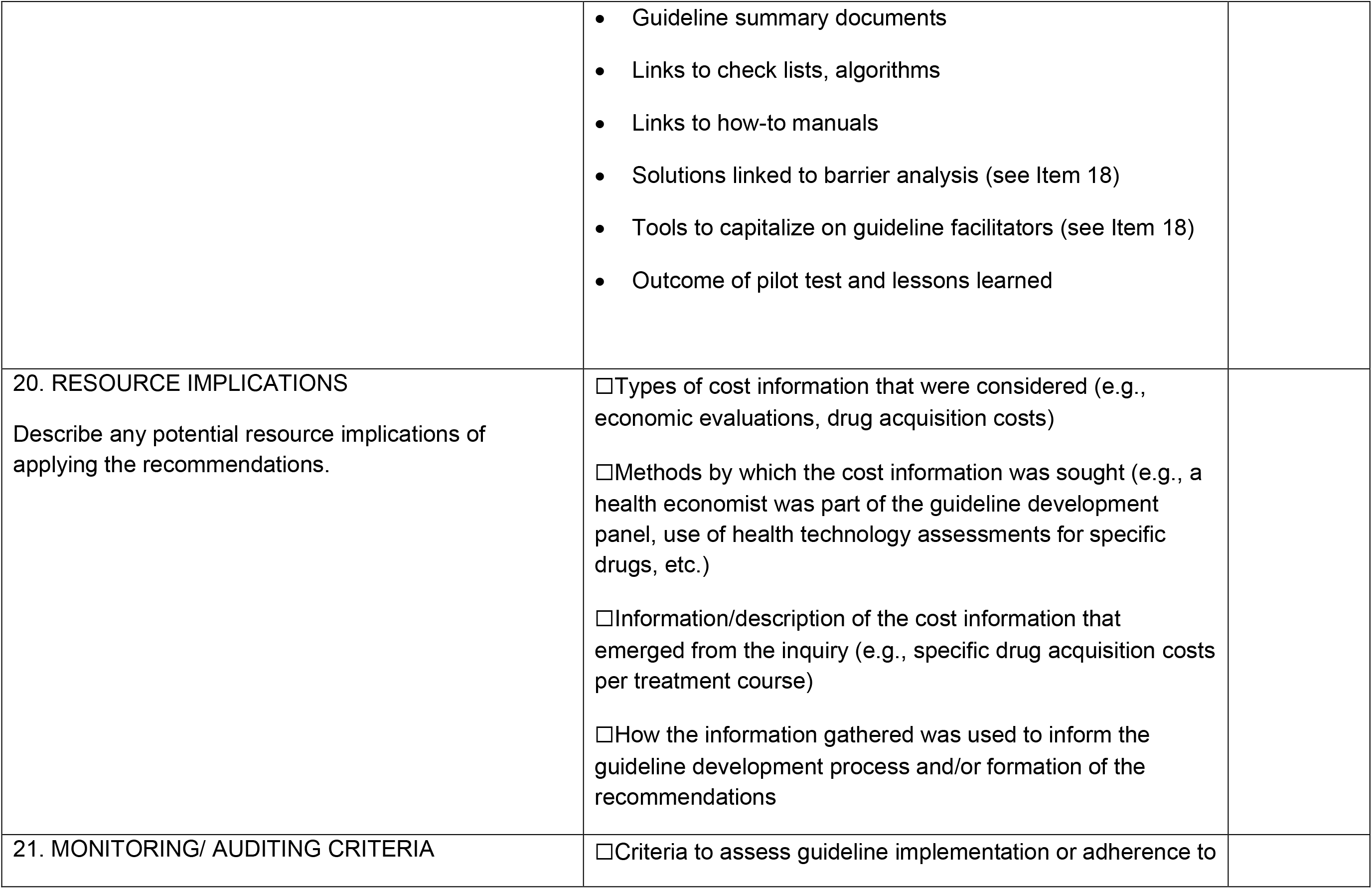

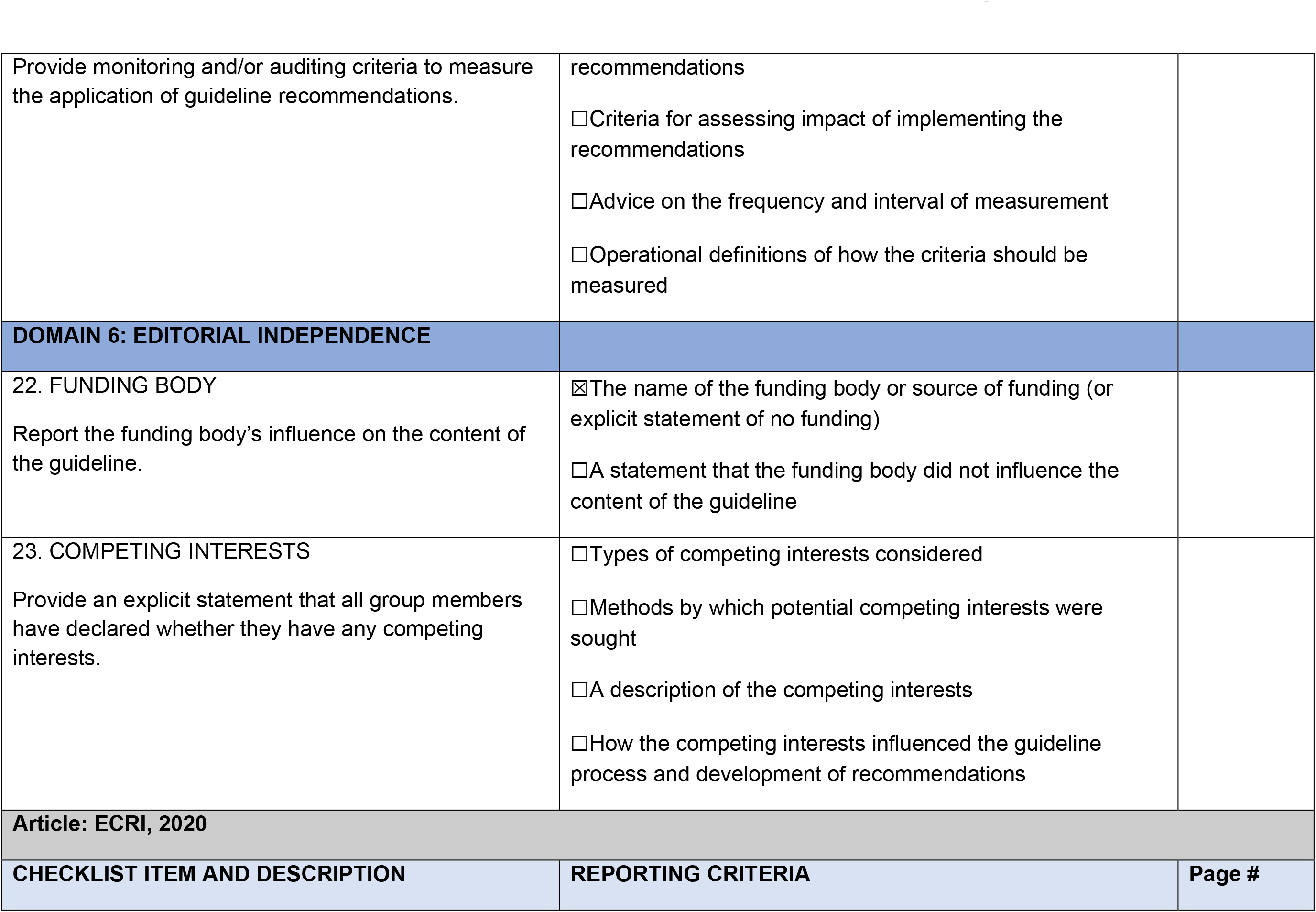

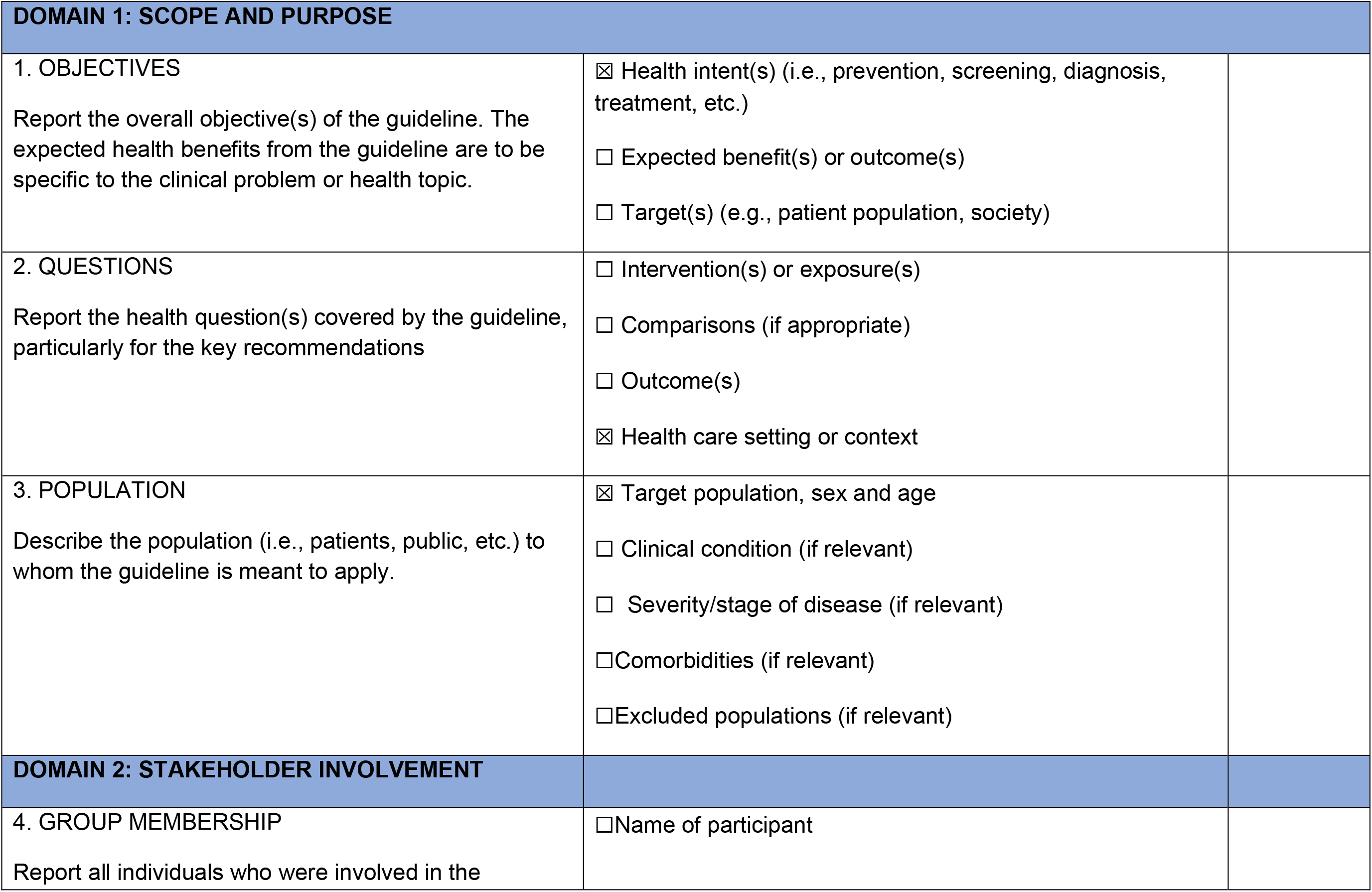

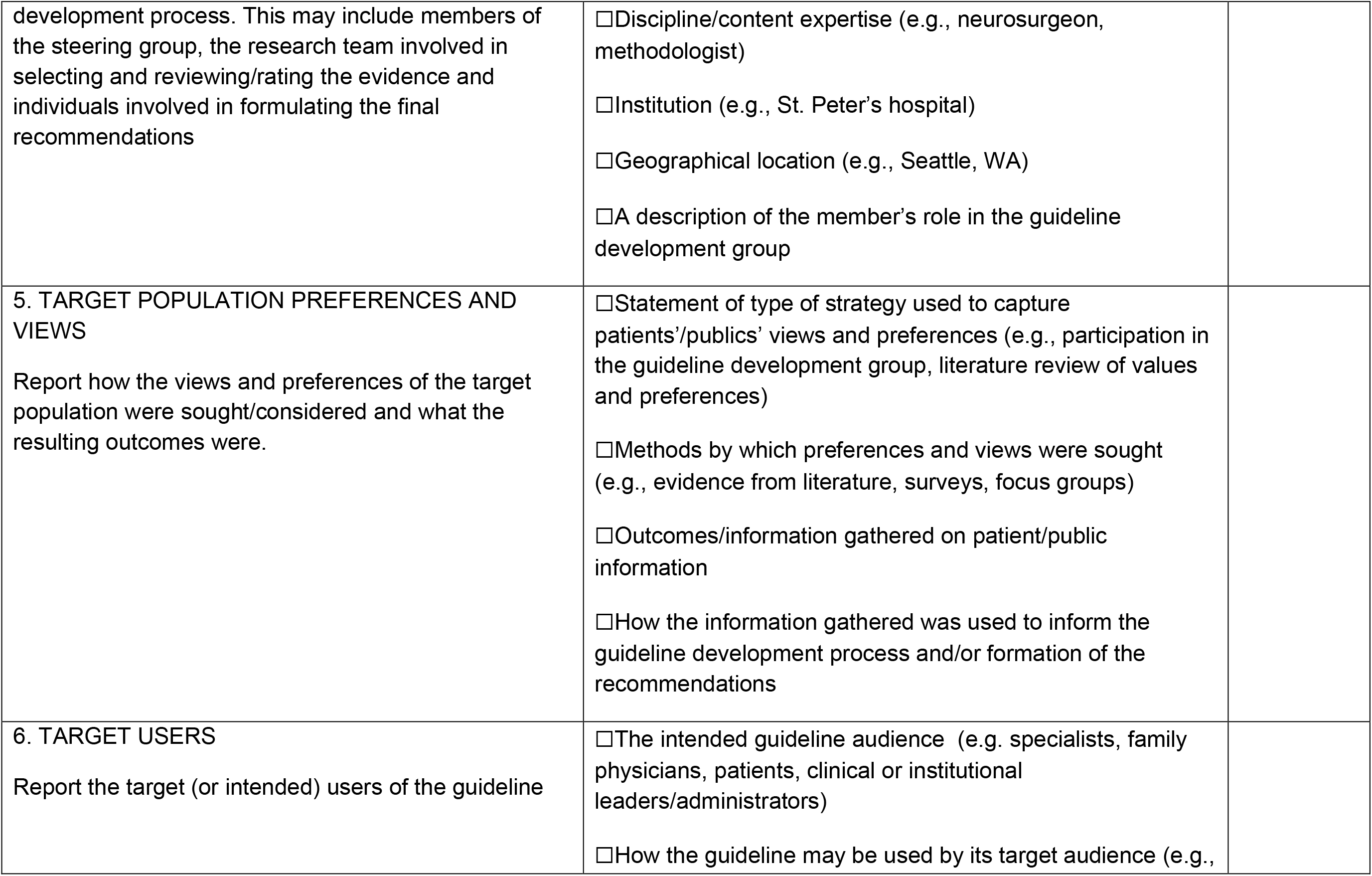

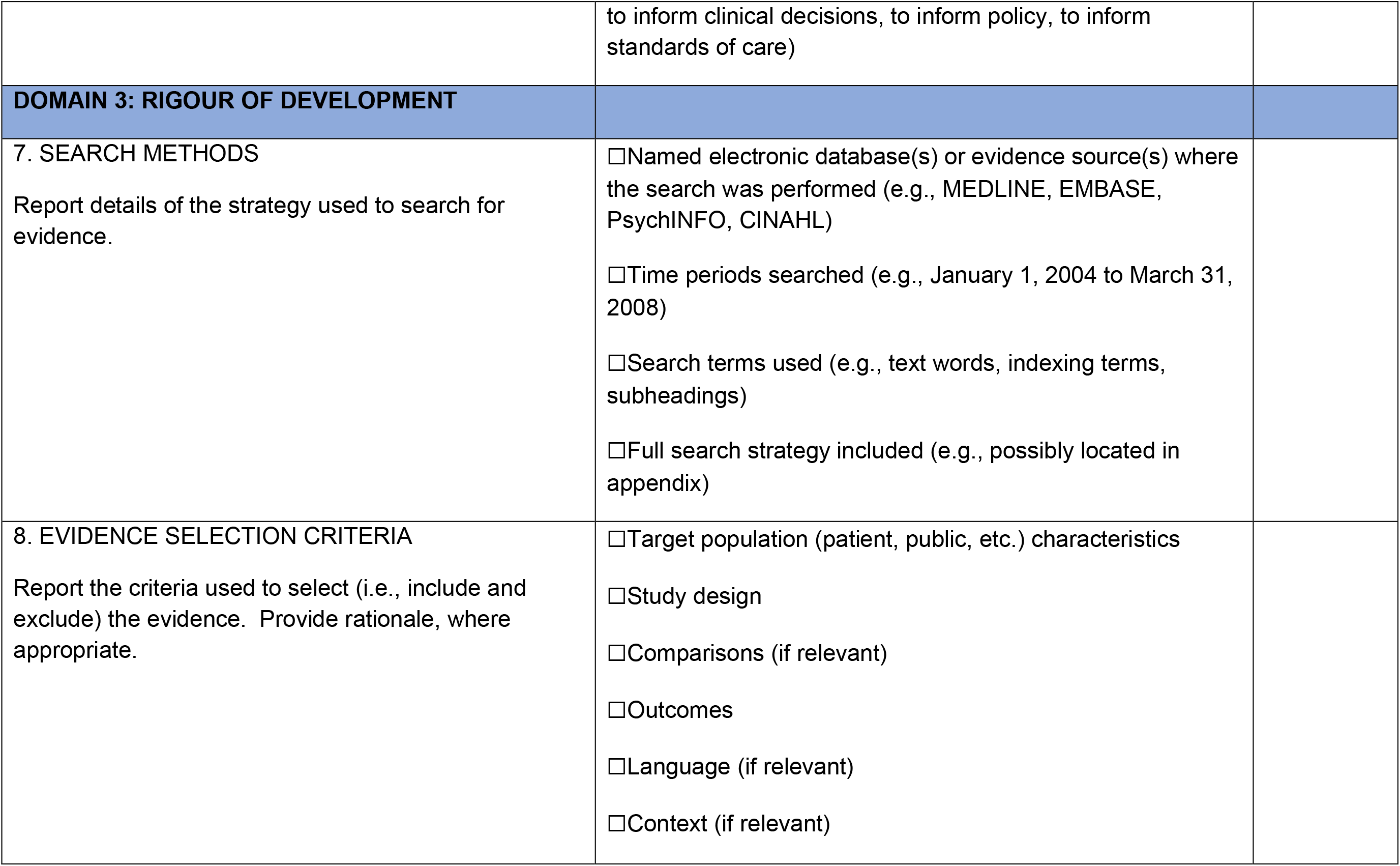

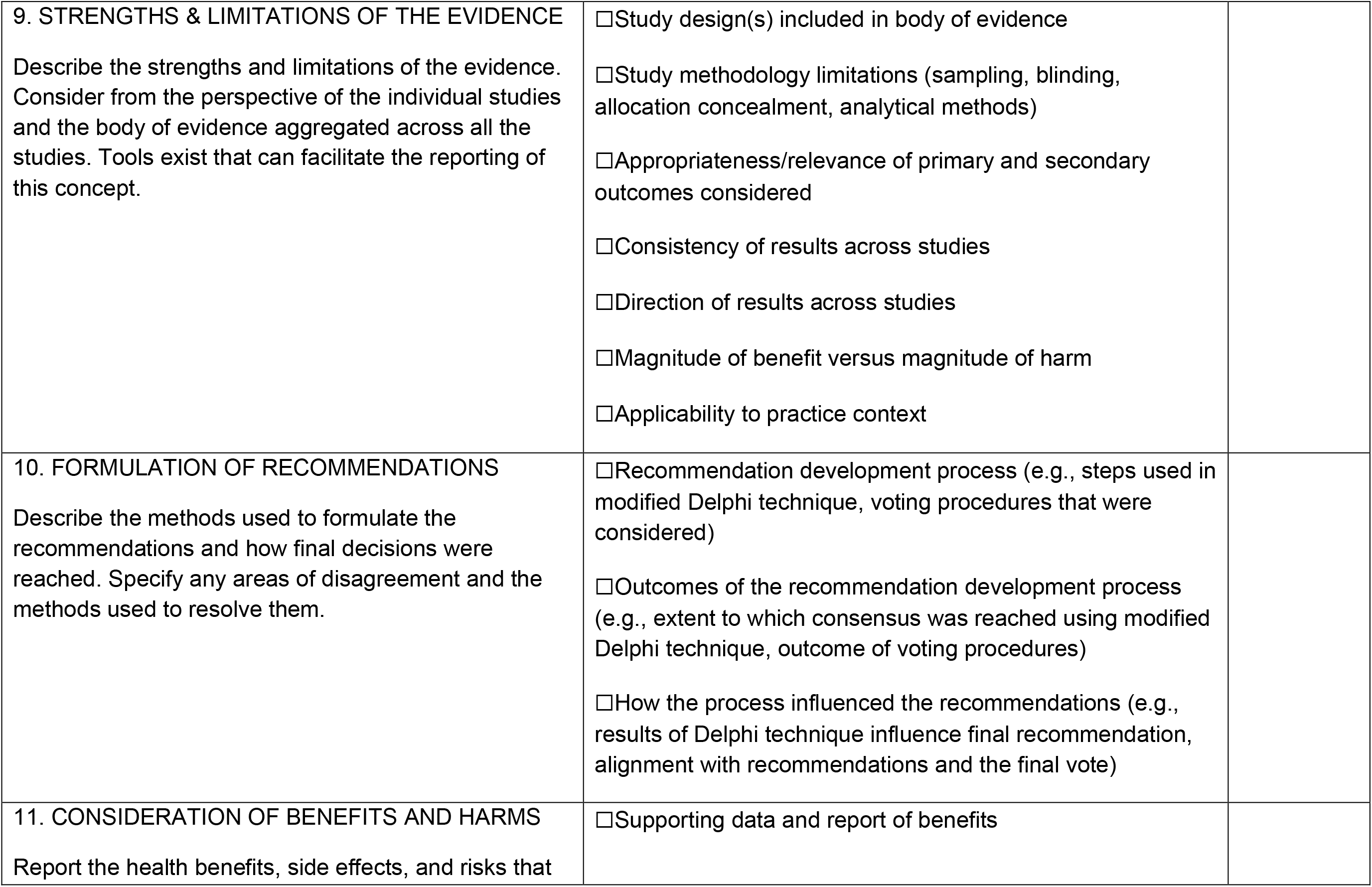

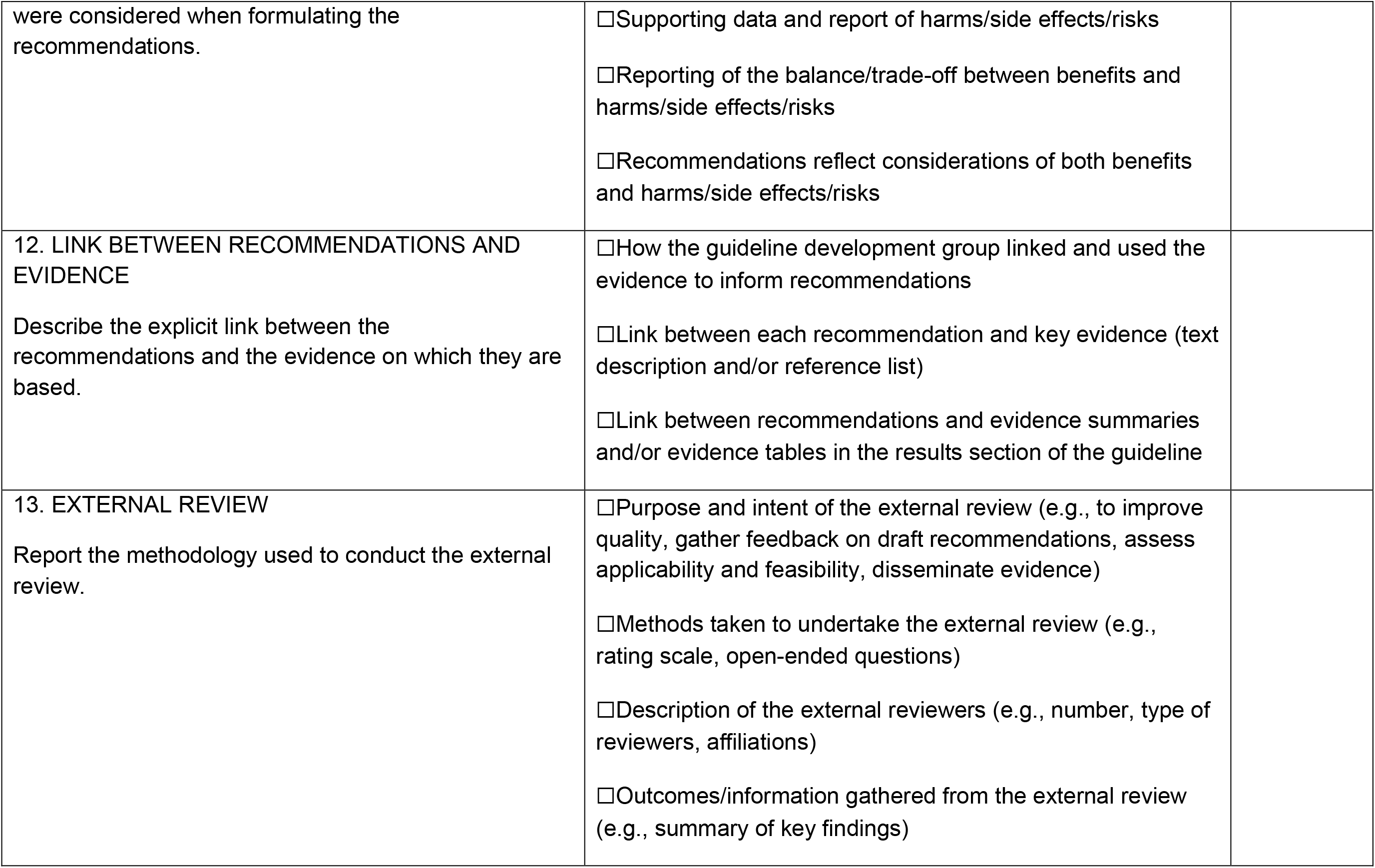

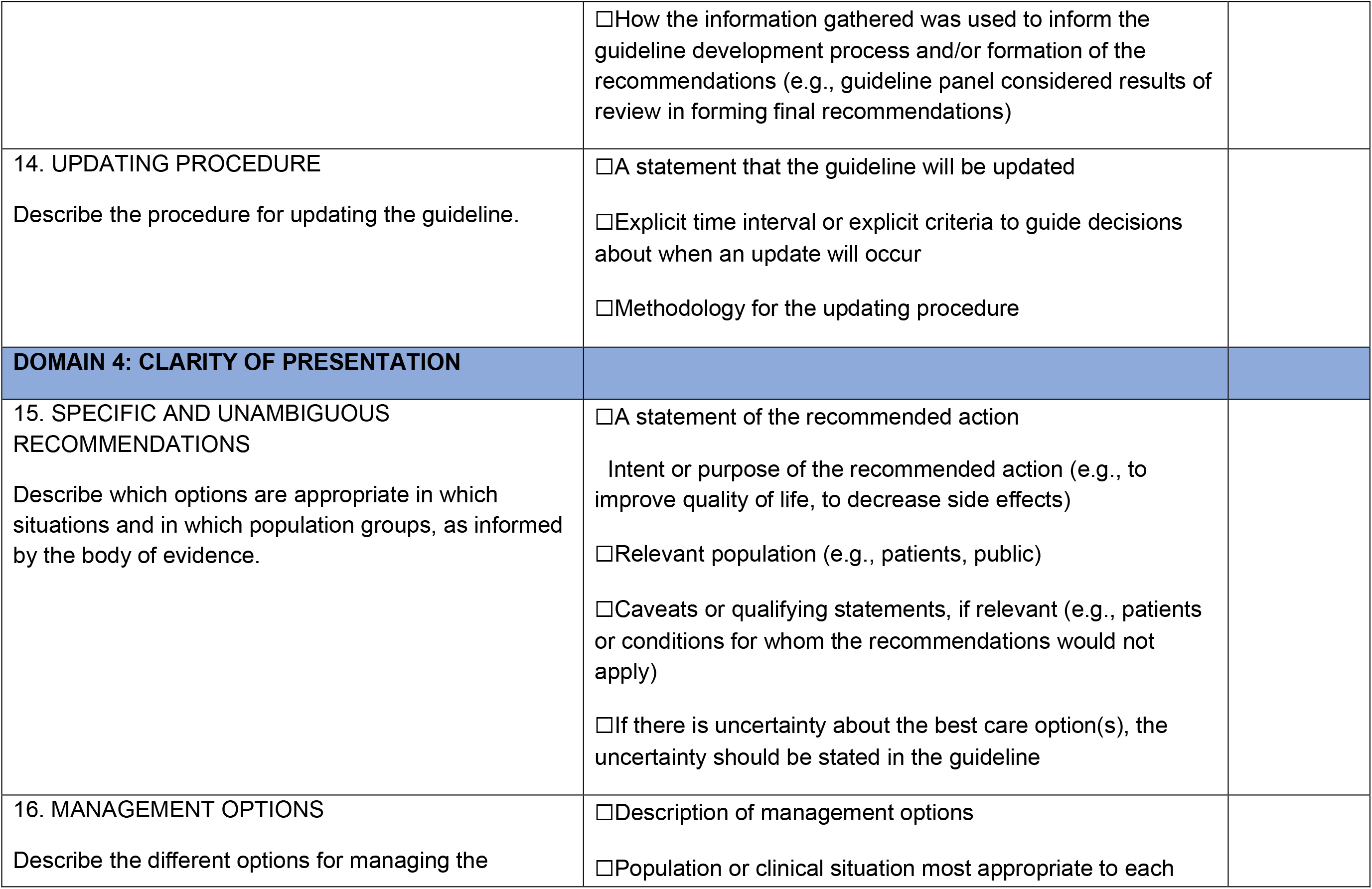

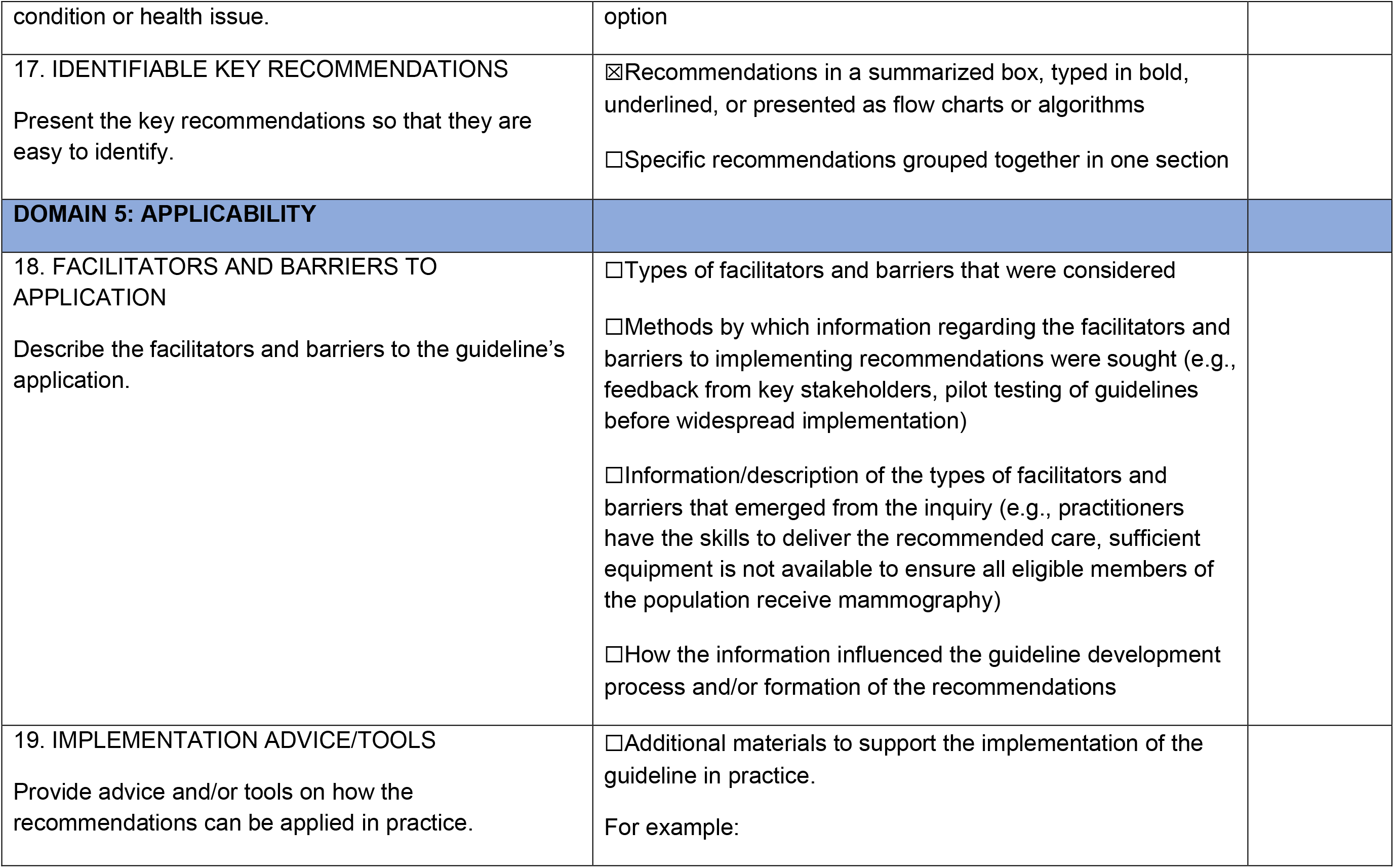

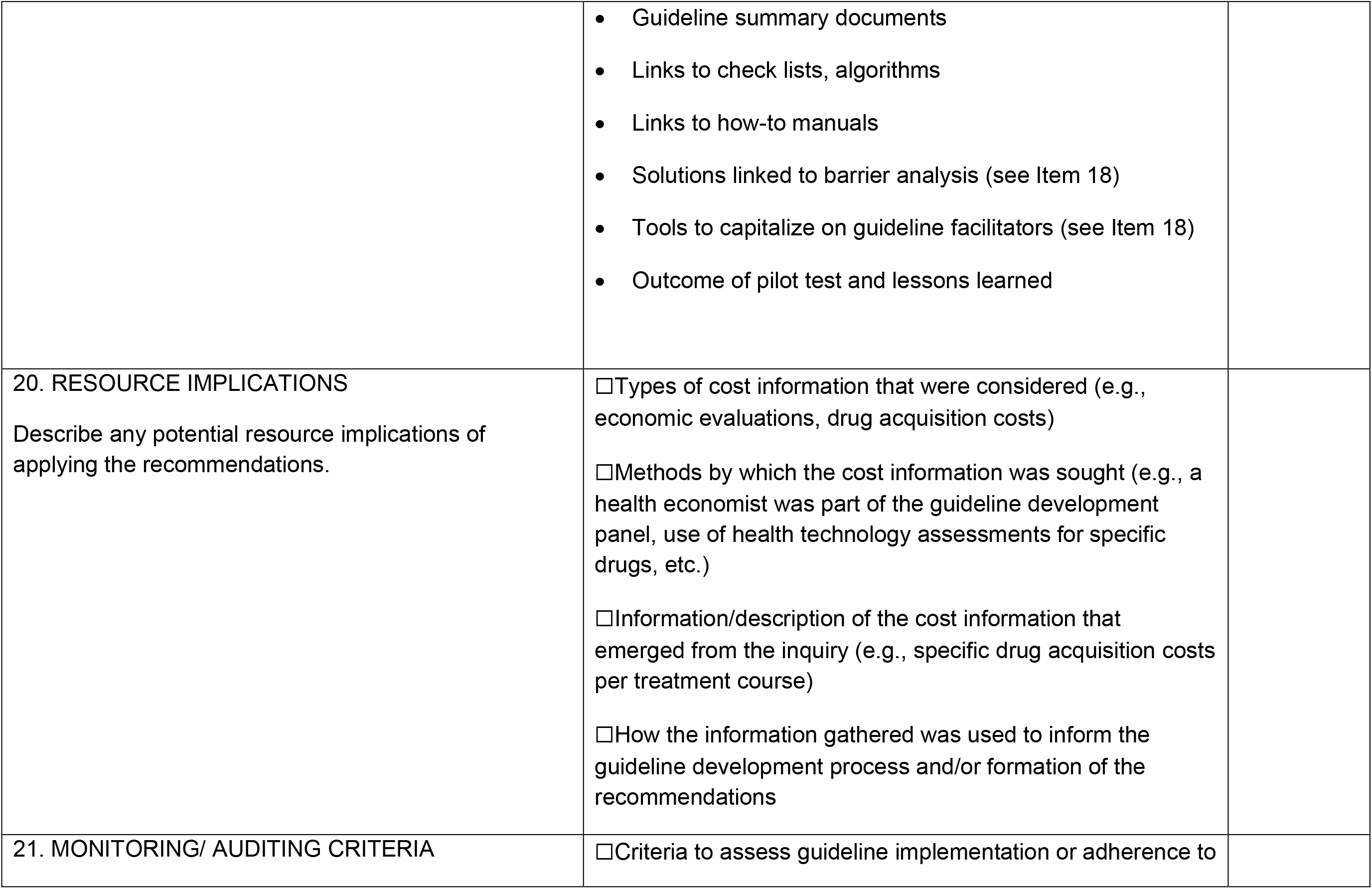

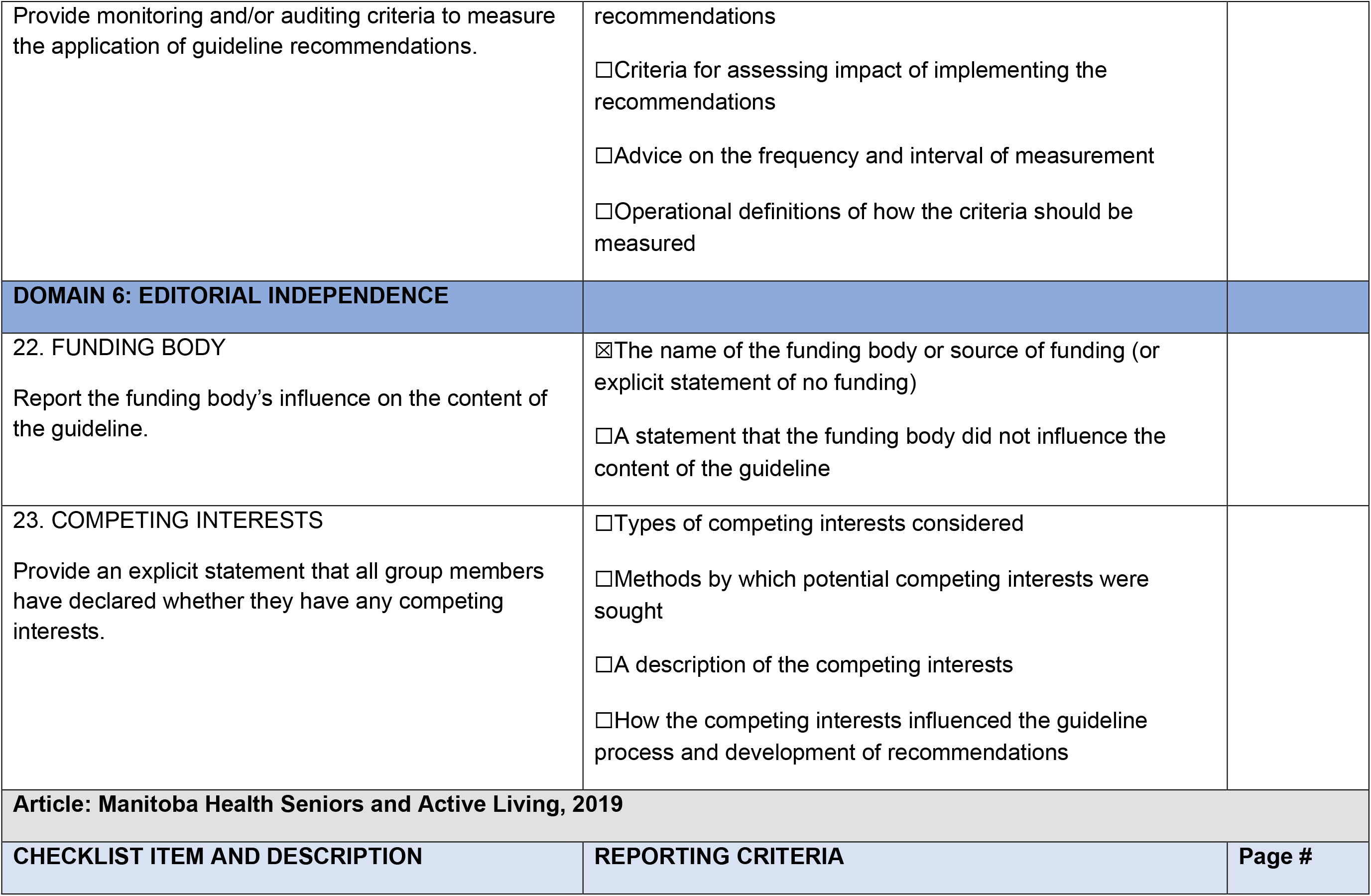

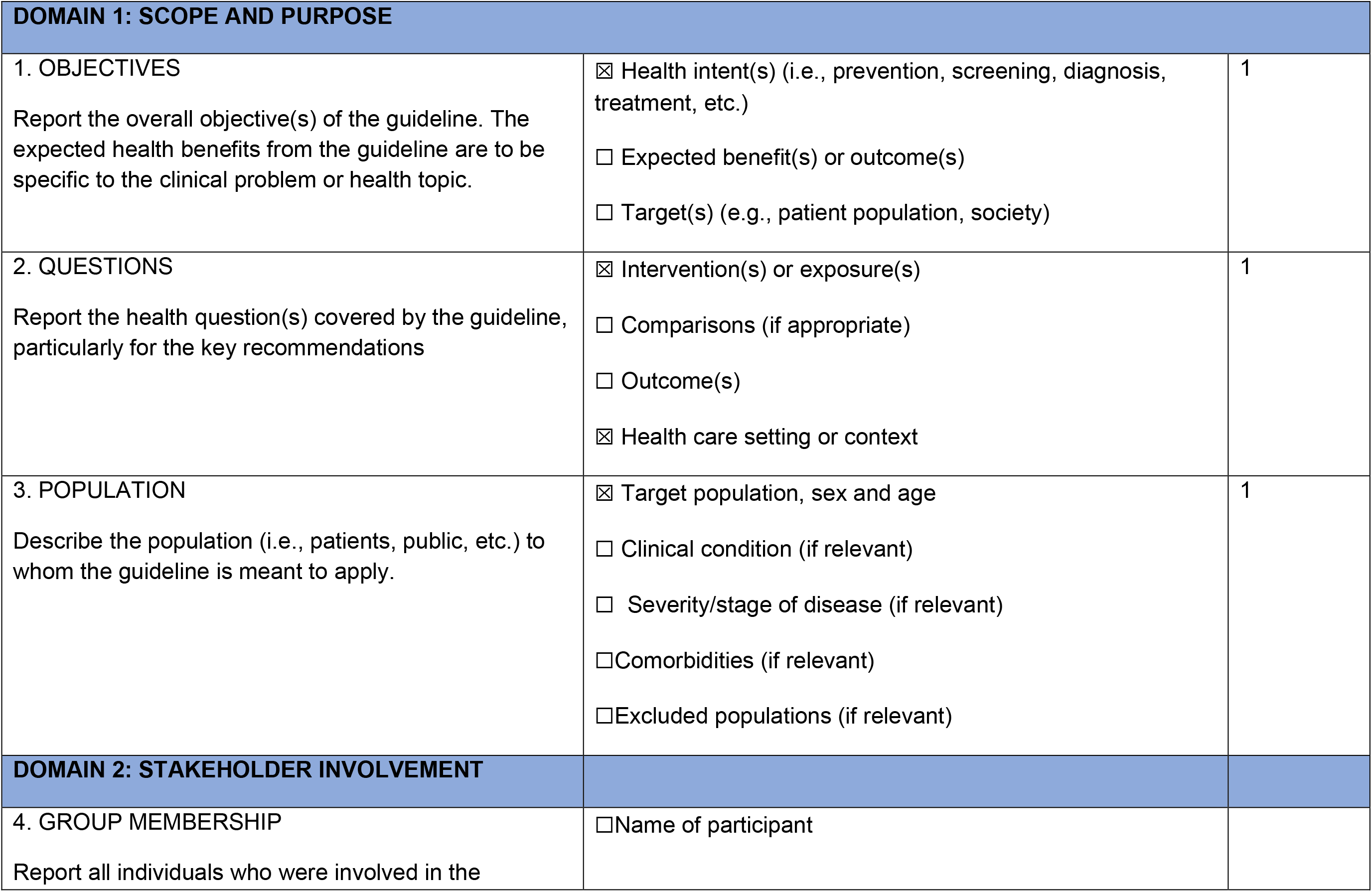

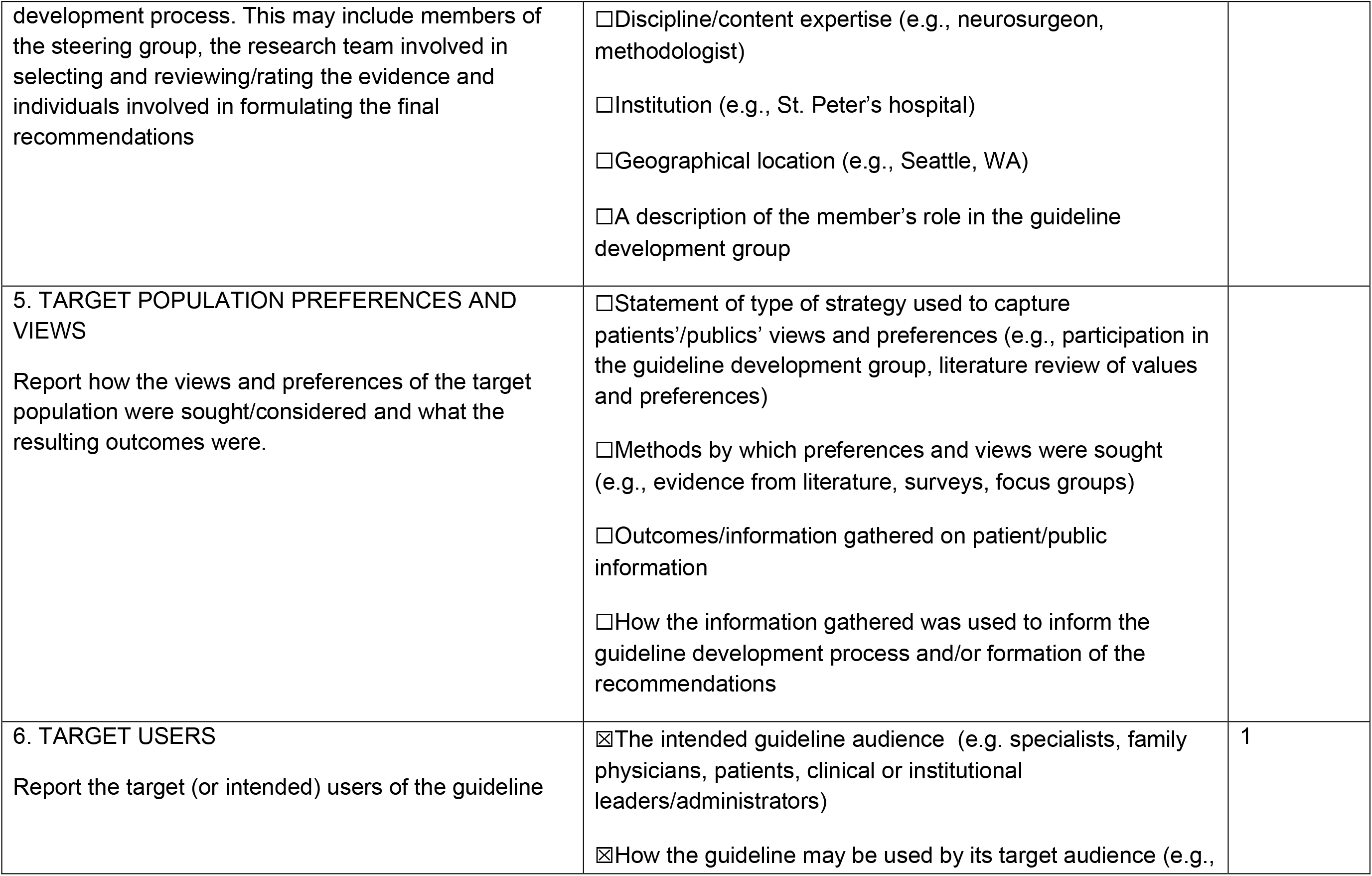

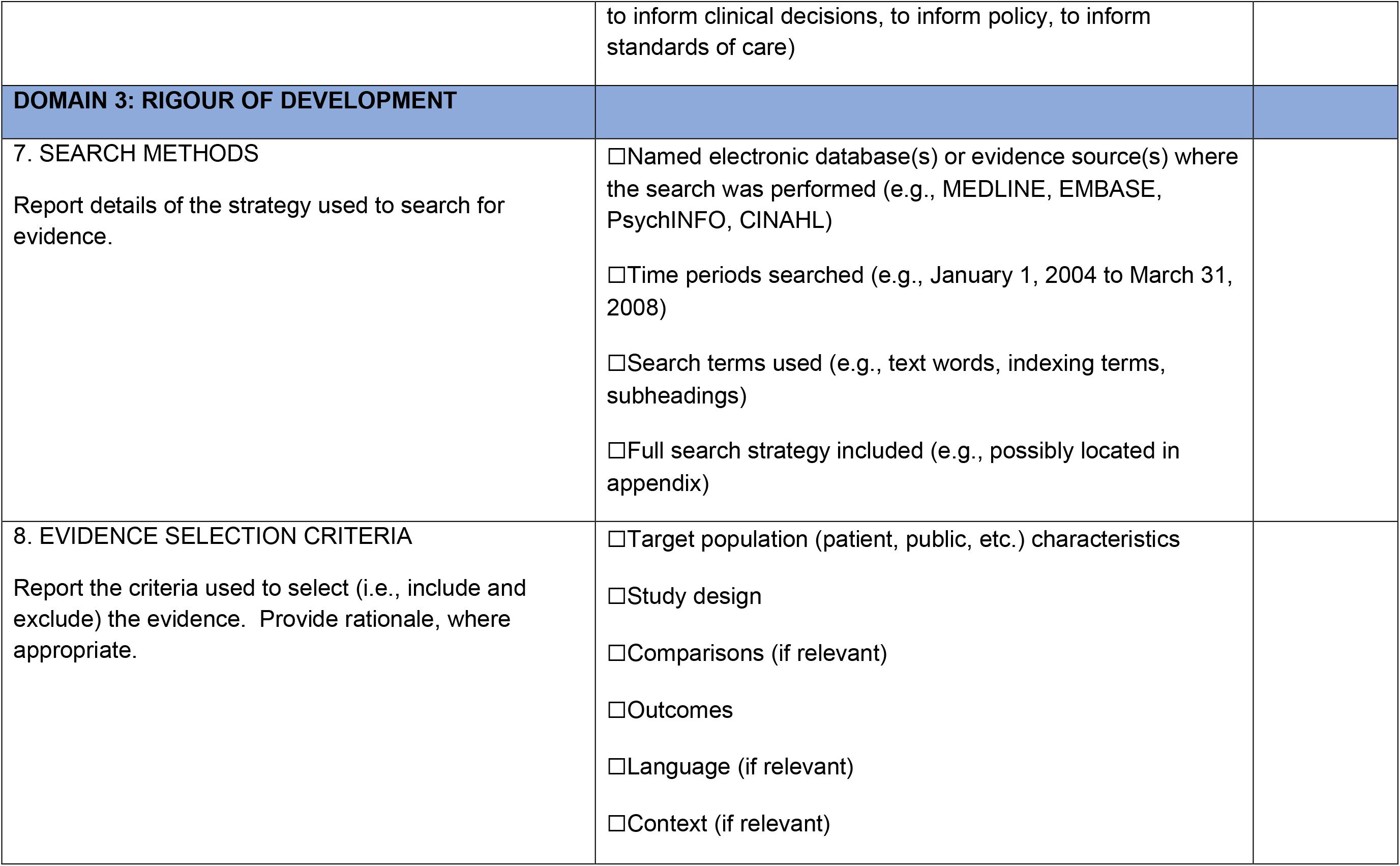

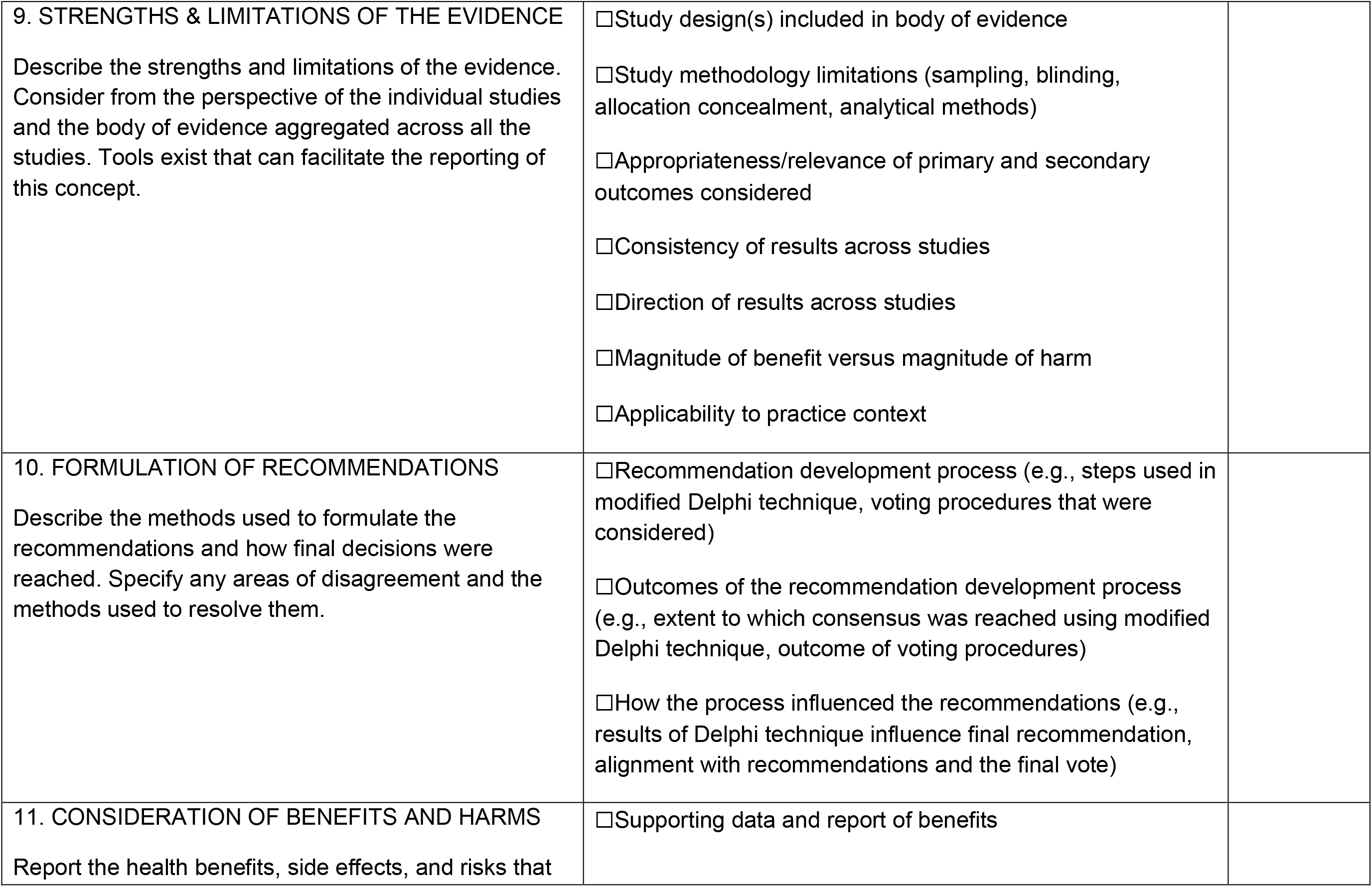

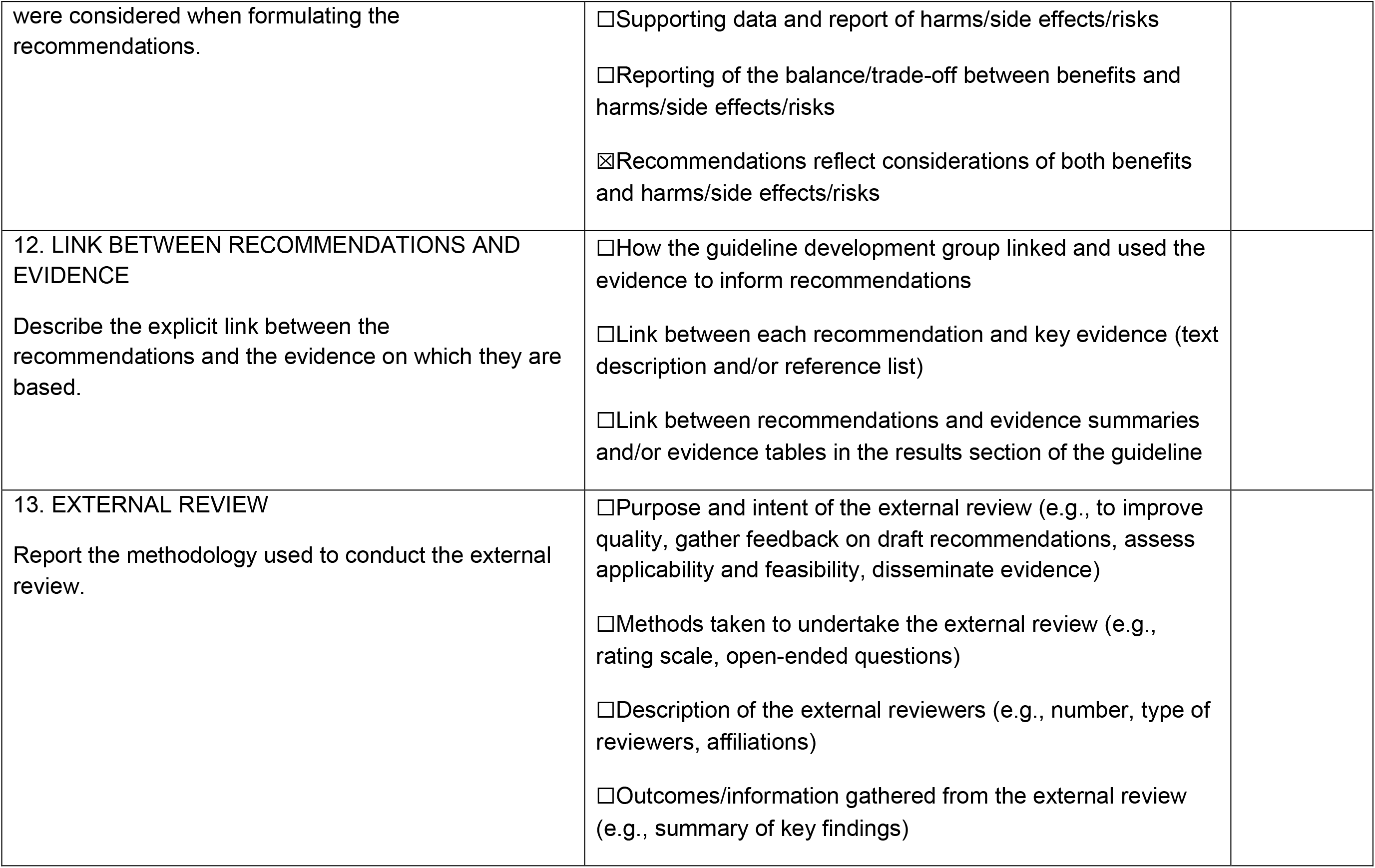

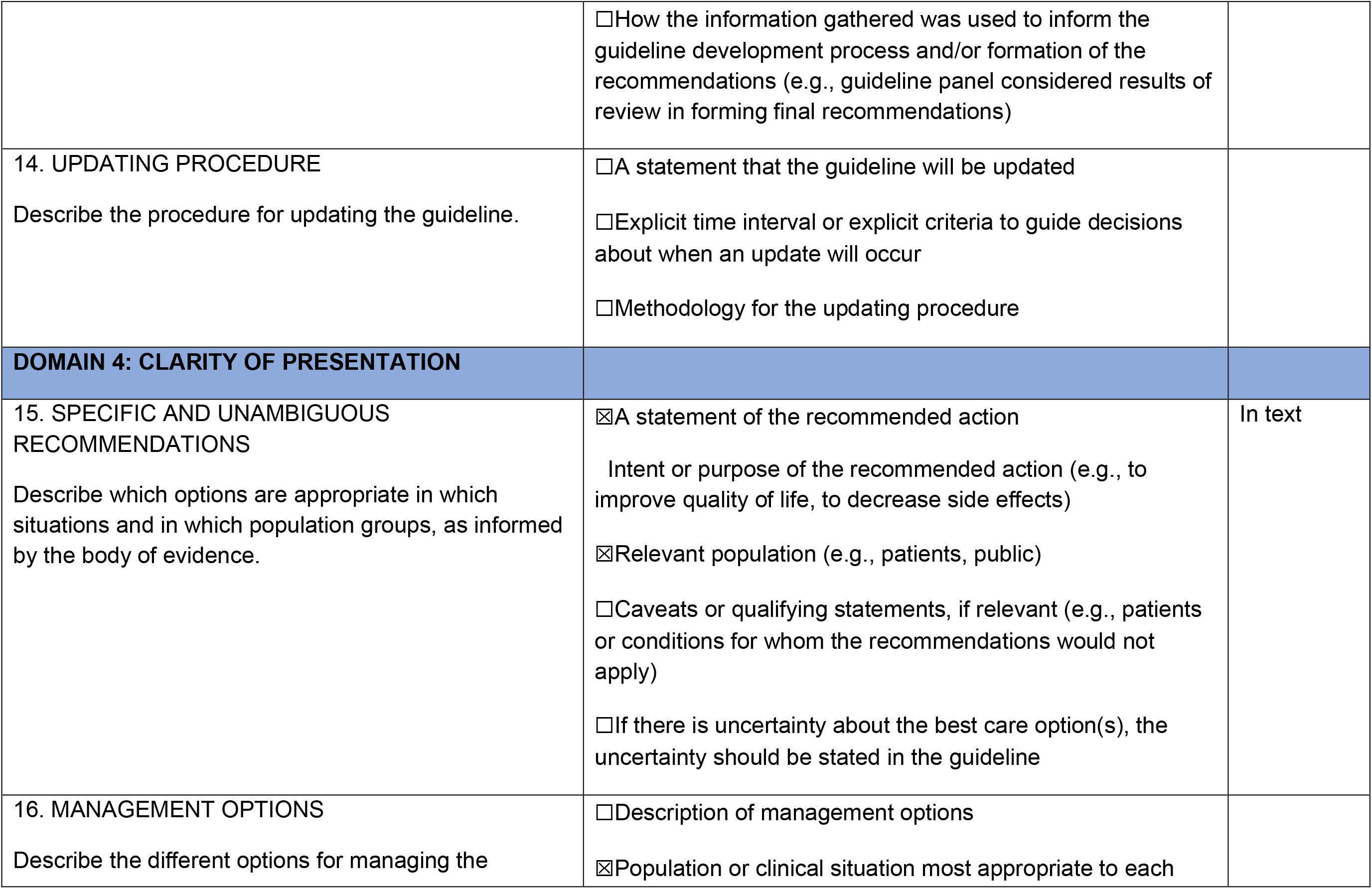

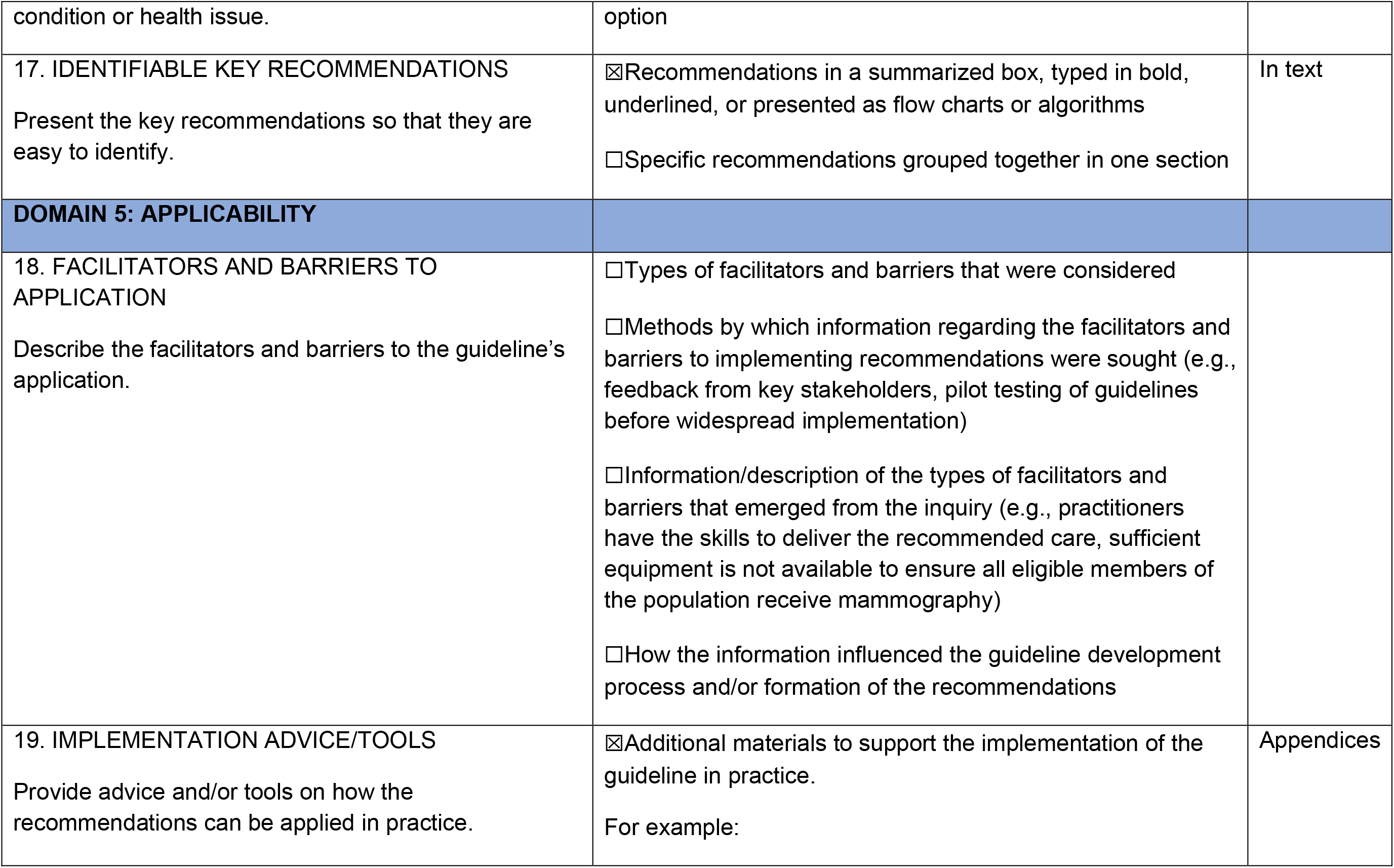

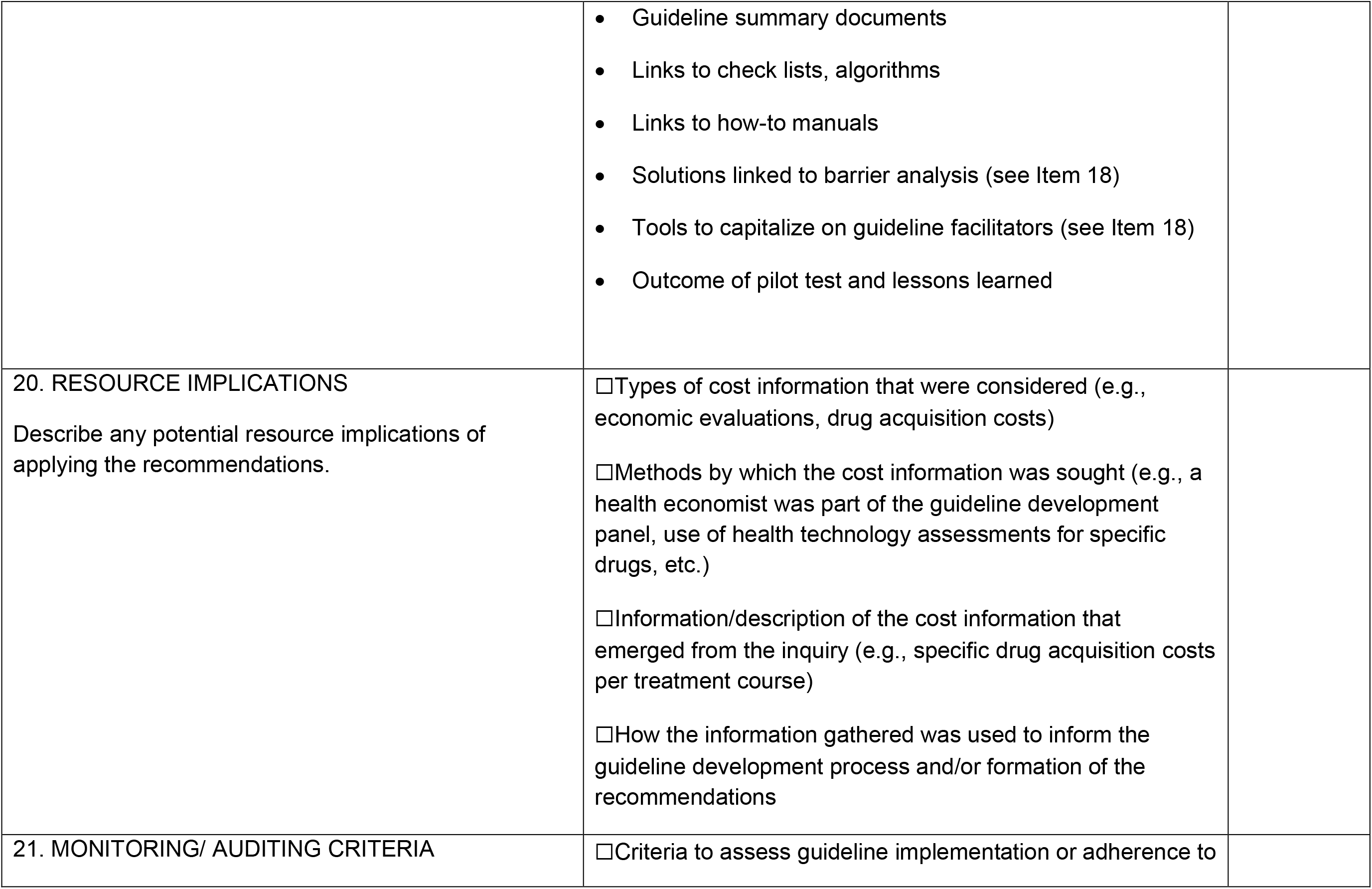

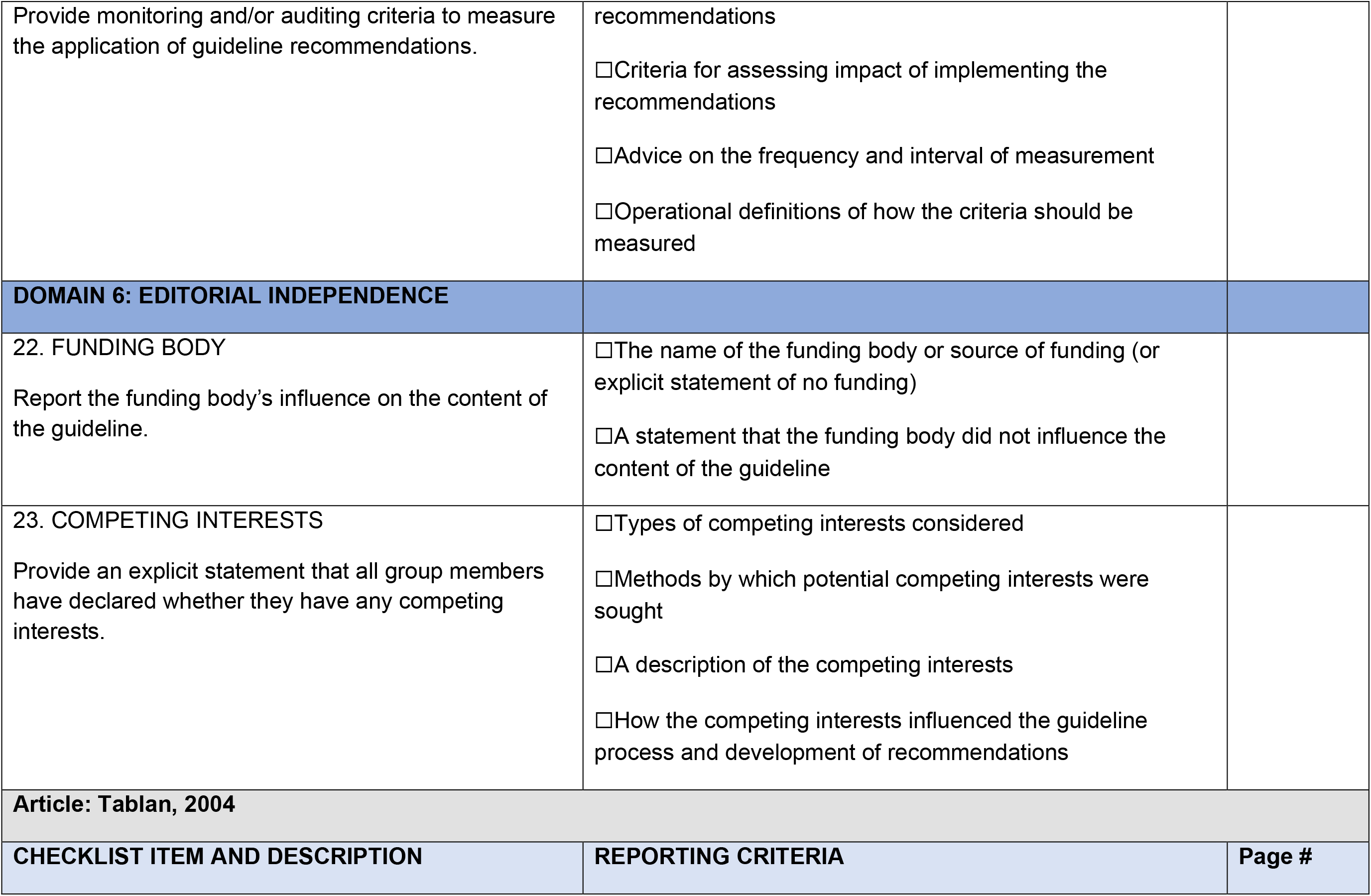

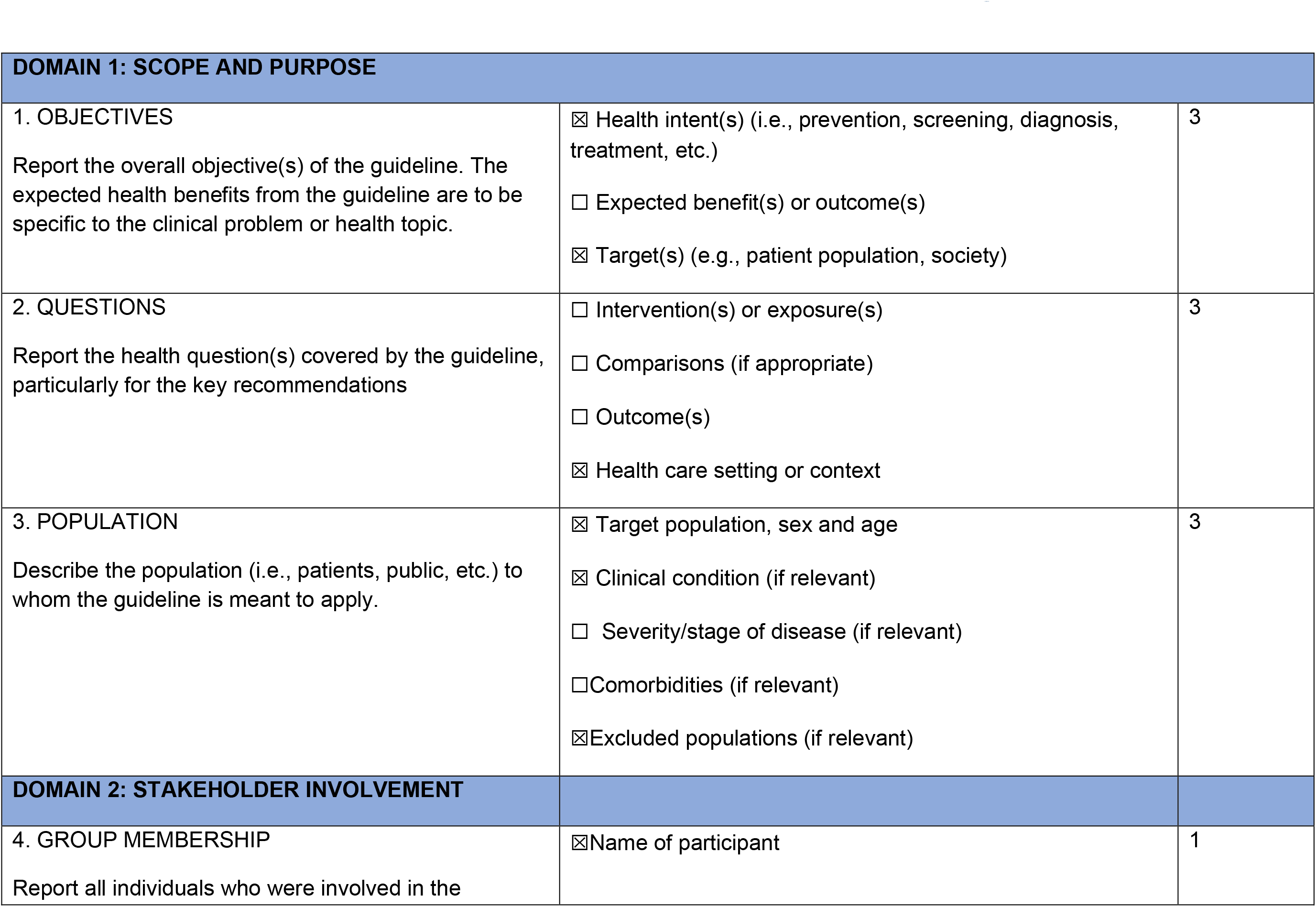

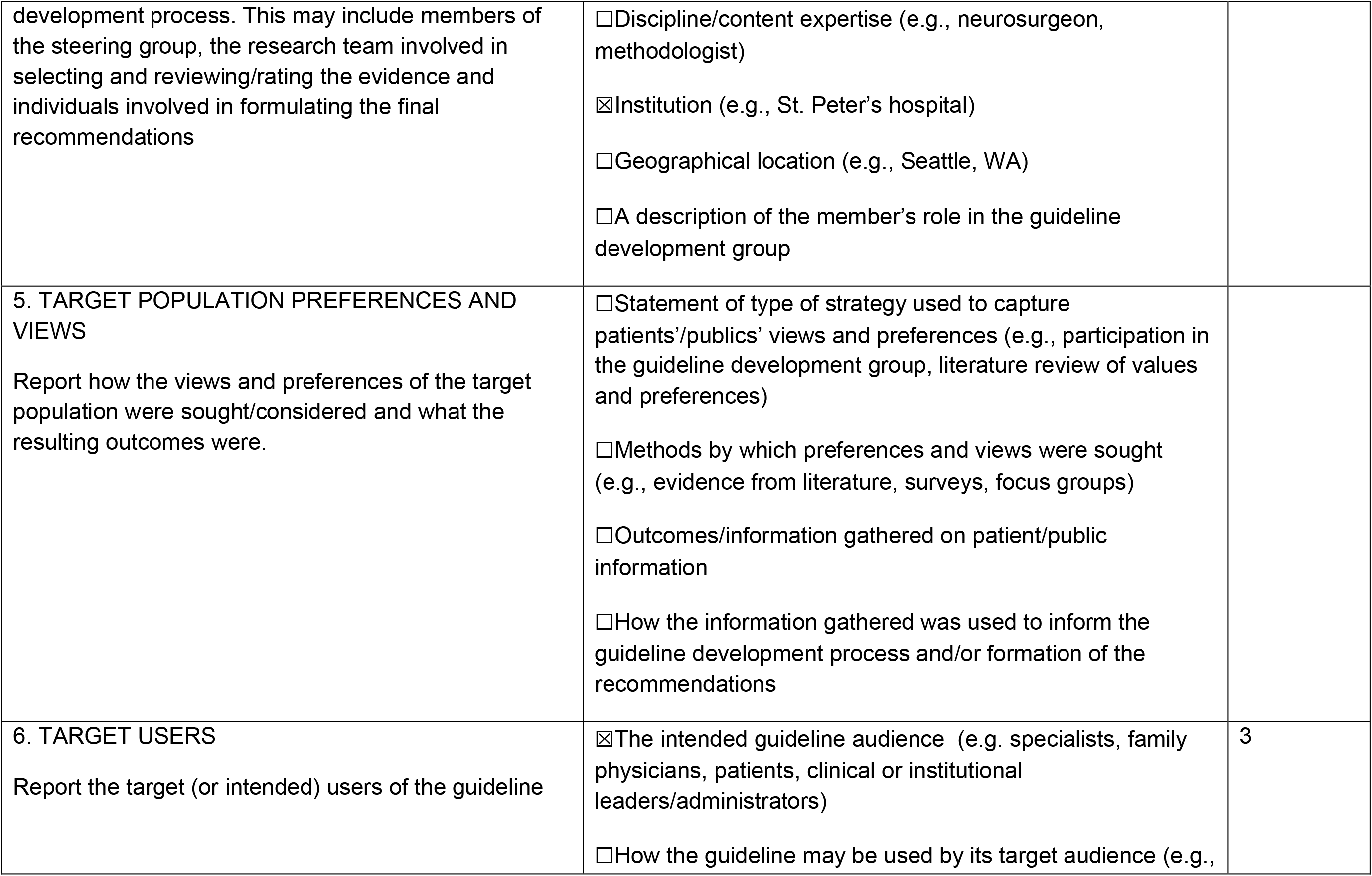

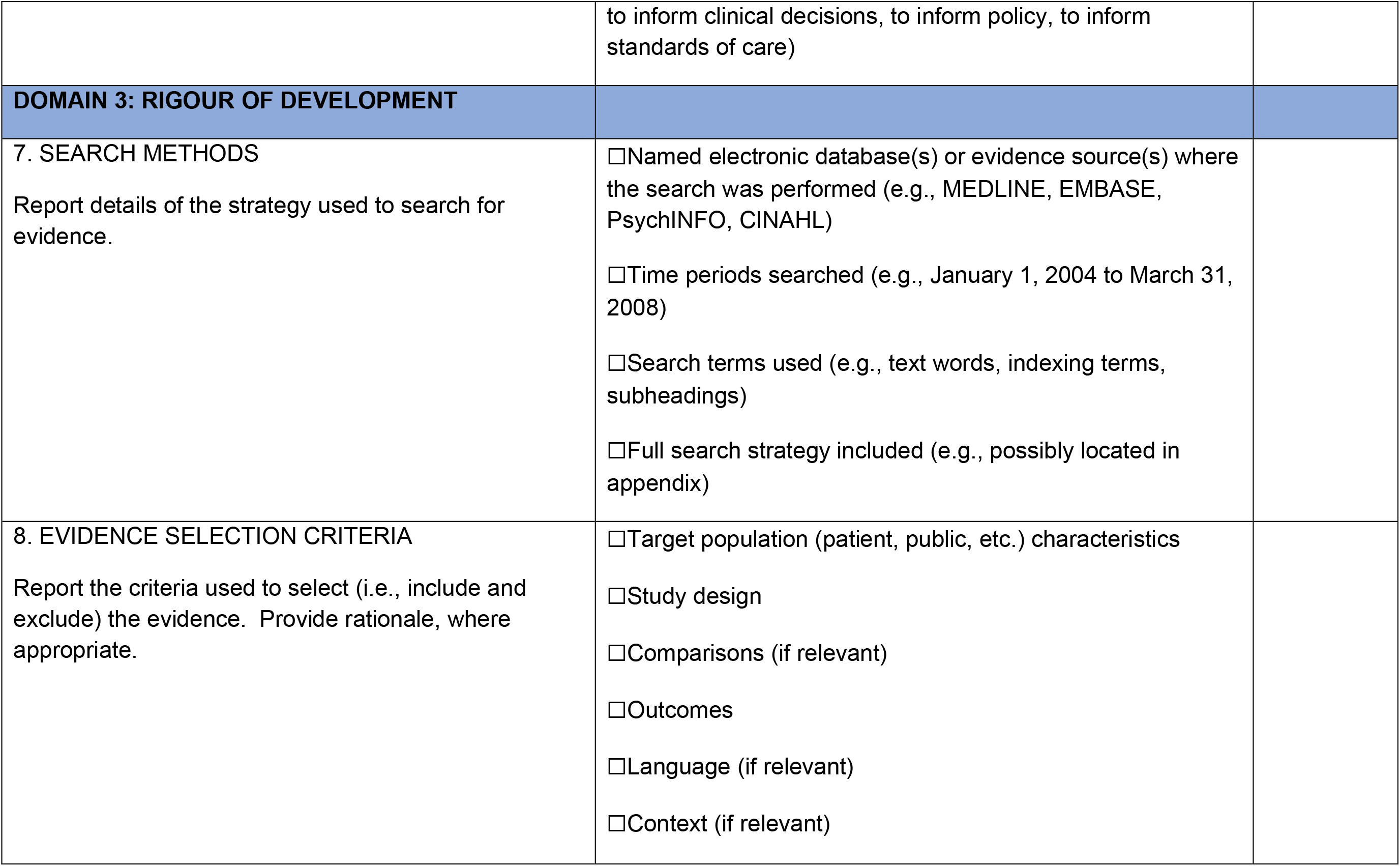

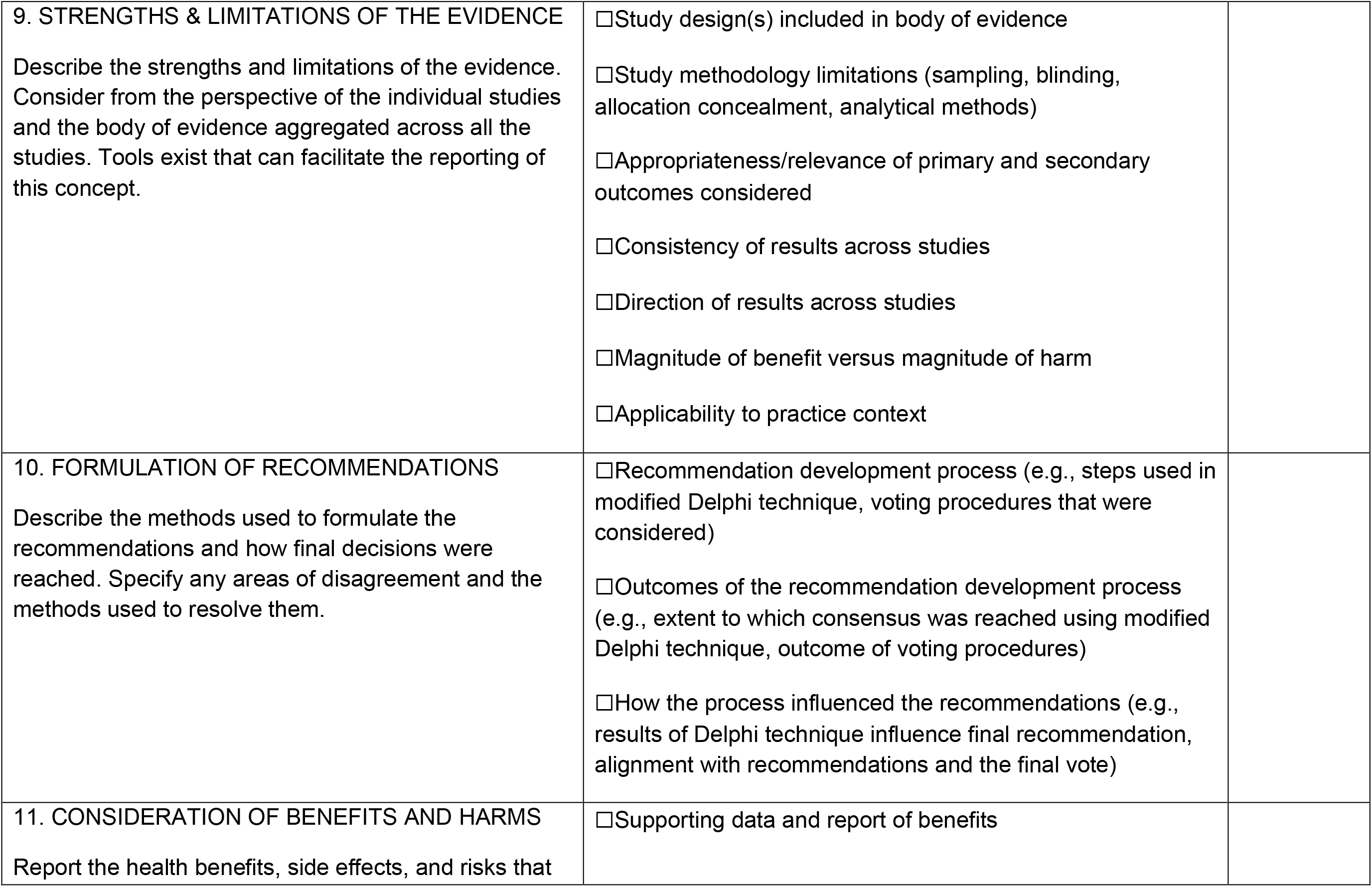

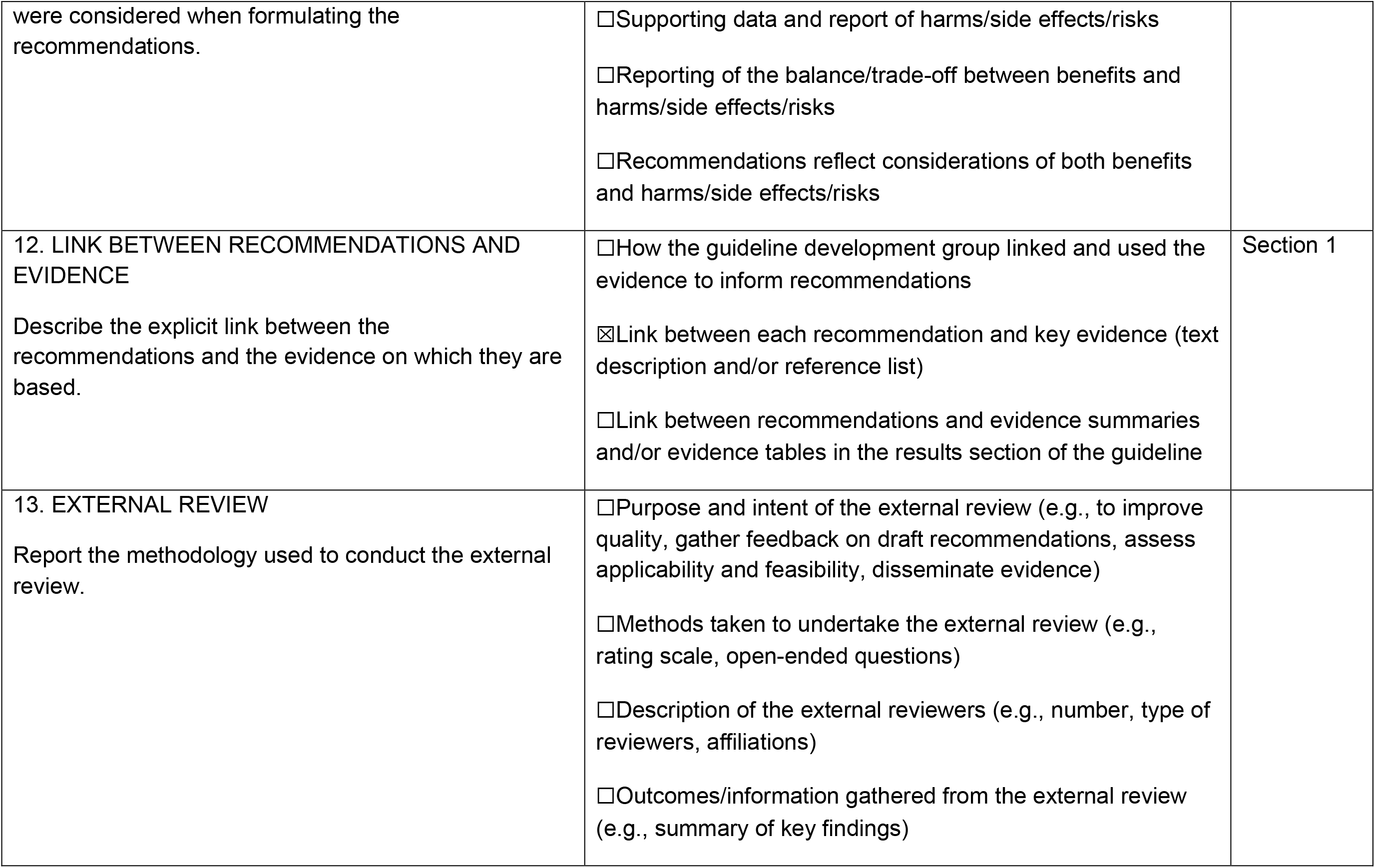

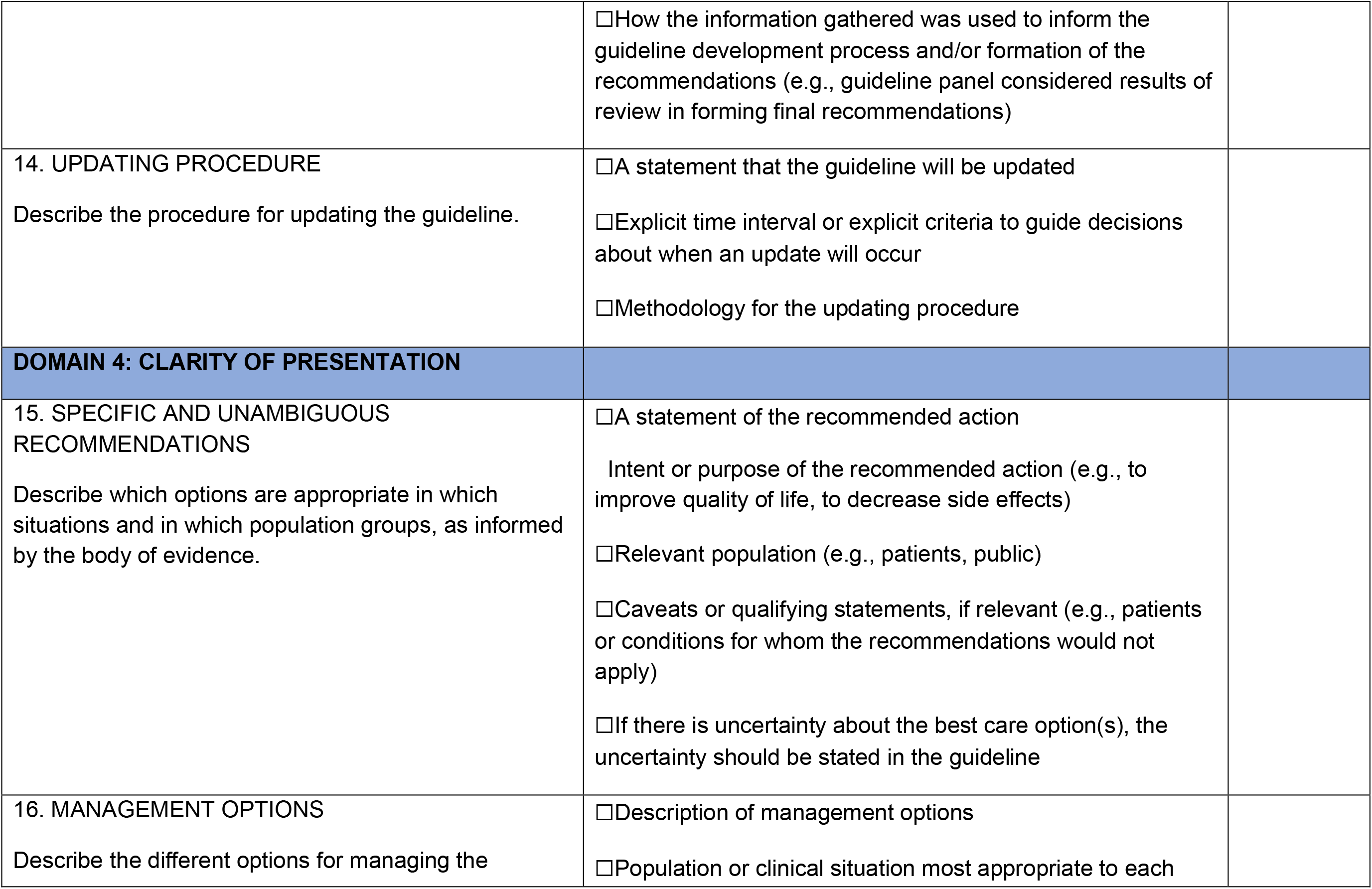

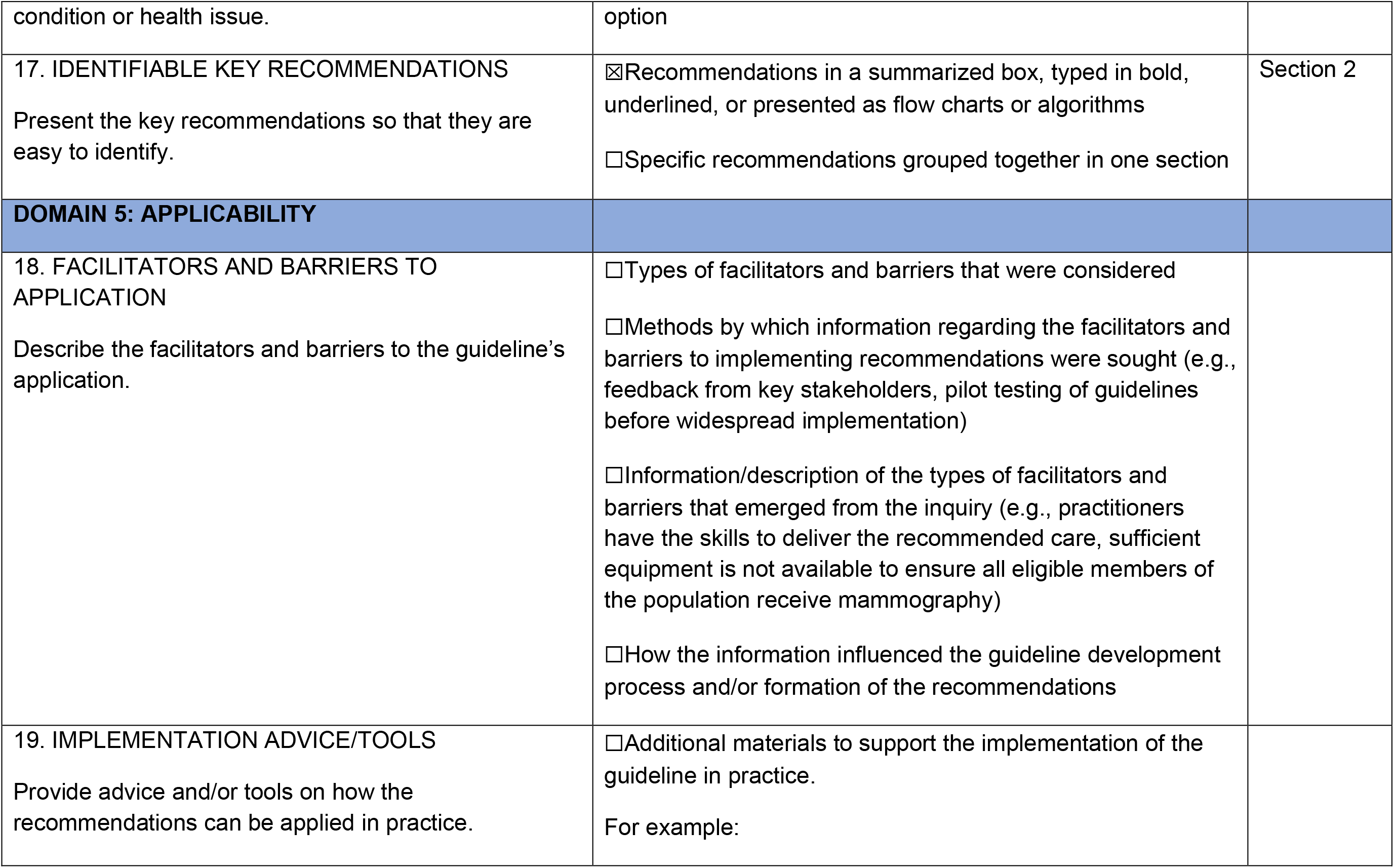

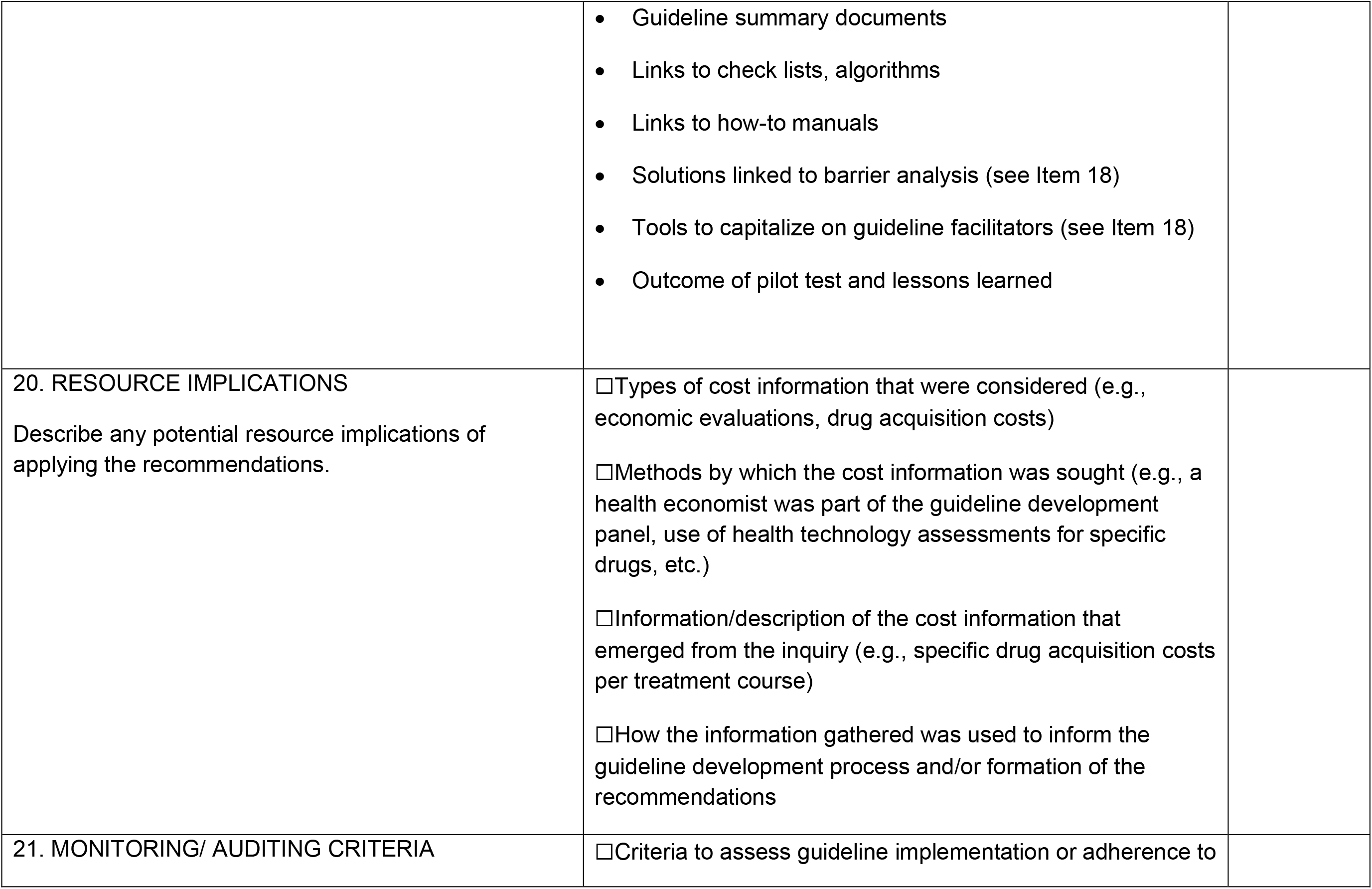

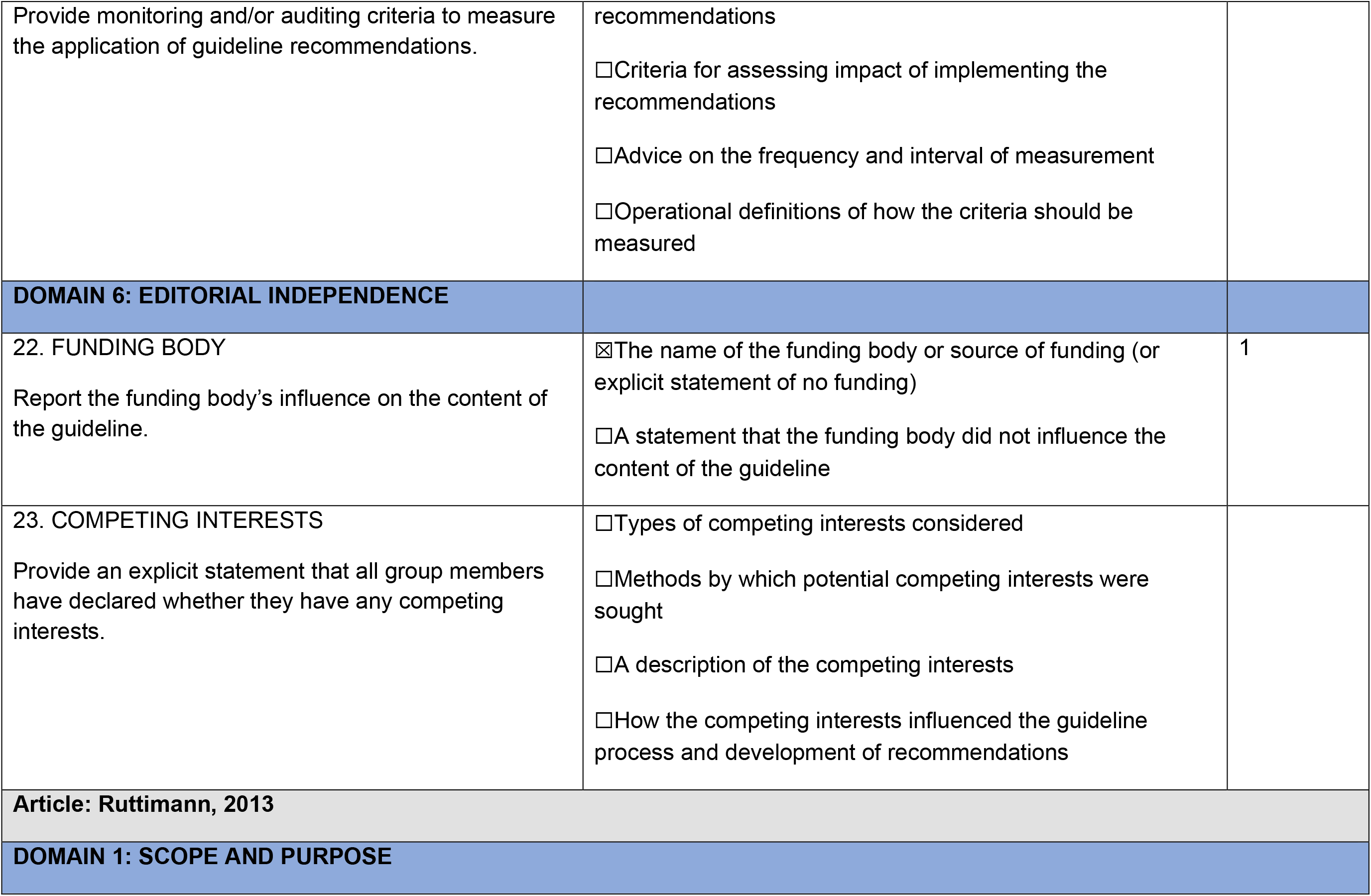

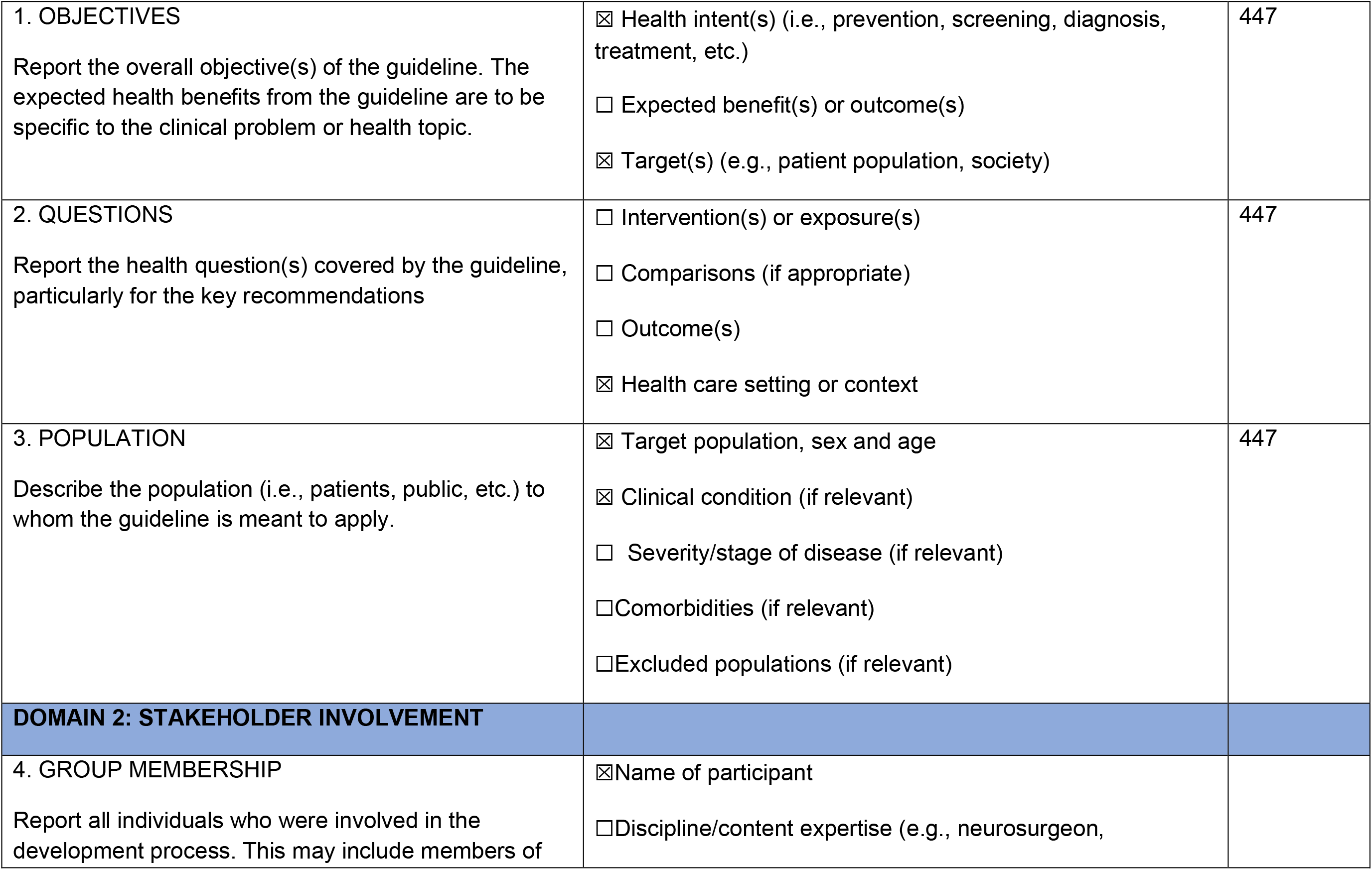

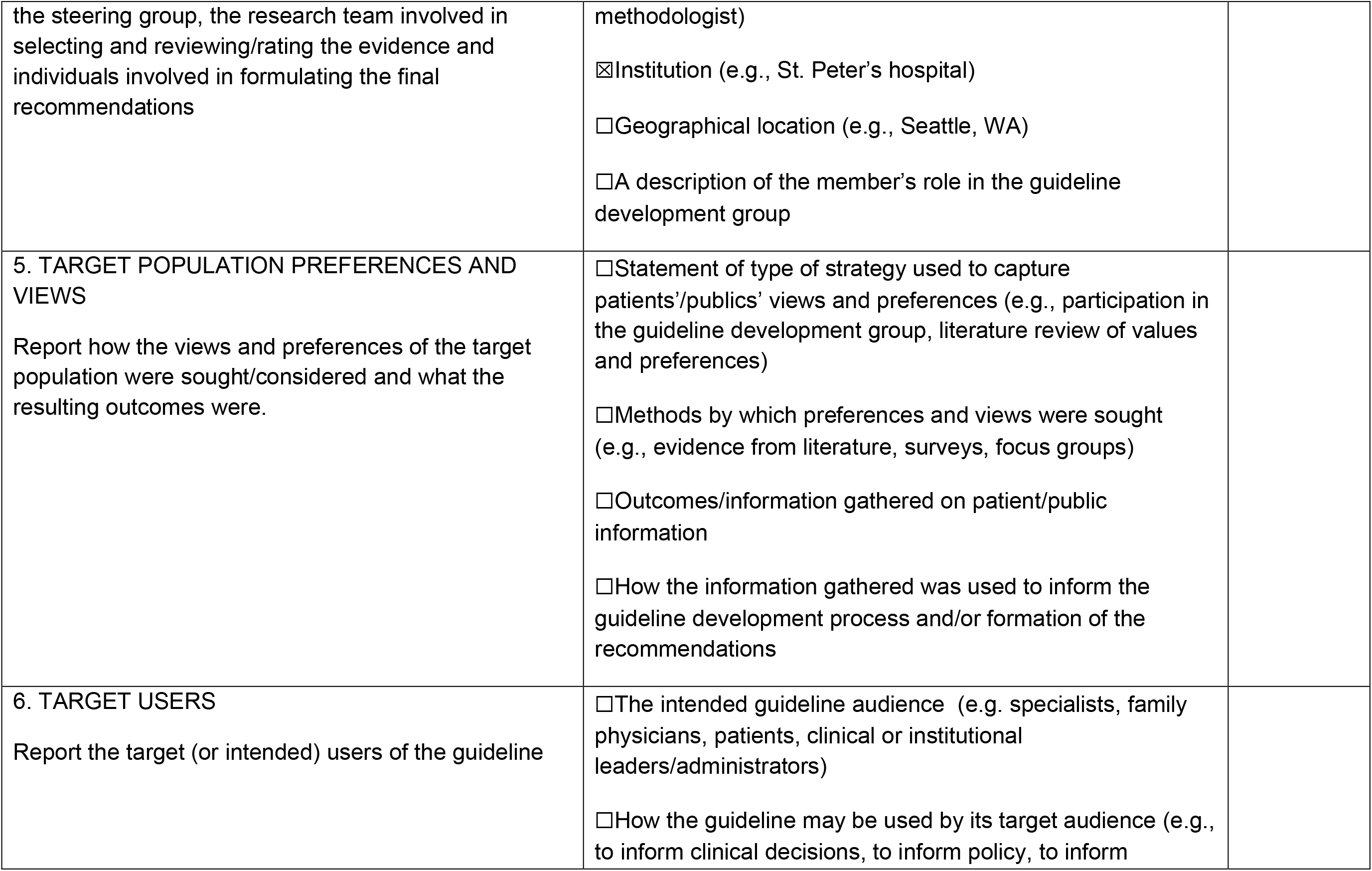

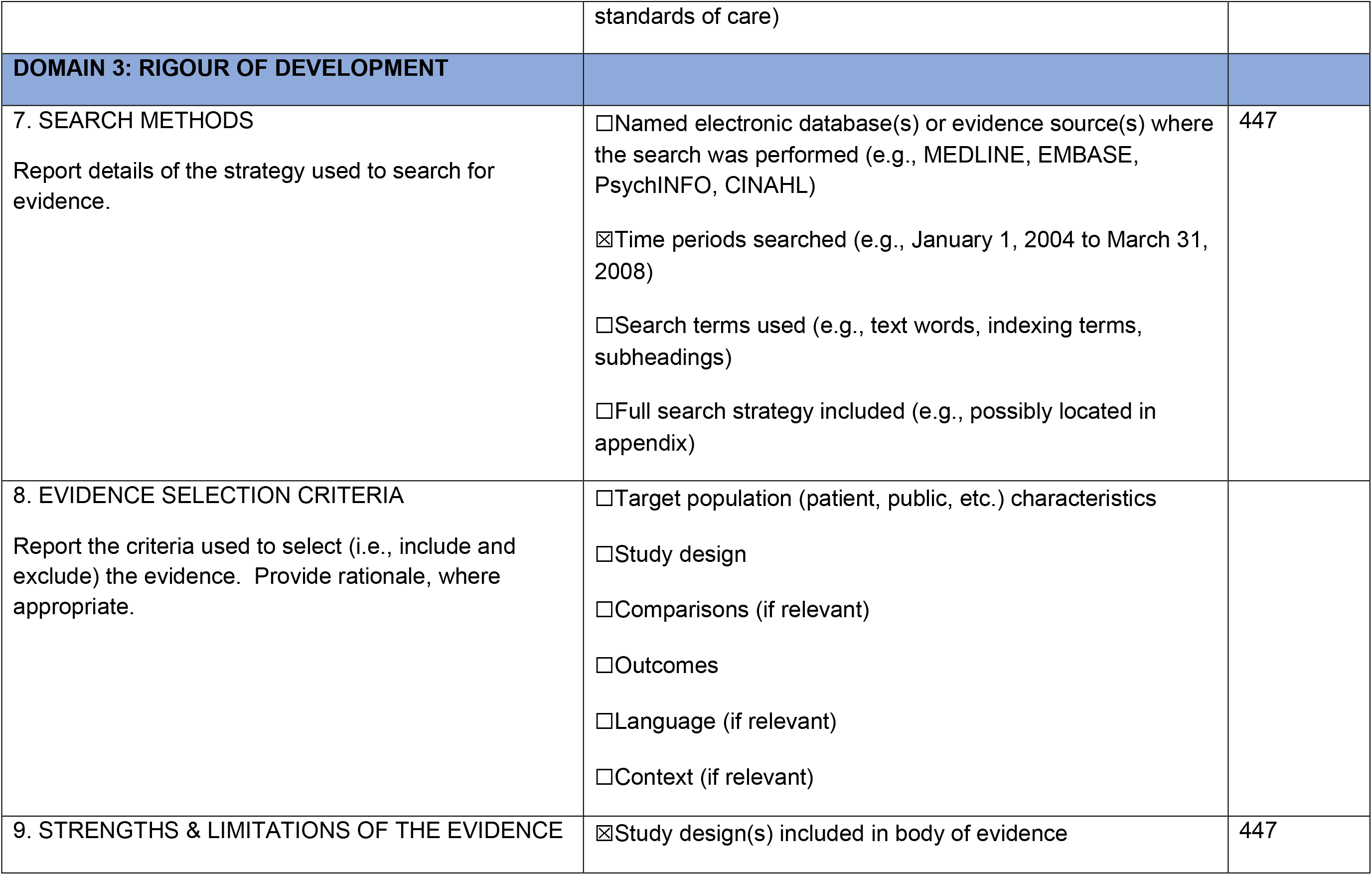

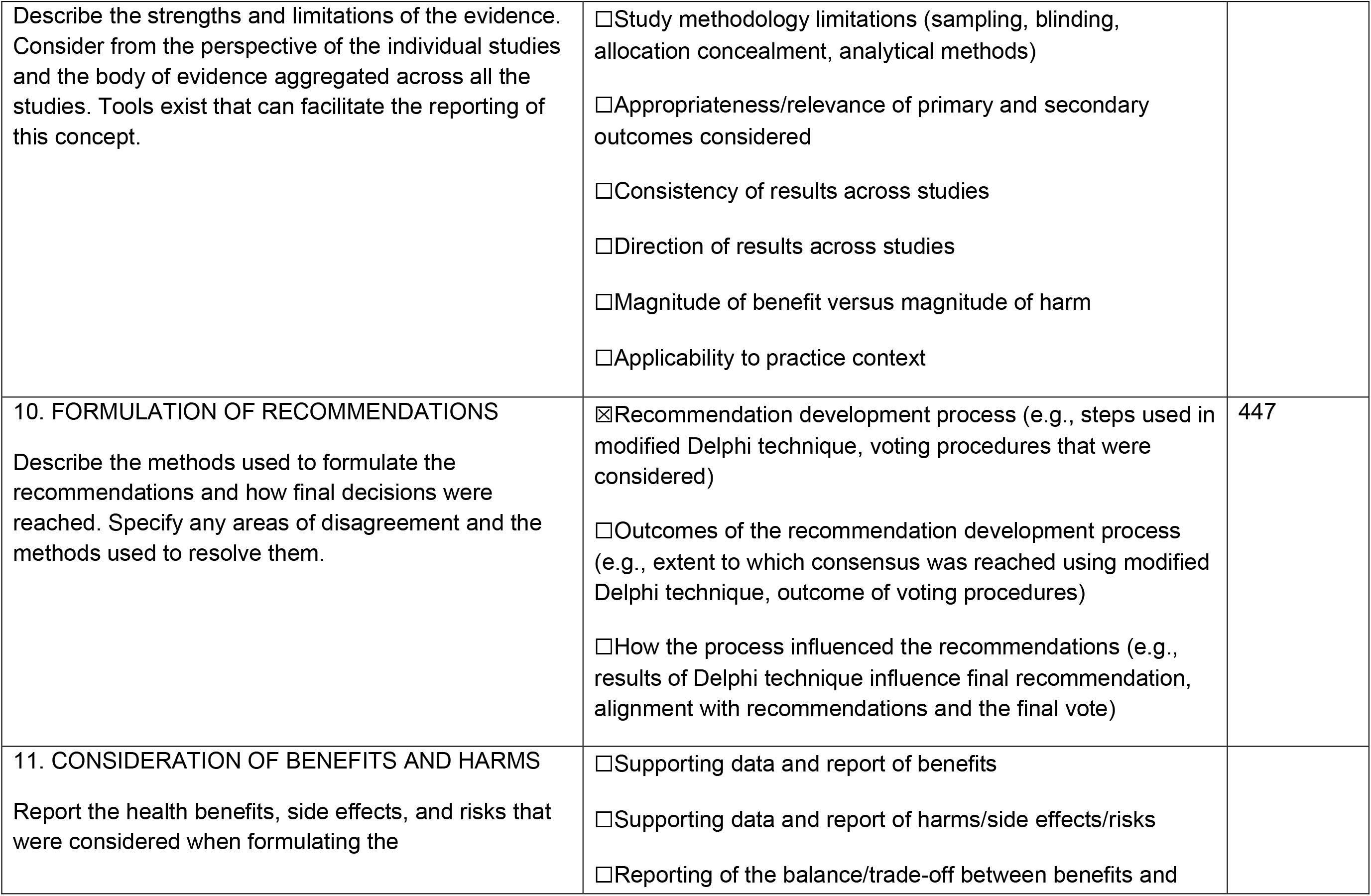

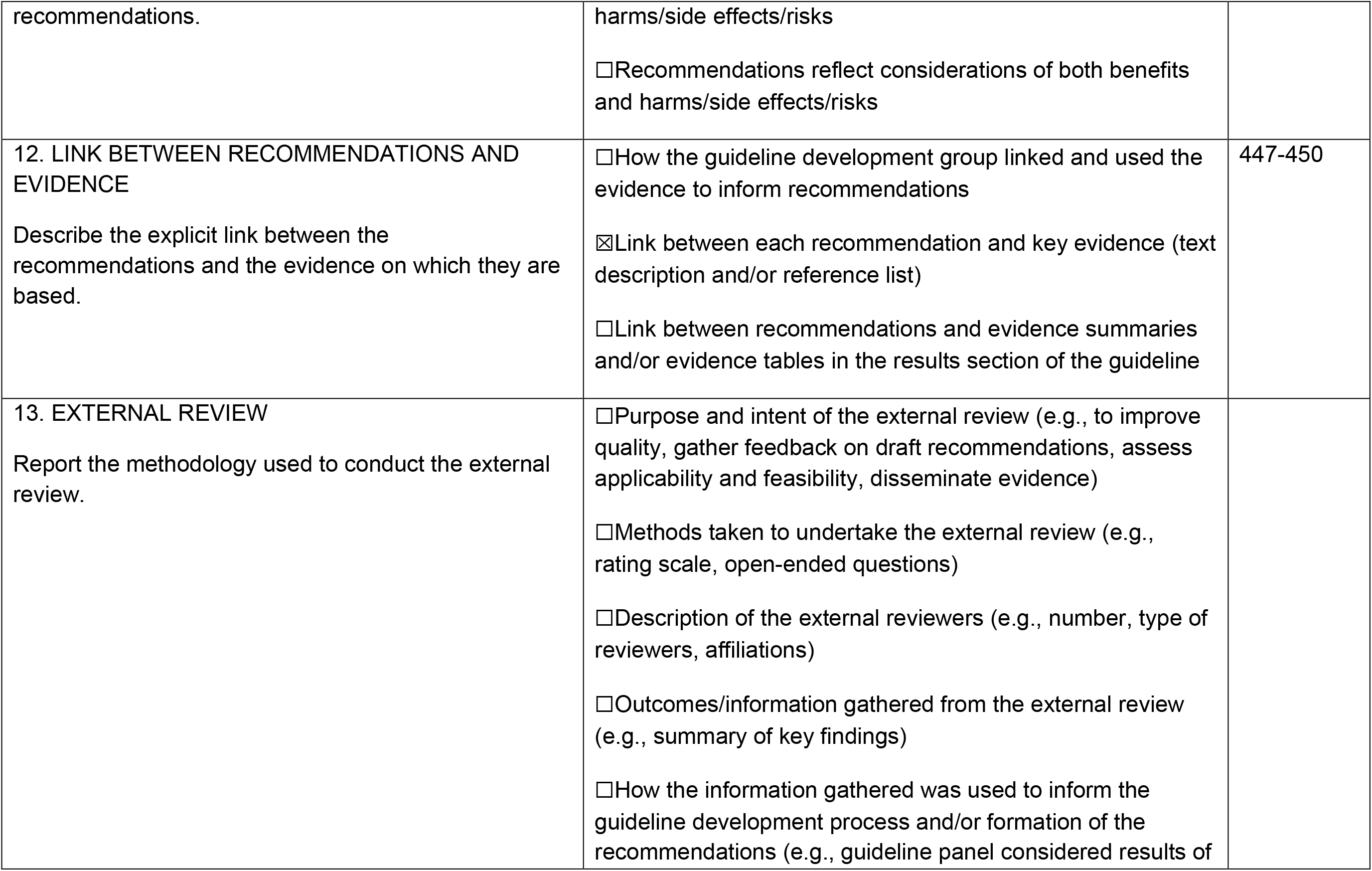

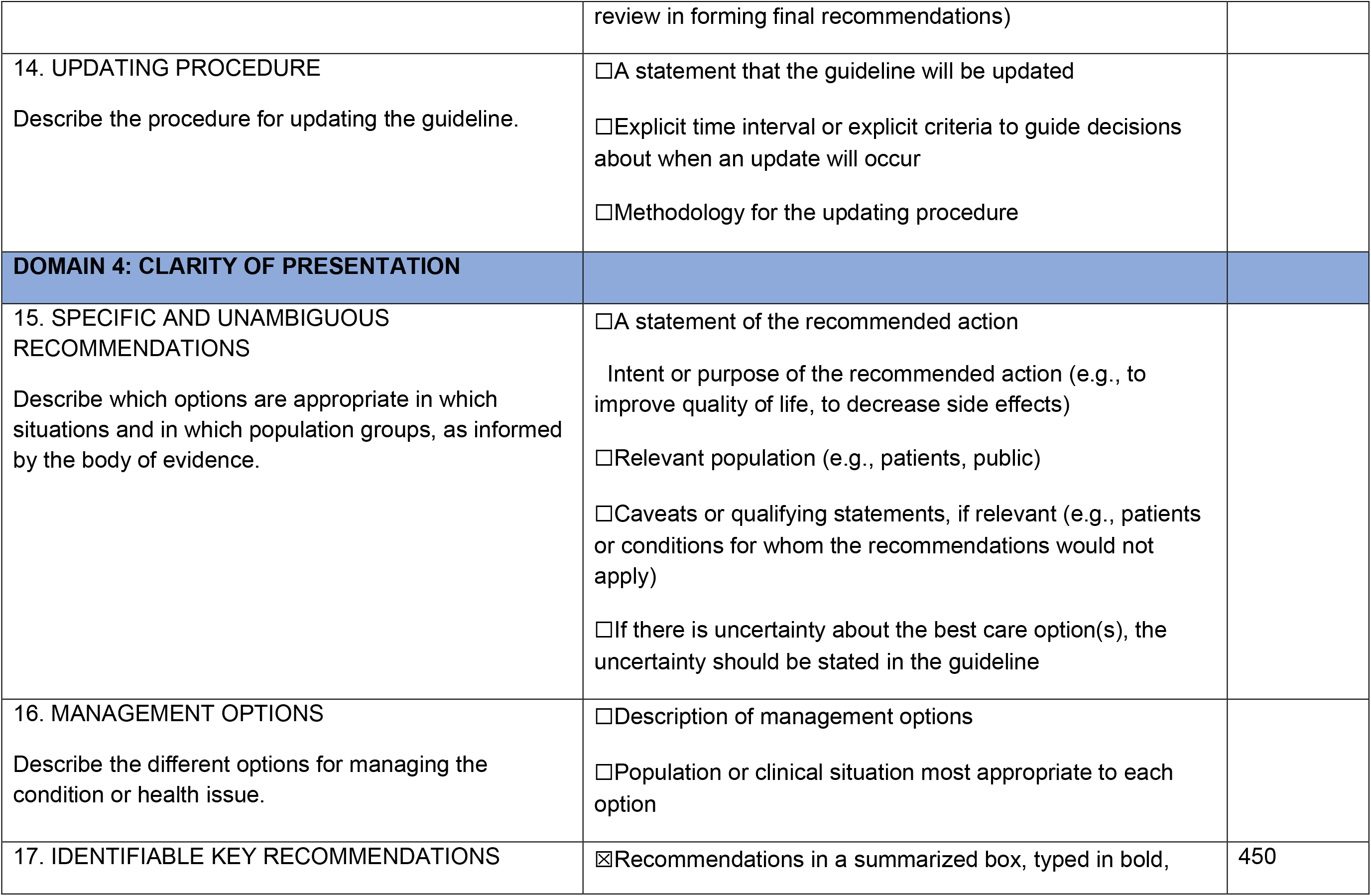

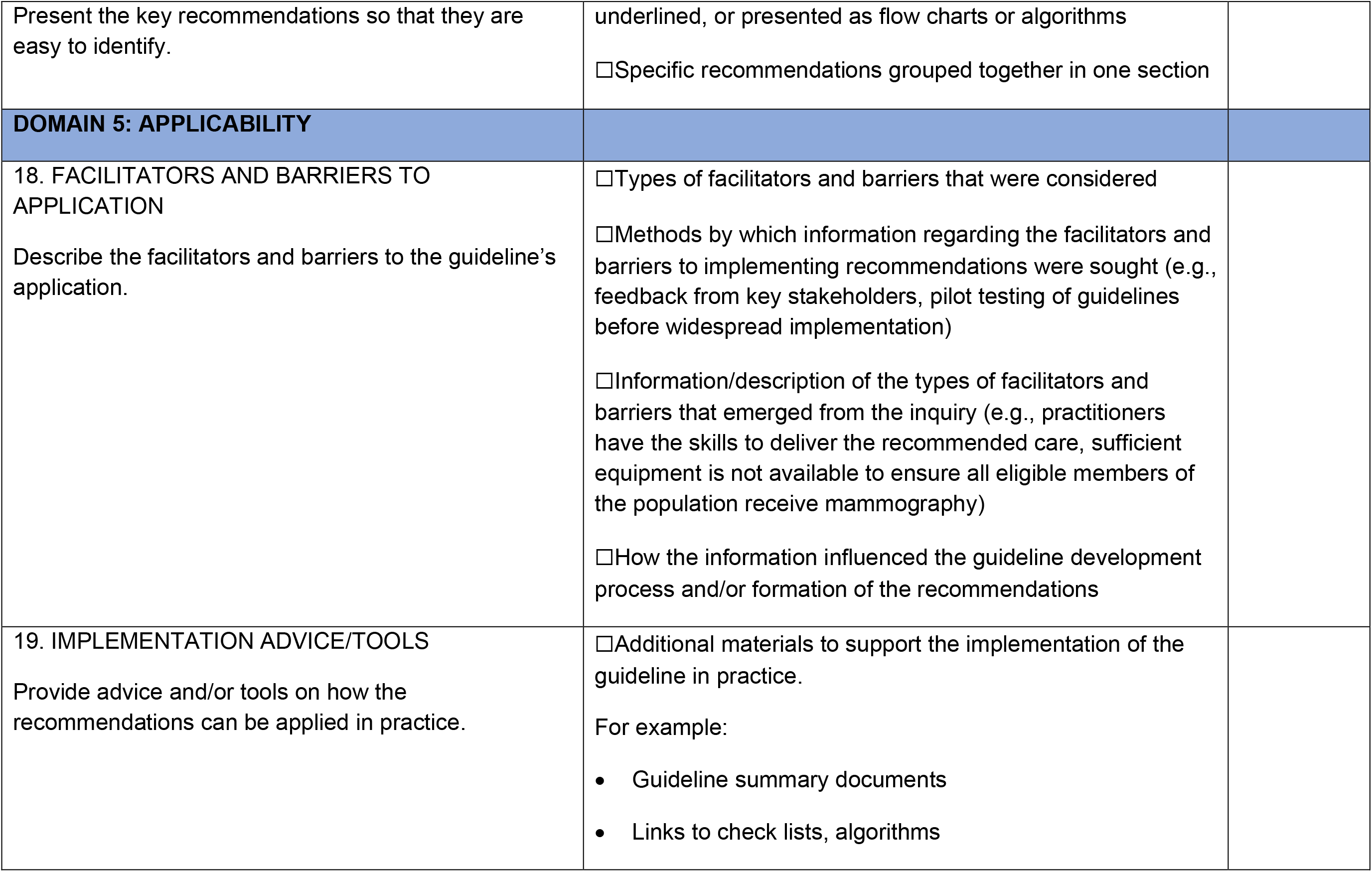

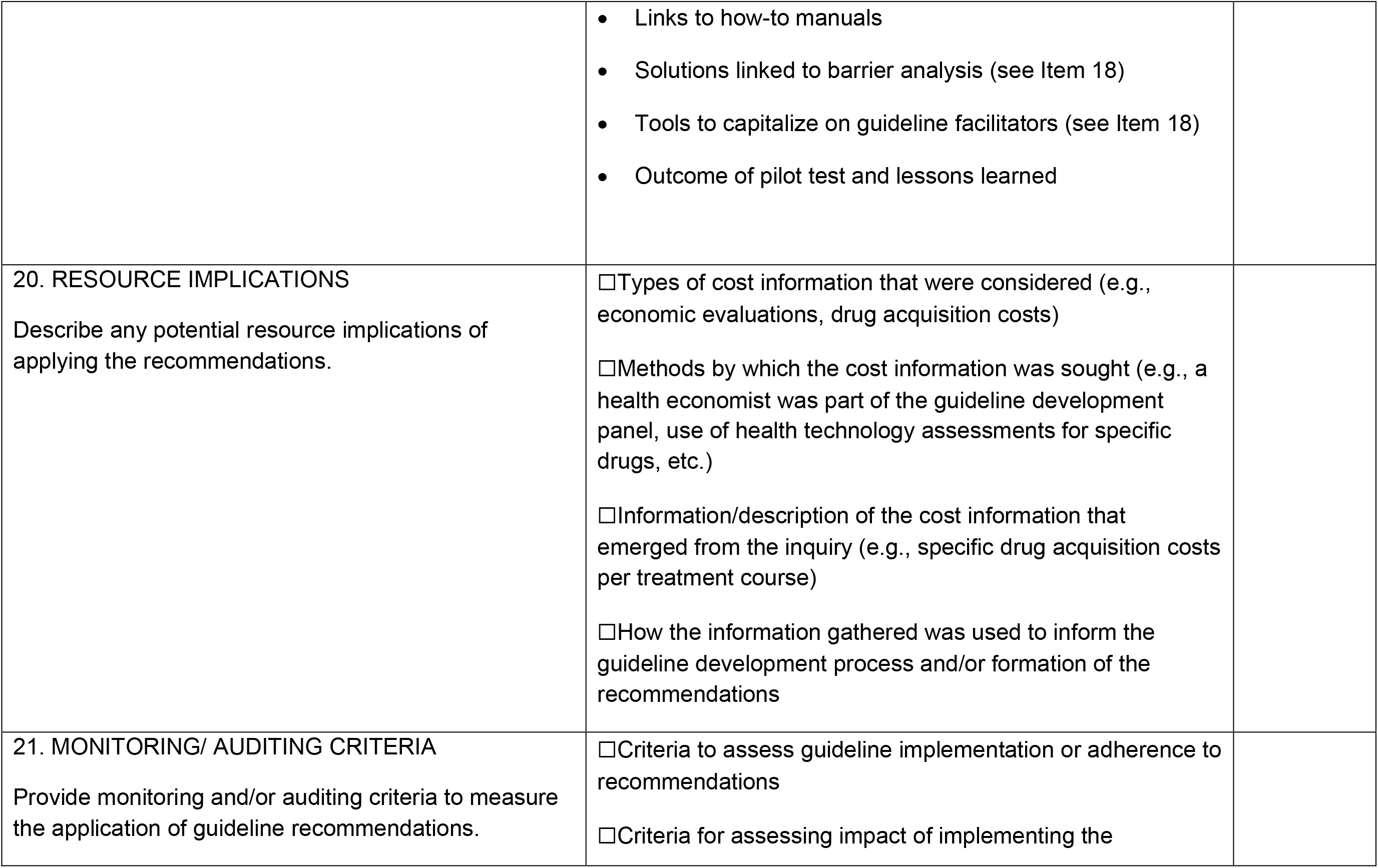

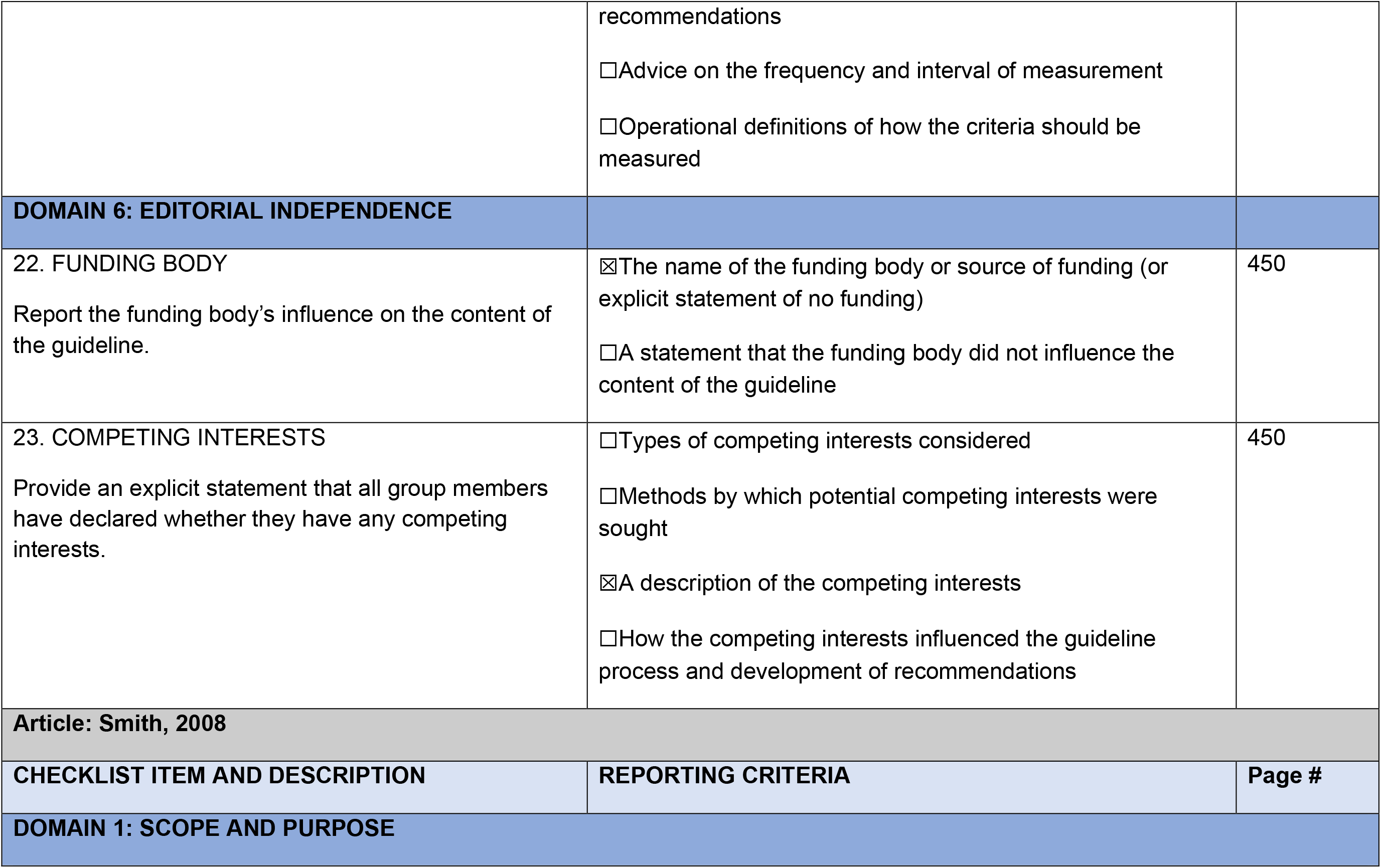

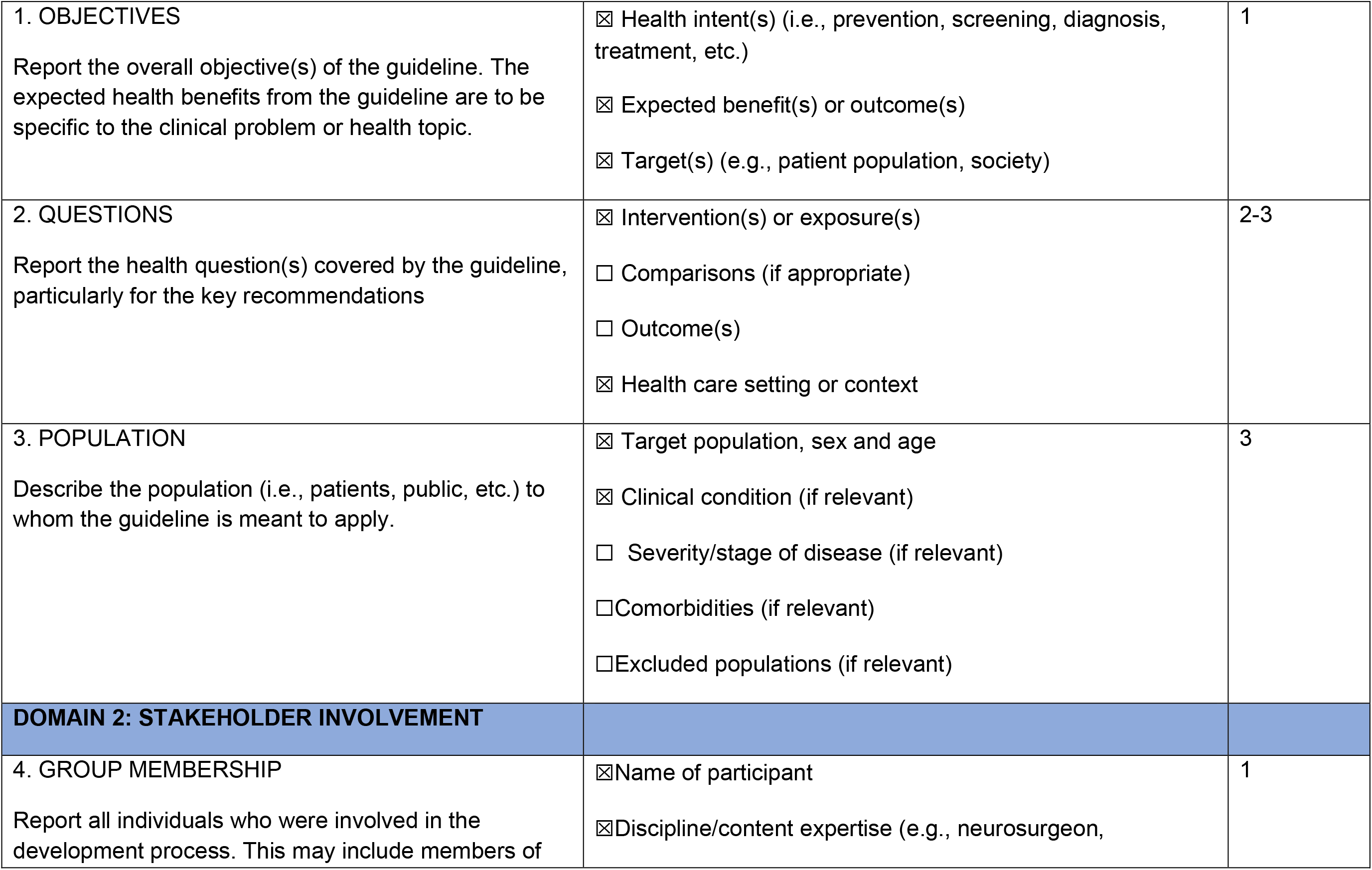

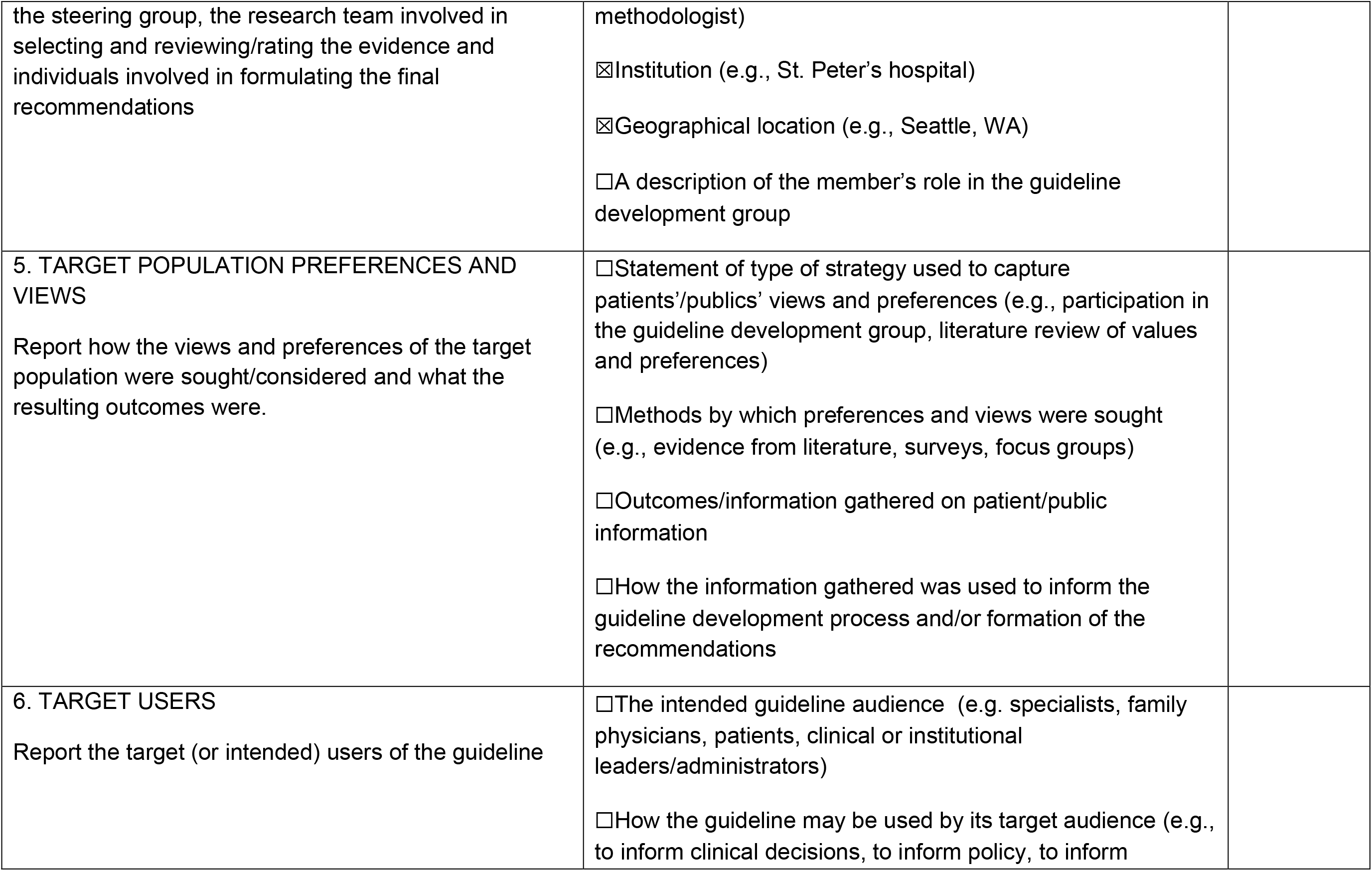

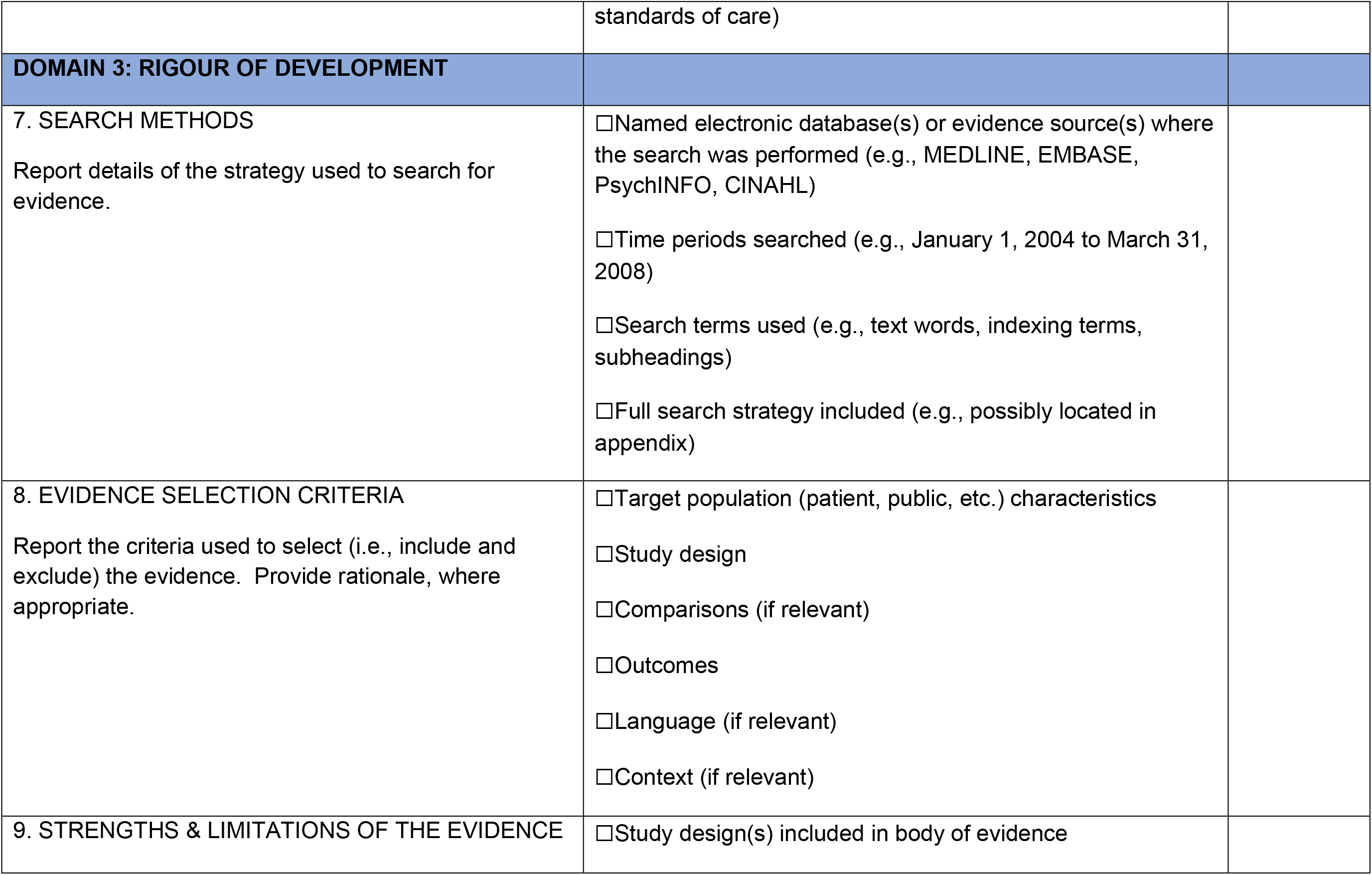

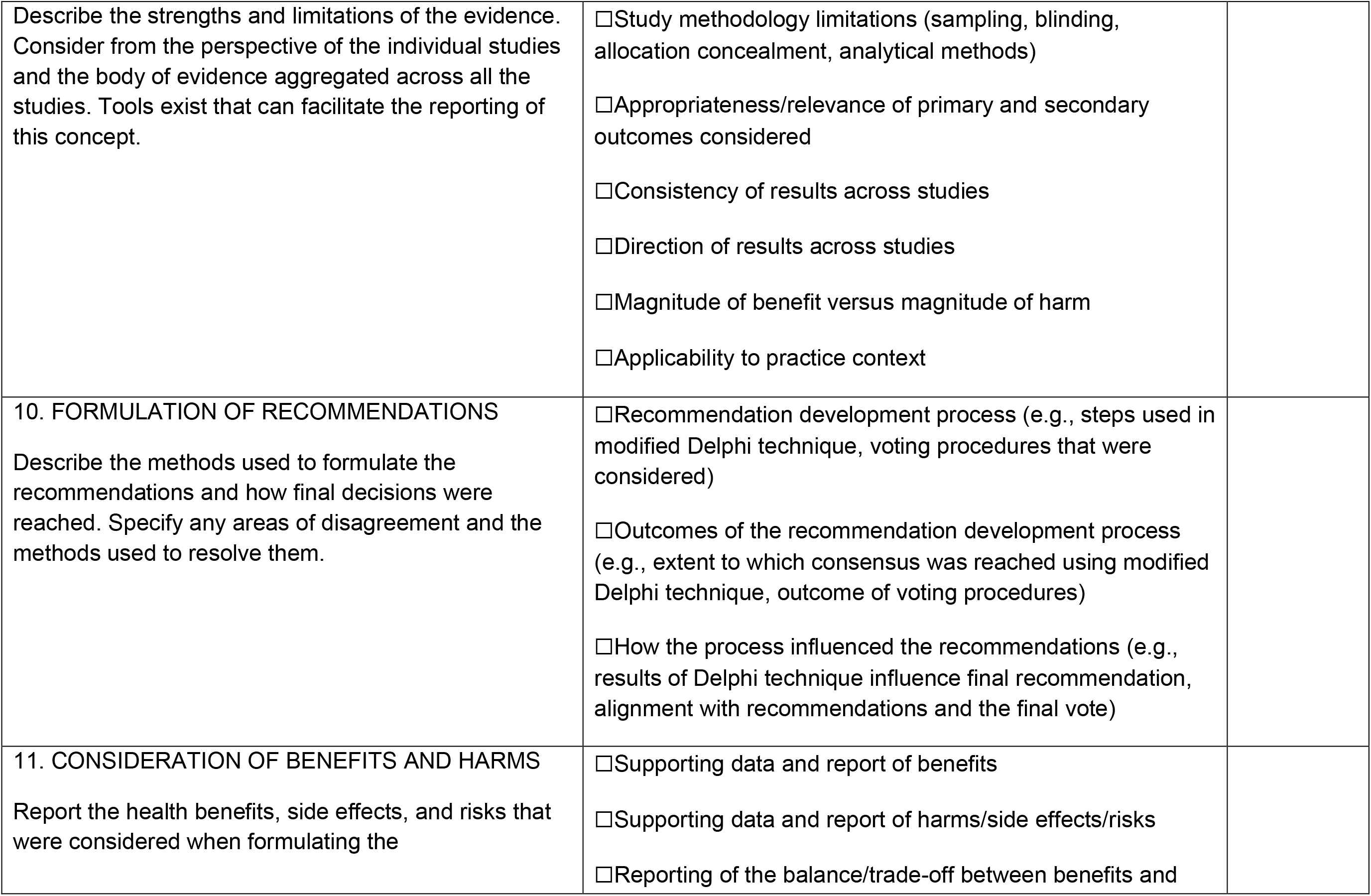

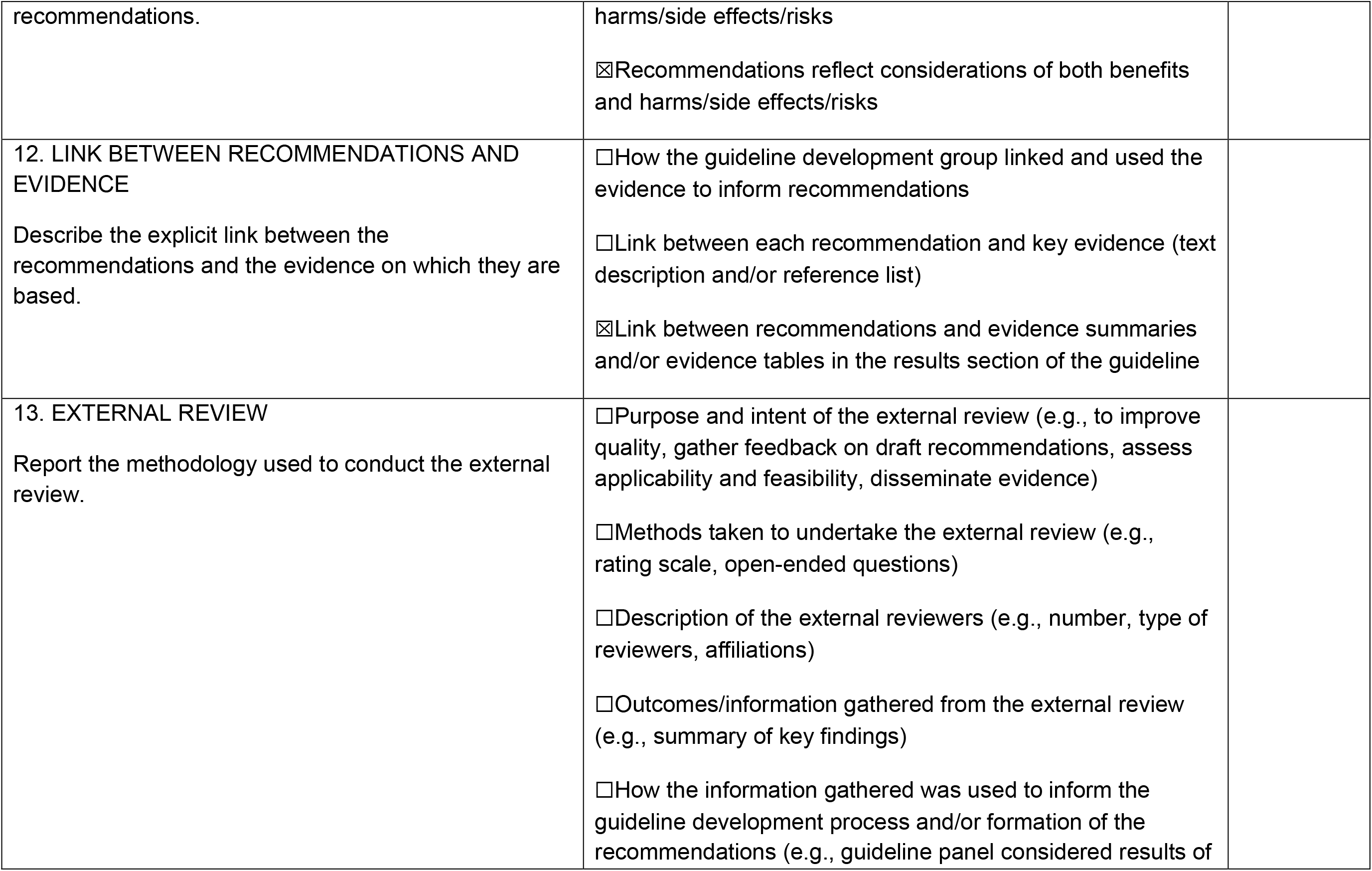

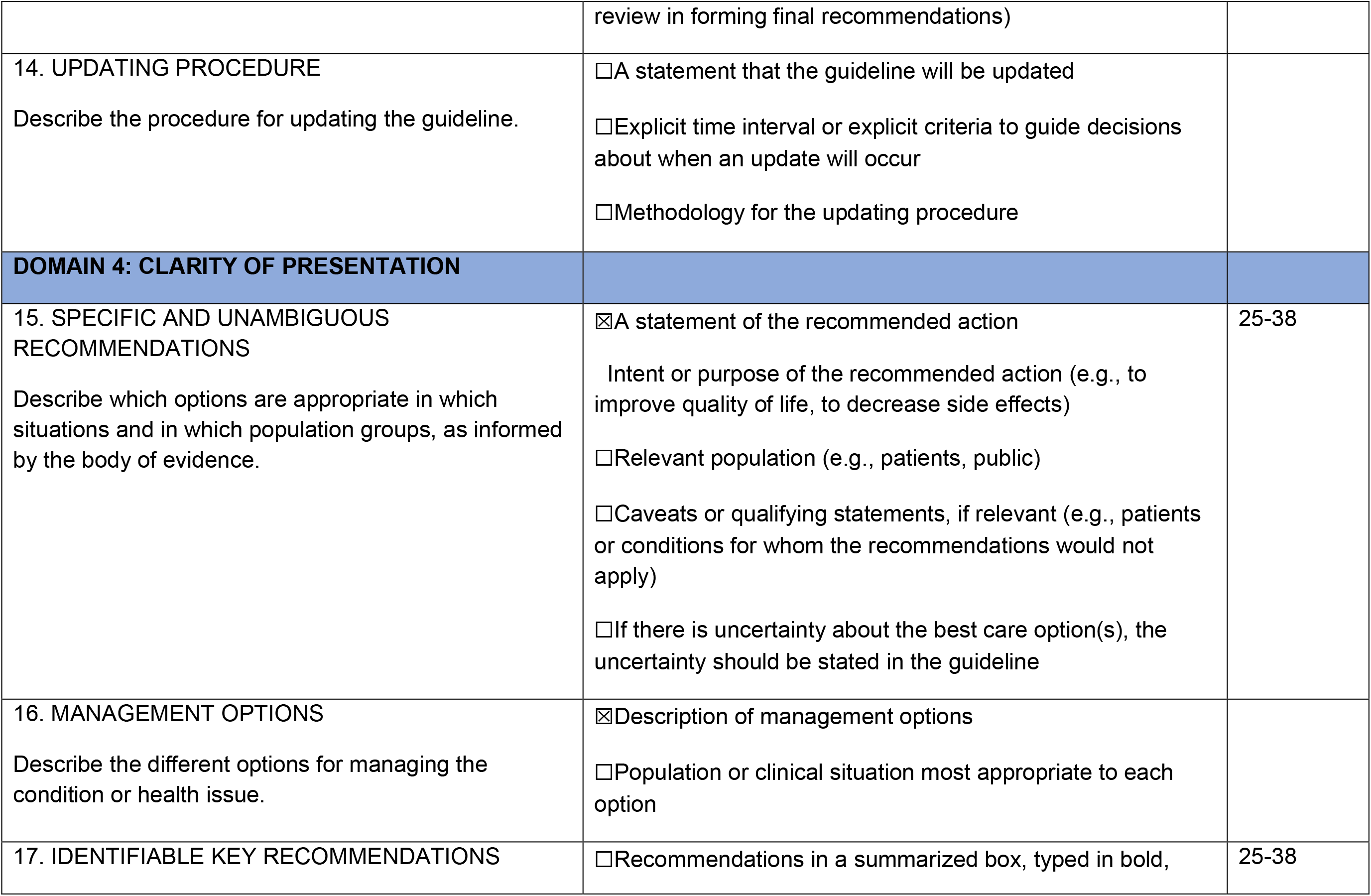

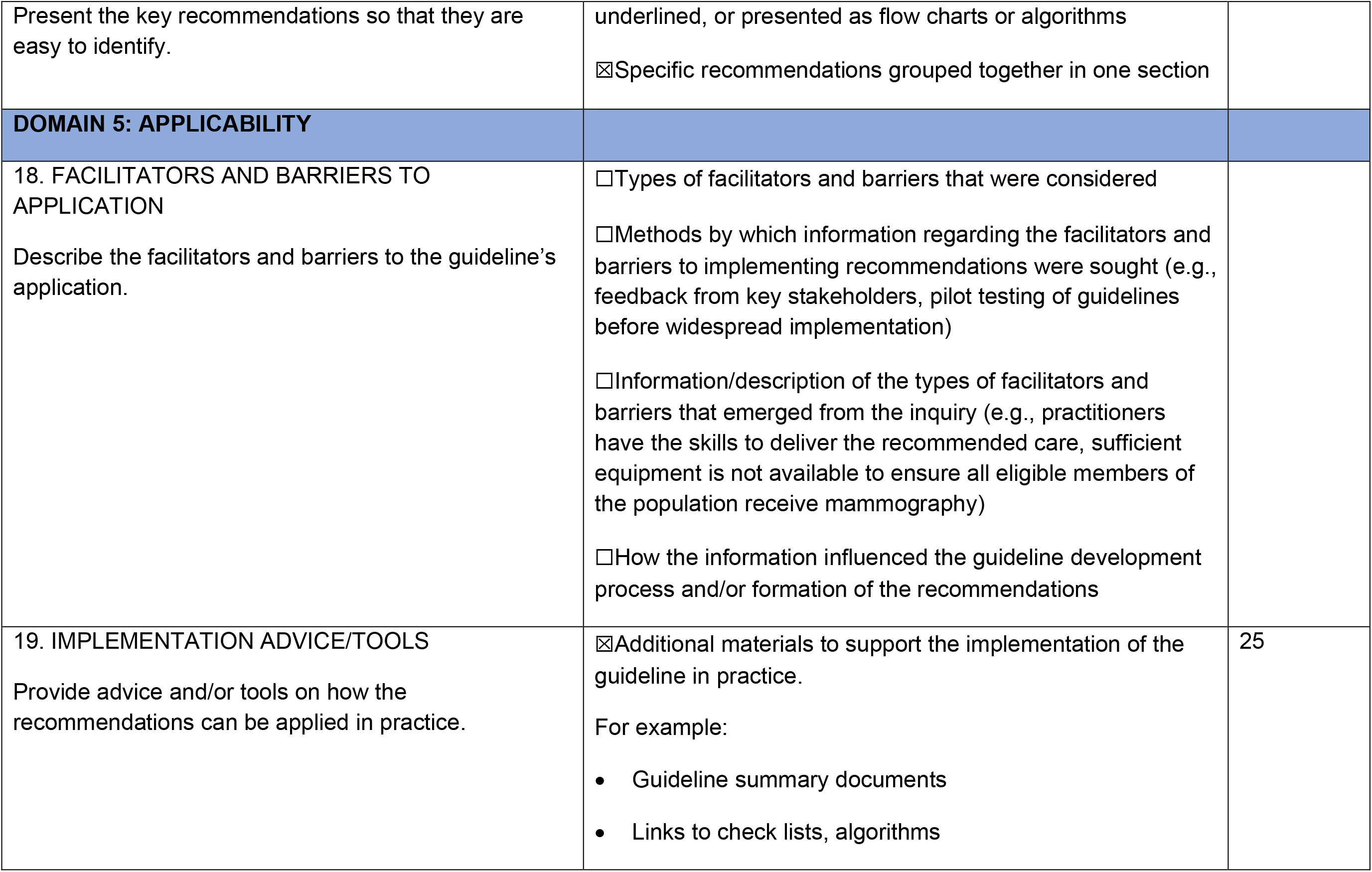

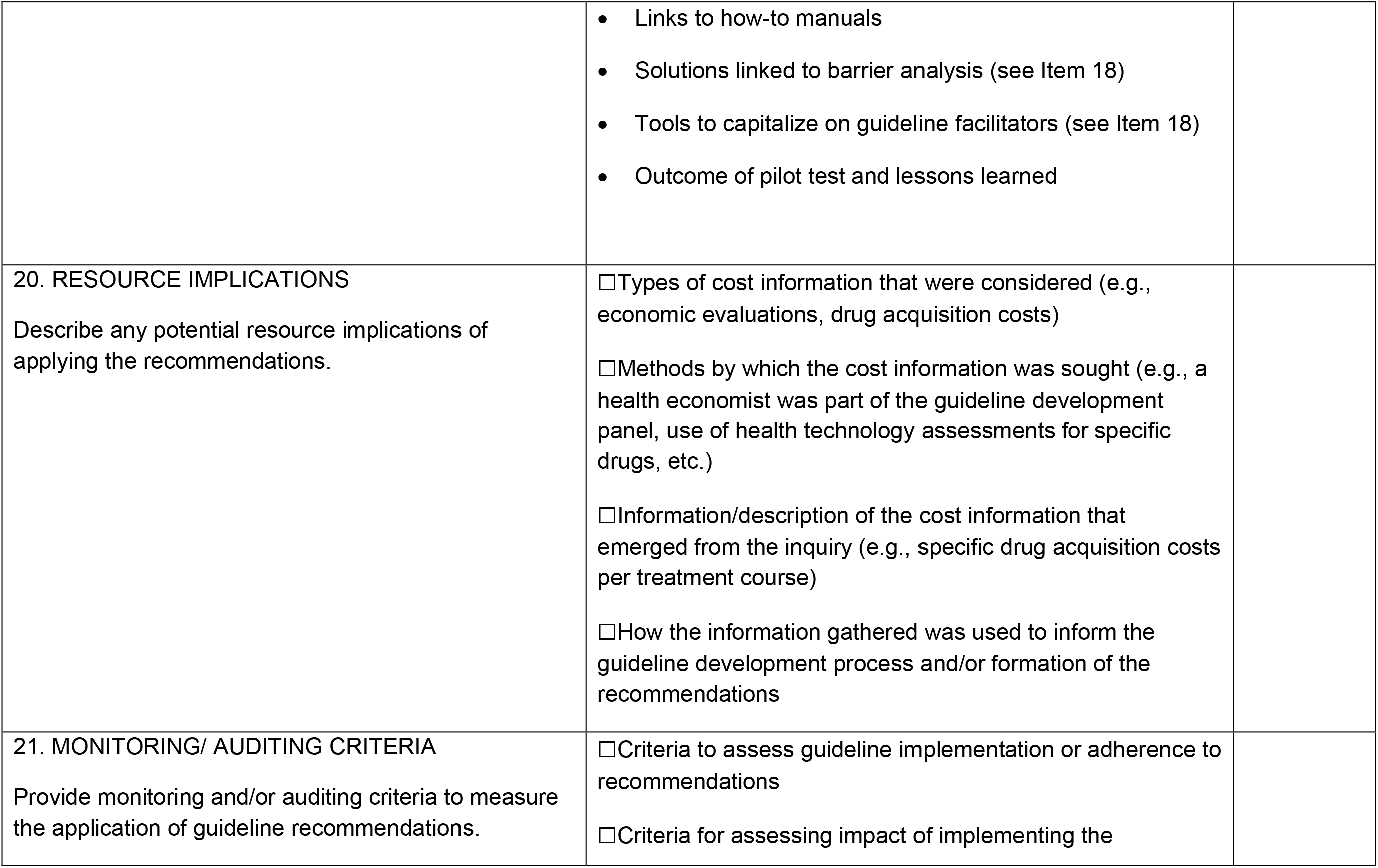

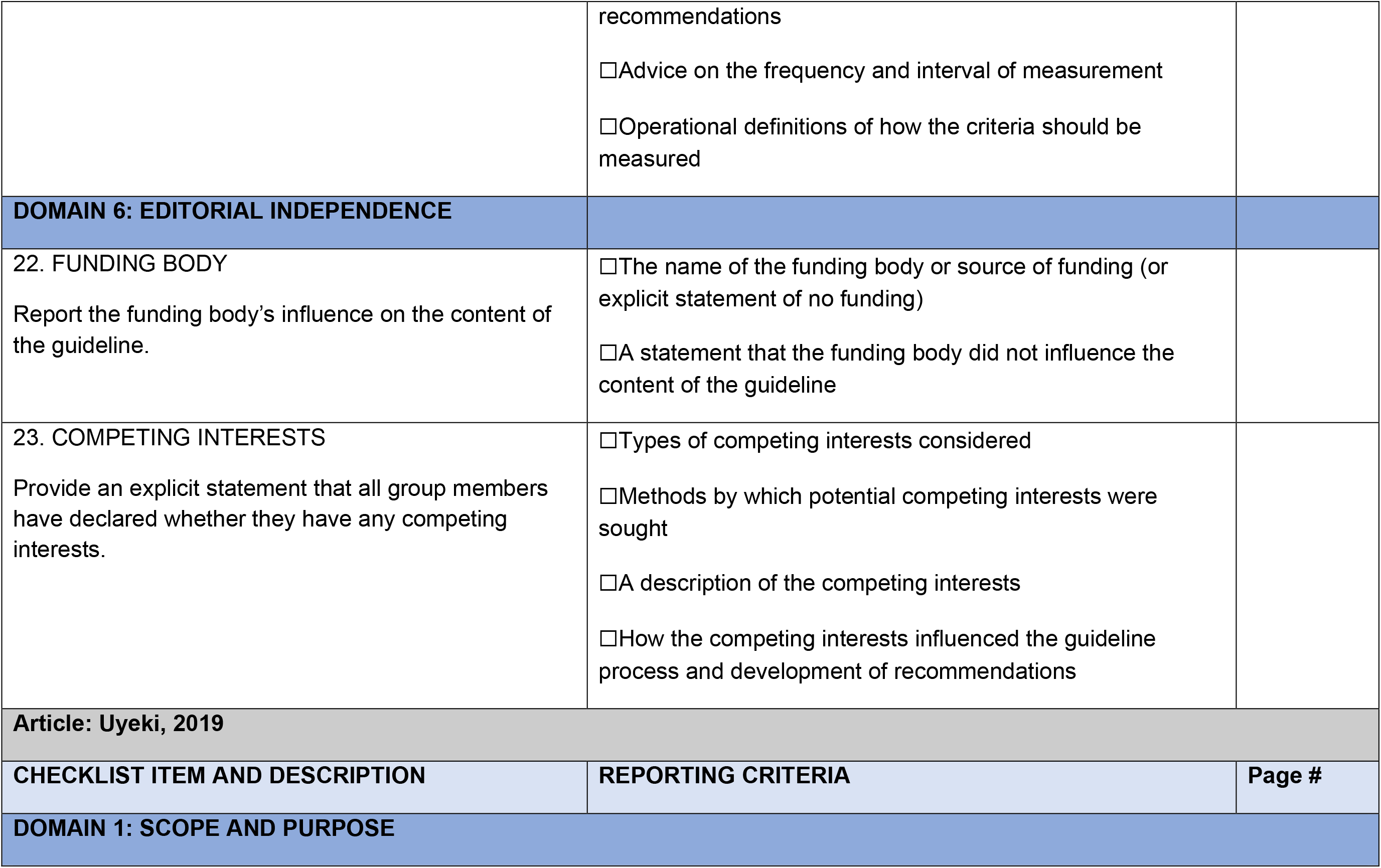

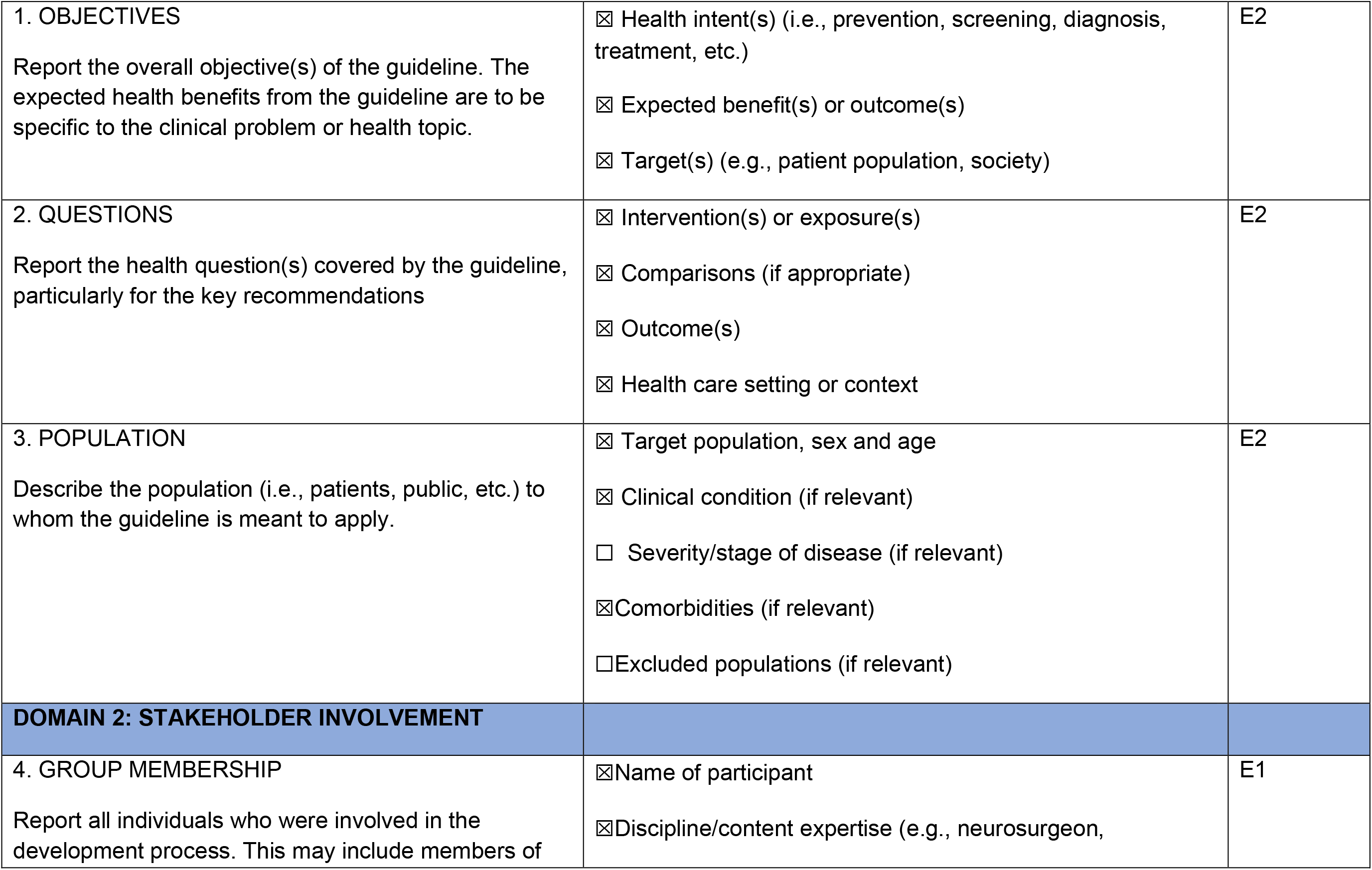

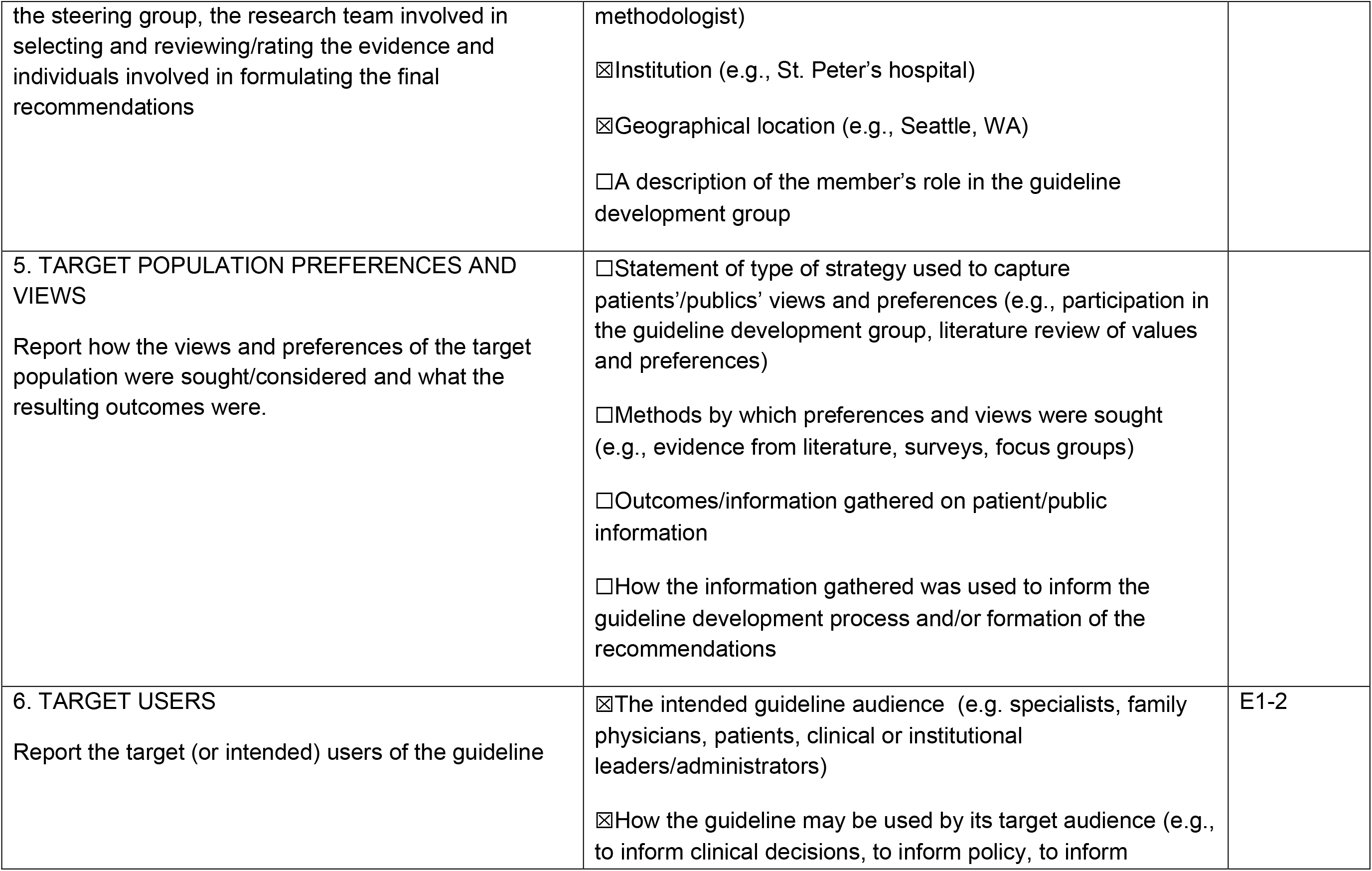

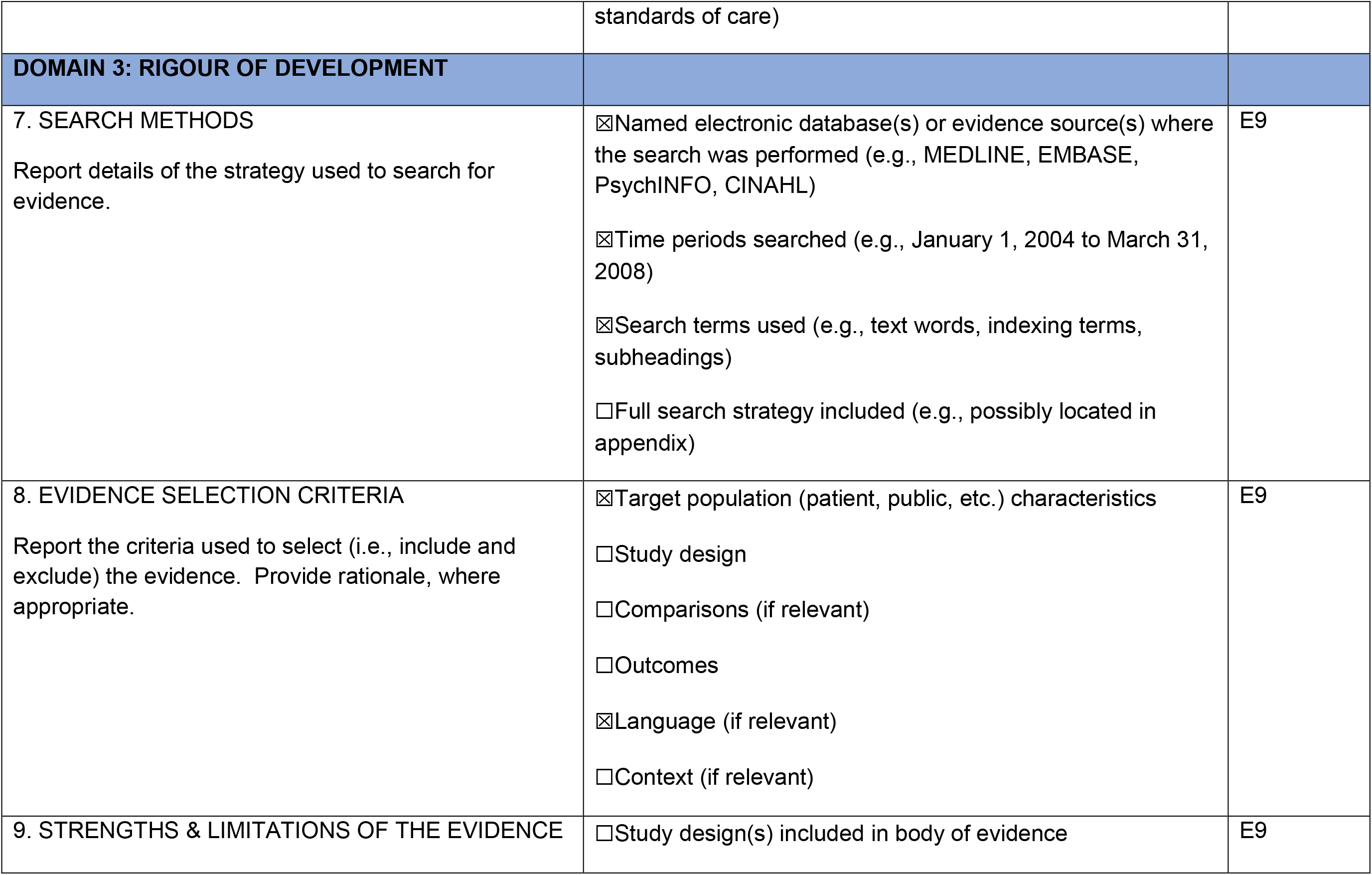

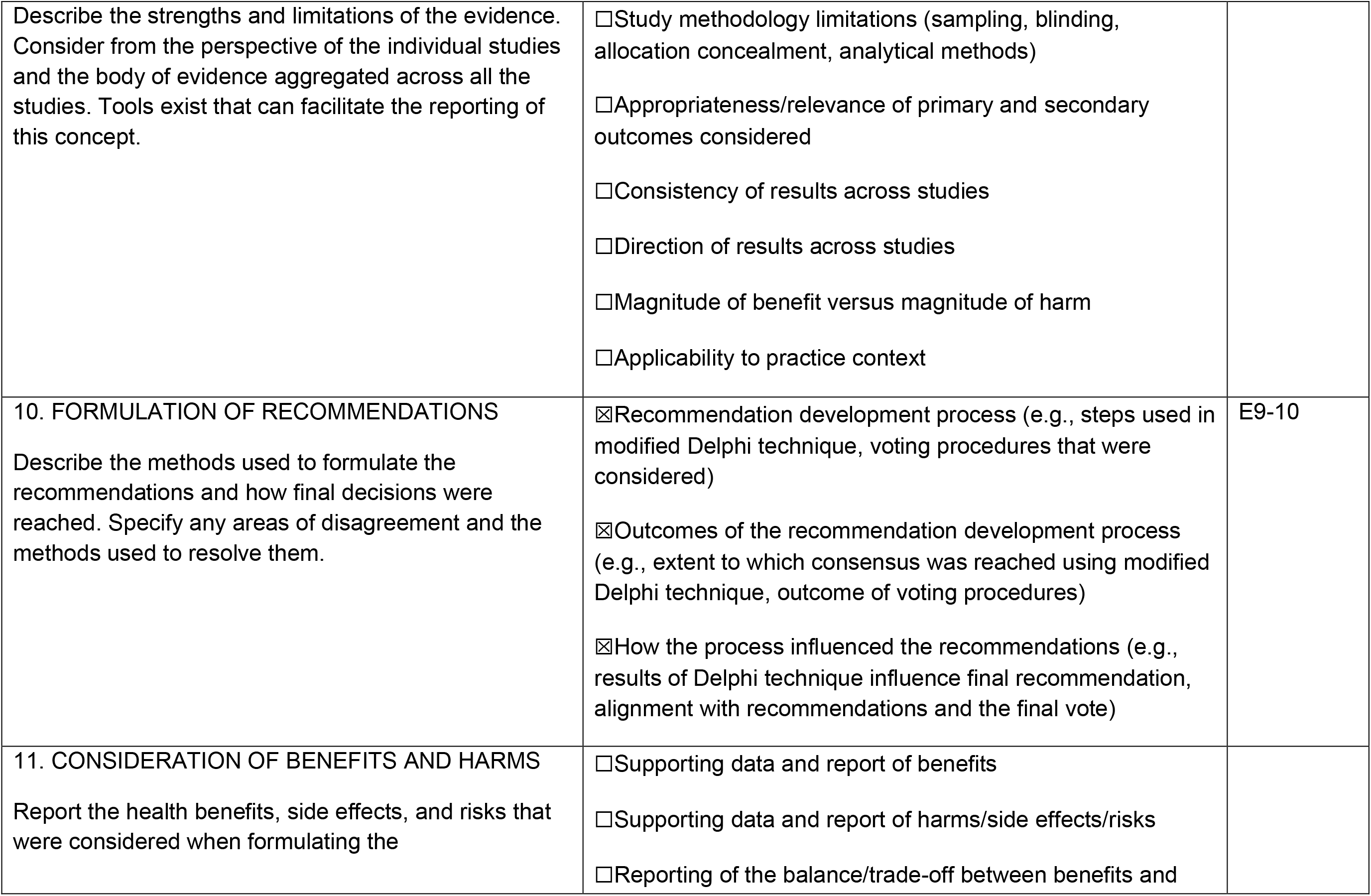

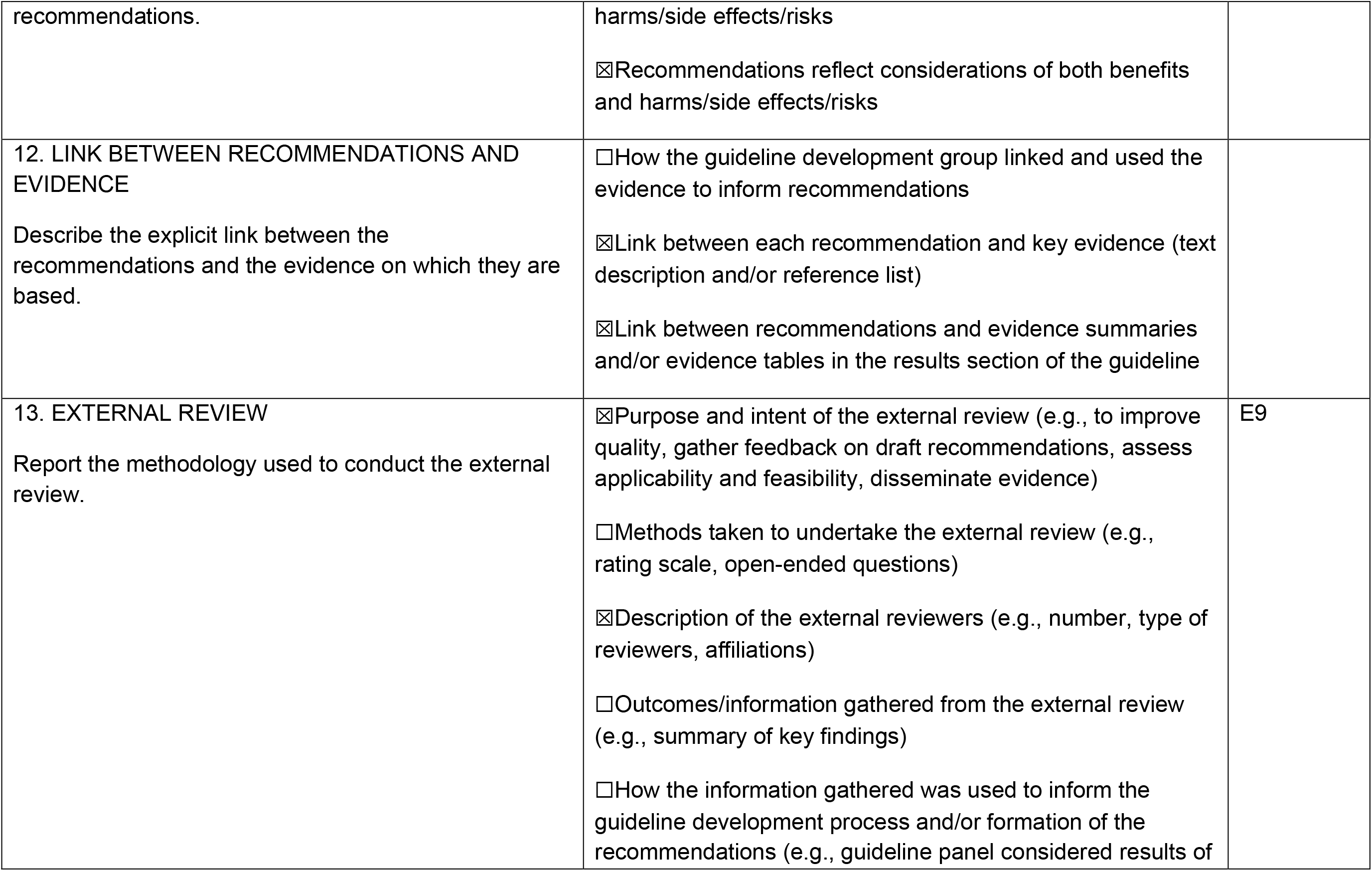

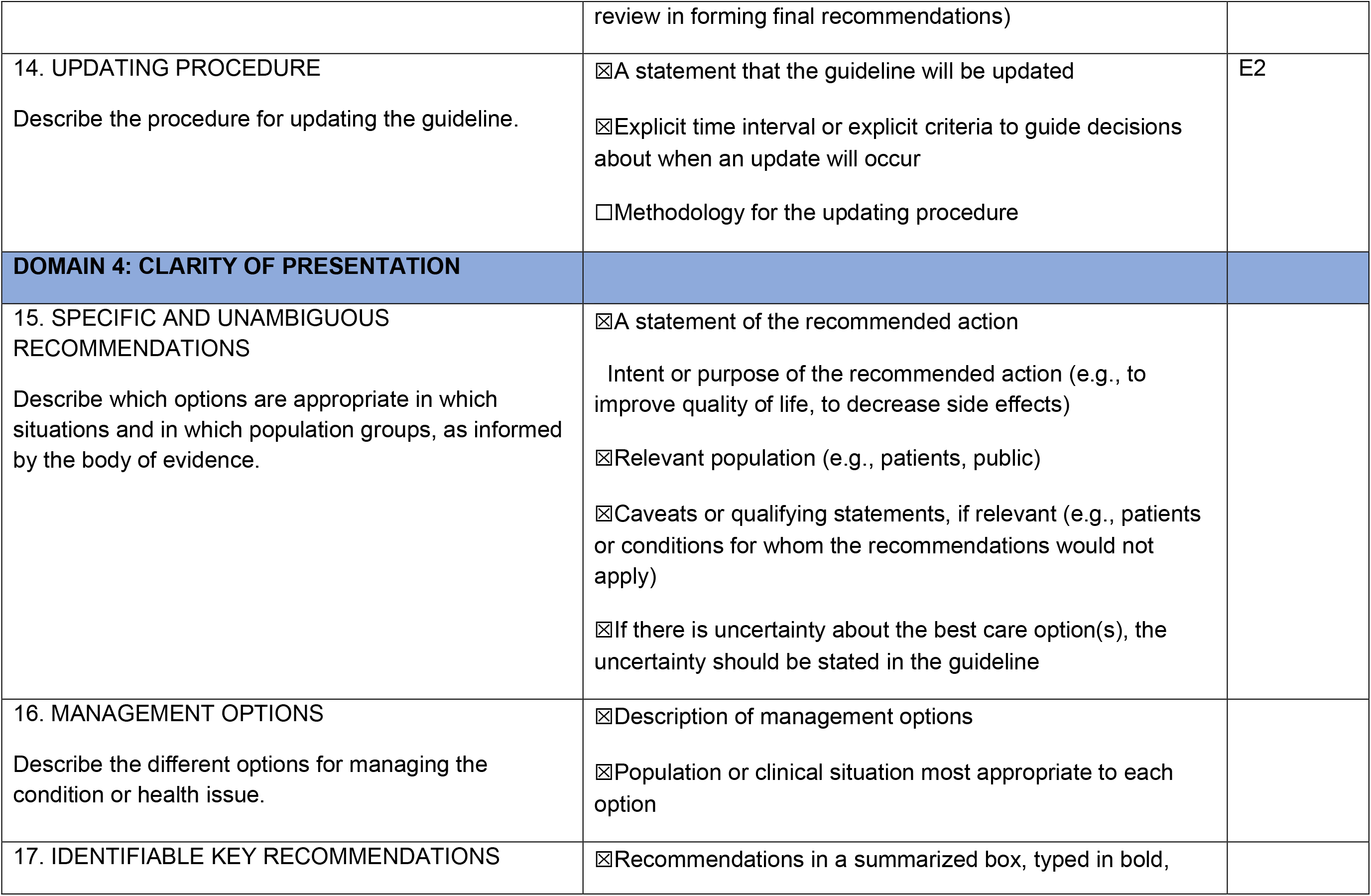

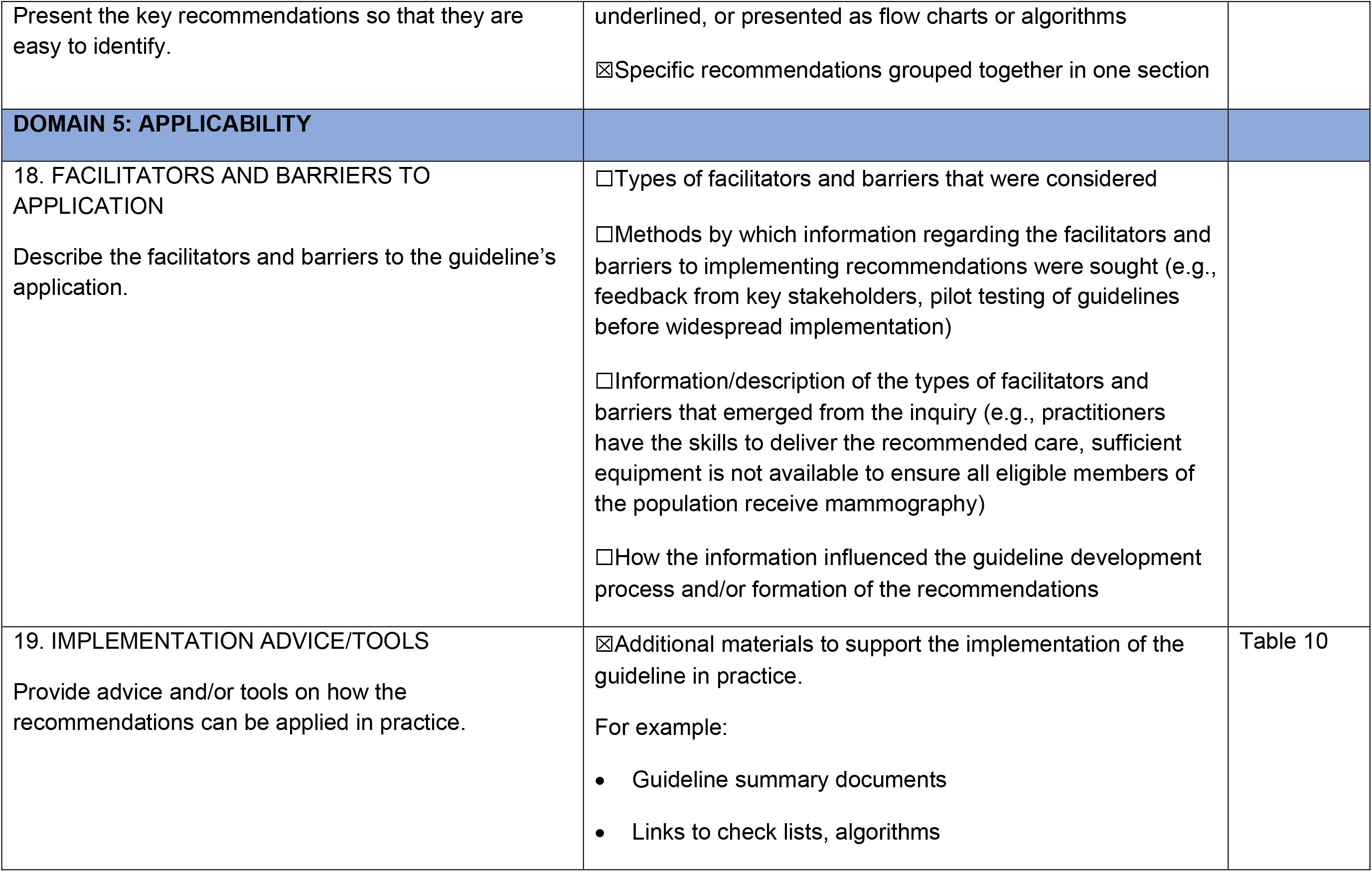

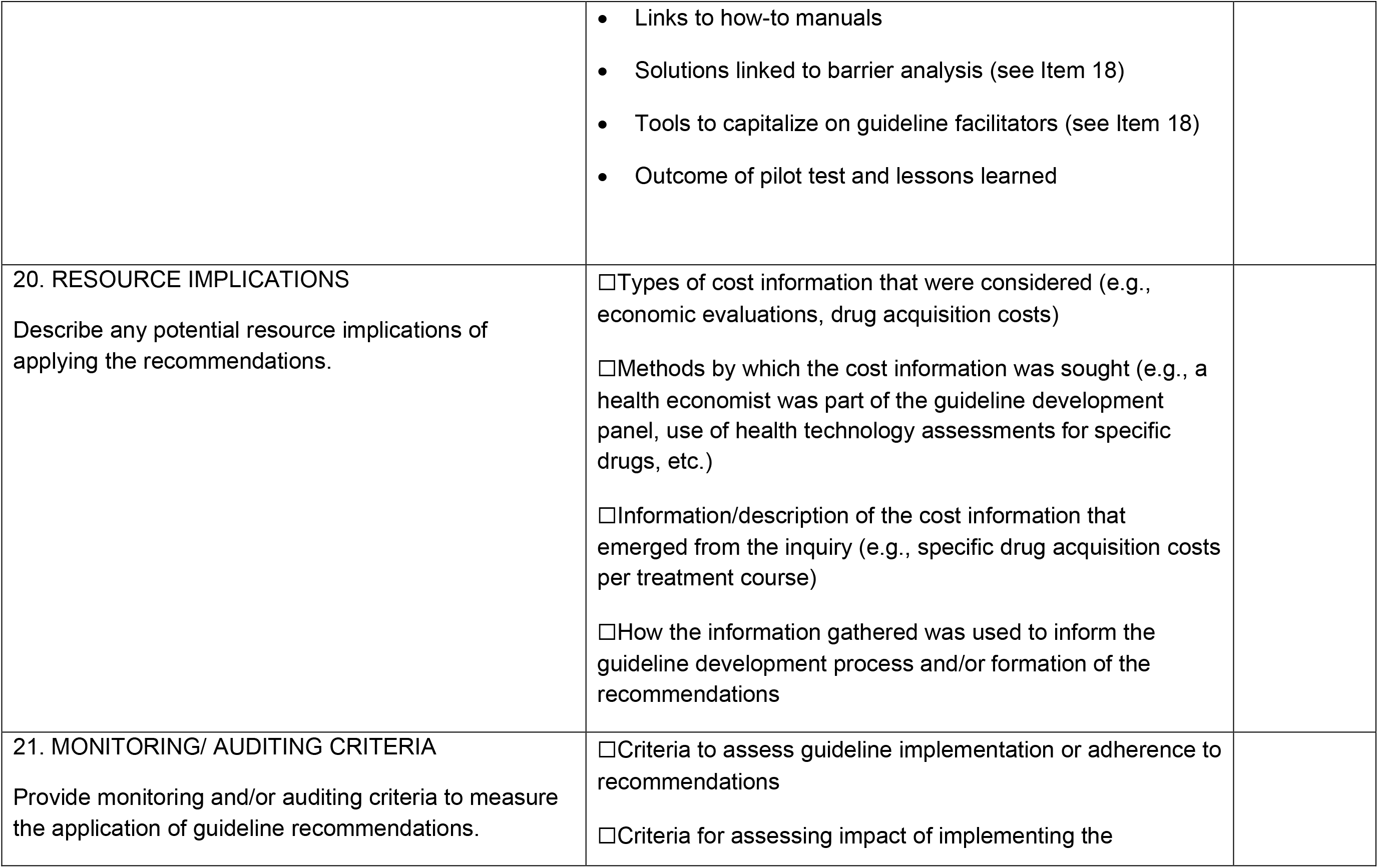

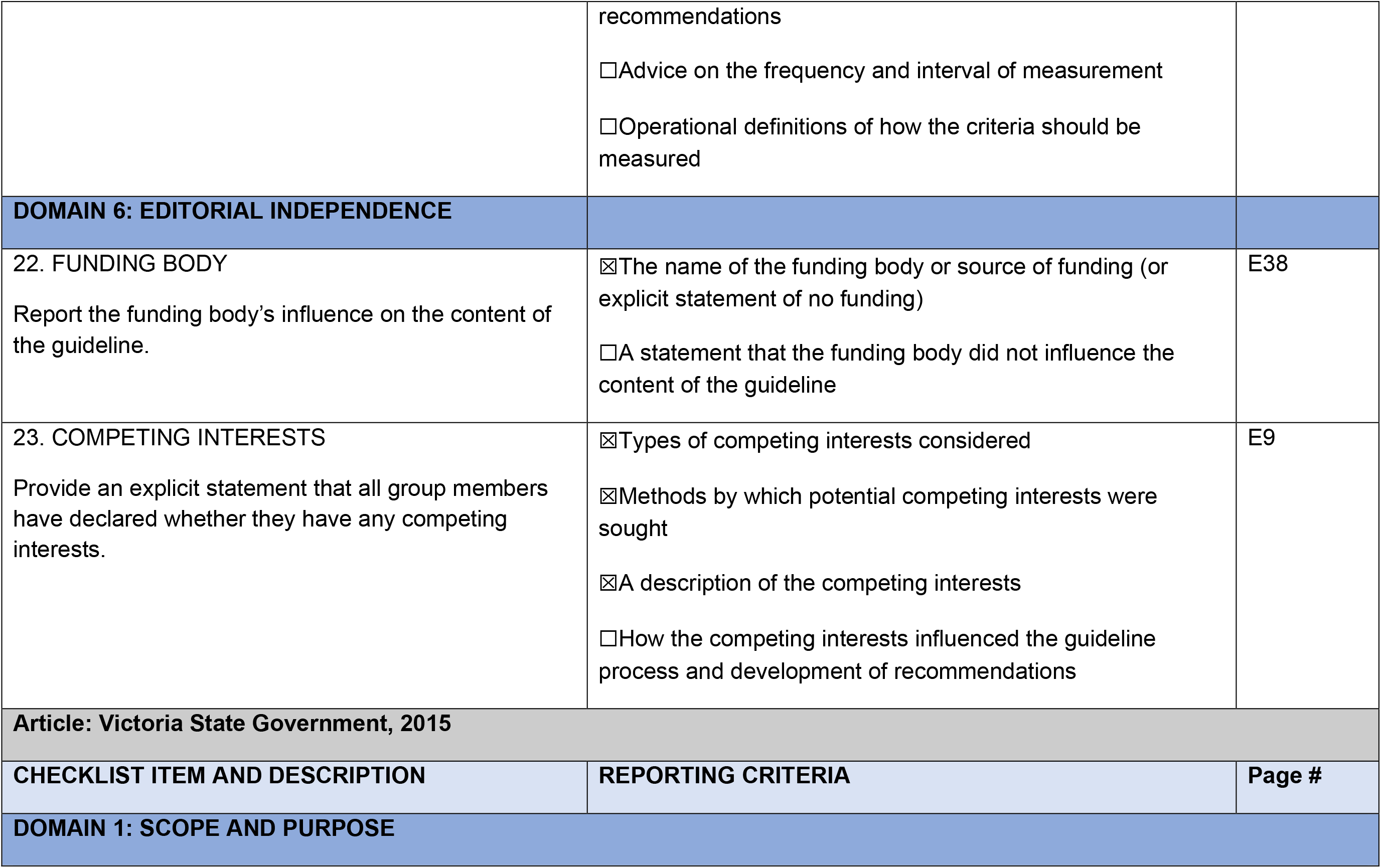

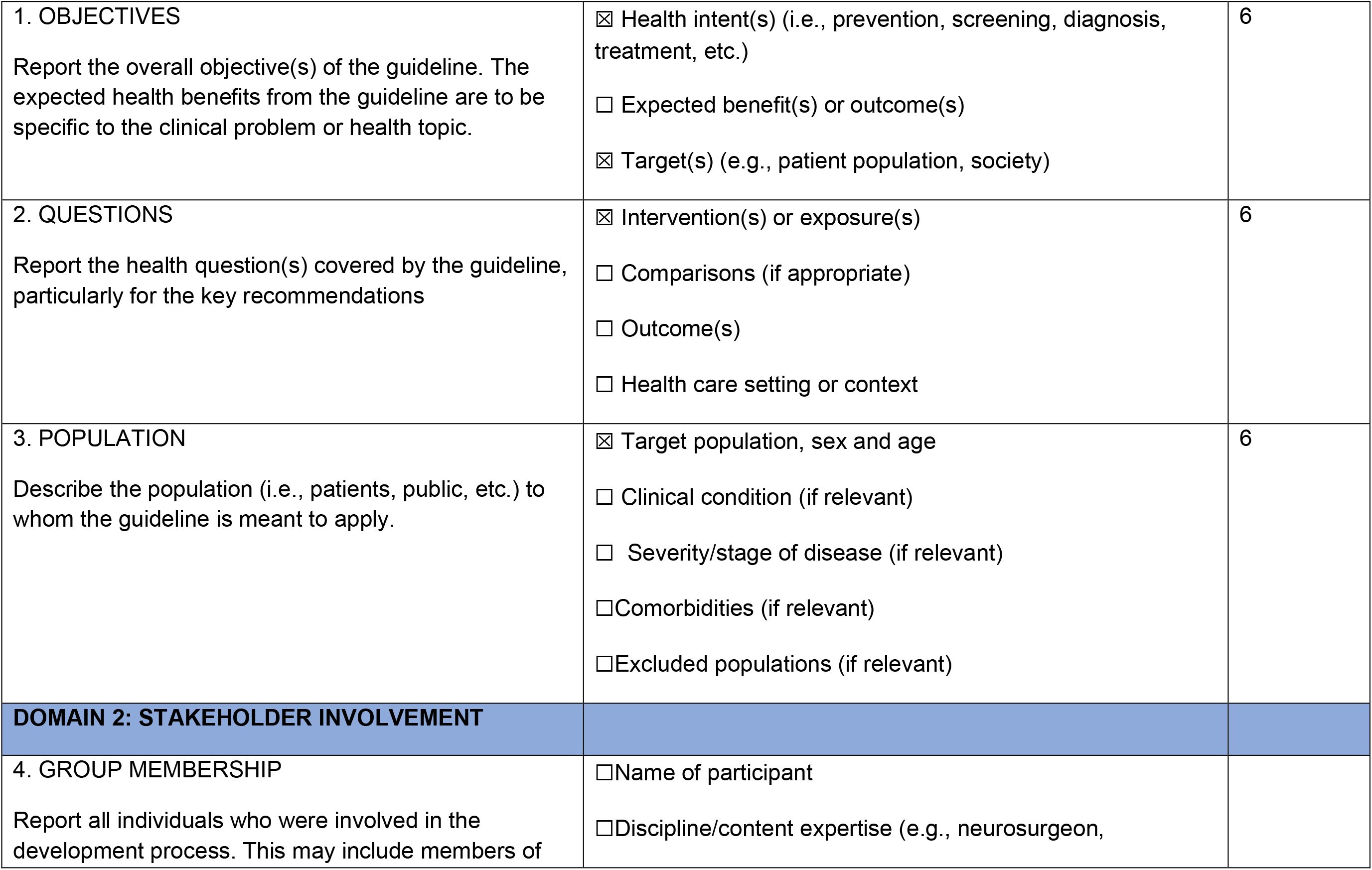

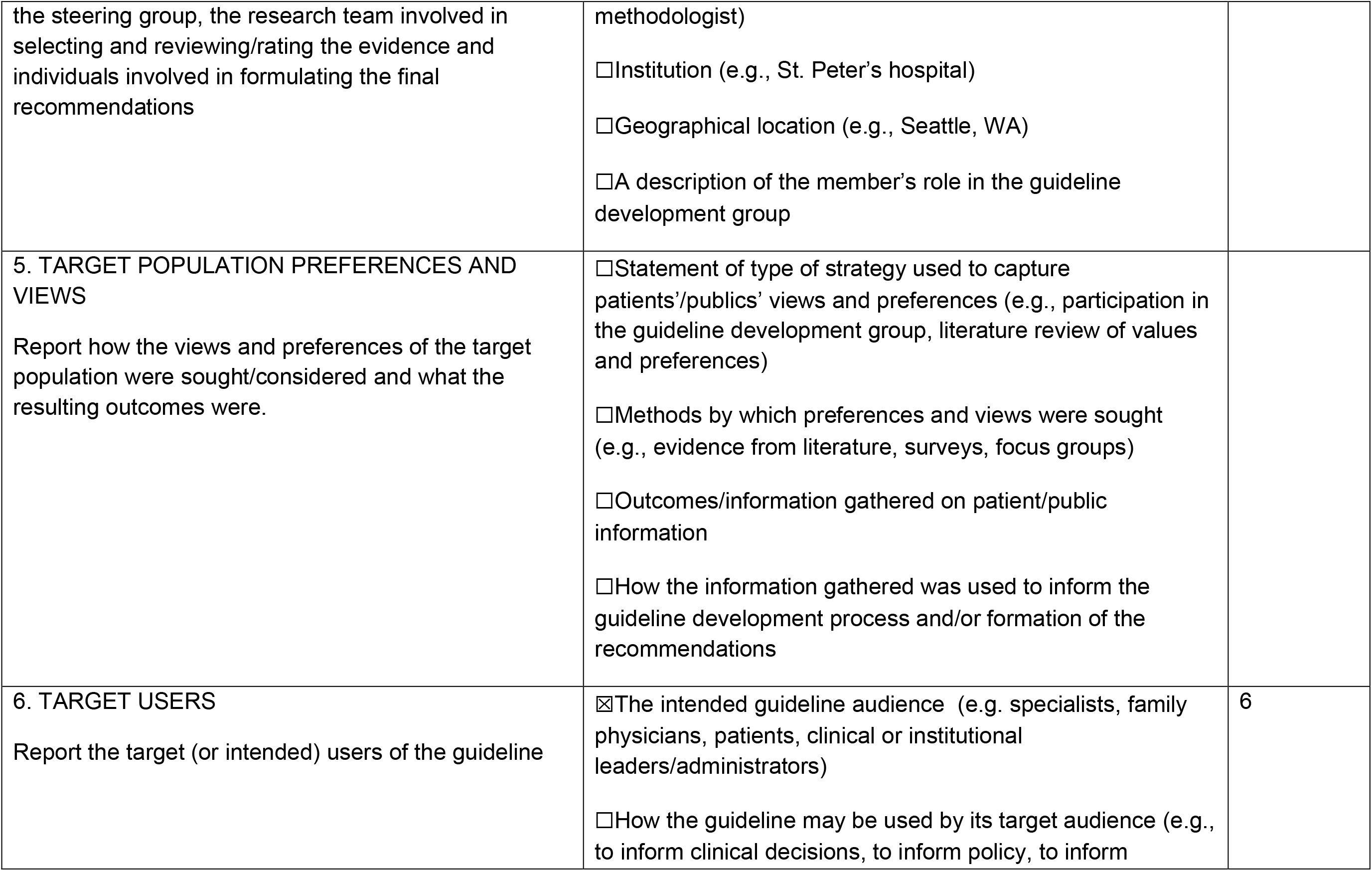

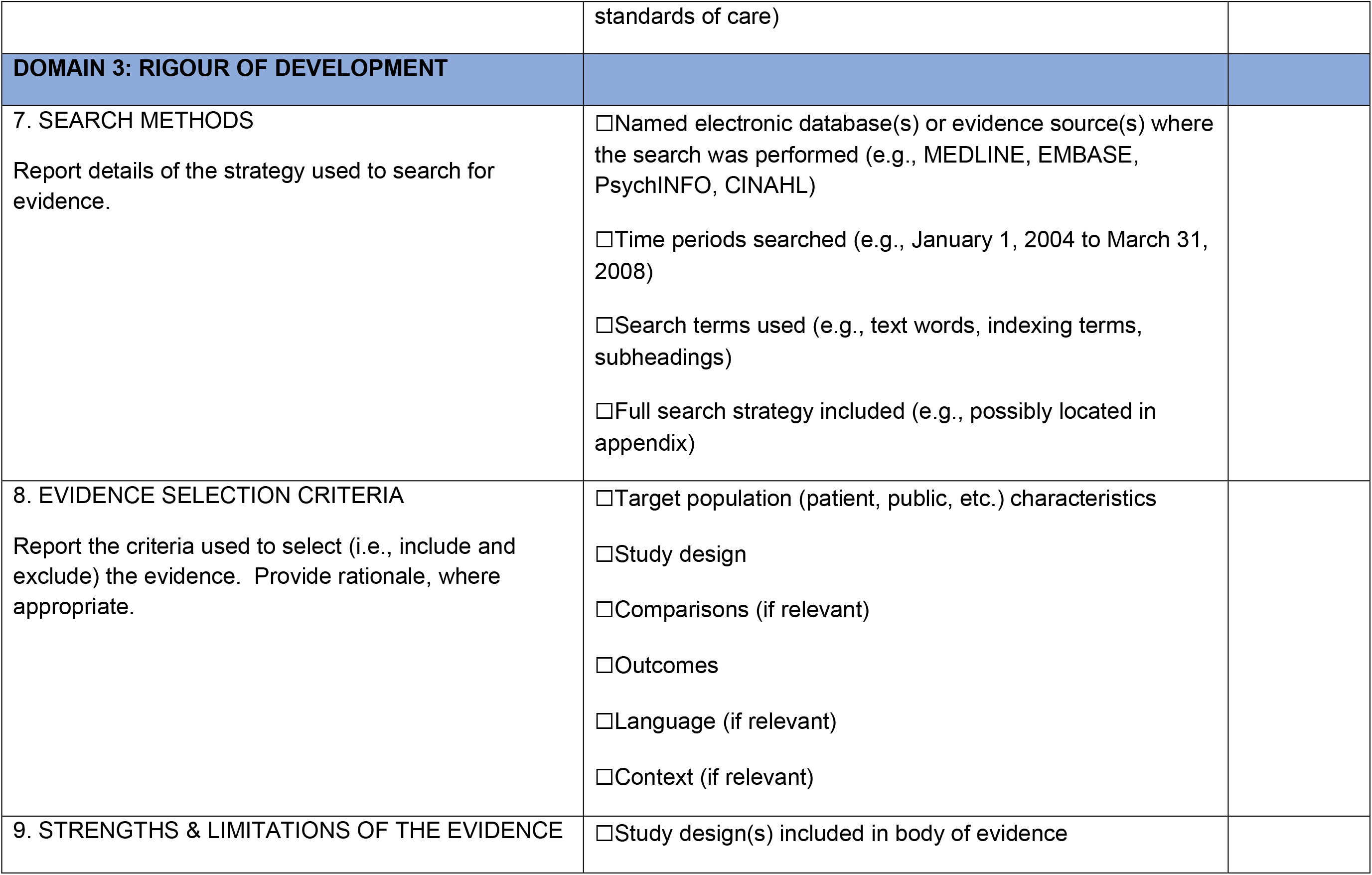

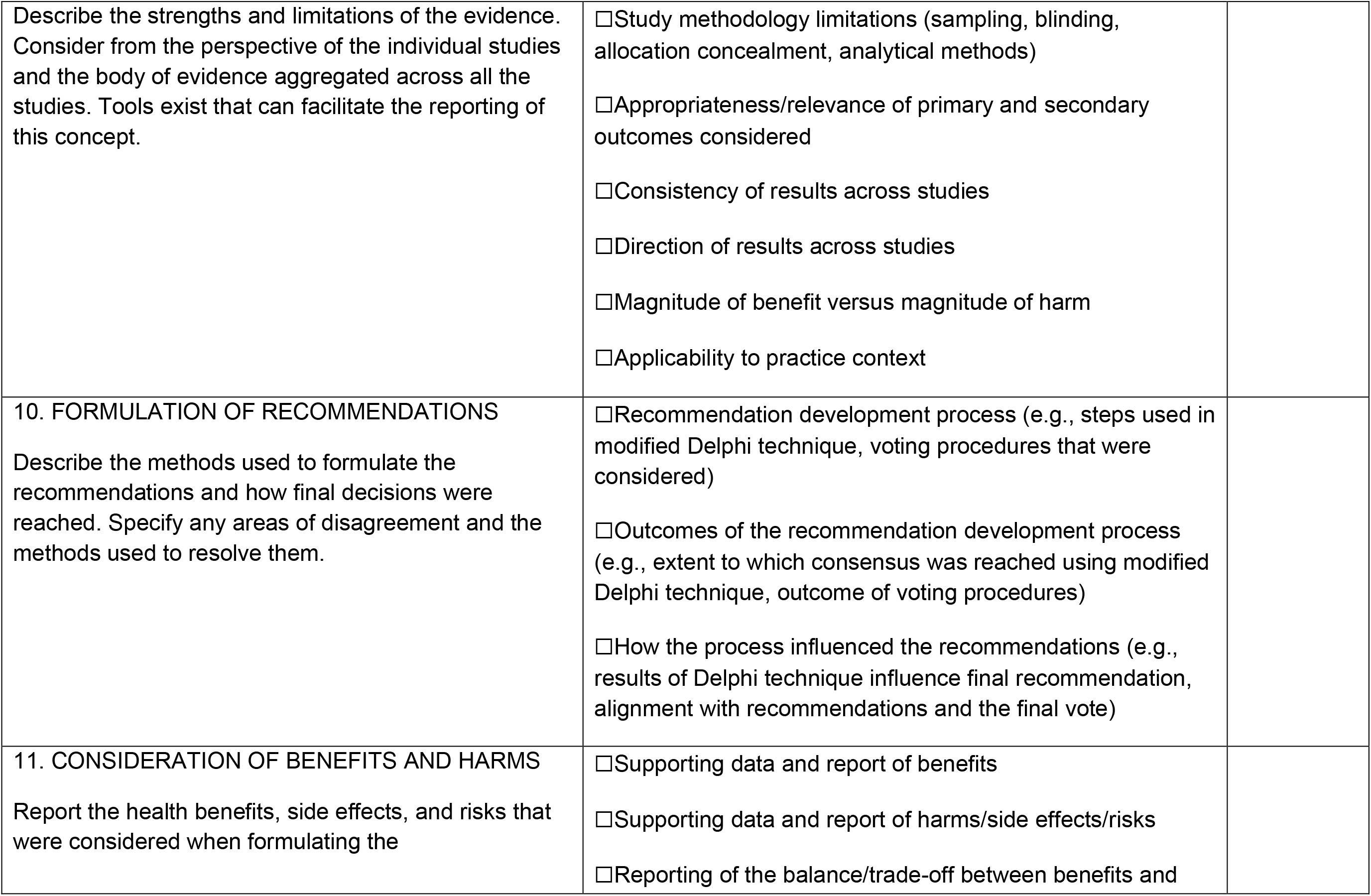

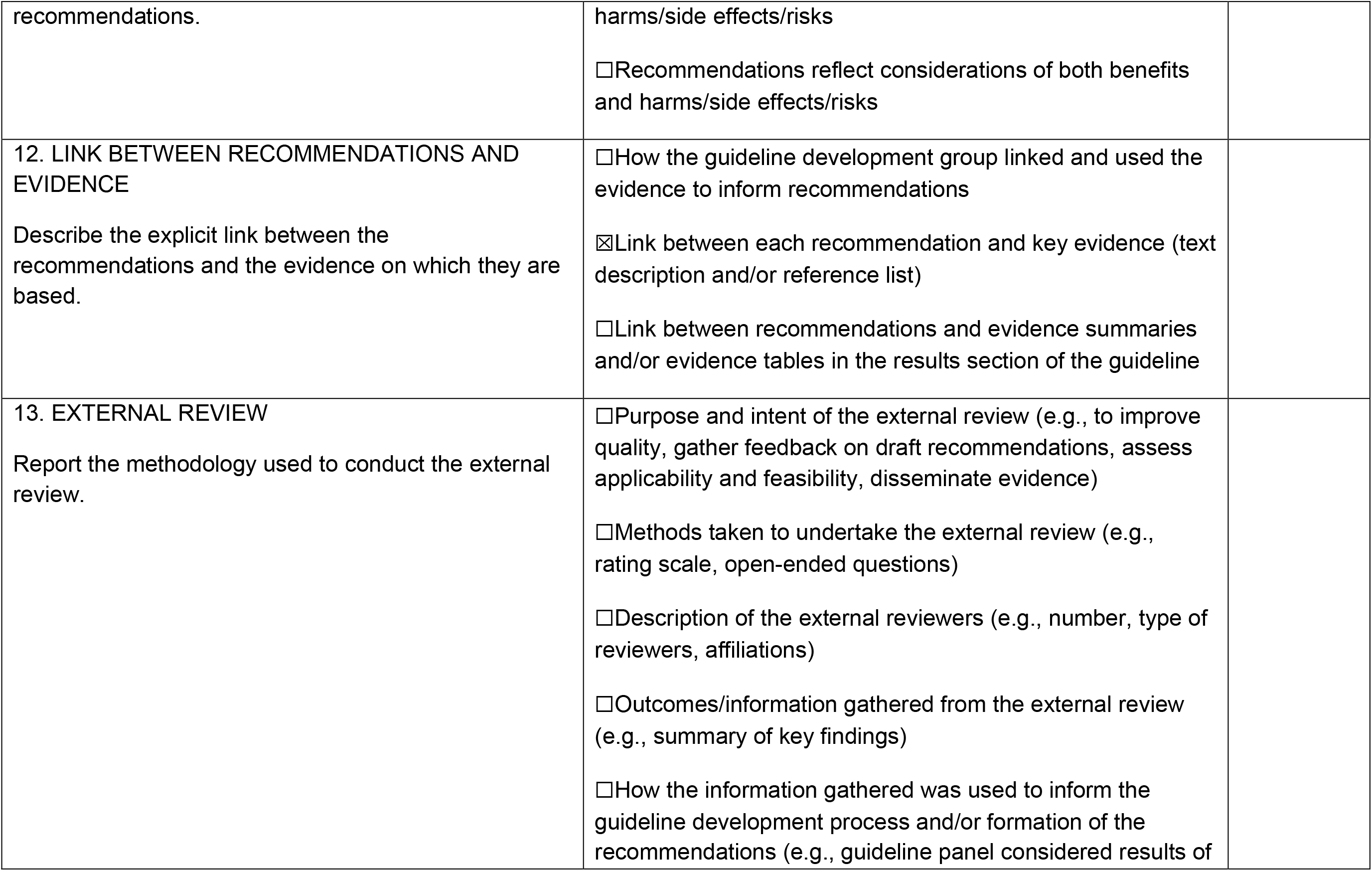

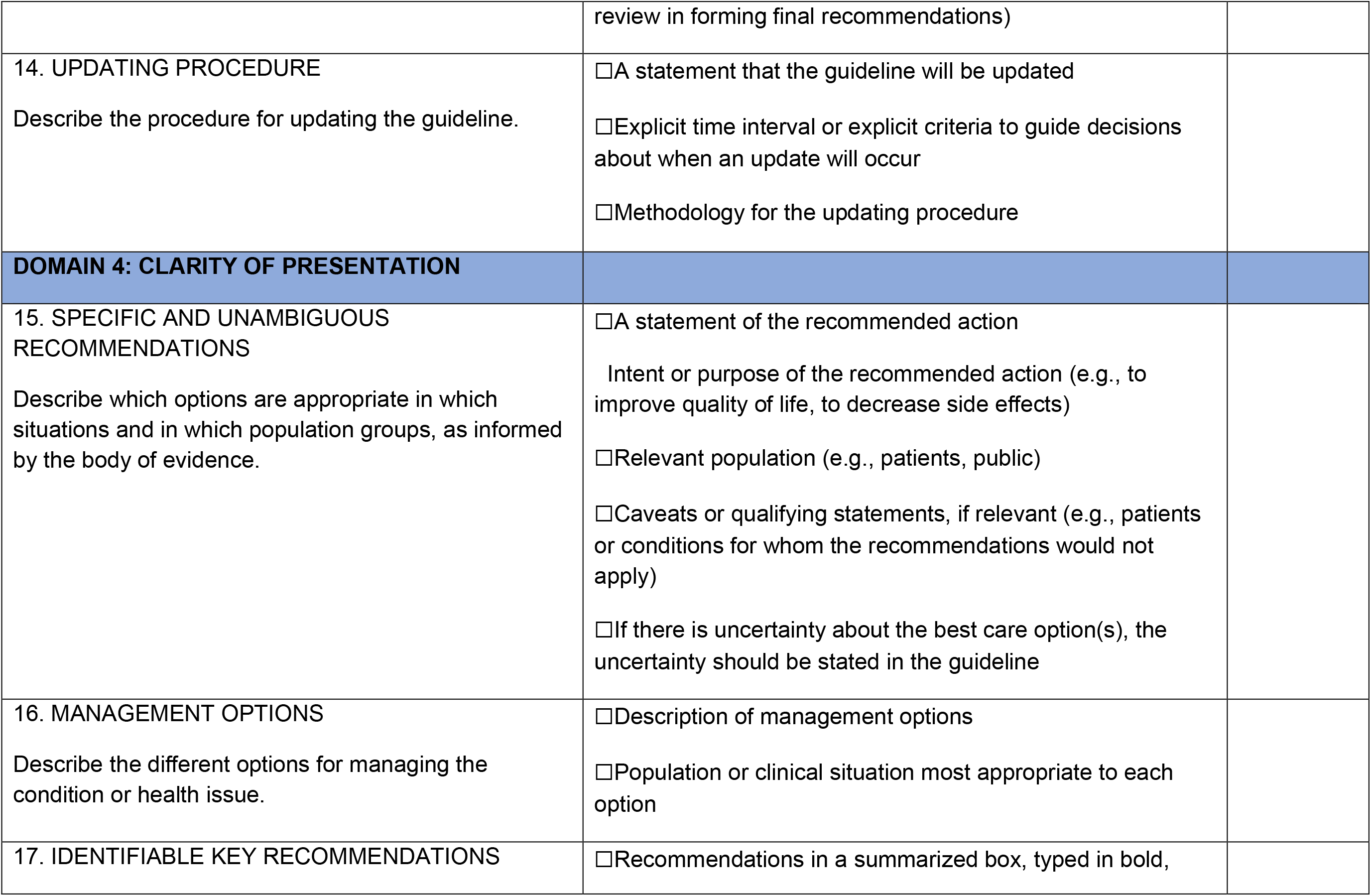

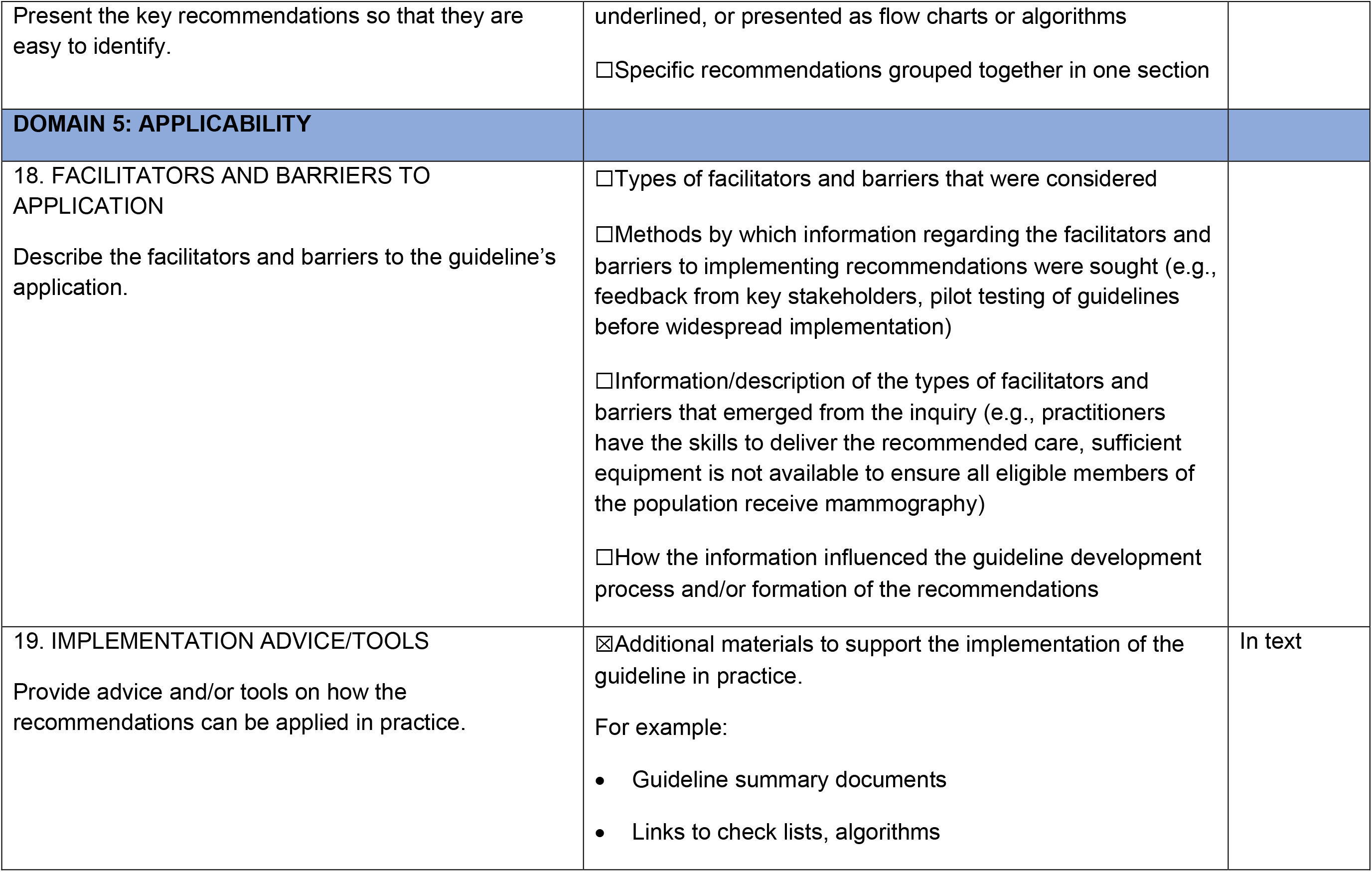

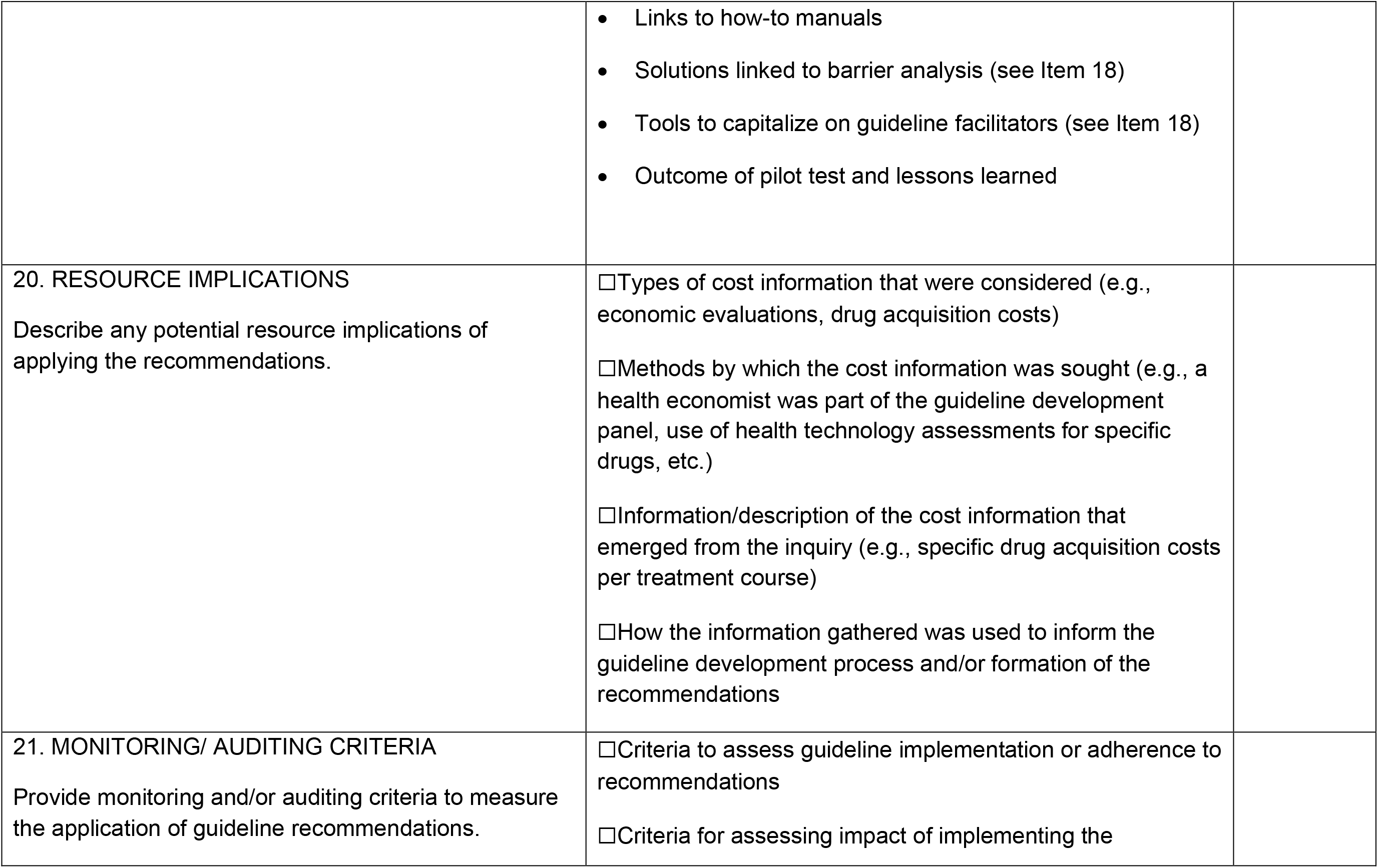

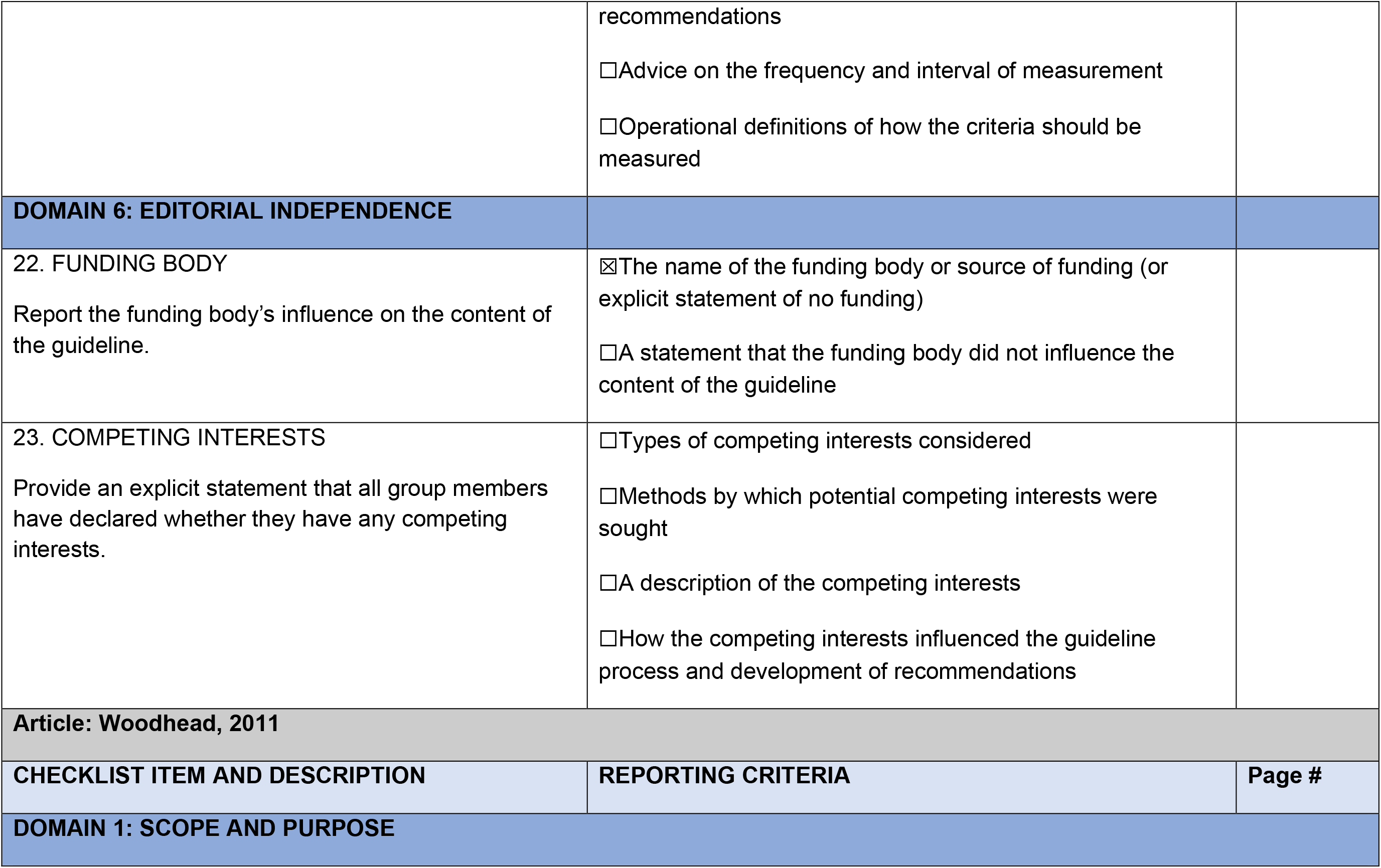

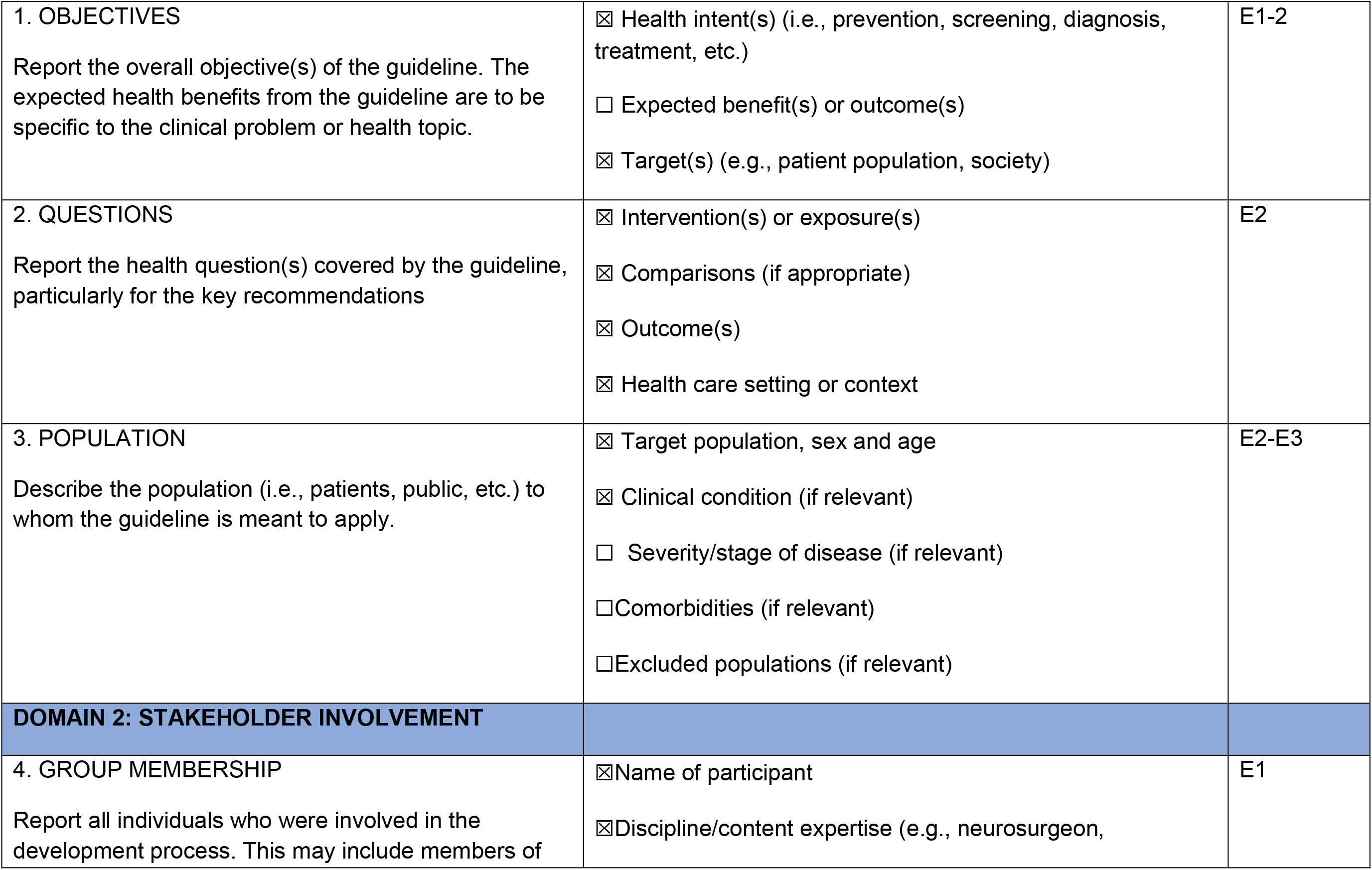

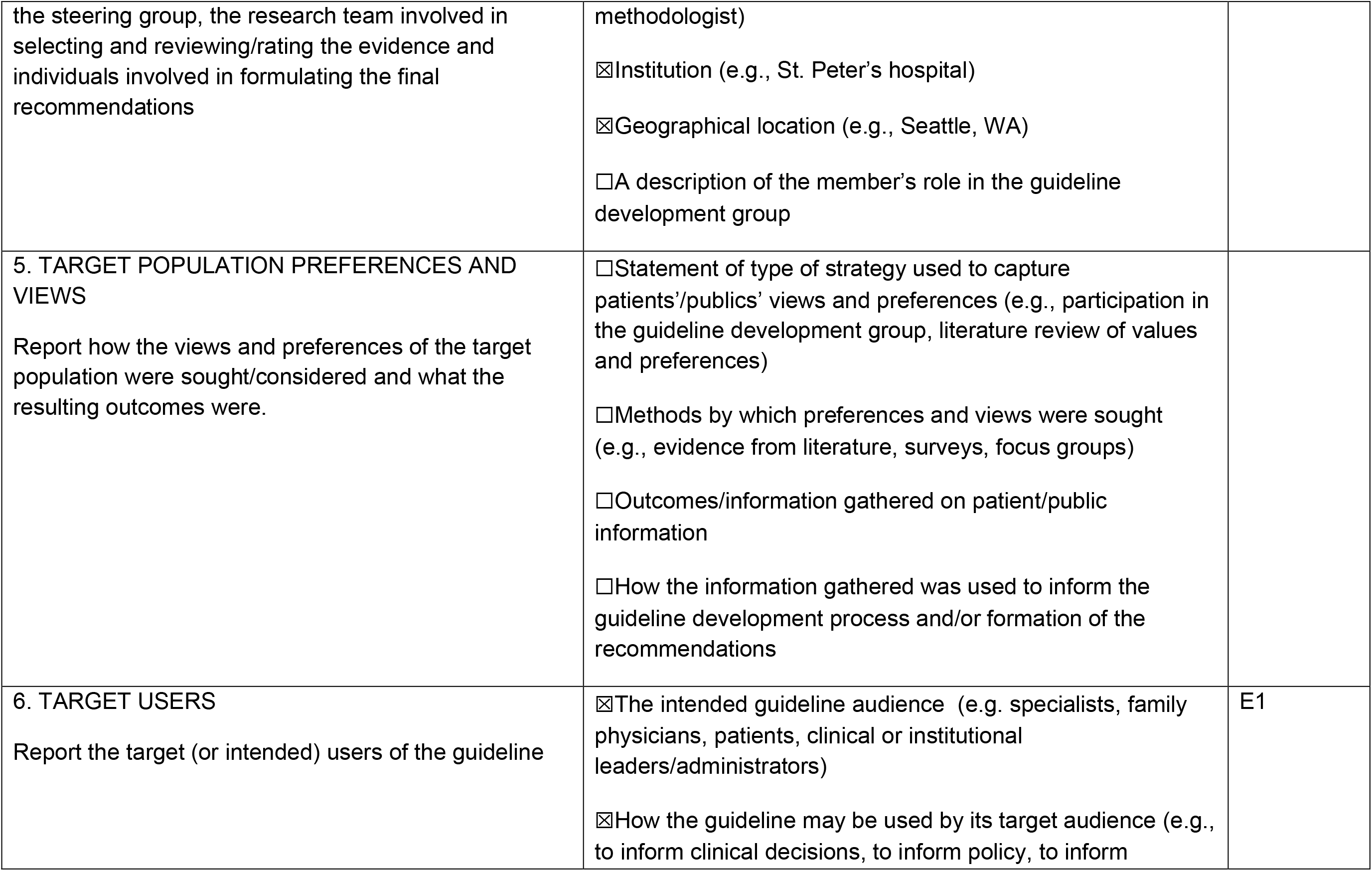

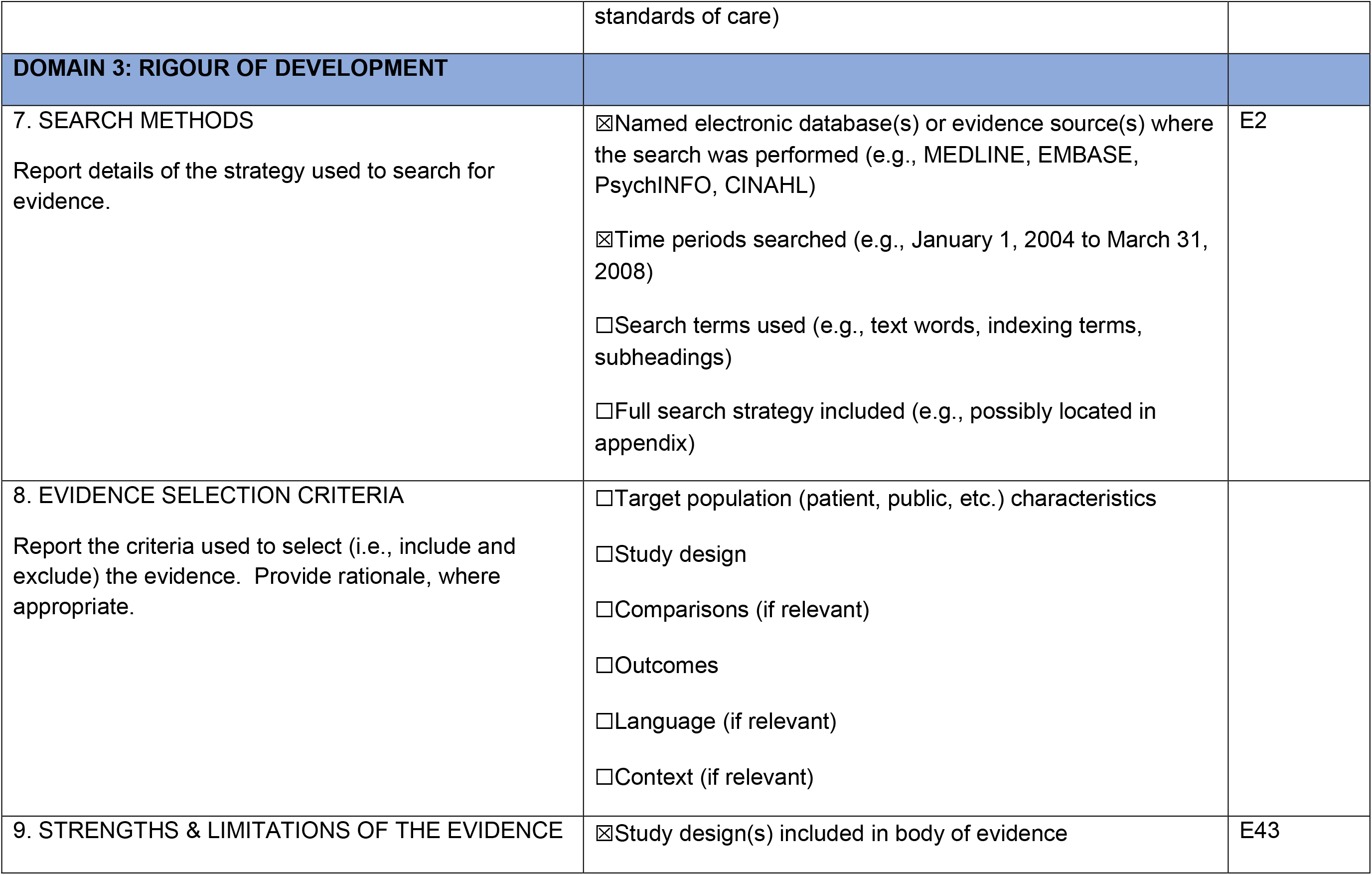

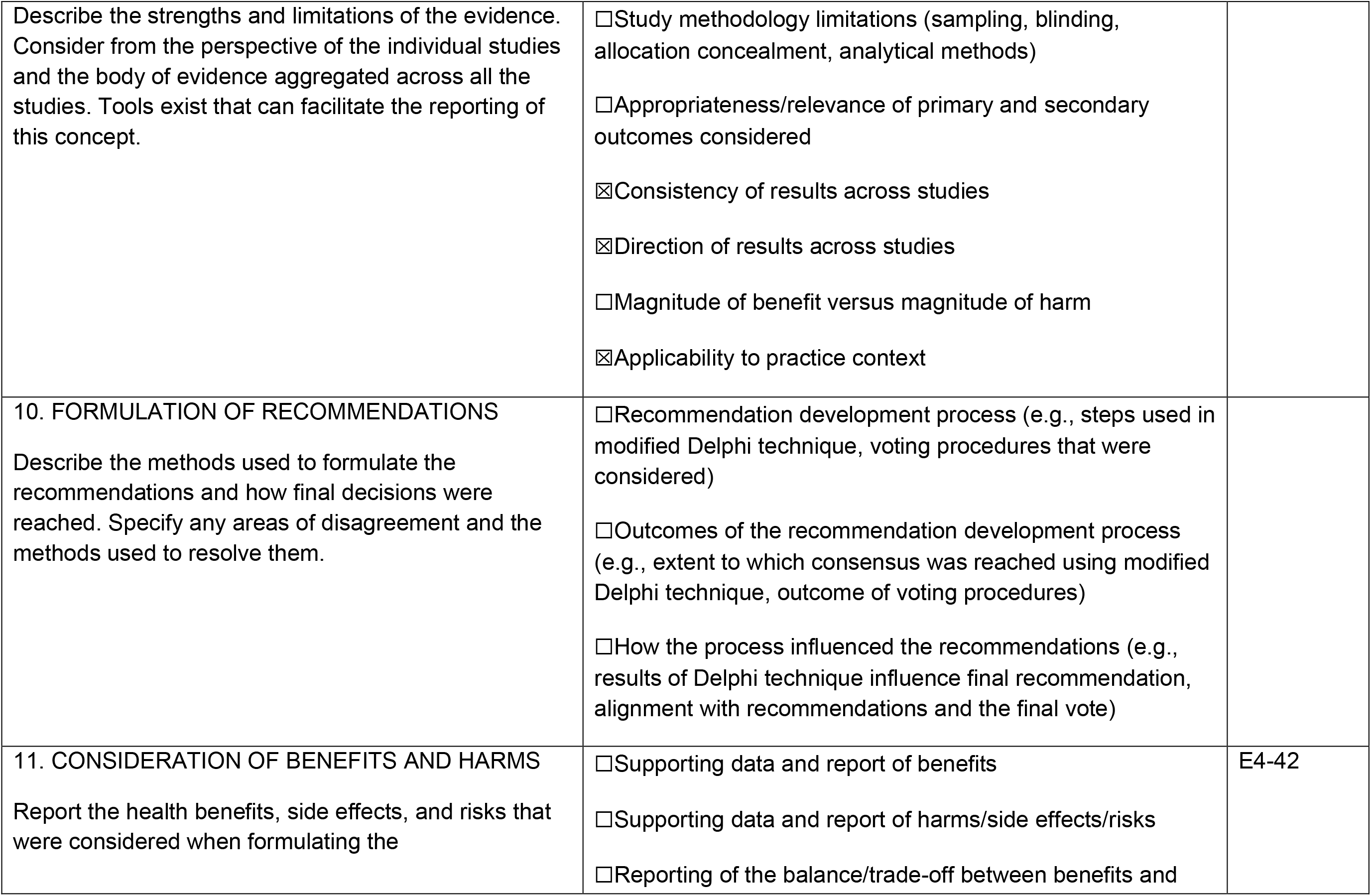

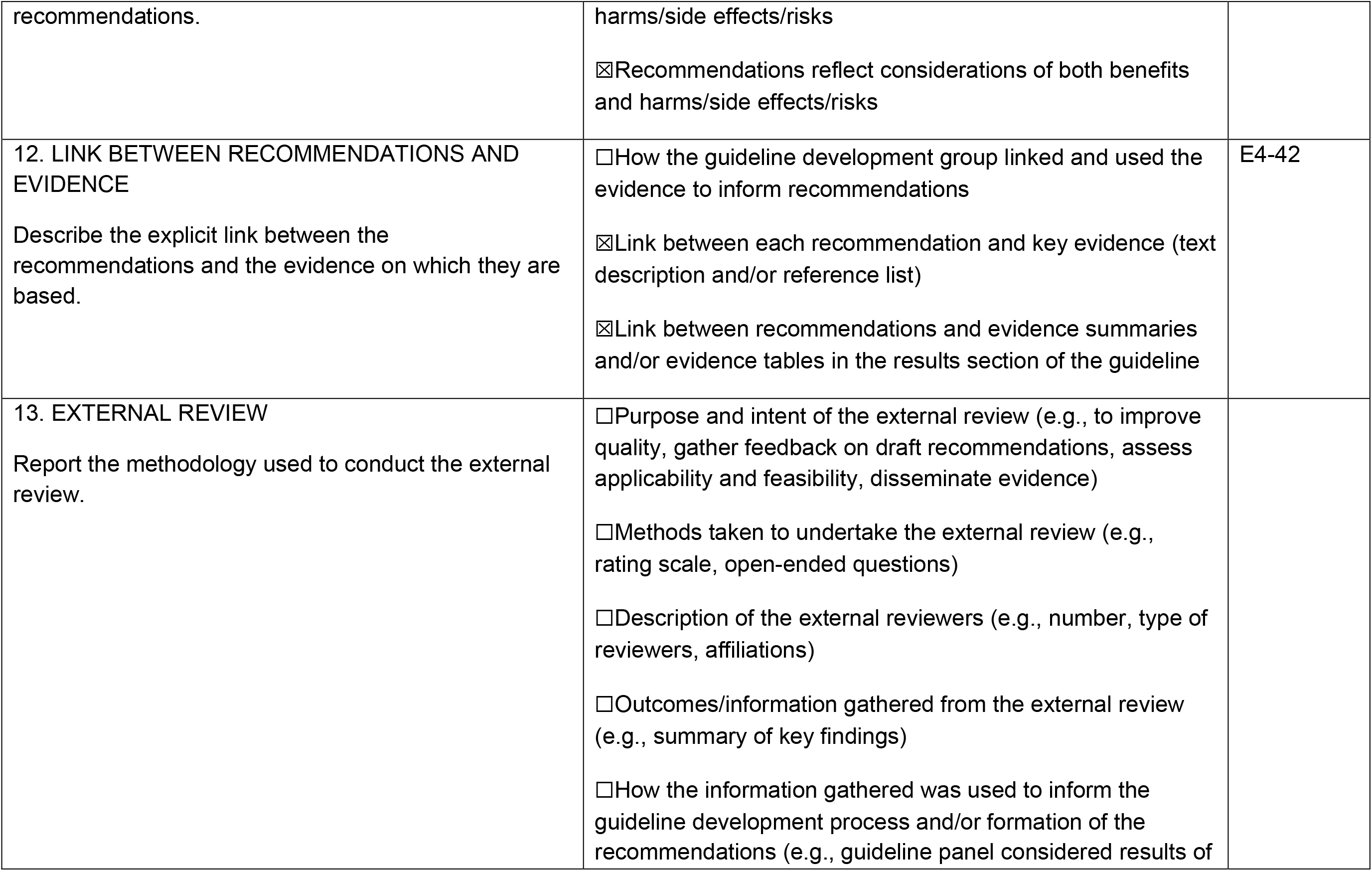

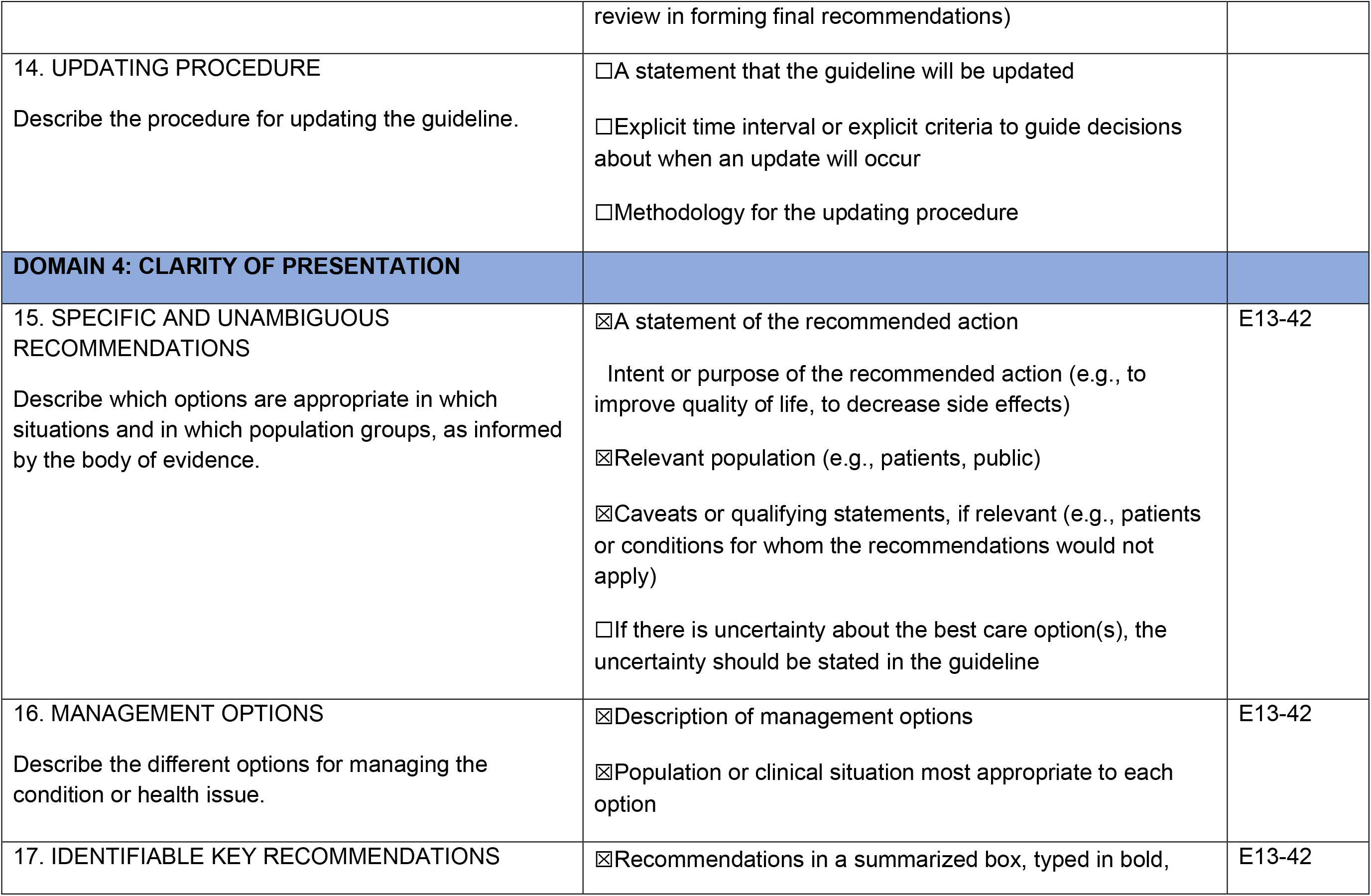

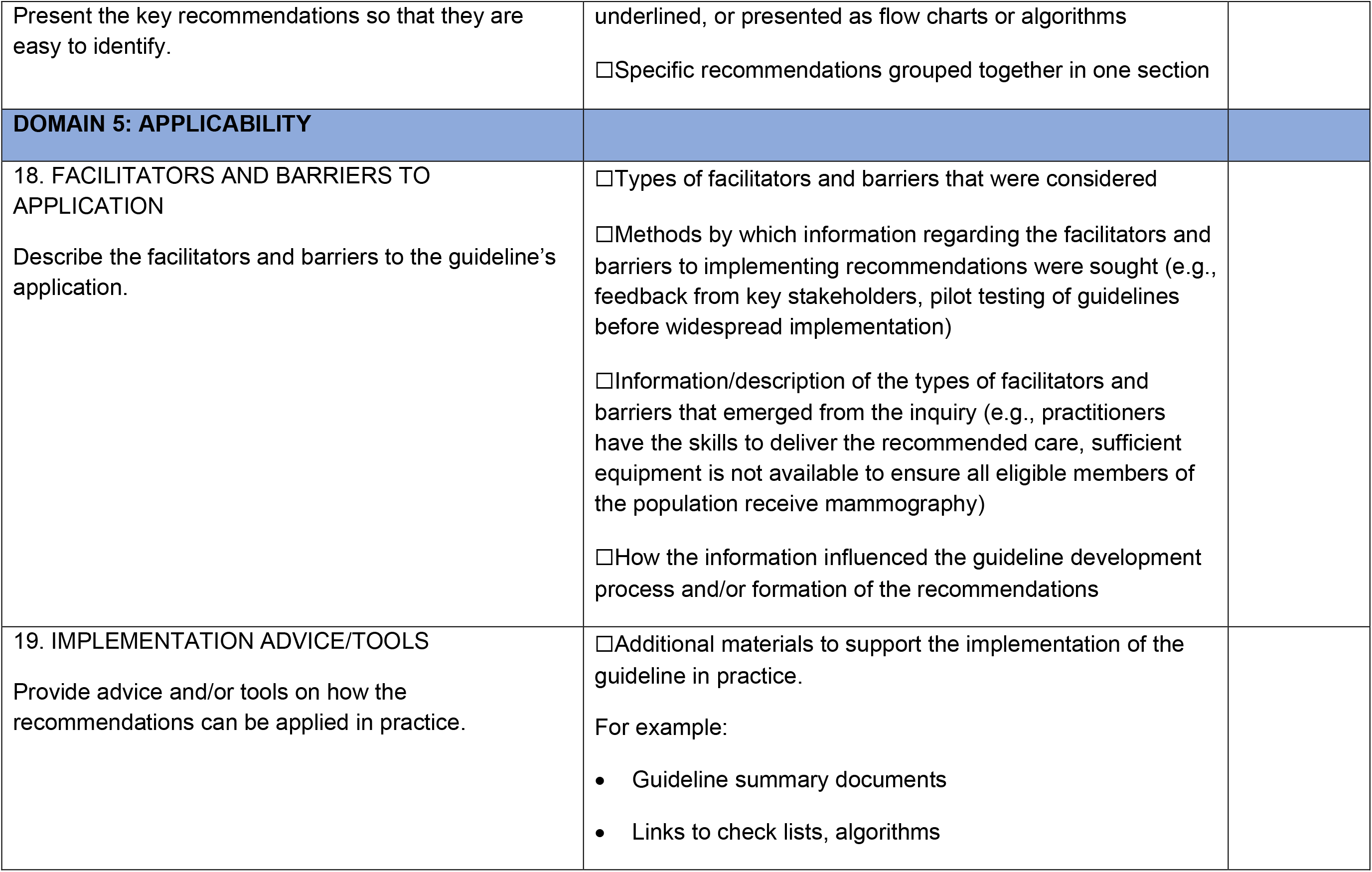

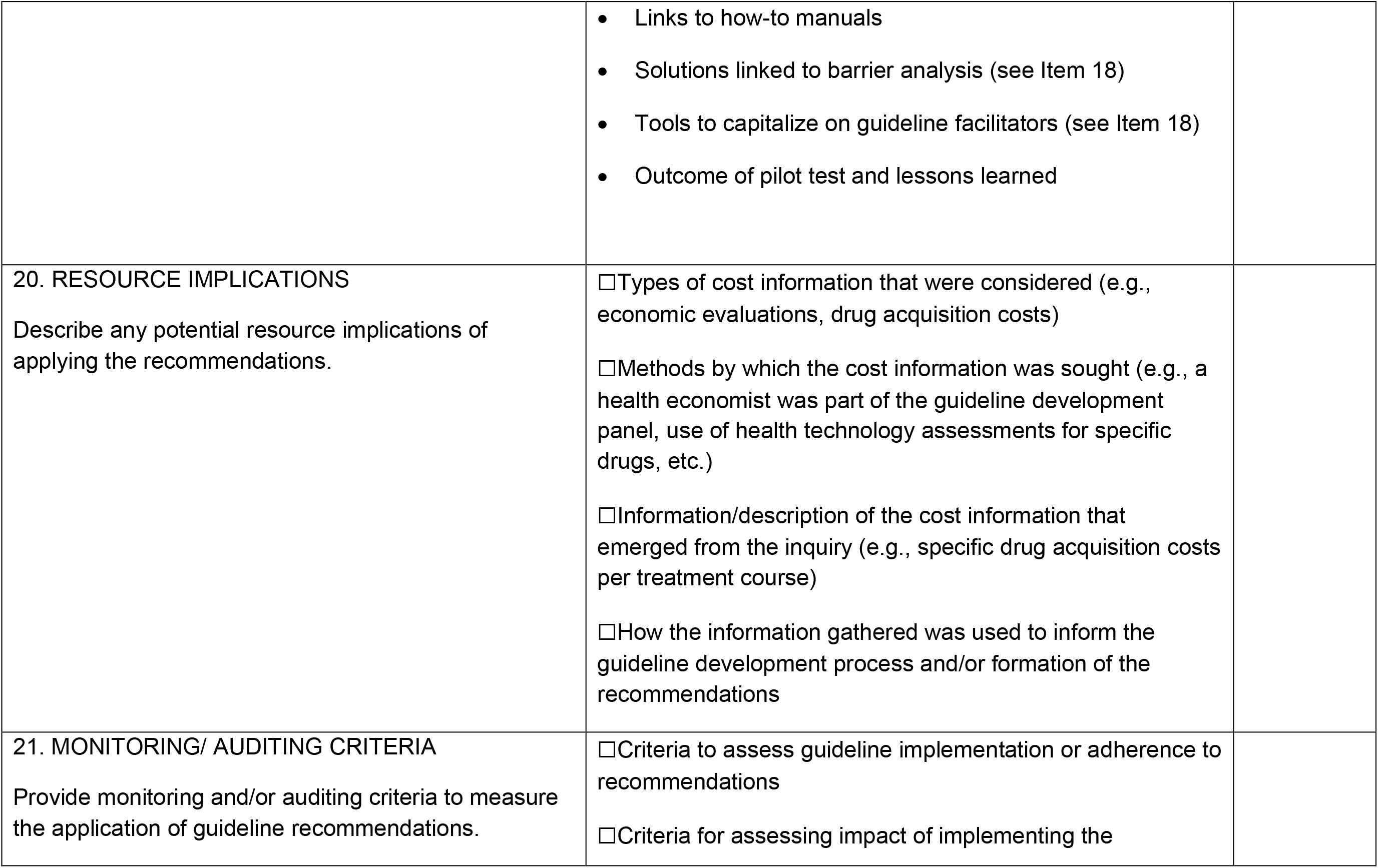

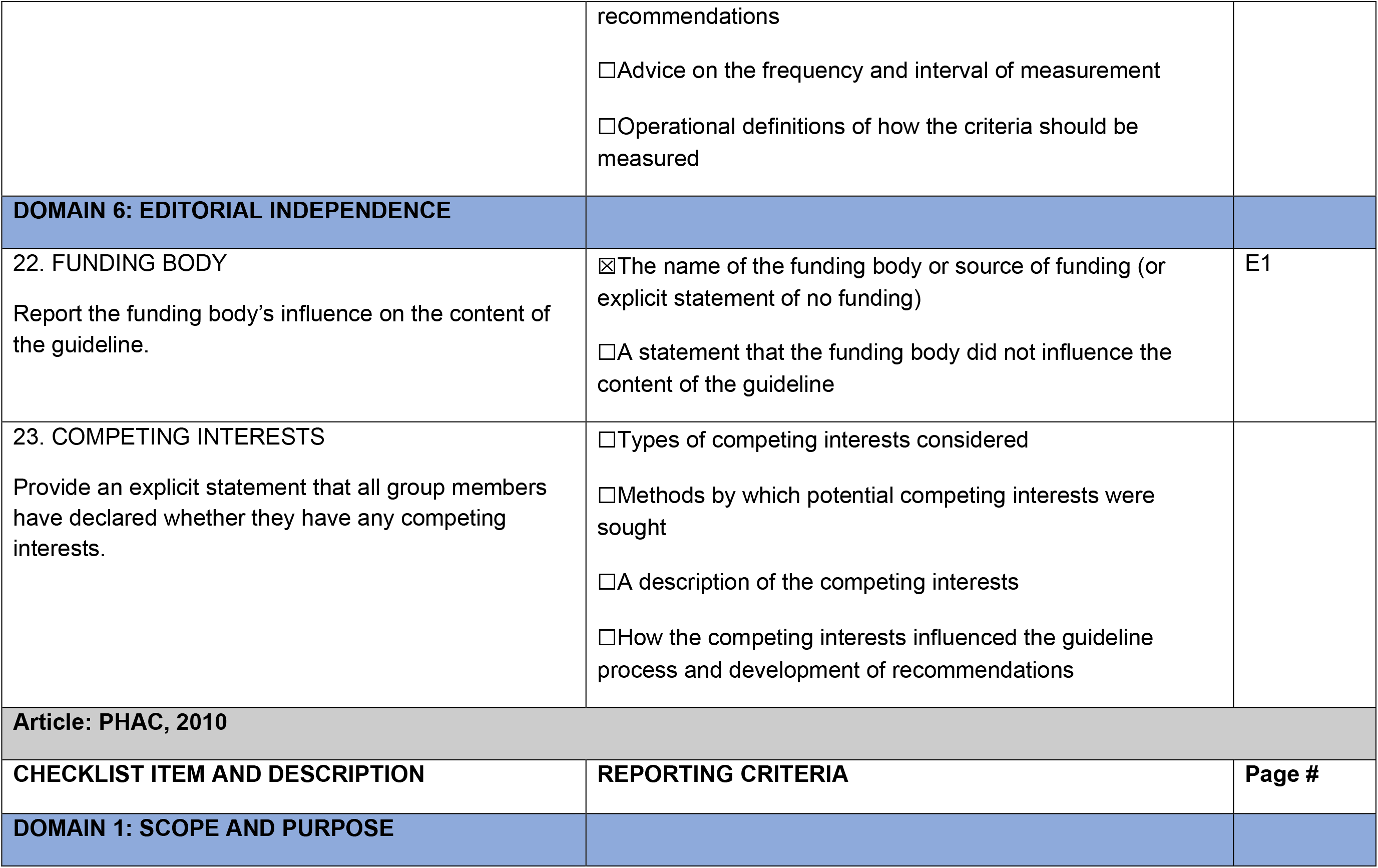

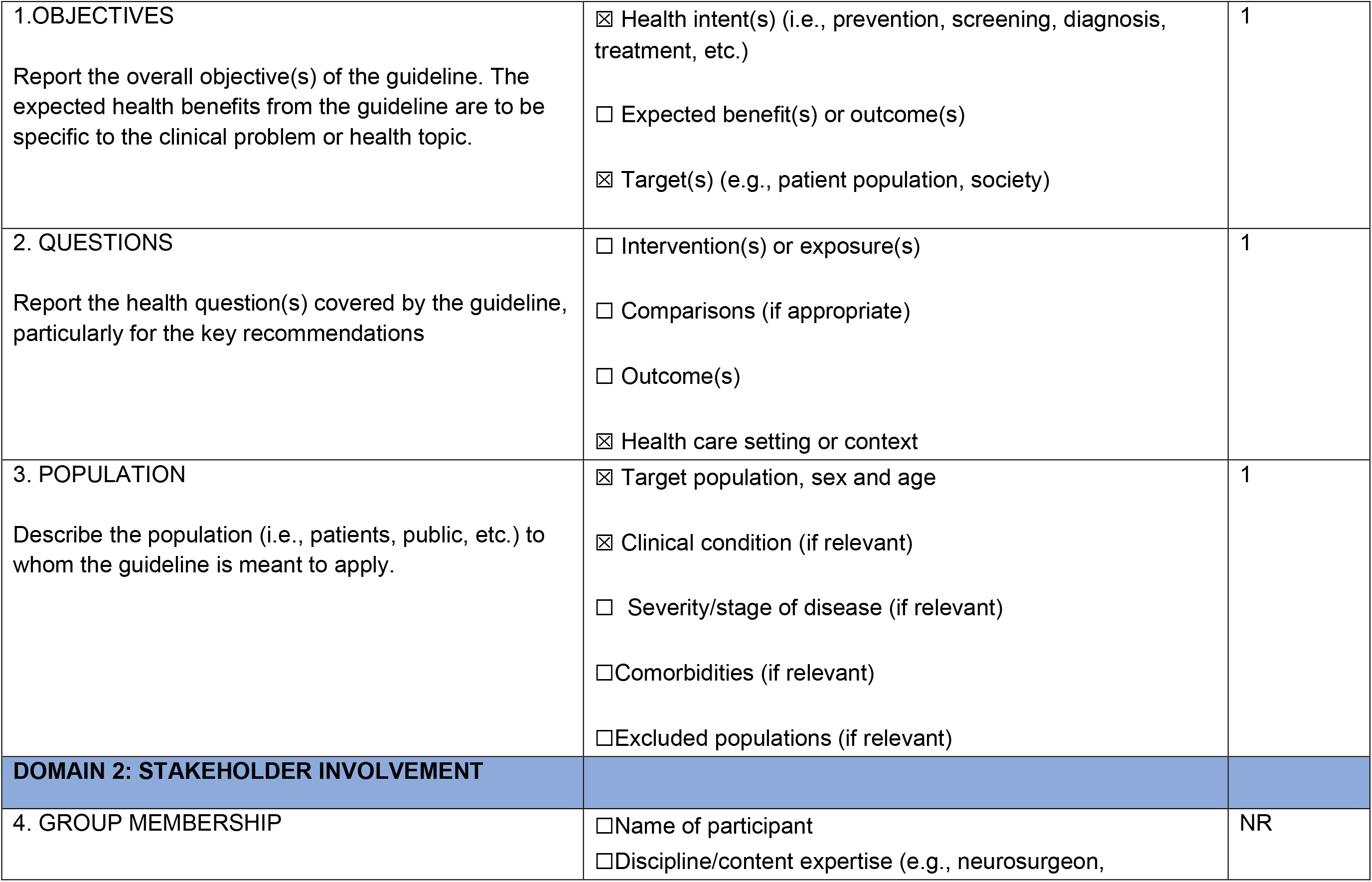

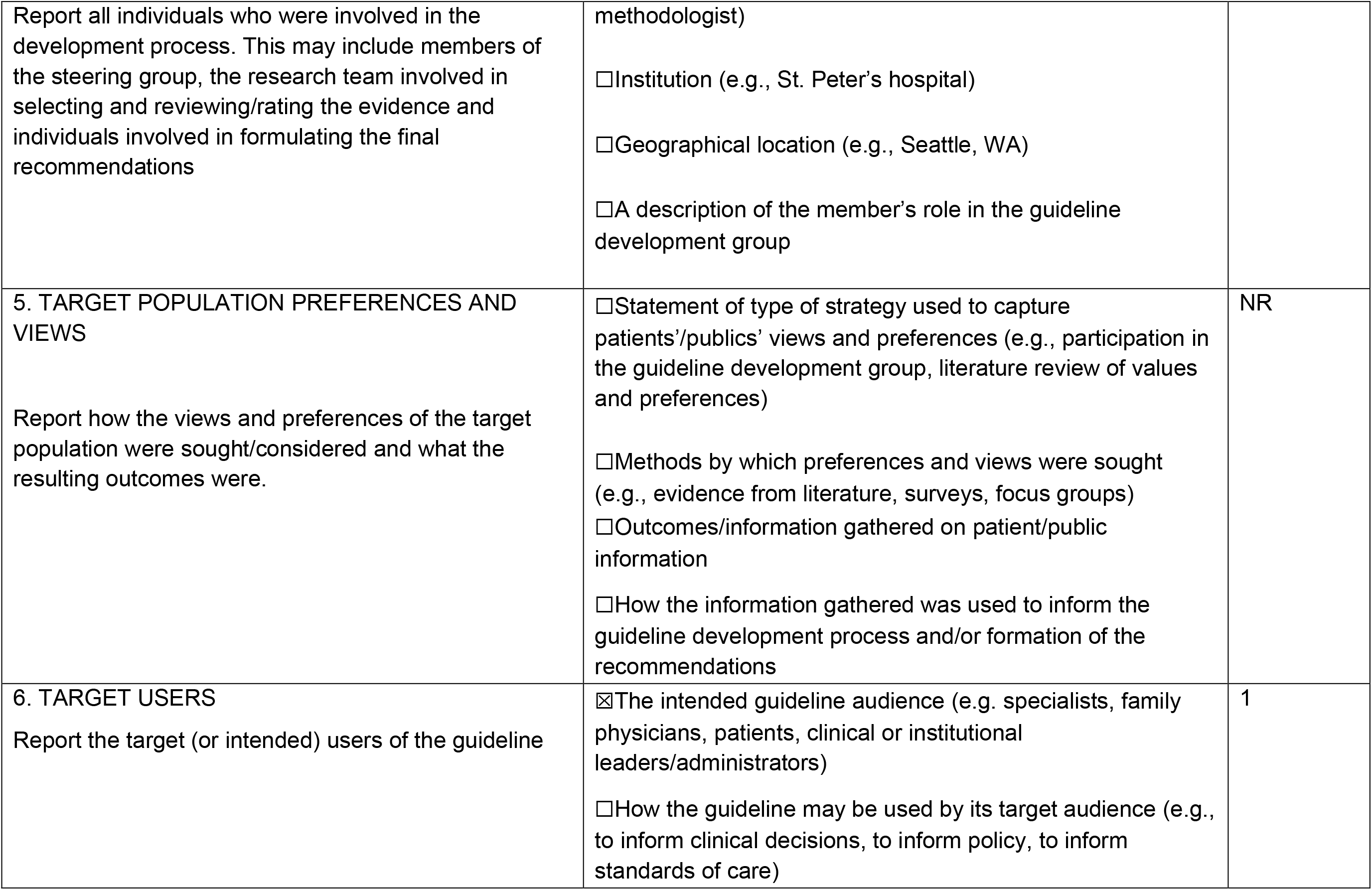

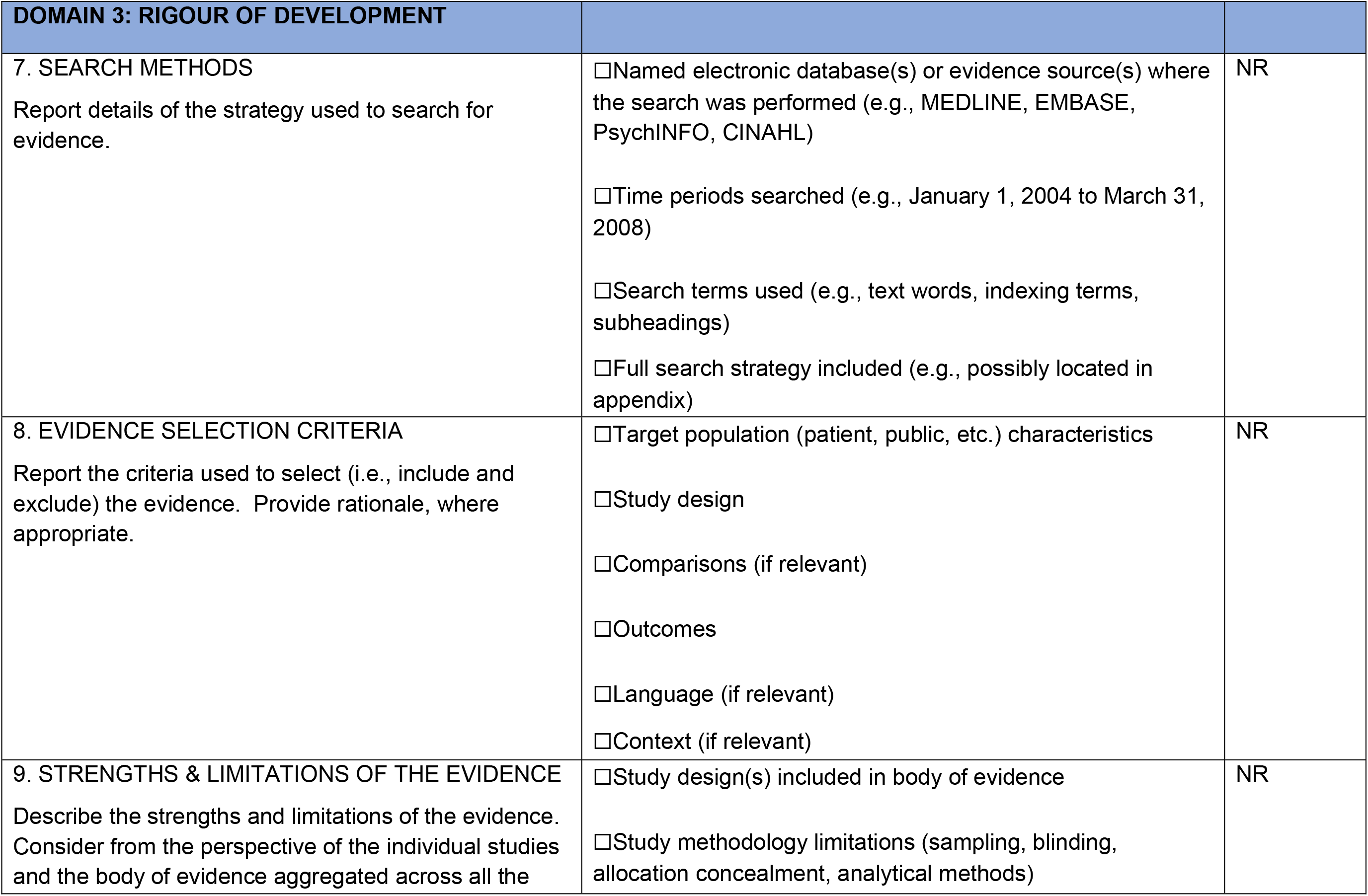

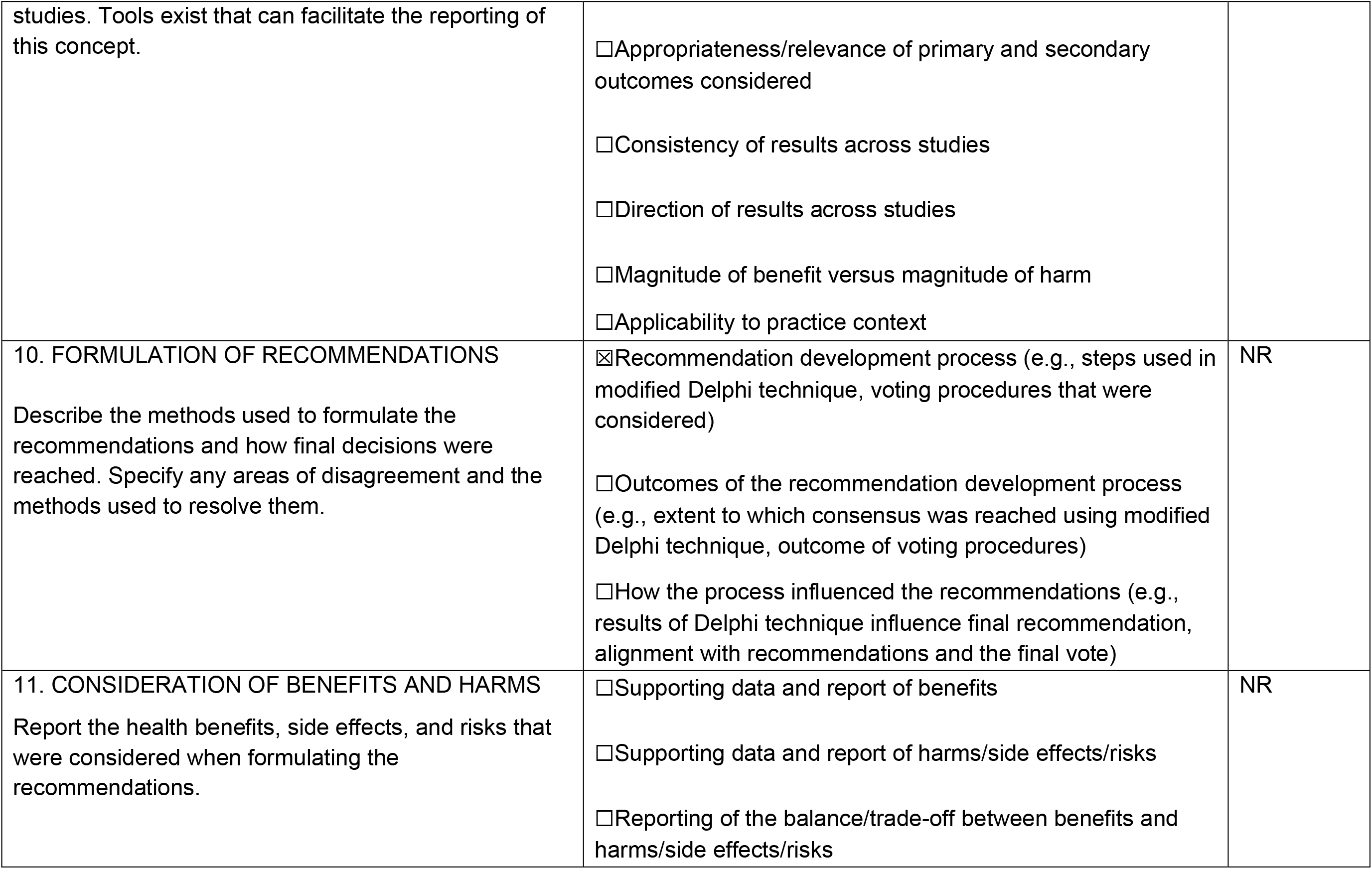

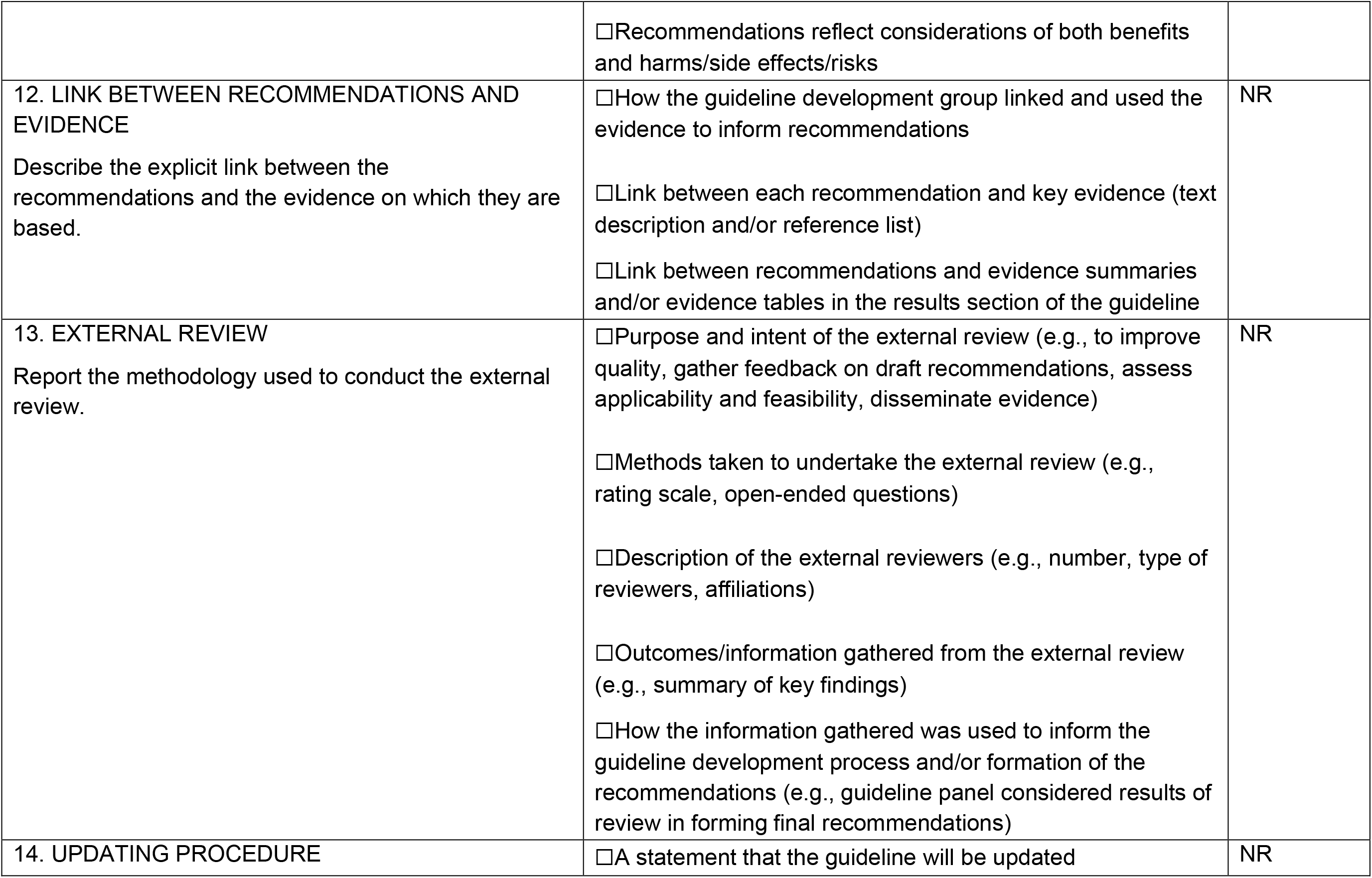

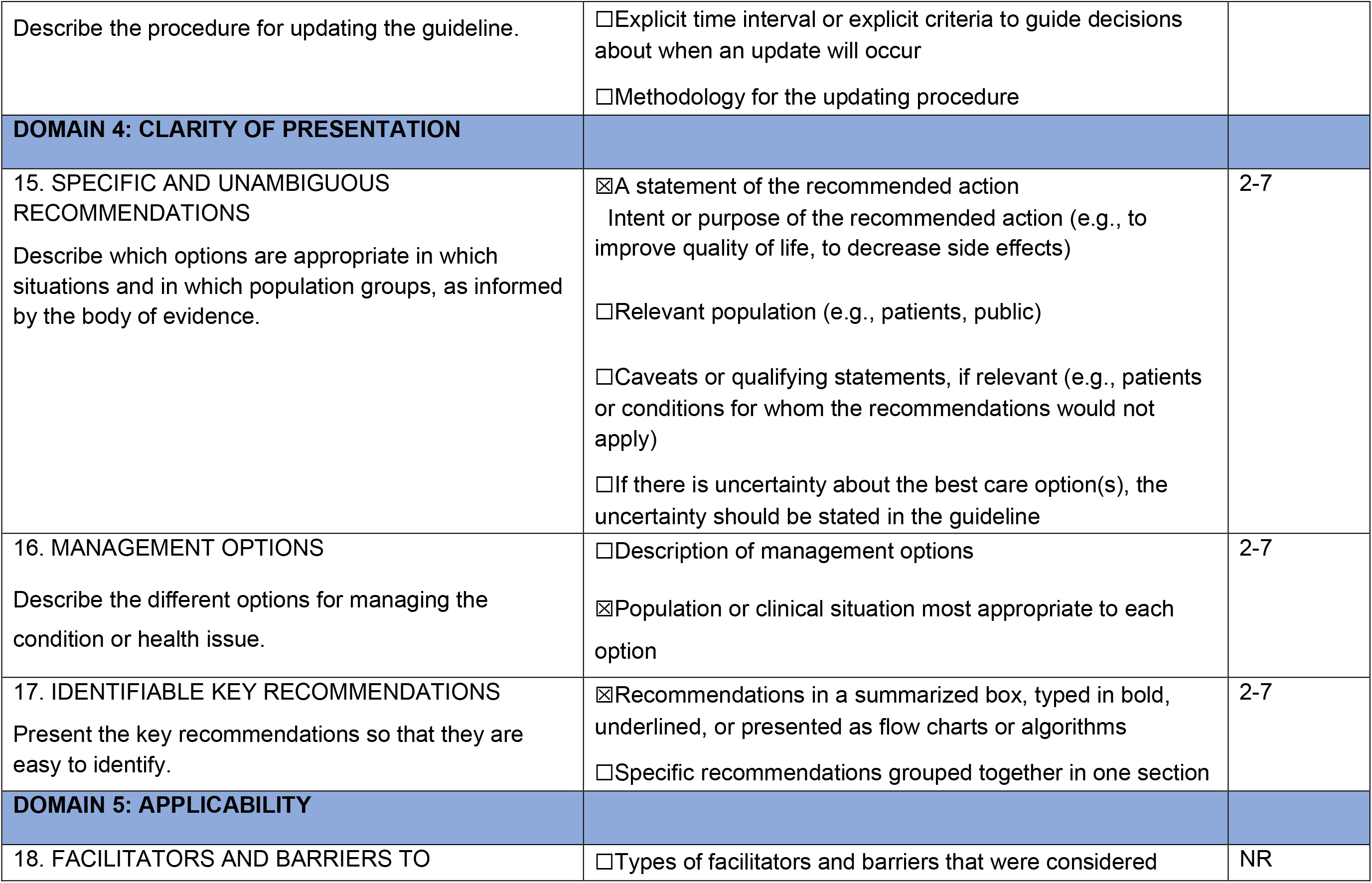

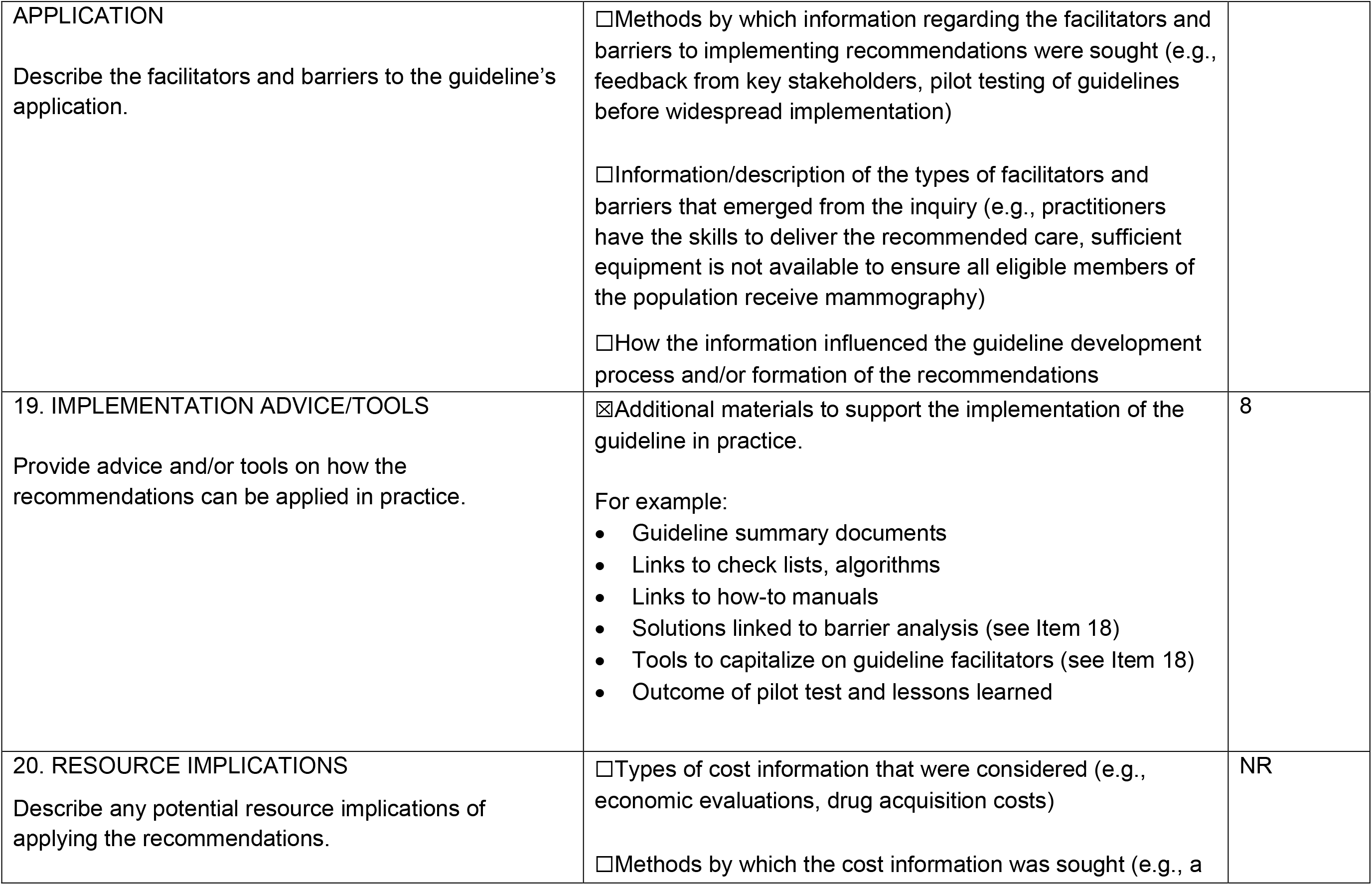

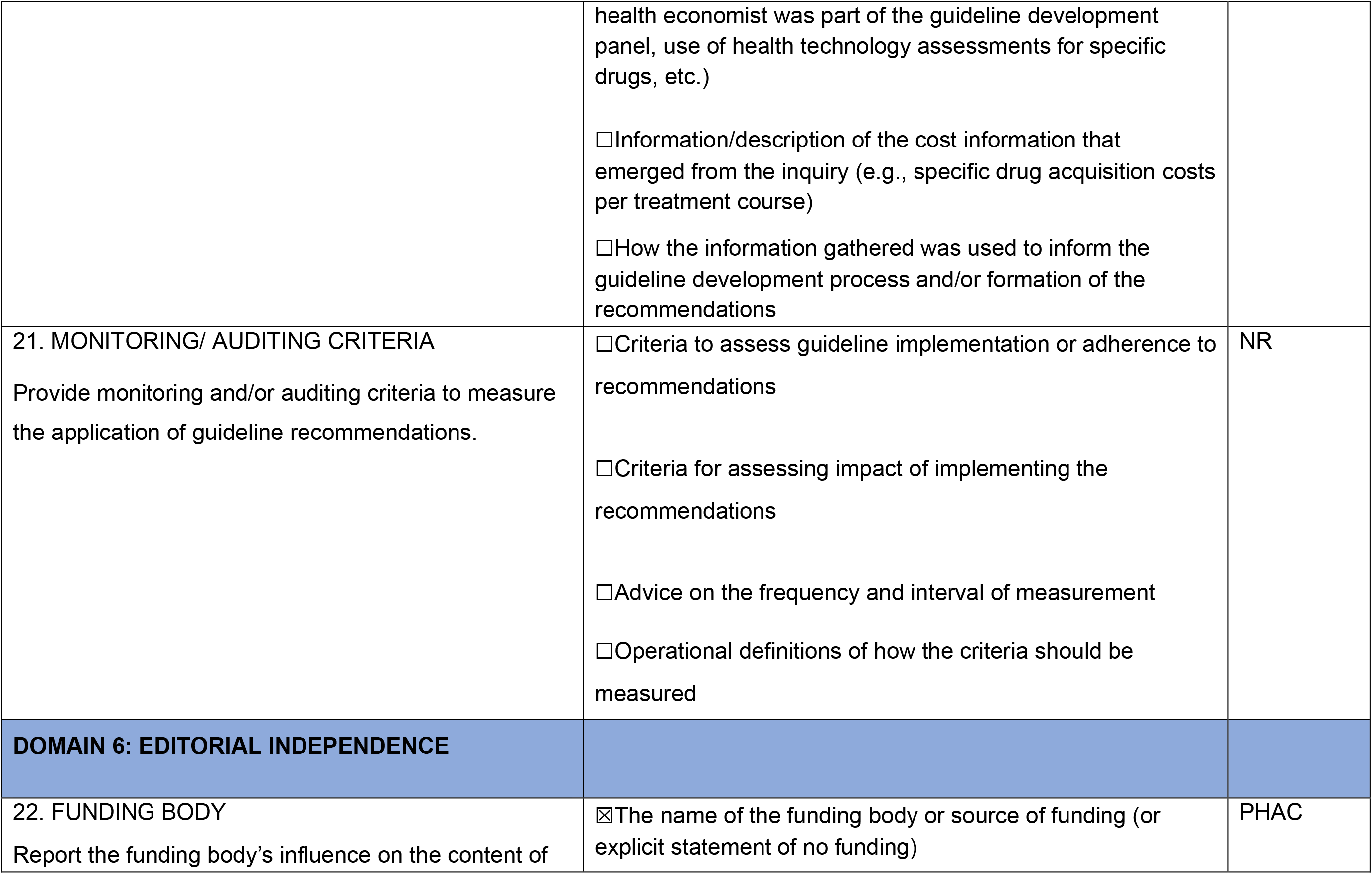

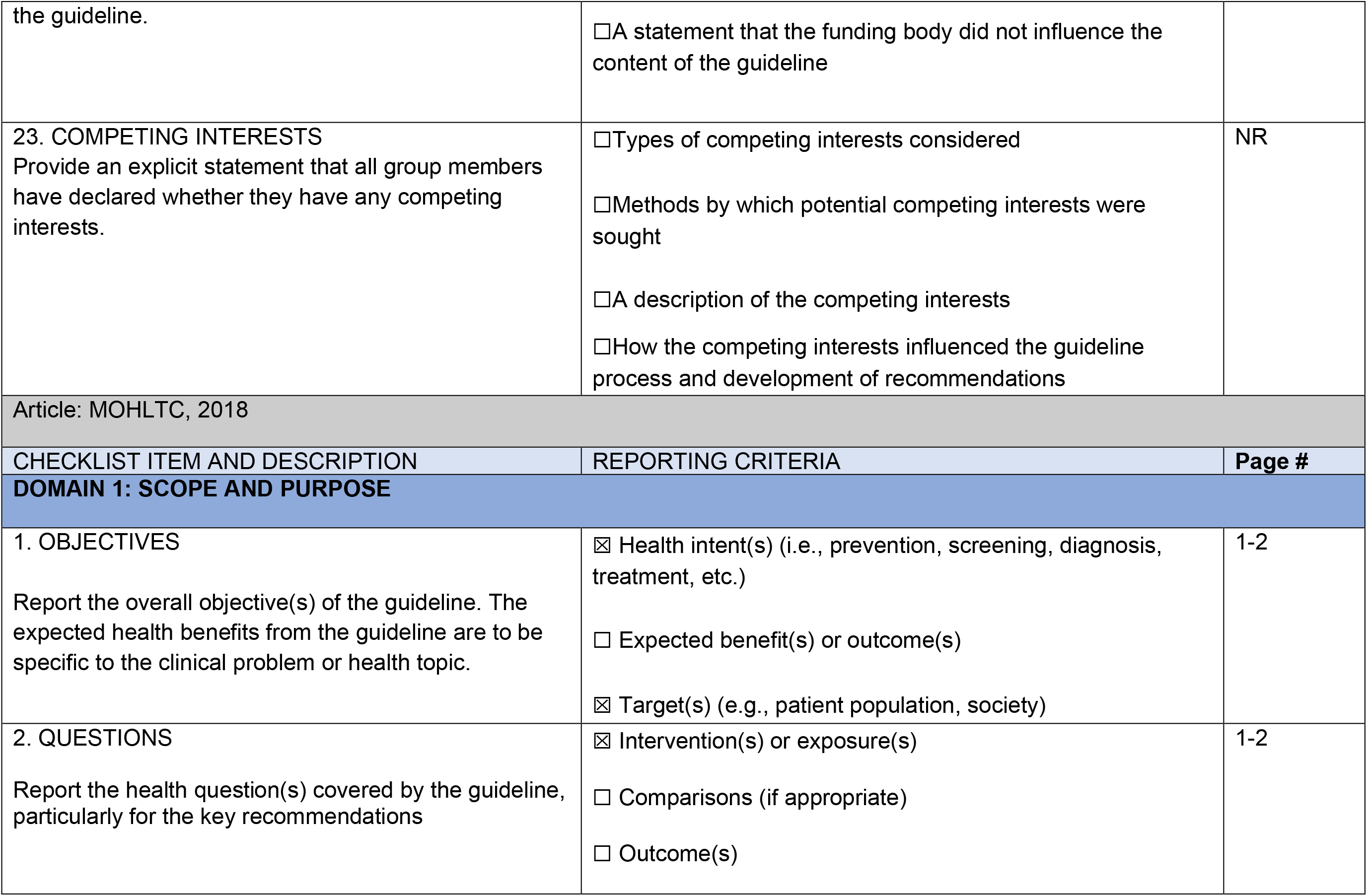

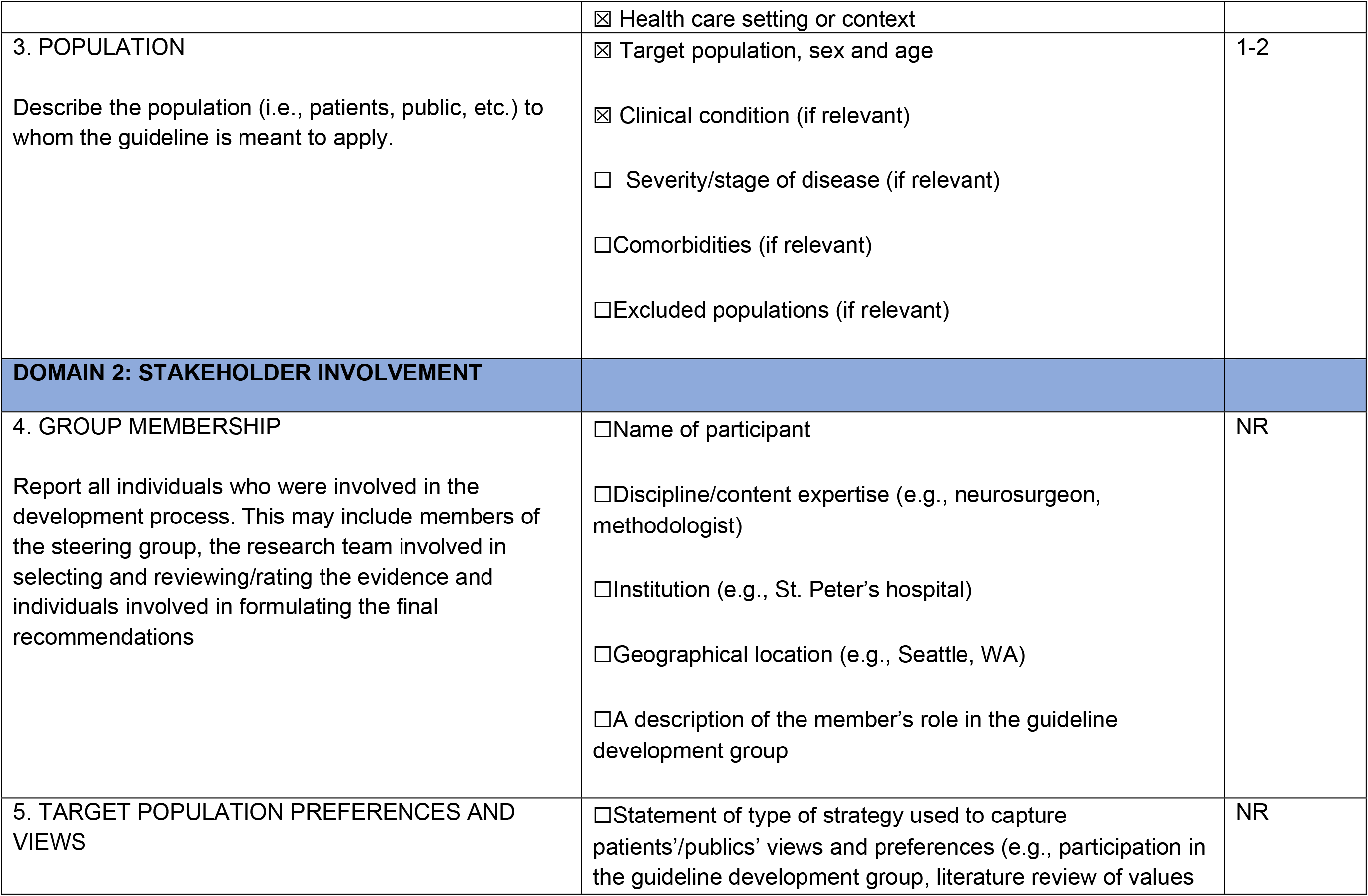

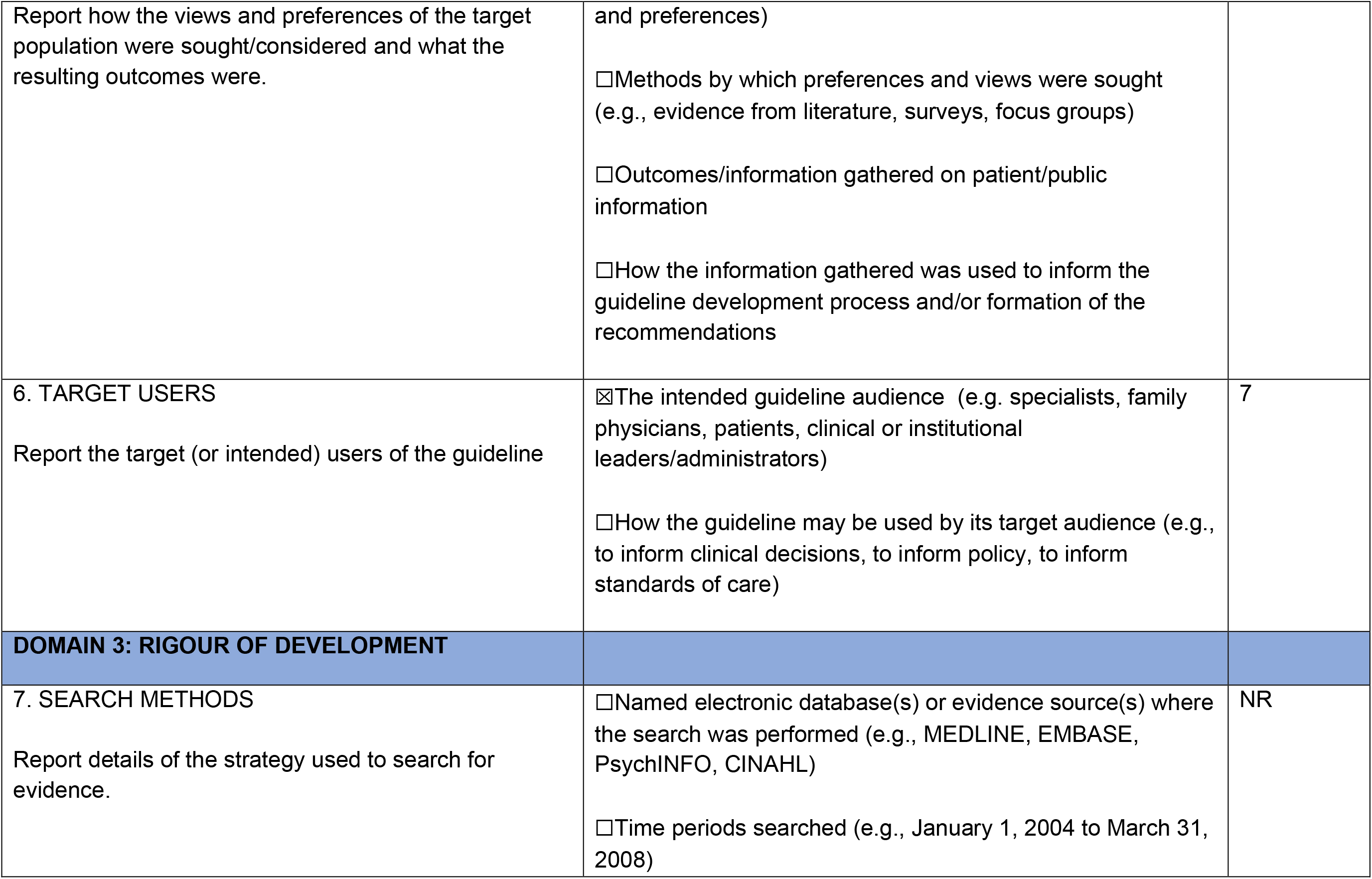

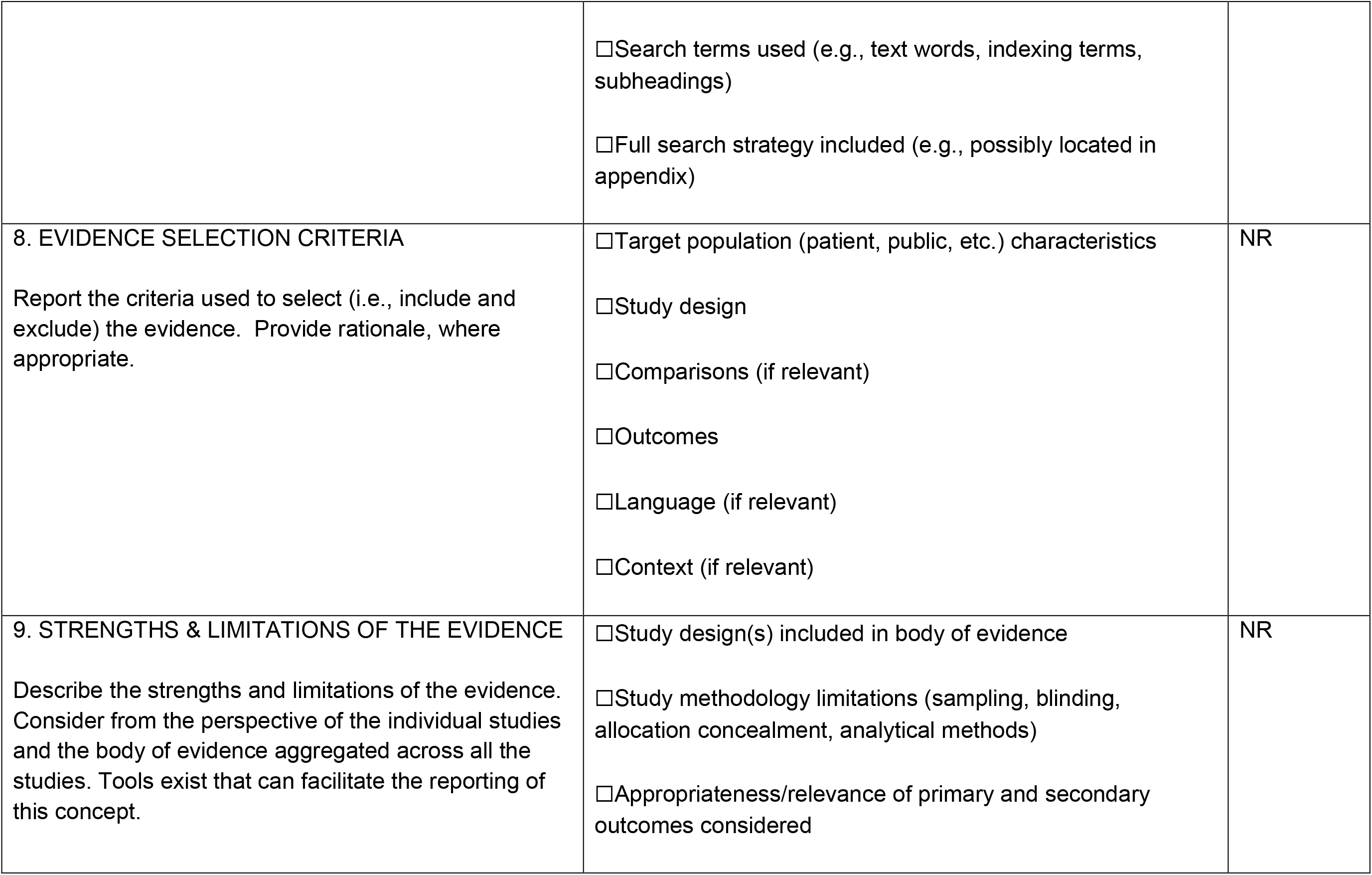

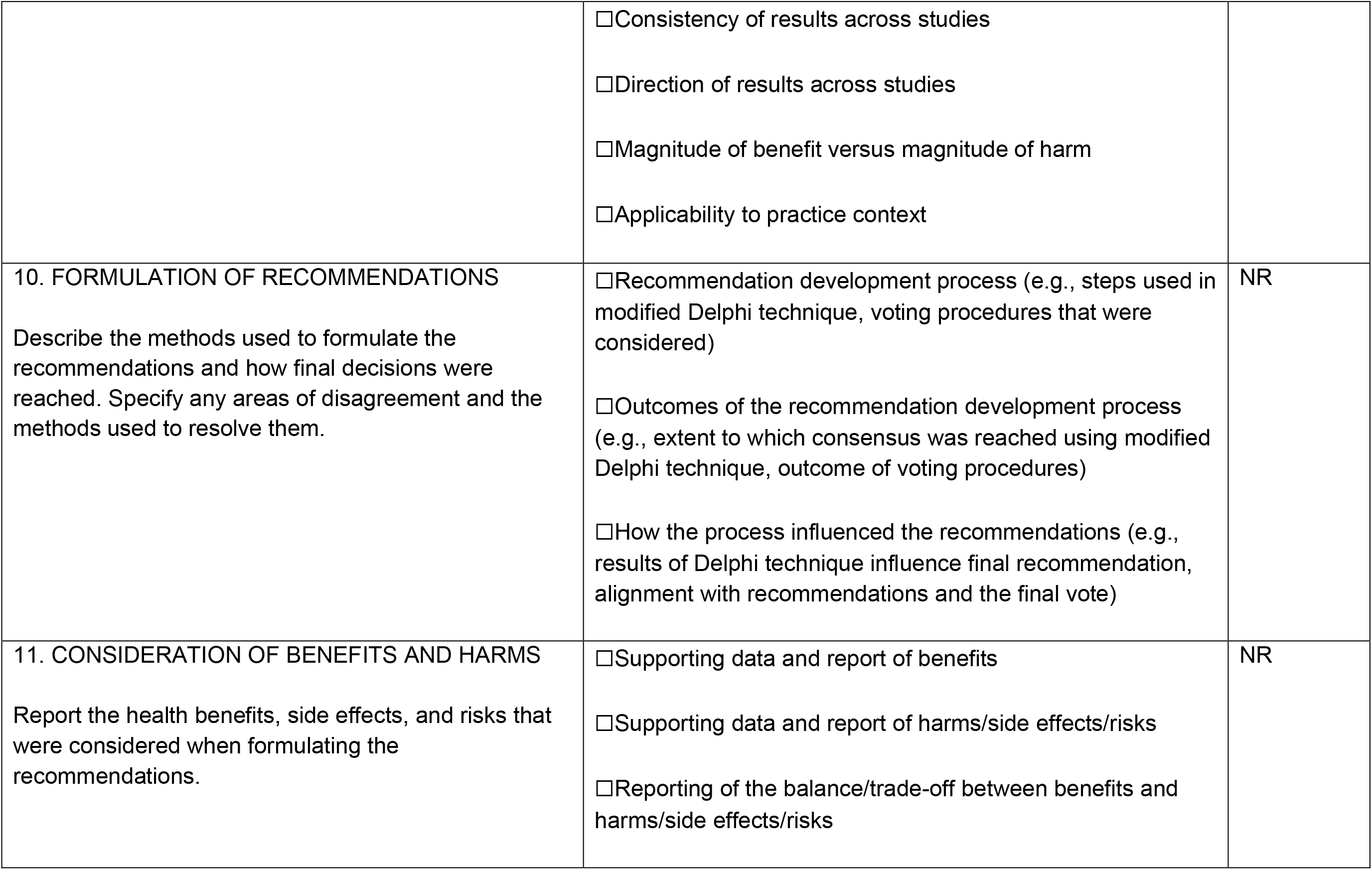

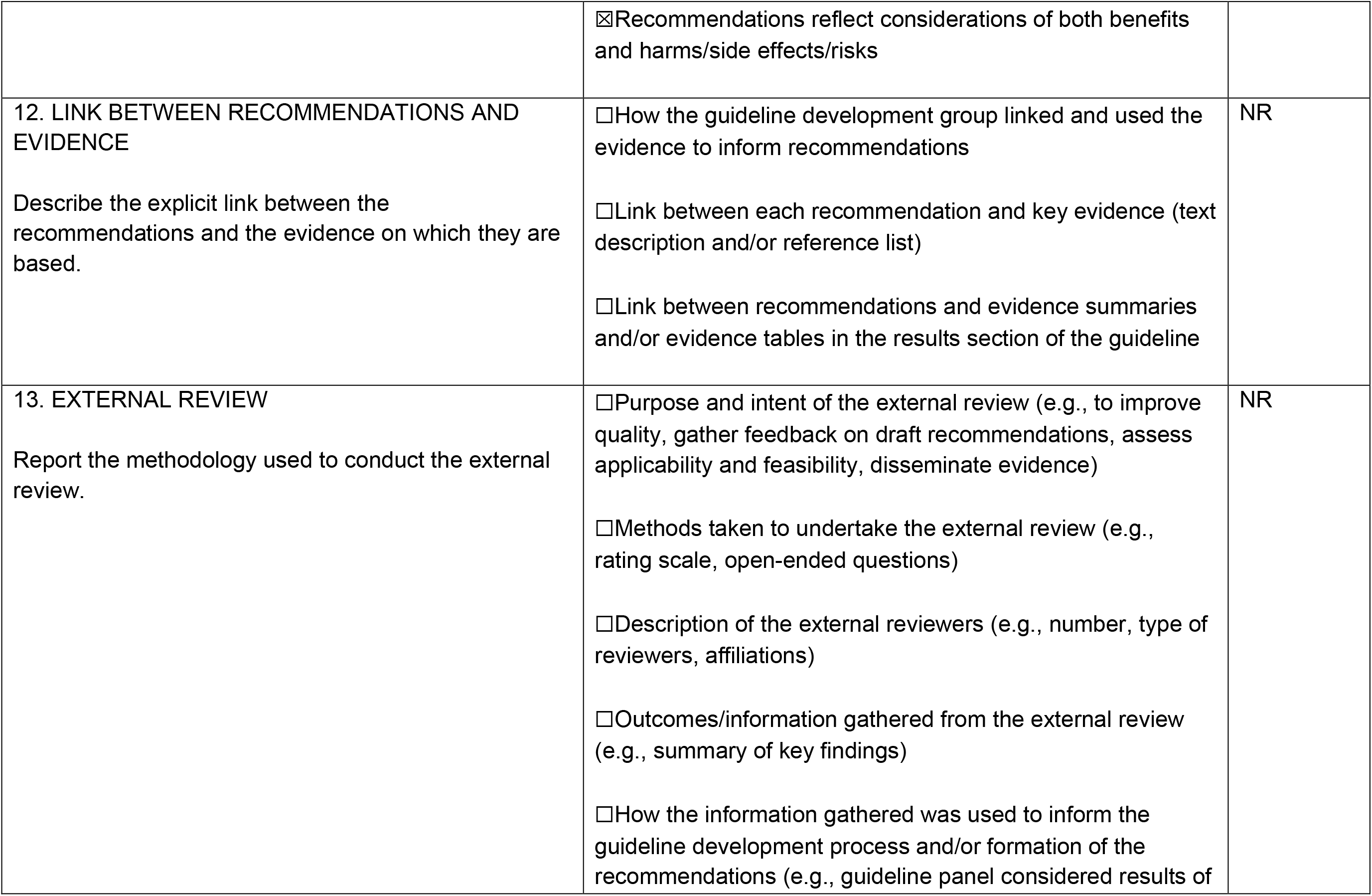

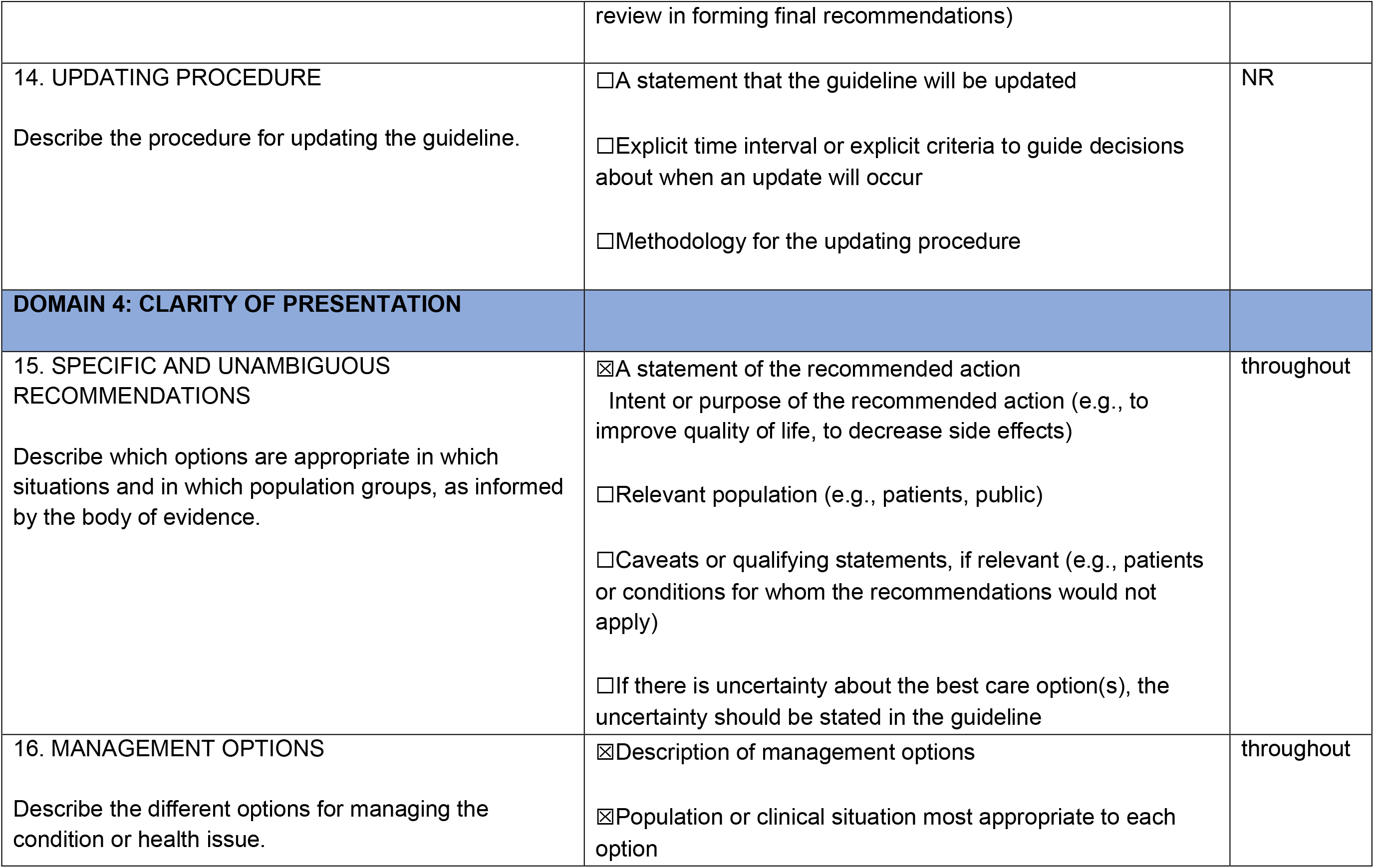

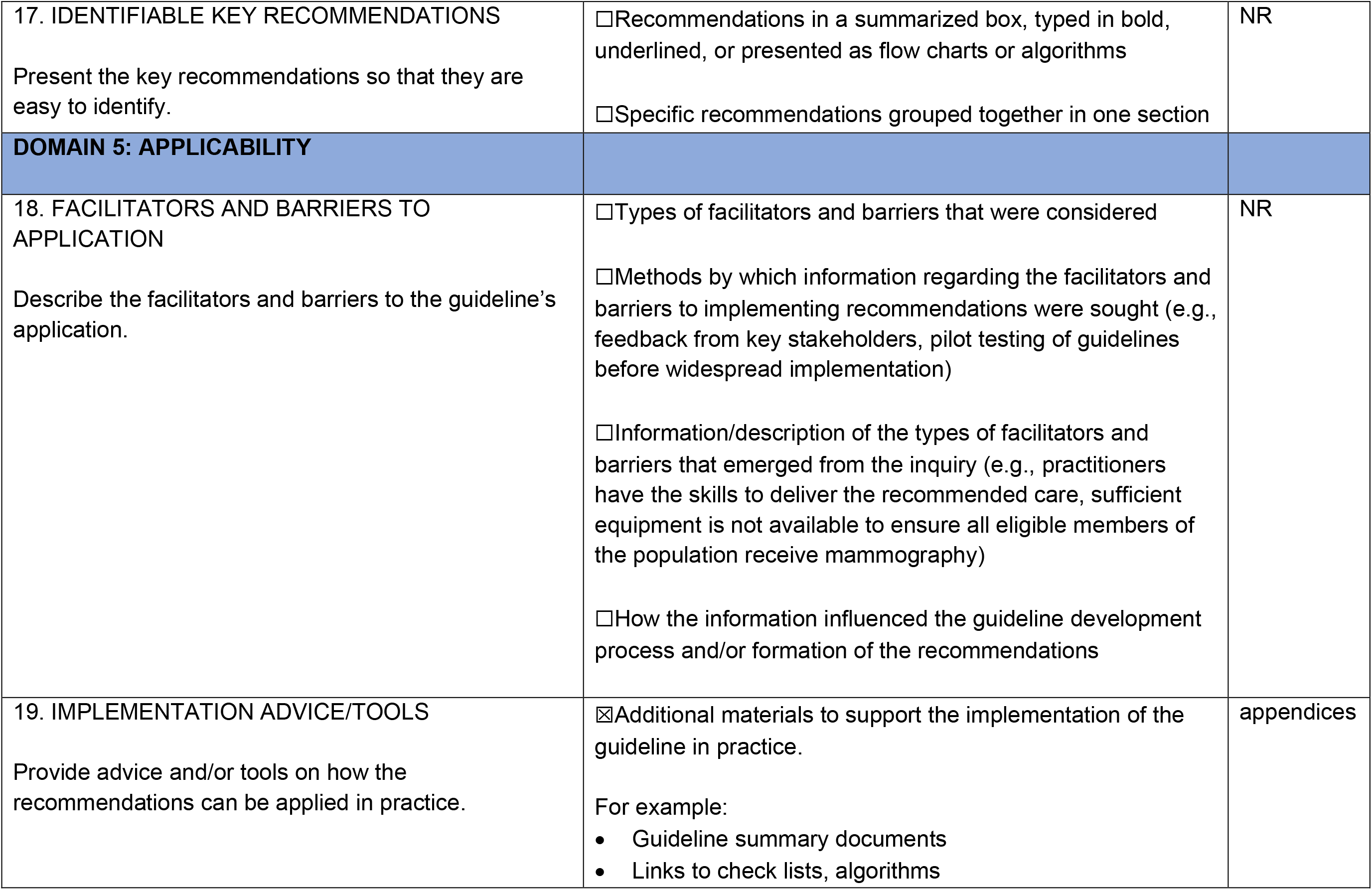

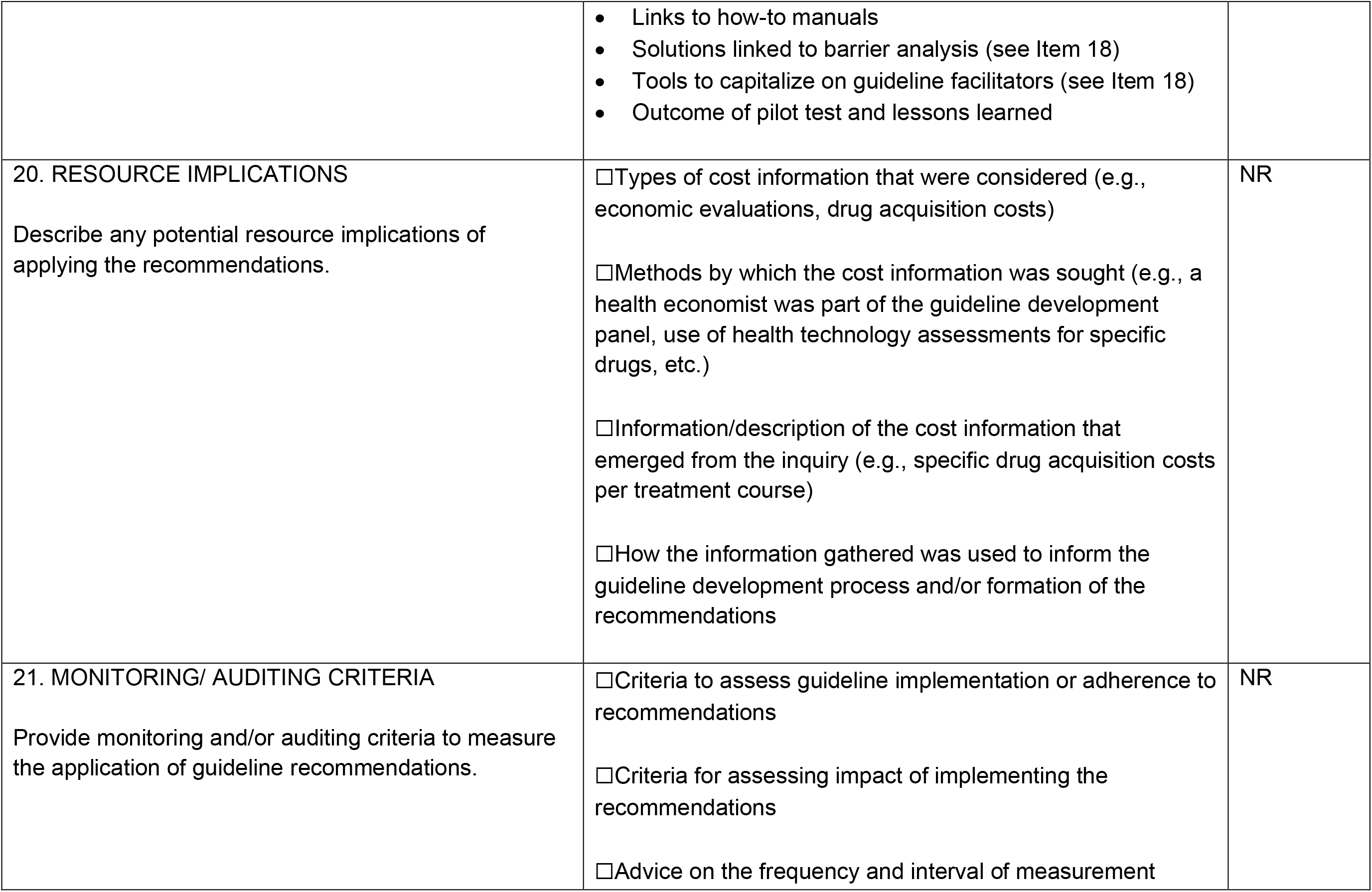

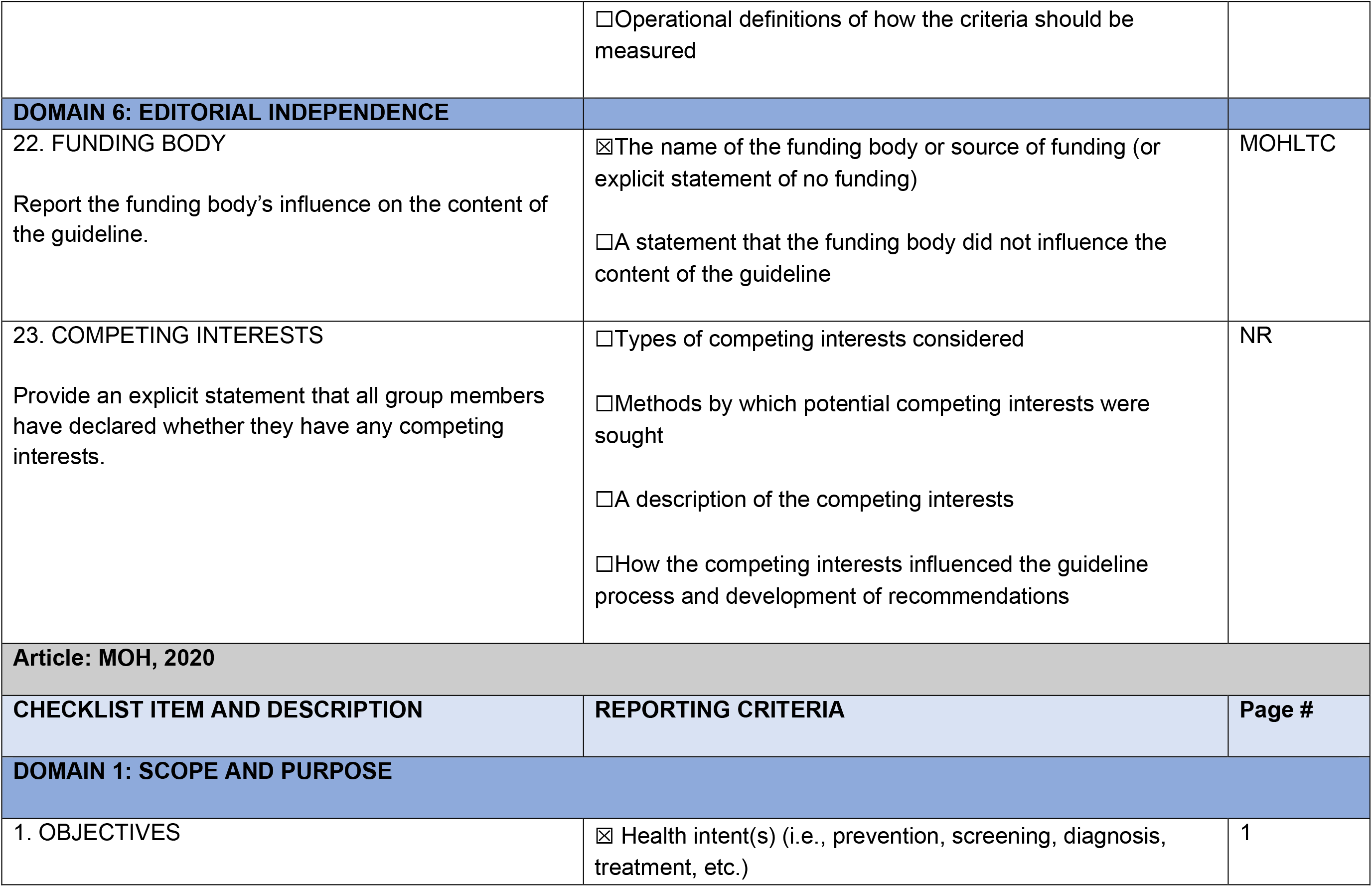

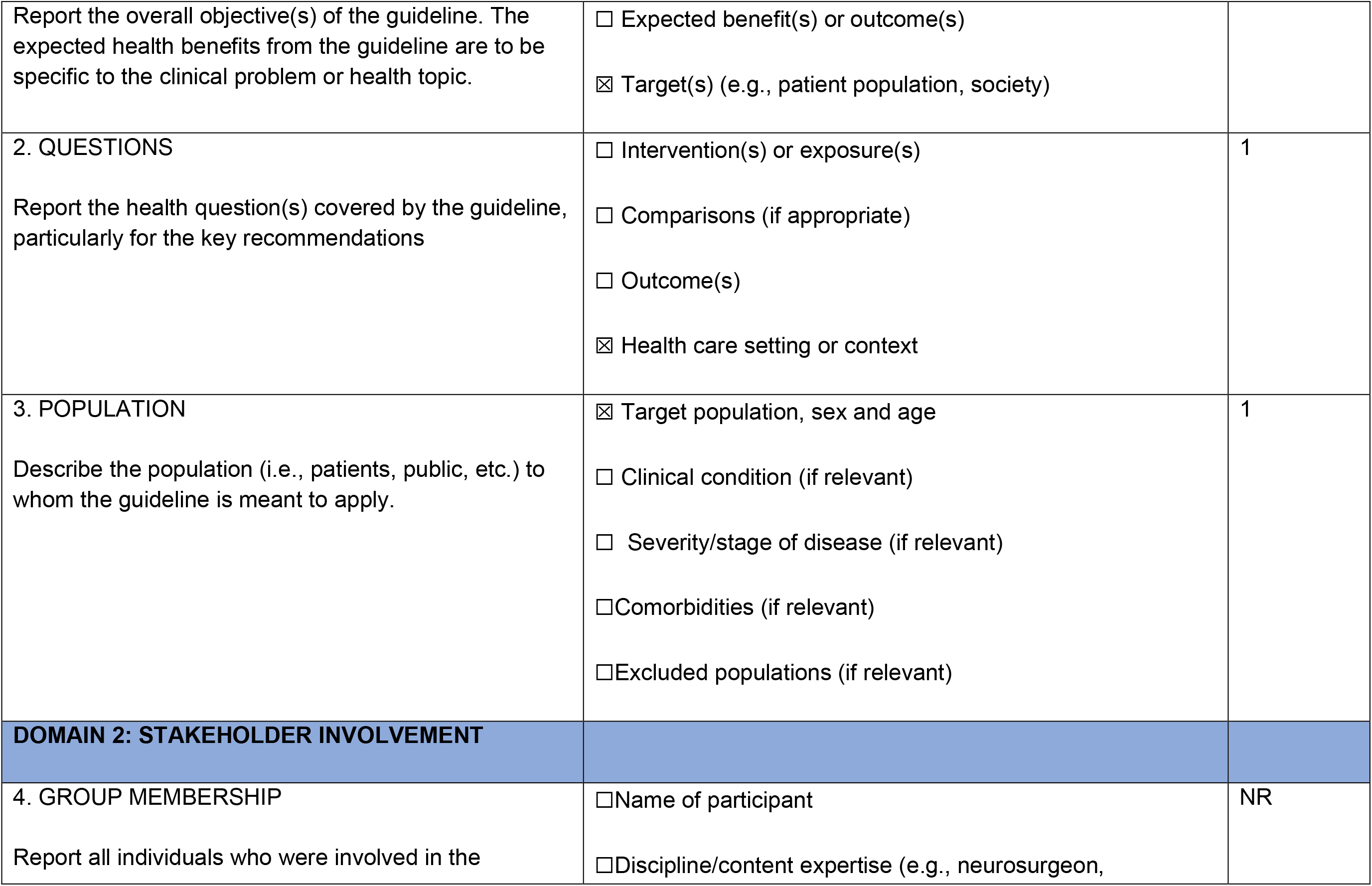

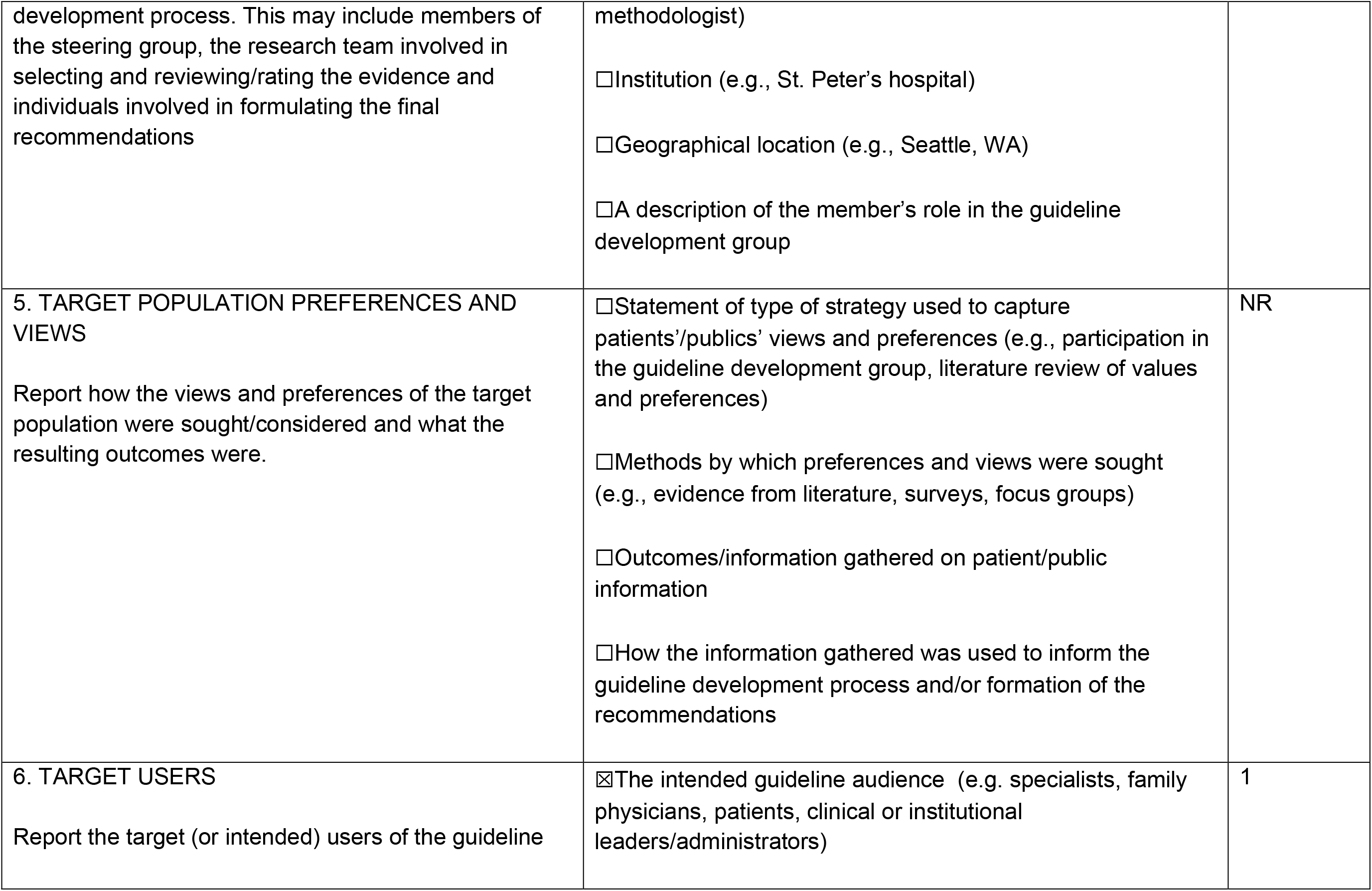

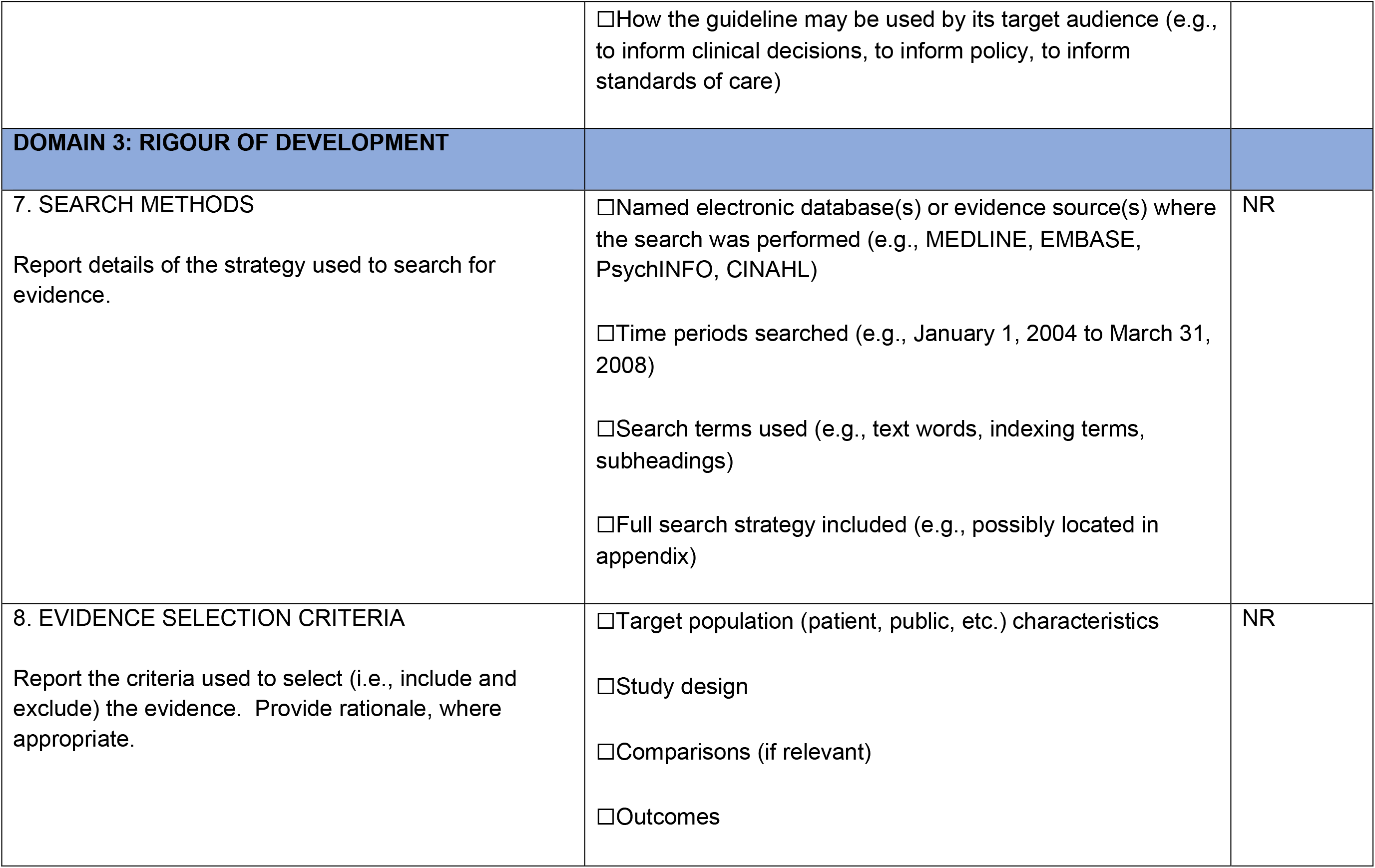

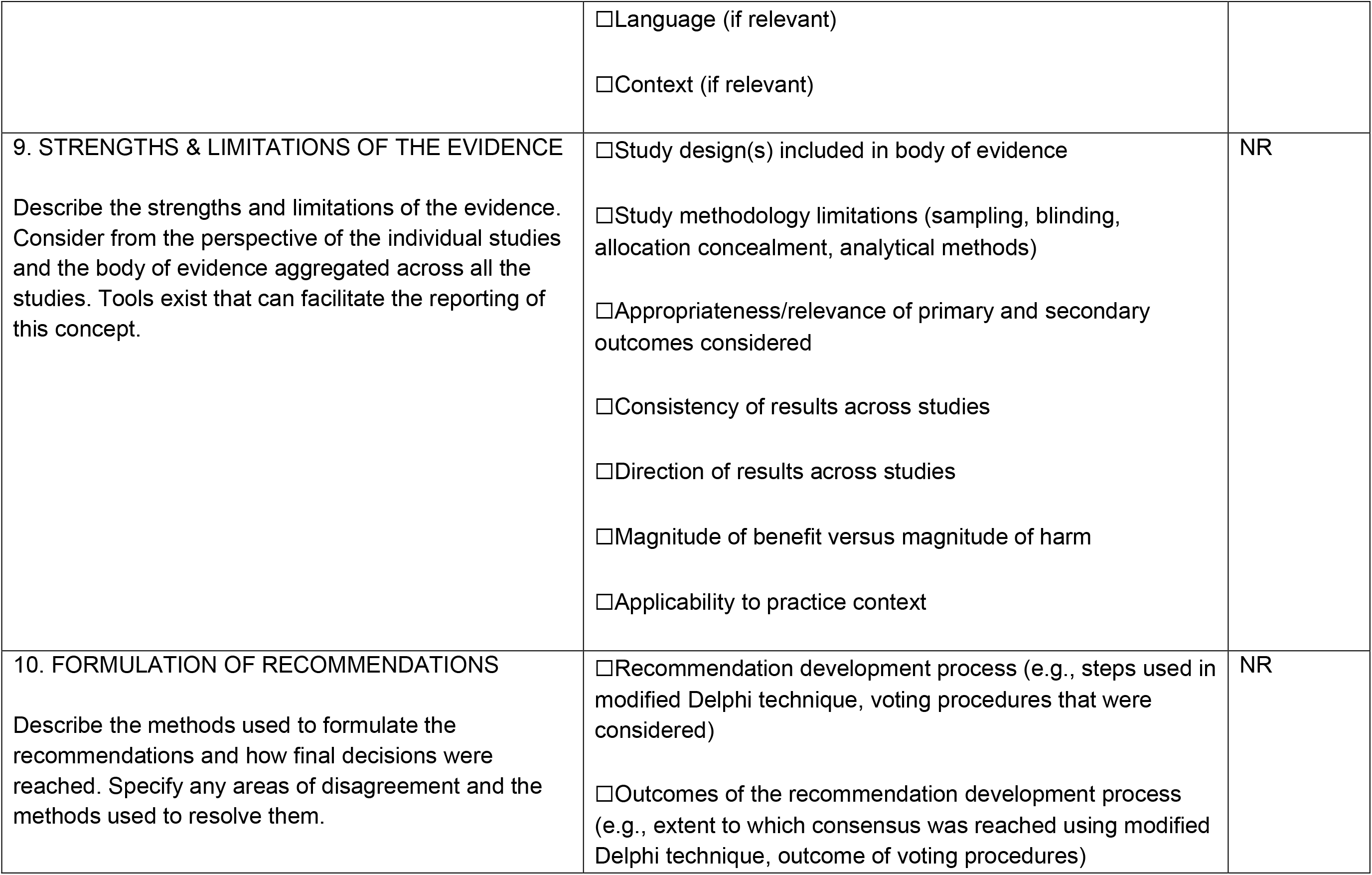

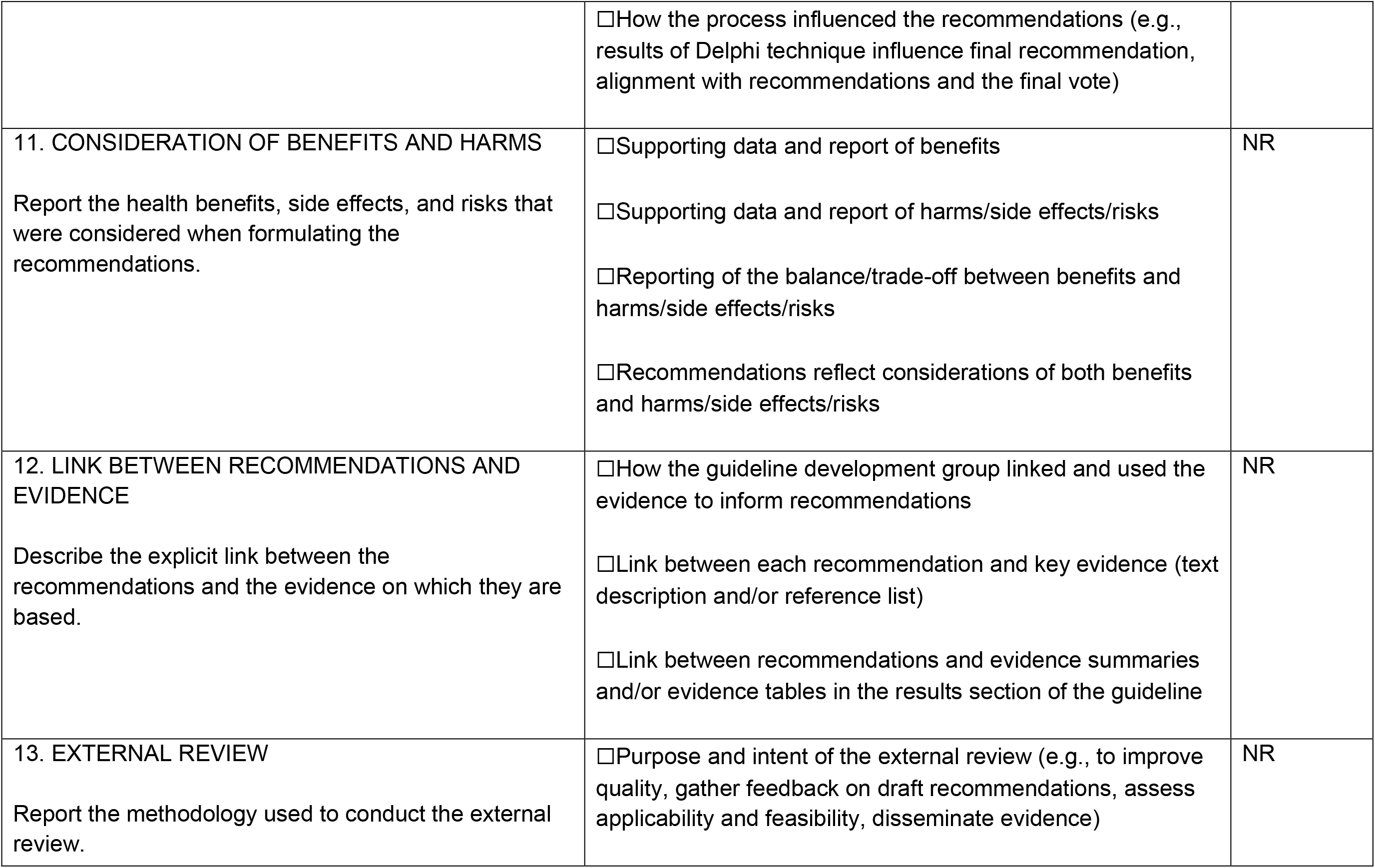

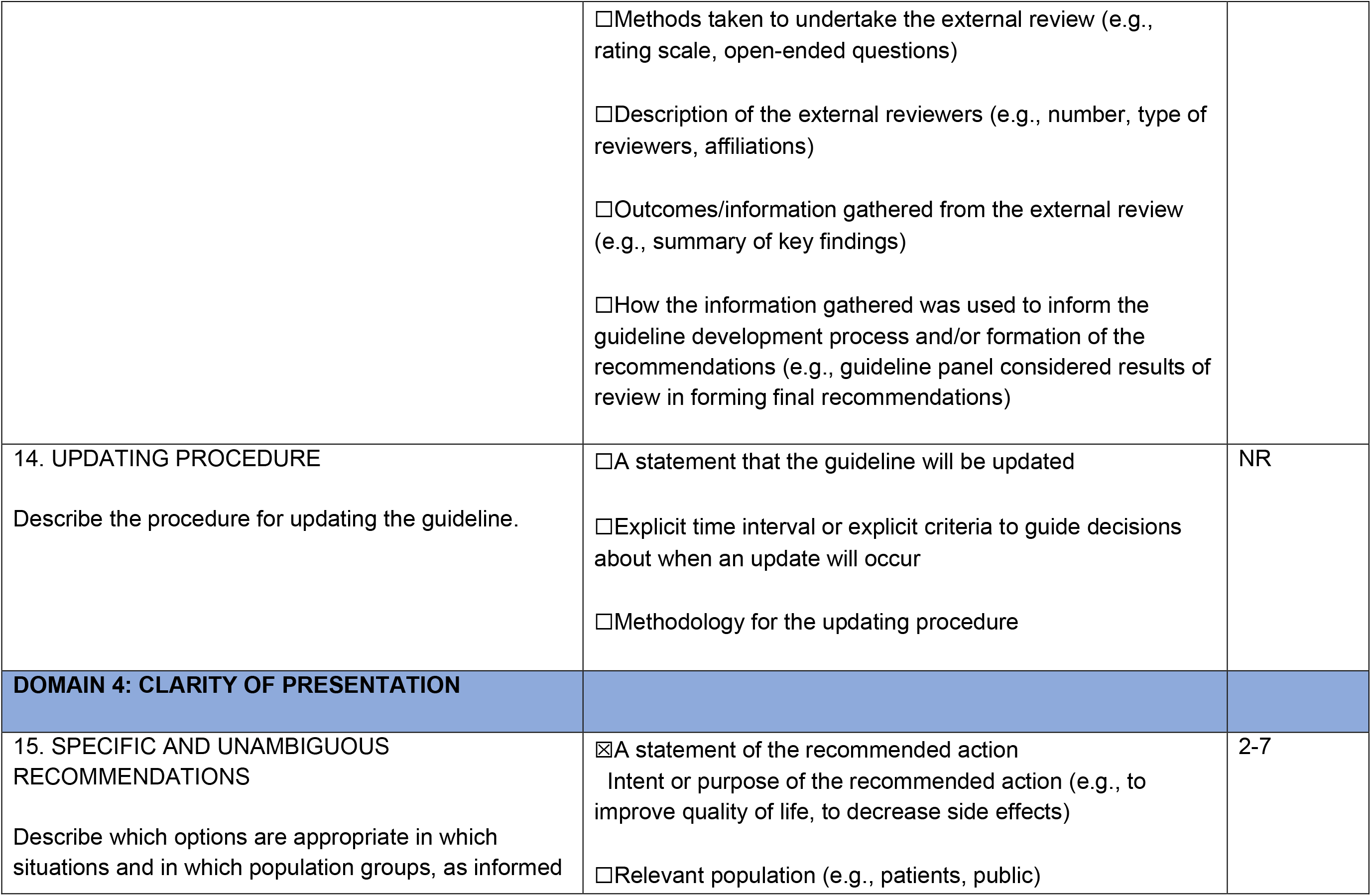

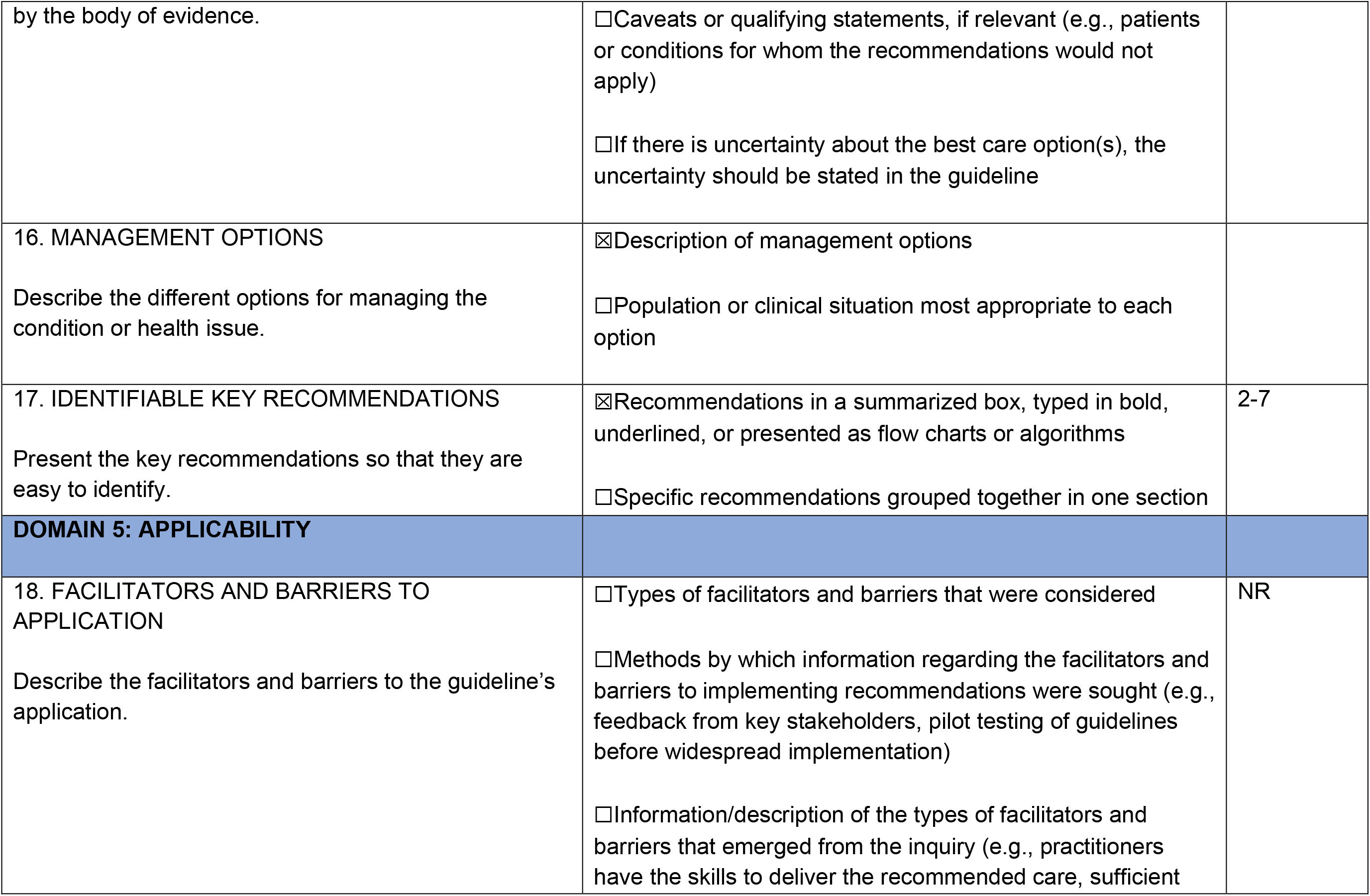

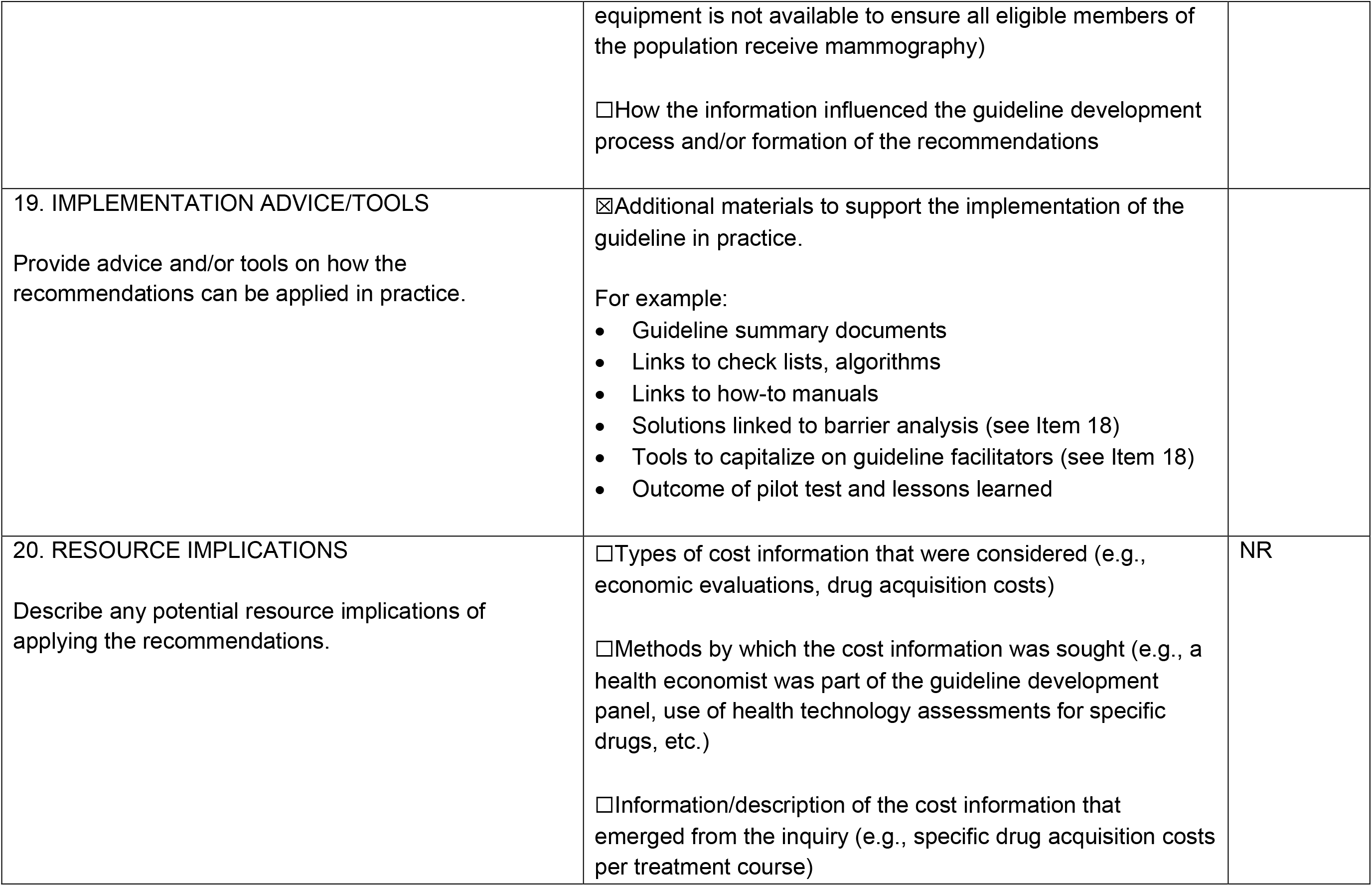

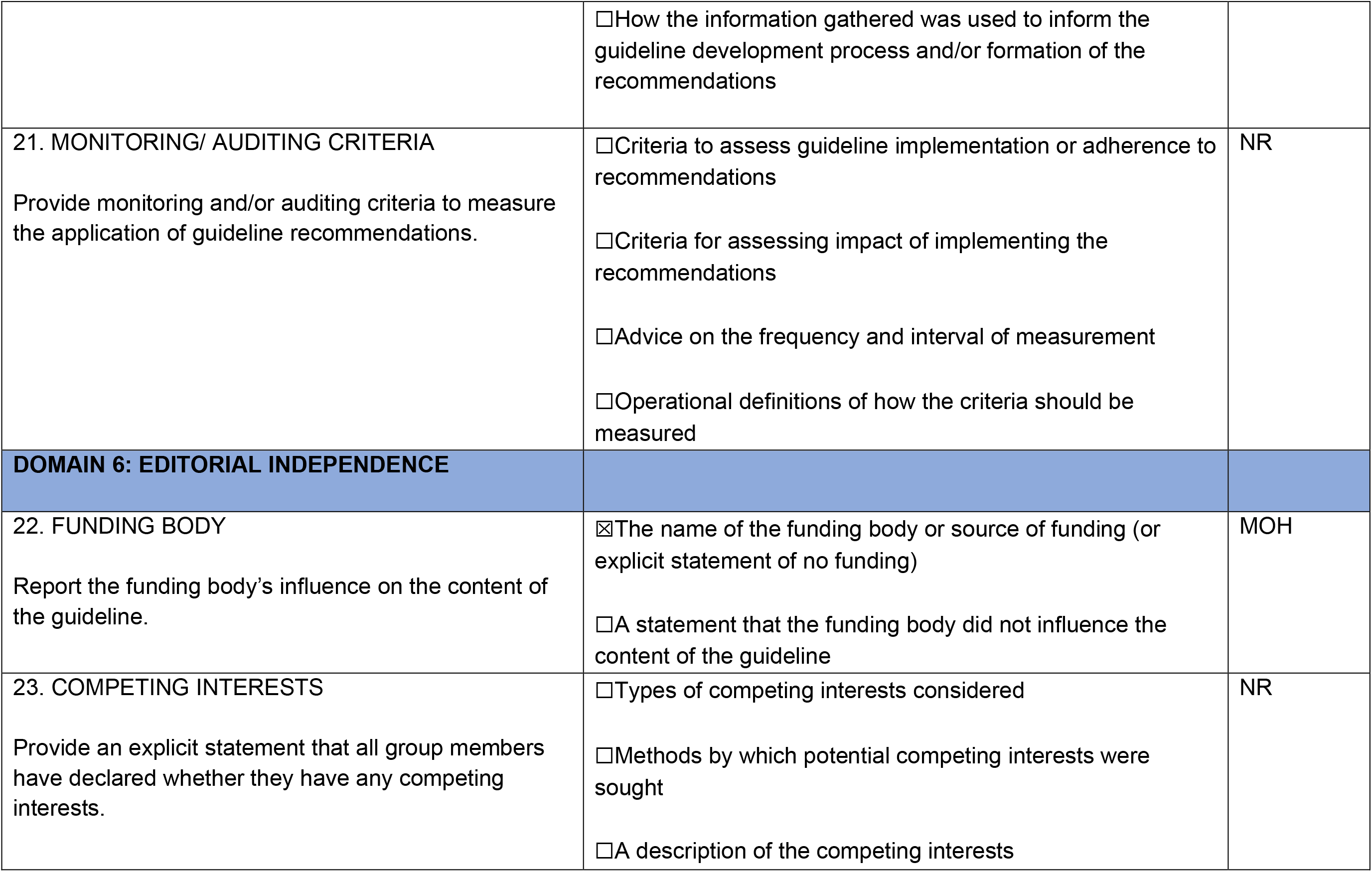

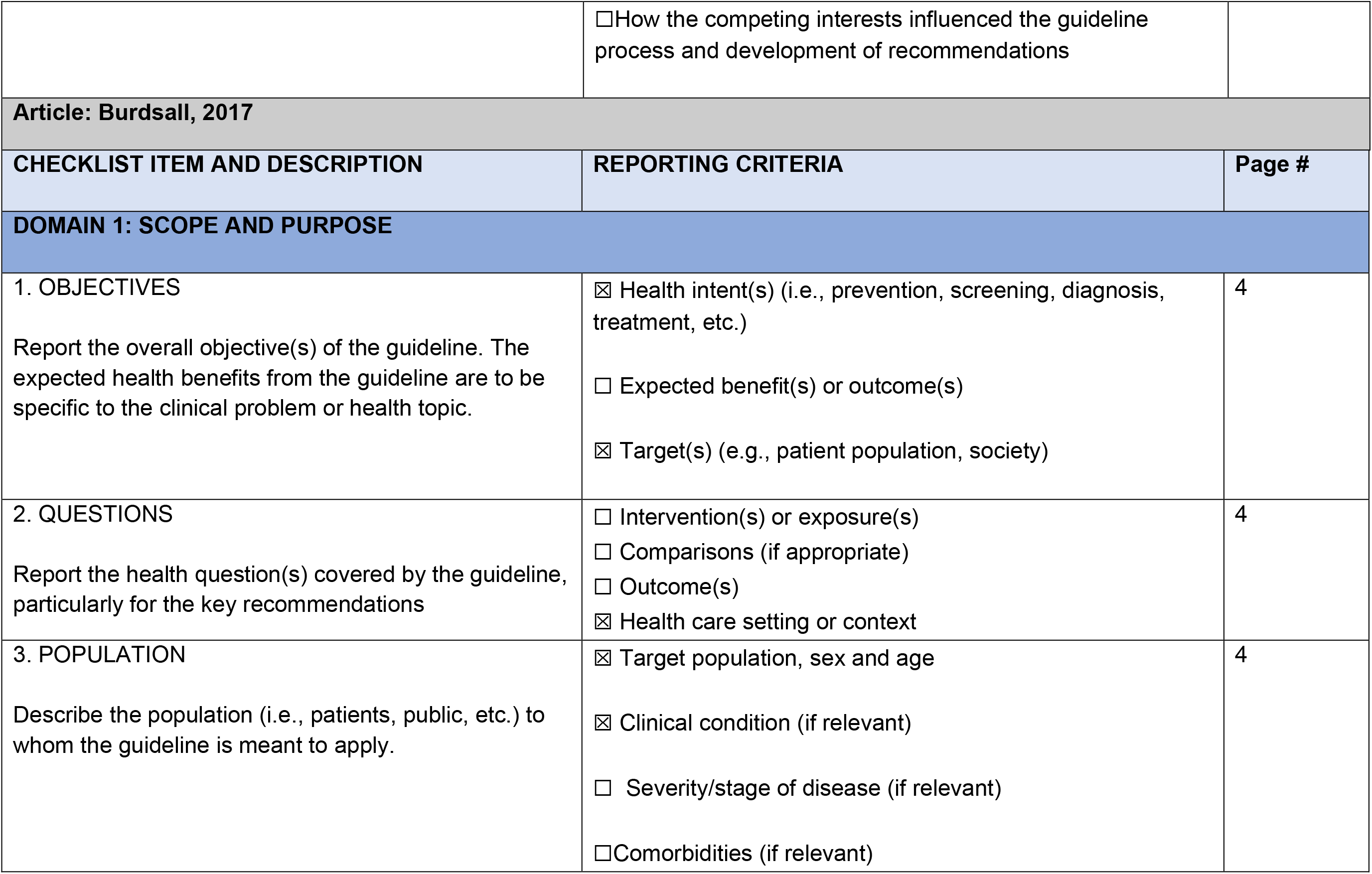

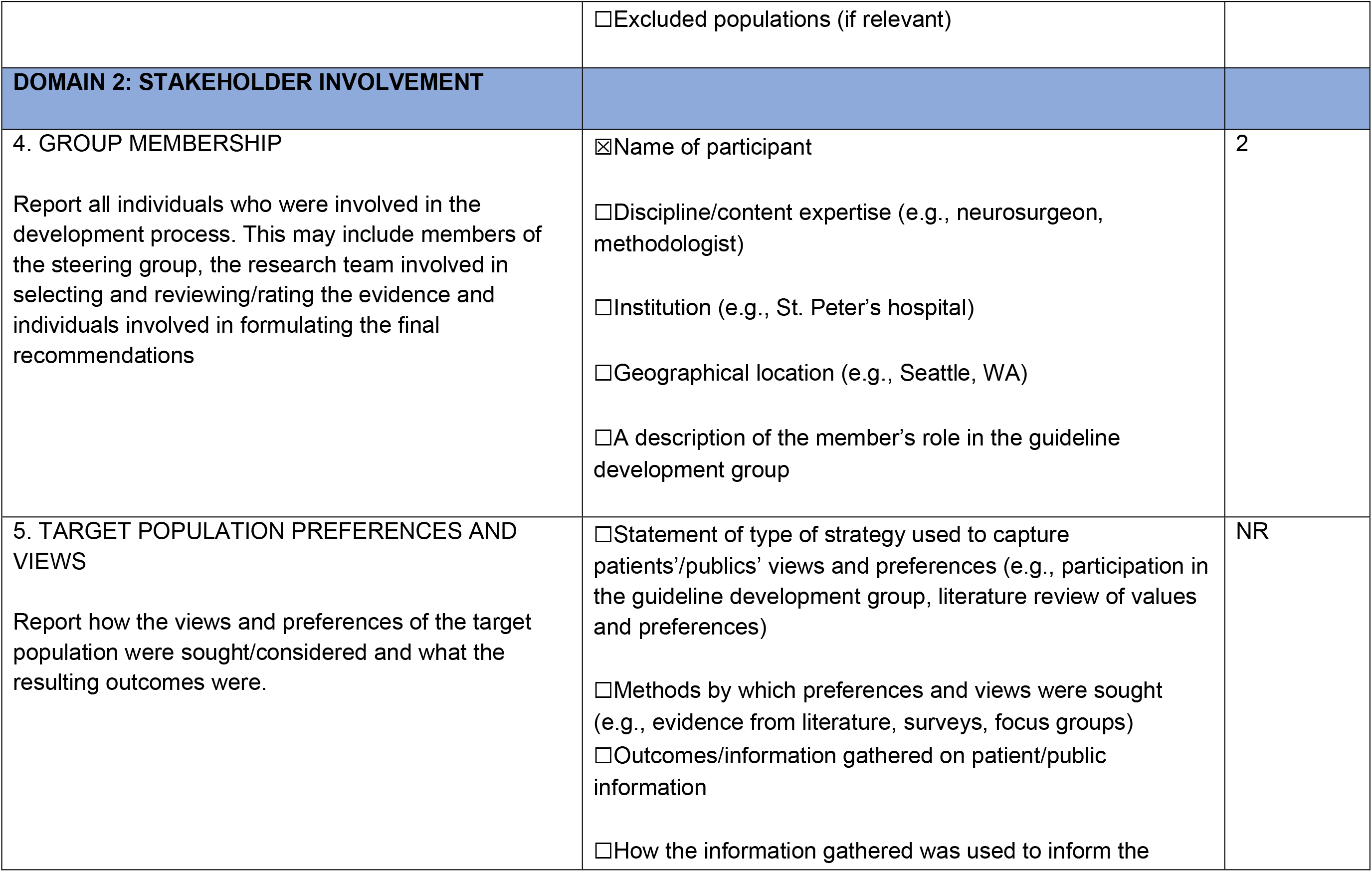

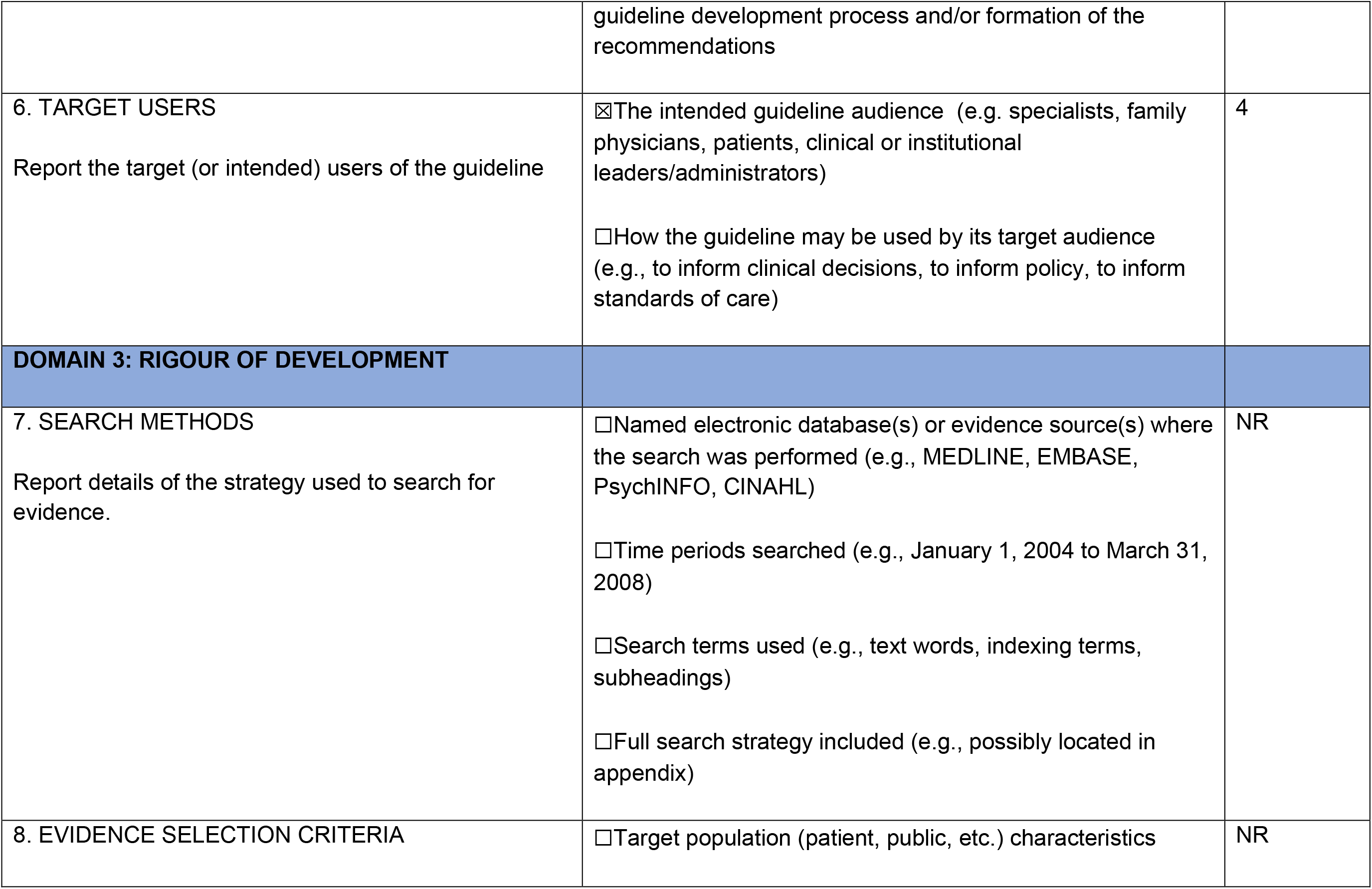

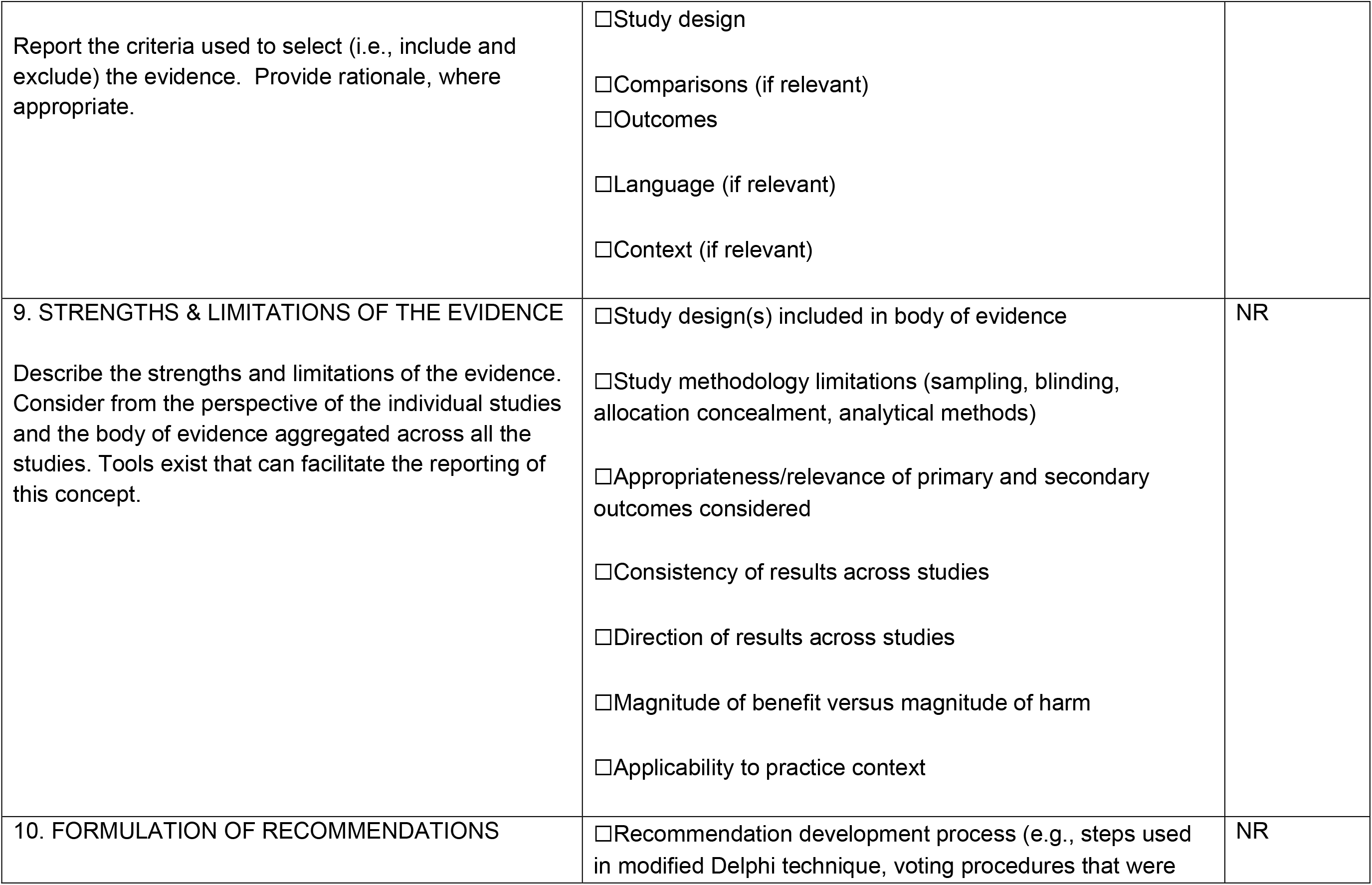

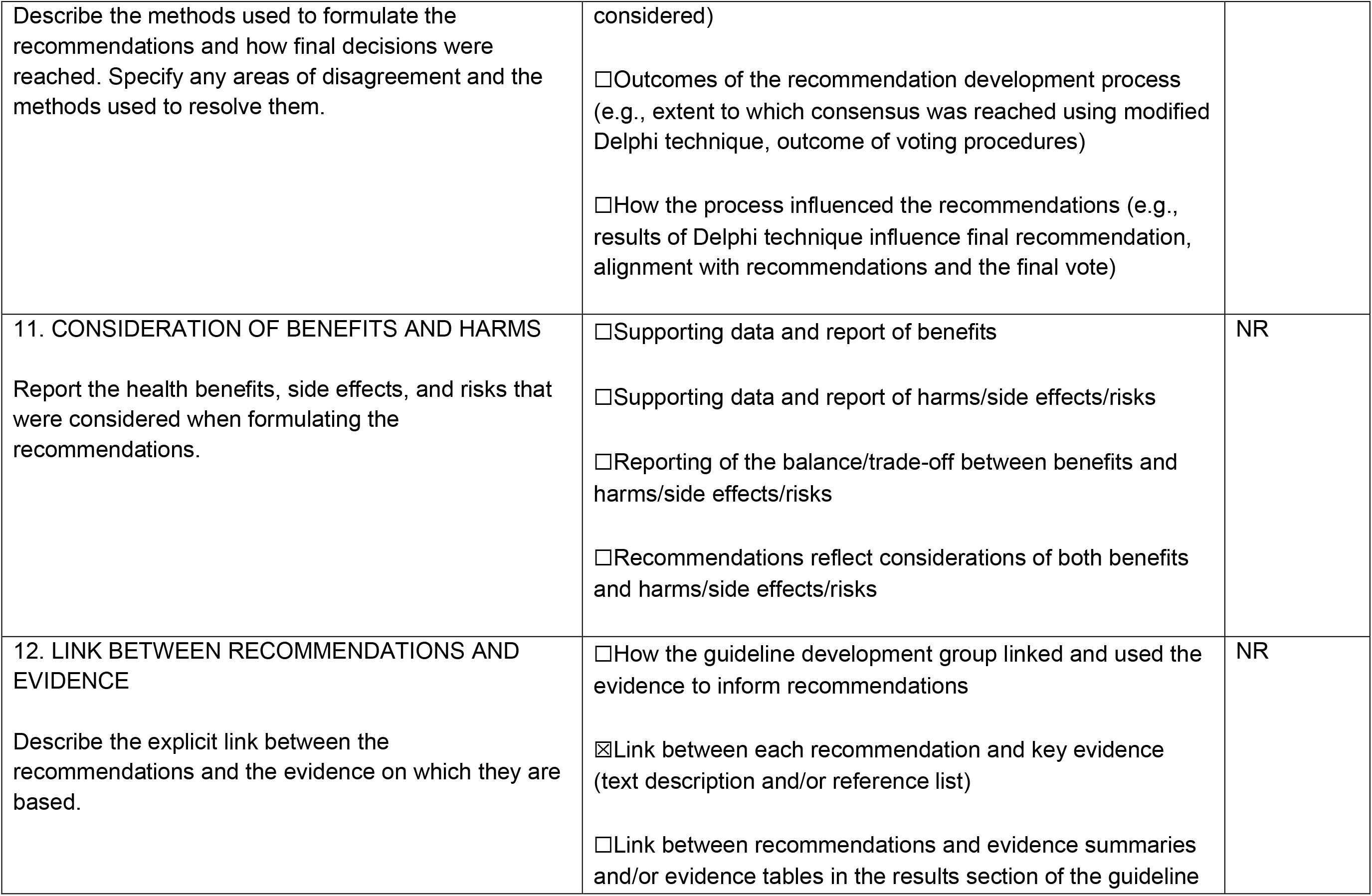

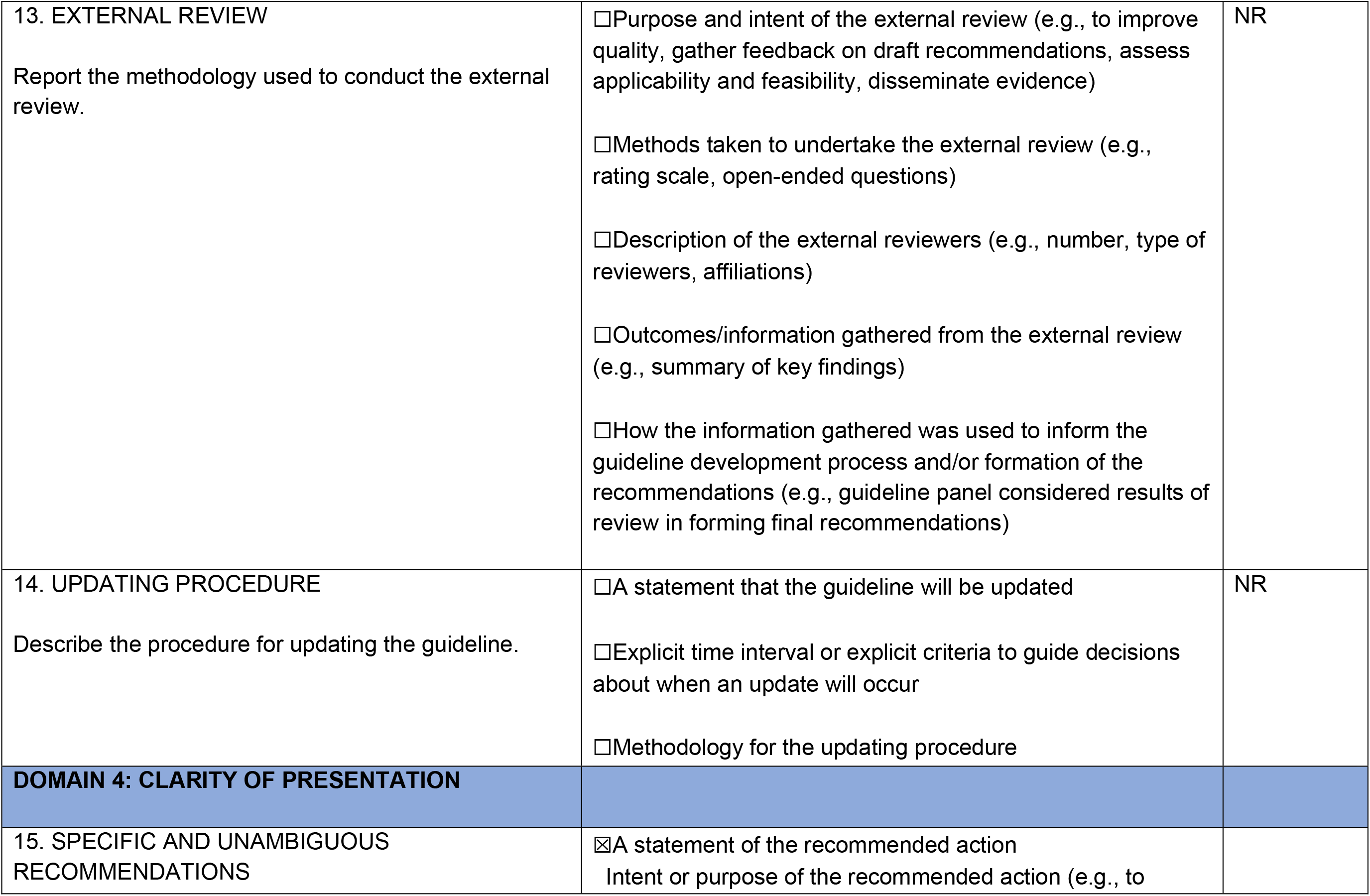

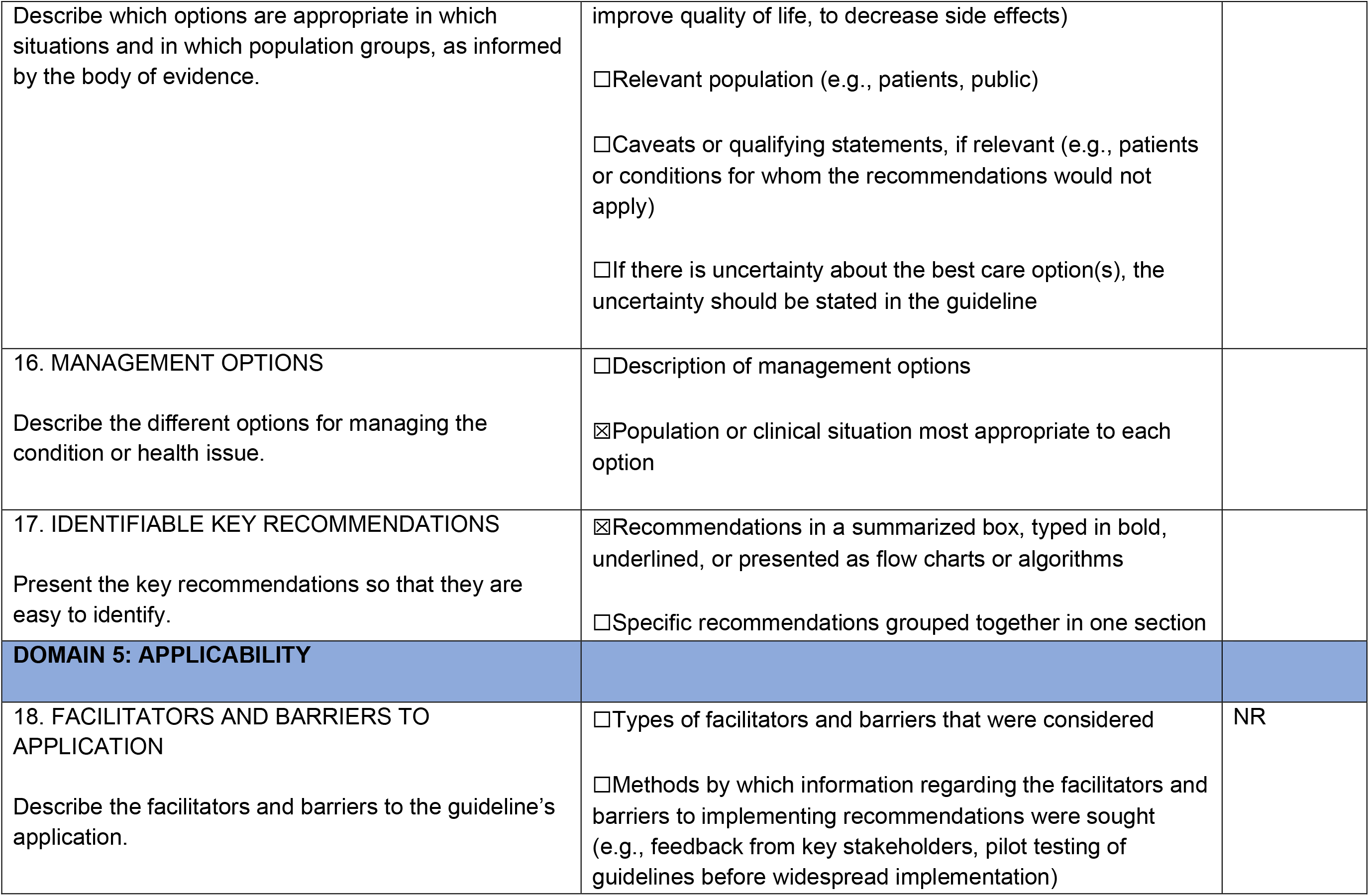

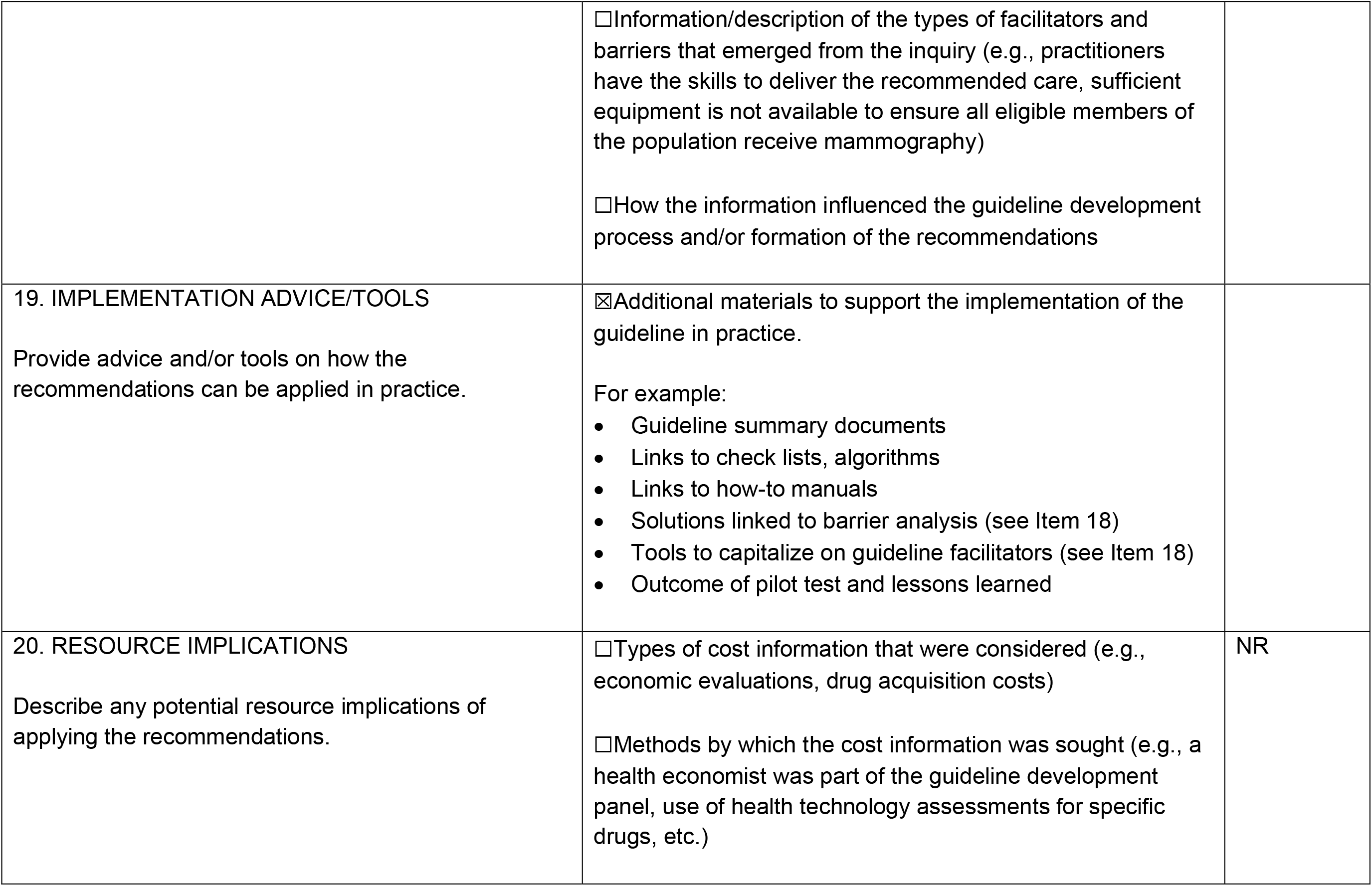

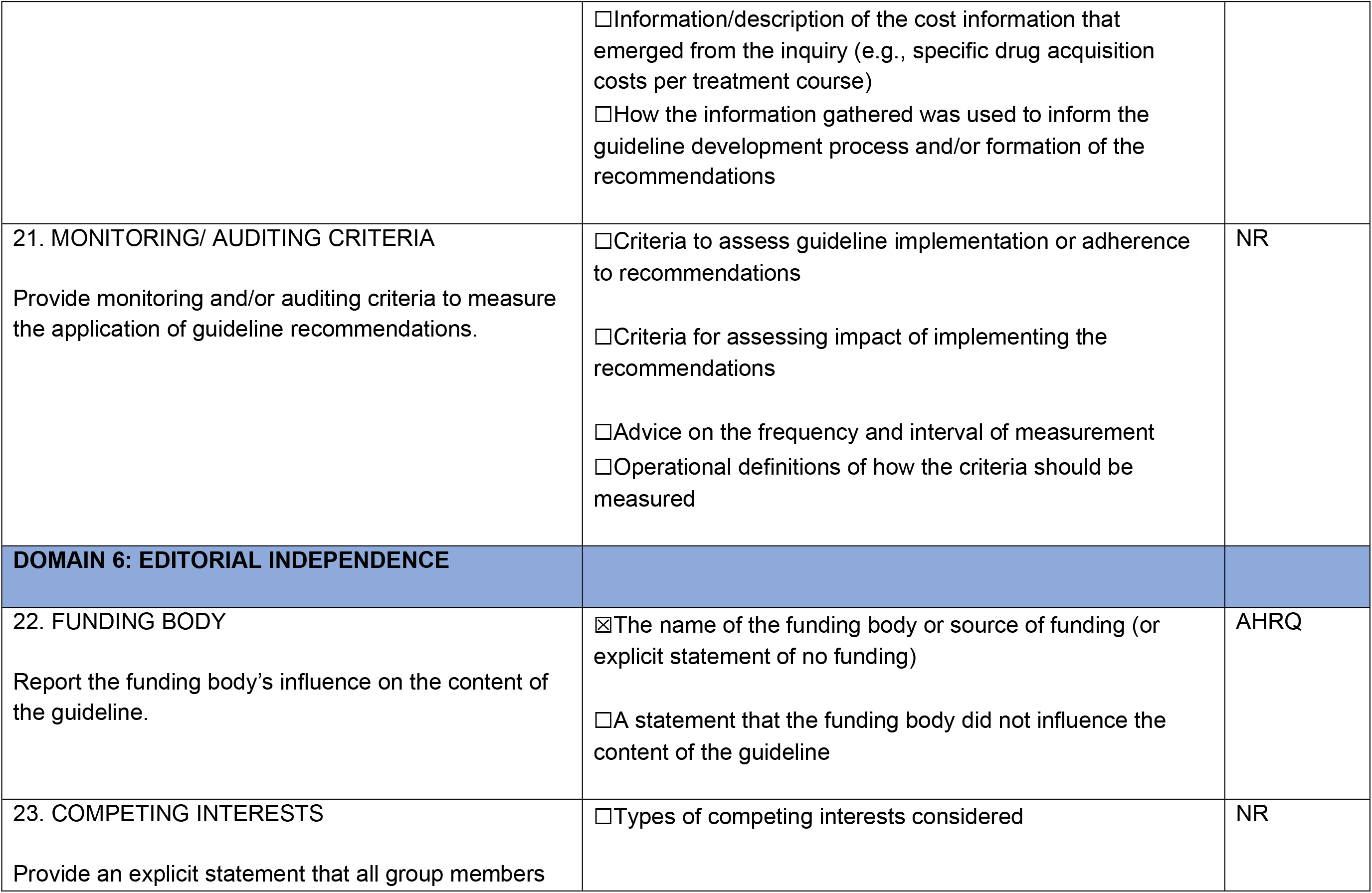

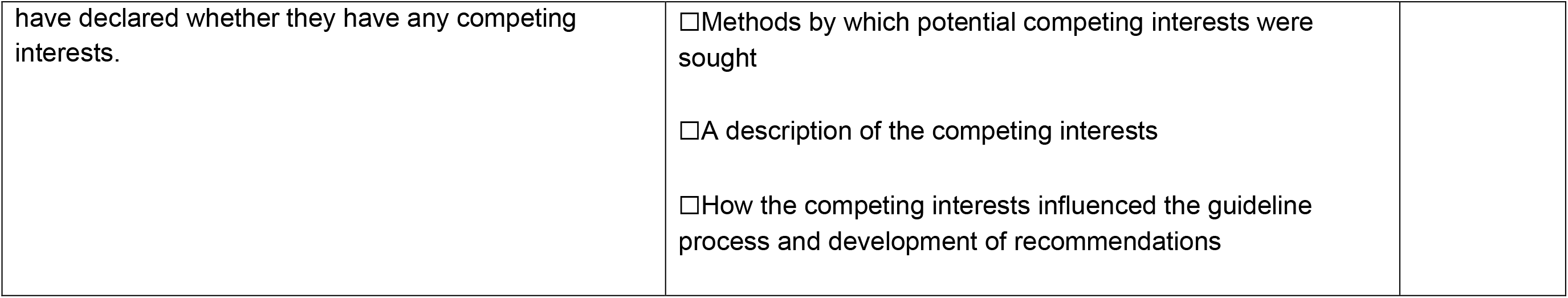

## APPENDIX 6 – CLINICAL PRACTICE GUIDELINE RESULTS

AMA, 2008; Country: Canada, Sponsor: Alberta Medical Association

Scope: Nursing home acquired pneumonia (NHAP)

ISSUES

- Treatment for NHAP should take into account the individual’s personal directives
- There is a lack of well-designed studies in this patient population
- Chest radiography is not widely available or practical in many locations
- Microbiologic diagnosis of NHAP has significant limitations and as such, treatment of NHAP is usually empiric
- Delay in administration of antibiotics for the empiric treatment of NHAP may lead to increased patient morbidity and mortality
- Inappropriate use of antibiotics may adversely affect patient outcomes and may increase antimicrobial resistance

Prevention

- Limit the spread of infections (e.g., hand washing and attention to outbreak management guidelines)
- Influenza and pneumococcal vaccines are recommended (see Appendix 2)
- Smoking cessation and avoidance of environmental tobacco smoke

Diagnosis

- Although a new infiltrate seen on chest X-ray with compatible clinical signs is the gold standard for the diagnosis of NHAP, in nursing home settings the diagnosis must often be made on clinical grounds alone.
- The physical examination must include blood pressure, heart rate, respiratory rate and auscultation of the respiratory system
- Ideally the diagnosis of pneumonia should be supported with chest X-ray, oxygen saturation, complete blood count and differential, blood cultures, and sputum cultures.
- As these tests are frequently unavailable in the nursing home setting, refer to management below.
- Note: There is still value in performing these tests even after treatment has been initiated.

Management/Assessment

- Determine the degree of medical treatment desired by the patient or legal decision maker such as a guardian or agent named in an enacted personal directive.
- Review vital signs
  - Consider transfer to hospital if impending respiratory failure or hemodynamic compromise
- Oxygenation
- Oxygen therapy is indicated for hypoxemia (e.g., O2 <90%)
- If oxymetry is not available consider oxygen at 2 litres/minute
- Note: COPD baseline oxygenation may be lower and therefore must be individually assessed
- Hydration
- Ensure adequate hydration (1 litre in a 24 hour period is required to replace insensible losses under most circumstances).
- Note: Consider hypodermoclysis
- Note: Fluid requirement for older persons without cardiac or renal failure is 30ml/kg/day in addition to estimated fluid deficit.

General Management

- Analgesics/antipyretics for pain and fever
- Cough suppressants are not routinely recommended

CONTINUING MANAGEMENT

In the nursing home setting, the care team needs to be involved in daily assessments to alert the physician to significant changes in patient status:

- Mobility
- Hydration: 1 litre/day
- Nutrition: weight loss of >5-10% is related to increased morbidity (Significant weight loss in the nursing home >5% in 30 days or >10% in 6 months)
- Review medication profile and consider holding or adjusting dosage where appropriate: psychoactive drugs, including hypnotic sedative drugs, cardiovascular drugs
- Review antibiotic treatments at 48 to 72 hours for evidence of response to therapy:
  - temperature stabilization or lower respiratory rate
- If failure of therapy occurs, consider change in antibiotics or transfer to hospital if:
  - Hemodynamic compromise
- Clinical deterioration after 72 hours of antibiotic therapy
- No improvement after completion of antibiotic therapy
- Consider:
  - Host-related factors:
    - non-infectious pulmonary pathology, immunosuppression
  - Pathogen-related factors
    - antibiotic resistance, Non-bacterial etiology, viruses, Mycobacterium spp, fungi
  - Drug-related factors
    - Adherence, malabsorption, drug-drug interactions, drug fever

Aoki, 2015; Country: Canada, Sponsor: Association of Medical Microbiology and Infectious Disease (AMMI) Canada Scope: Long-term care facility outbreak control

NI drugs, oseltamivir (Tamiflu®, Hoffmann-La Roche Ltd, Canada) and zanamivir (Relenza®, GlaxoSmithKline Inc, Canada), are the antiviral medications to be prescribed for both treatment and prevention of influenza, including H3N2 viruses. Amantadine is not to be prescribed due to resistance of the H3N2 virus to its inhibitory action.

In view of vaccine mismatch for H3N2 viruses in 2014-2015 and anticipated reduced VE for that component, it is recommended that antiviral chemoprophylaxis be considered for all staff working at the site of a declared influenza A(H3N2) LTCF outbreak regardless of whether they have received this year’s 2014-2015 influenza vaccine.

Controlling outbreaks of influenza in LTCF requires a multifaceted approach including:

1. Surveillance for influenza-like illness (ILI)
2. Laboratory testing to identify the cause of ILI
3. Promotion of, and adherence to, infection control guidelines and practices including respiratory etiquette and routine practices, and the use of personal protective equipment
4. Timely communication
5. Influenza immunization for residents and staff
6. Exclusion of ill staff, visitor exclusion and new admission deferral
7. Antiviral drug therapy for ill residents and staff
8. Antiviral drug prophylaxis of non-ill residents and staff

1. Antiviral drug therapy
  i. Residents:
    a. Oseltamivir or zanamivir treatment should be administered as soon as the clinical diagnosis of influenza has been made because residents of LTCF are, by definition, at increased risk of complications (6).
    b. Nasal/nasopharyngeal secretions should be tested to confirm the diagnosis but treatment initiation should not wait for the test result.
    c. Antiviral therapy works best when initiated within the first 48 h after symptom onset. However, these medications can still help even if begun more than 48 h after illness onset.
    d. The dosage regimens for adults are oseltamivir 75 mg PO BID or zanamivir two inhalations BID.
    e. The recommended duration of therapy is five days.
    f. There is no need to reduce doses in patients with mild to moderate reduced renal function and drug-drug interactions are also not generally a concern (6).
  ii. Staff:
    a. Staff with ILI during an outbreak should be offered antiviral treatment and sent home until their symptoms have resolved.
    b. Dose is per E1id, above.
2. Antiviral drug prophylaxis
  i. Residents:
    a. Upon diagnosis of an influenza outbreak, all residents becoming ill should be treated (vide supra) while all non-ill residents regardless of whether they have received the current seasonal vaccine should be started on chemoprophylaxis with either oseltamivir or zanamivir
    b. Generally, it is preferable to wait to initiate chemoprophylaxis until laboratory confirmation has been received.
    c. The prophylactic doses are oseltamivir 75 mg PO once daily or zanamivir two inhalations once daily for adults.
    d. Prophylaxis should be continued for 14 days minimum or until the outbreak has been declared over (see section D above).
  ii. Staff:
    a. In the context of significant antigenic drift and/or vaccine mismatch for which suboptimal VE may reasonably be anticipated, and in particular in relation to H3N2 viruses this season, it is recommended that staff who provide resident care or conduct activities where they may have the potential to acquire or transmit influenza (21) should also take prophylactic antiviral medication during the outbreak, regardless of whether they have received the current season’s influenza vaccine. This is because, despite some vaccine protection anticipated, a substantial proportion of vaccinated individuals, including healthy working-age adults, are anticipated to remain susceptible to drifted H3N2 viruses.
    b. Antiviral prophylaxis recommendations should be reinstituted whenever an outbreak is declared even for the same subtype and within the same setting and season. The maximum duration of continuous prophylaxis should be eight weeks, but outbreaks managed with antivirals should generally be terminated well within this period. In the unusual event that the outbreak is more prolonged, control measures should be reassessed in consultation with the local Medical Officer of Health and other experts.
    c. Doses and durations should be as in section E2ic above.
3. Continued surveillance for ILI during an outbreak:
  a. Residents or staff developing ILI while on prophylactic doses of an antiviral agent need to be considered for treatment doses if influenza is diagnosed or considered likely.
  b. Residents or staff receiving chemoprophylaxis who develop ILI should be assessed and tested to determine the cause of their illness, which may be due to the current virus, another viral agent or an oseltamivir-(or zanamavir)-resistant influenza virus. Expert consultation is suggested to address: the cause, organize testing for NI resistance, and to assess the possible need to possibly switch to another NI.

Buynder, 2017; Country: Australia, Sponsor: Not reported Scope: The purpose of this document is to provide national best practice guidelines for, preparing for, preventing, identifying and managing outbreaks of influenza in residential care facilities (RCFs) in Australia.

#### 5.1 Infection control measures to prevent spread of influenza

Standard precautions are a group of infection prevention practices always used in healthcare settings, and in RCFs with a suspected or confirmed influenza outbreak. Standard precautions include performing hand hygiene before and after every episode of resident contact, the use of PPE including gloves, gown, mask and eye protection, depending on the anticipated exposure, good respiratory hygiene/cough etiquette and regular cleaning of the environment and equipment.

Transmission-based precautions are work practices used in addition to standard precautions to reduce transmission opportunities due to the specific route of transmission of a pathogen. These practices are implemented depending on the type of spread. For example, respiratory infections are commonly spread by droplet and airborne routes. For influenza, droplet precautions are required.

Key elements of droplet precautions are to use PPE, maintain a 1 metre distance between the infected resident and others, encourage good cough etiquette, use resident-dedicated equipment where possible; and allocate ill residents to single rooms or cohort (put in a shared room) those with confirmed influenza. Additionally, enhanced cleaning and disinfection of the ill resident’s environment and minimising transfer of residents within and between facilities may help reduce spread. Detailed advice on these precautions is presented later in this chapter.

All staff in a residential care facility should receive general education on policies, including the principles of infection prevention and control. This would include a review of hand hygiene and infection control precautions, along with refresher training, as required. The use of antivirals should be stated in the RCF’s influenza outbreak management policy. During outbreaks, information and education should be provided for residents and their families, and should include their specific condition, and practices necessary to reduce the risk of infection.

Practical infection control information, tips, diagrams and signs are in Appendix 1.

##### 5.1.1 Hand hygiene

As influenza viruses can be spread by hands, frequent hand cleansing is important. Alcohol based hand rubs are the gold standard for hand hygiene practice in healthcare settings. However, if hands are visibly soiled or had direct contact with body fluids they should be washed with liquid soap and running water then dried thoroughly with disposable paper towel (refer to Appendices 1.1-1.4, and Hand Hygiene Australia (HHA) (http://www.hha.org.au/home/)).

Influenza viruses can persist on hard surfaces and remain viable for up to 24 hours on hard, non-porous surfaces. Infectious influenza virus can be transferred to hands from these surfaces for at least 2 - 8 hours after contamination of the surface. Virus transfer from porous materials to hands is less efficient due to rapid drying of the virus.23 Increased environmental cleaning can help interrupt virus transmission.

Hands should be cleaned with an alcohol-based hand rub or water and liquid soap solution before and after caring for a resident.

- All staff must perform hand hygiene after every contact with an ill resident.
- Even when hands are visibly clean.
- After being in contact with contaminated surfaces.
- Whether or not gloves are worn.

When visibly soiled with body fluids and/or substances, use water and liquid soap for hand washing. Alcohol-based hand rub solutions can be used when performing procedures whenever hands are not visibly soiled.

2 Refer to Hand Hygiene Australia 5 moments (http://www.hha.org.au/home/5-moments-for-hand-hygiene.aspx).

##### 5.1.2 Respiratory hygiene and cough etiquette

The importance of respiratory hygiene and cough etiquette for respiratory illnesses should be explained to residents as part of droplet precautions (refer to Appendices 1.4, 1.5). Encourage residents with respiratory symptoms to cover their nose and mouth when they cough or sneeze, use tissues, and dispose of them into a rubbish bin /receptacle. Bin contents can be disposed of as general waste.

##### 5.1.3 Personal protective equipment (PPE)

Appropriate PPE is important for all staff caring for infected residents requiring standard and droplet precautions (refer to Appendix 1.6). Staff must change their PPE and perform hand hygiene after every contact with an ill resident, when moving from one room to another, or from one resident care area to another.

##### 5.1.4 Surgical face masks

Any RCF staff member providing direct care to a resident with an ILI or influenza should wear a surgical mask. Surgical face masks must meet Australian Standards 18 and be fluid resistant, protecting the wearer from droplet contamination of the nasal or oral mucosa (refer to Appendix 1). All staff and visitors entering the room of a person with a respiratory illness should wear a single-use surgical face mask for close contact (less than (<)1 metre).

Note. Single-use surgical face masks protect the user against droplets. However, P2 respirators (known as N95 respirators or facemasks (USA equivalent rating)) protect the user against aerosols as well as droplets. Insufficient evidence exists to support the use of P2 respirators to further reduce the risk of infections transmitted by the droplet route.

Aerosol generating procedures should be avoided, or if necessary, performed in a single room that is properly cleaned before further use.25 This advice extends to nebulisers, which should not be used; use individual patient spacers instead.

Single-use surgical face masks should be worn by RCF staff when exposure to respiratory droplets is likely, that is, when within 1m of an affected resident:

- The mask should be put on when entering the room.
- Remove the mask after leaving the room, handling only by the tapes, and place in a clinical (yellow) waste bin. Perform hand hygiene after disposing of the mask.
- Never re-use masks.

When undertaking activities that require an infected resident to leave their room, the resident should wear a mask if tolerated.2 For example, during transfer within or between facilities. RCF staff members and well residents may be required to wear a mask while these activities are undertaken, based on likely exposure.

##### 5.1.5 Gloves and gown

Gloves and gowns should be used as described in standard precautions. After use, they should be removed in a manner which prevents contamination of the hands or surfaces or the workers clothing, then placed in the appropriate waste bin. Hand hygiene should be performed after removing PPE. Refer to Appendix 1

##### 5.1.6 Eye protection

Eye protection includes the use of safety glasses, goggles or face shields but does not include personal eye glasses. Goggles or other protective eyewear must be disposed of, or where approved for re-use, cleaned after use.2

Eyes should be protected where there is potential for splattering or spraying of blood, body fluids, secretions or excretions, including coughing; or when undertaking aerosol-generating procedures such as nasopharyngeal aspiration.

#### 5.2 Environmental measures

Regular, scheduled cleaning of all resident care areas is essential. Frequently touched surfaces are those closest to the resident, and should be cleaned more often (for example - bedrails, bedside tables, commodes, doorknobs, sinks, surfaces and equipment close to the resident).

During a suspected or confirmed influenza outbreak, an increase in the frequency of cleaning with a neutral detergent is recommended, especially for frequently touched surfaces. Ensure appropriate availability, quantity and placement of disposal units for tissues, used PPE, etc., as well as appropriate cleaning for reusable items.

Suggested minimum cleaning frequencies for many items are specified in the Minimum Cleaning Frequency table in the Australian Guidelines for the Prevention and Control of Infection in Healthcare (p.159, B 5.1).2 As a guide, the risk profiles of most RCFs during an influenza outbreak would be similar to general hospital wards. Refer to Appendix 2 for further information on cleaning.

Ideally, any care equipment should be dedicated for the use of an individual resident. If resident care equipment must be shared, the items must be cleaned and disinfected between each resident use.

Linen should be laundered using hot water and detergent. Linen should be dried on a hot setting in a dryer. There is no need to separate the linen of ill residents from that of other residents. Appropriate PPE should be used when handling soiled linen.

Crockery and cutlery should be washed in a dishwasher or if not available, by hand using hot water and detergent, rinsed in hot water and dried. Separation of cutlery and crockery from ill residents is not required.

#### 5.3 Exclusion of staff

RCF staff members with ILI should be excluded from work while they are infectious, that is, at least 5 days after onset of acute illness, or until they are symptom free, whichever is longer. As unvaccinated staff are at higher risk of acquiring influenza, during a confirmed influenza outbreak, unvaccinated staff are recommended to work only if asymptomatic and wearing a mask, or asymptomatic and taking appropriate antiviral prophylaxis, in keeping with the RCF’s influenza outbreak management policy. Any antiviral use by staff should be documented.

If issues arise regarding compliance with work exclusions, options should be reviewed by the Outbreak Management Team (refer to Section 6).

##### 5.3.1 Rationale for allowing staff on antivirals to return to work

Antiviral prophylaxis for staff members works to protect residents from infection, firstly, by reducing the acquisition of infection by staff (and ability to further transmit the virus) and secondly, by reducing viral shedding from asymptomatic infected staff (refer to Appendix 16: Antiviral Prophylaxis in residential care facilities decision tool).

#### 5.4 Isolation and cohorting

##### 5.4.1 Resident placement

Any residents with a respiratory illness should be cared for in a single room, where practicable. Isolating sick residents in single rooms reduces the risk of resident-to-resident transmission. The importance of respiratory hygiene and cough etiquette should be explained to all residents.

If single rooms are not available, the following principles can guide decision-making on resident placement:

- As a priority, place residents with excessive cough and sputum production in single rooms.
- Place together in the same room (cohort) residents infected with the same pathogen and who are assessed by the RCF as suitable roommates.
- Importantly, ensure that residents sharing a room are physically separated (more than (>1 metre apart) from each other. Draw the privacy curtain between beds to minimise the risk of droplet transmission.

Note: Clinical ILI may be caused by influenza A virus or influenza B virus. Only laboratory tests distinguish between these causes. Separation of ill residents is important unless it is clear their infection is caused by the same viral pathogen.

##### 5.4.2 Staff

Once resident isolation measures are in place, to further reduce the risk of transmission, it is preferable to allocate specific (vaccinated) RCF staff to the care of residents isolated in rooms. These staff members should not move between their section and other areas of the facility, or care for other residents. Staff members should self-monitor for signs and symptoms of respiratory illness and self-exclude from work if unwell.

When ILI is apparent in an RCF, influenza can be spread within the facility by unvaccinated staff, who should work only if well and wearing a mask, or taking antiviral prophylaxis. Preferentially, only staff vaccinated against influenza should care for residents with confirmed influenza or suspected ILI.

Unvaccinated staff who have been working in an outbreak-affected area should not be moved to other parts of the RCF, as they may be incubating influenza.

##### 5.4.3 Droplet precaution sign

Place droplet precaution signs (refer to Appendix 3) outside ill residents’ rooms to remind staff and visitors about the requirement for transmission-based infection control work practices.

#### 5.5 Resident movement during an outbreak

There is some evidence for the benefits of restricting internal movement of residents and visitors during an outbreak.26 RCF outbreak response plans should consider restricting resident mingling in communal living/dining areas, and prepare a guide for restrictions based on the RCF layout and usual practices. To minimize the direct interaction of residents within the facility during an outbreak, it is important for RCFs to also consider suspending group social activities for residents.

##### 5.5.1 New admissions

Admissions of new residents into the facility should be restricted. Depending upon the extent of the outbreak and the physical layout of the building, the restriction on admissions might be applied to one floor, one wing or the entire facility. The 21 RCF_Guidelines rationale for restriction on admissions is related to both the risk of infection for the newly admitted resident and potential to prolong the outbreak by adding new, potentially susceptible residents. Though normally not recommended, if important for resident well-being, admissions may be reconsidered once outbreak controls are having effect. To clarify the extent to which restrictions may be relaxed, consult with your PHU.

##### 5.5.2 Re-admission of residents who have had influenza

The re-admission of residents who had influenza and were transferred to hospital or another facility, requires the provision of appropriate accommodation, care and infection control.

##### 5.5.3 Re-admission of residents without influenza

The re-admission of residents who have not had suspected or confirmed influenza in the outbreak (i.e. who are not known cases) is generally not recommended during an outbreak. If required, admissions may be considered once the outbreak is under control. To clarify relaxation of admission restrictions, consult your public health unit.

Factors to consider if re-admission of non-cases is being sought, include:

- The availability of appropriate accommodation for the returning resident.
- Vaccination status of the person for re-admission.
- The ability to protect the readmitted resident from infection.
- Infection control measures currently in place.
- Provision of antiviral prophylactic medications.
- Availability of adequate staff at the RCF to care for the re-admitted resident.

##### 5.5.4 Non-infected residents

In some circumstances, it may be feasible to transfer residents who definitely do not have ILI to other settings for care (e.g. family care). In these circumstances, the family should be aware that the resident may have been exposed to influenza and is at risk of developing disease. NB: In residential Aged Care settings, security of tenure provisions of the Aged Care Act 1997 will need to be considered.

#### 5.6 Transfers

If transfer to hospital is required, notify the ambulance service and receiving hospital of the outbreak and the suspected or confirmed diagnosis. A template for resident transfer advice is available at Appendix 4.

#### 5.7 Visitor restriction and signage

During an outbreak, preferably, minimize the movement of visitors into and within the facility. If recommended by the outbreak management team, RCFs should:

- Suspend group social activities that involve visitors such as musicians.
- Postpone visits from non-essential external providers, if possible.
- Inform regular visitors and families of residents and of the outbreak of influenza and request they only undertake essential visits; discourage unnecessary visitors.
- Ask those who do visit an ill resident, to: o Visit only one person.
  - Enter and leave directly without spending time in communal areas.
  - Use an alcohol-based hand rub or wash their hands before and after visiting.
  - If giving direct care, use PPE as directed by RCF staff.
- Initiate passive screening for respiratory symptoms using “Attention Visitors” signage (refer to Appendix 3), and reminding visitors: o Not to visit if unwell.
  - To limit visiting to one resident.
  - To follow signs for the use of PPE, as indicated.
  - To practice hand hygiene and respiratory hygiene/cough etiquette.
- Post “Attention Visitors” signs at the entrance(s) and other strategic locations in the facility (refer to Appendix 3).
- Initiate active screening (incoming visitors report to the desk) as required.

#### 5.8 Education and training

The RCF should provide ongoing education to staff, residents and volunteers, and opportunistic education to regular visitors (e.g. residents’ families) about outbreak prevention, infection control and related policies. Topics for staff (some apply to residents) include:

- Personal hygiene, particularly hand hygiene, sneeze and cough etiquette.
- Appropriate use of PPE such as gloves, gowns, eye protection and masks, including how to don and doff PPE correctly.
- Persons experiencing symptoms of influenza (do not work or visit an RCF).
- Handling and disposal of sharps and clinical waste.
- Processing of reusable equipment.
- Environmental cleaning.
- Laundering of linen.
- Food handling and cleaning of used food utensils.

Note: This information is based on resources developed by the National Health and Medical Research Council: Australian Guidelines for the Prevention and Control of Infection in Healthcare. 2010.

Section 6: Outbreak management

Effective outbreak management has four phases:

1. Preparation: ensure a comprehensive outbreak management plan is in place and roles and responsibilities are clear, assigned and understood.
2. Response: activate the RCF’s outbreak management plan.
3. Monitor outbreak progress: assess and modify outbreak control activities.
4. Conclusion: declare the outbreak over, review events and lessons learned for future outbreaks. Update outbreak management plan.

#### 6.1 Preparing for influenza outbreaks

Seasonal influenza typically peaks over winter and early spring, although sporadic cases occur year-round. Outbreaks of influenza are not uncommon in RCFs.

To protect the health of staff and residents and to ensure the RCF is well prepared to manage an outbreak of influenza, it is important to consider the following actions:

- Plan for a possible outbreak:
  a. Develop easily accessible internal policies and procedures on infection control and an outbreak management plan. A copy of these guidelines and the Influ-Info Kit produced by the Department of Social Services should be used as references.27
  b. Include a medical practitioner (or equivalent) in the development of the outbreak management plan; this is essential for consideration of use of antiviral medications.
  c. Acquire adequate stocks of materials such as personal protective equipment (PPE) and cleaning materials (refer to Appendices 1 and 2). Consider having an outbreak kit / box specifically for use in an outbreak.
  d. Ensure RCF staff know the symptoms and signs of influenza, and are trained in infection control procedures for use of PPE (Appendix 1).
  e. Policies supporting use of antiviral drugs for prophylaxis should reflect the reduced effectiveness of antivirals with either delays in implementation or low levels of compliance, and a response option for poor compliance.
  f. If there is an intention to use influenza antivirals for prophylaxis in the management of outbreaks, RCFs may develop processes with attending GPs for antiviral prescriptions/ standing orders prior to the influenza season (see 6.2.9), as to be effective antiviral medication must be used in a widespread fashion at the same time.
  g. For residents with renal failure, GP assessment of the renal function is needed before or soon after prophylaxis commences (Appendix 16). Note. Legislation relating to the use of standing drug orders may differ between jurisdictions.
- Develop a systematic method for detecting and recording residents in the facility who develop symptoms of ILI, such as fever or cough. An ILI detection system may include increased frequency of temperature observation during winter and methods to alert the responsible clinician of symptomatic residents.
- Maintain a functional system for the timely detection of potential influenza outbreaks (3 residents/staff with ILI within 3 days) in the RCF. This may be a daily summary of all ILI records. Ensure daily hand-over time for ILI monitoring and outbreak detection for those assigned to this important task.
- Early in the year, plan to ensure at least 95% of staff and residents are vaccinated (refer to Section 3: Vaccination). Encourage staff, family members and regular visitors such as hairdressers etc., volunteers, allied health and support workers to be vaccinated. This helps keep them healthy and reduces the likelihood of influenza being brought into the facility.
- Keep vaccination records for staff and residents up-to-date, and ensure they are easily accessible when needed for an outbreak response.

Preparation and training in outbreak management and infection control for RCFs is relevant to outbreaks caused by any pathogen. A checklist of outbreak preparedness actions for RCFs can be found in Appendix 9.

#### 6.2 RCF response to an outbreak of ILI

Actions in response to an outbreak: NB - some are simultaneous:

6.2.1 Notify - ALL staff, residents, PHU, GPs, visitors (and others).

6.2.2 Implement infection control measures.

6.2.3 Arrange testing of residents with ILI.

6.2.4 Collate information onto a line list.

6.2.5 Confirm and declare an influenza outbreak.

6.2.6 Form an outbreak management team.

6.2.7 Continue infection control during the outbreak.

6.2.8 Vaccinate during an outbreak, as needed.

6.2.9 Use antiviral medication during an outbreak, as advised.

Outbreak preparedness & management checklists are in Appendices 9, 10, 11.

##### 6.2.1 Notifications

It is important to notify the local public health authority early!

PHU can assist with advice on investigation and management of potential outbreaks.

Contact details for regional public health authorities are available in Appendix 15

The RCF response to an outbreak will depend on the nature of the institution and the setting of the outbreak. Larger institutions with dedicated infection control practitioners (ICP) may require little assistance, whilst others may look to PHU for advice.

The public health unit (PHU) will require some essential information from RCFs including:

- The name, contact number and email address of a designated person or position at the RCF who will be the point of contact for the PHU. These include details for after hours and weekends, and the name and contact number of the primary ICP or delegate at the RCF.
- The number of residents in the RCF, as well as the number with ILI, their symptoms, date of onset of symptoms, whether reviewed by a GP or medical officer, likely diagnosis, and investigations requested.
- The total number of staff, and number of staff who are ill with ILI.
- The name of the testing laboratory and results of any tests completed or pending.
- The initial infection control measures that have been implemented.
- A floorplan, clearly showing the layout of different areas in the RCF.
- Current influenza vaccination rates and numbers for both residents and staff, i.e. those currently vaccinated.

To assess an outbreak, the RCF (and PHU) needs to determine the population at risk. This requires:

- The total number of residents in the RCF, and the total number of staff employed at the facility, including casual workers, and non-resident-care staff.
- If the outbreak is restricted to one area/unit, the number of residents and staff associated with that area/unit.

Refer to Appendix 14 for a generic PHU initial notification form.

Note. PHU: If requested, the PHU can provide advice regarding infection control, and assist the RCF to consider when to declare an outbreak. Refer to Section 4 for definitions of ILI, influenza and an outbreak of influenza.

The PHU can supply the RCF with additional information on testing for influenza, including advice on collecting and storing specimens, and laboratory submission of samples (see Section 6.2.3). PHU personnel may discuss results already reported, as well as request a line list of those with ILI (refer to Section 6.2.4 and Appendix 13).

It will be important for the RCF to promptly notify the following of the outbreak:

- All staff (everyone employed in the RCF) and residents.
- Visiting medical officers/GPs of residents, and arrange clinical review for ILI.
- The laboratory where specimens will be sent for testing, if needed.

To increase awareness and encourage cooperation:

- Warn visitors of the influenza risk including - scheduled allied health and service providers, and regular visitors such as family and friends.
- Place appropriate signage at the facility entrance and other strategic points.
- If transfer to hospital is required for any resident, notify the ambulance service and receiving hospital of the suspected/confirmed outbreak in the RCF, and clinical diagnosis status of the resident (well, ILI, or confirmed influenza).

##### 6.2.2 Implement preliminary infection control measures

The initial outbreak response actions in an RCF are to investigate and confirm whether the outbreak is due to influenza, and concurrently initiate appropriate management of the outbreak (regardless of the respiratory pathogen involved).

Ideally, standard precautions are in routine use. However, as soon as an influenza outbreak is suspected, transmission-based control measures (such as isolation, droplet precautions and respiratory hygiene) should be implemented as well. Refer to Section 5 for detailed advice on PPE and Infection Control, and Appendix 7 for an initial response flow chart.

##### 6.2.3 Testing of residents with ILI

Testing some residents with ILI for influenza is essential to establish a diagnosis. In an outbreak, several people meeting the ILI case definition should be tested (usually 4 to 6, but up to 10). Nose or throat swabs are collected for an influenza NAT/PCR test.

Once three or more cases of ILI occur within 3 days, and at least one has a positive laboratory test for influenza, the outbreak is confirmed. Further cases of ILI are assumed to be due to influenza and should be treated as such. No clinical or public health benefit is derived from continuing to test those with ILI in a confirmed RCF influenza outbreak, unless requested by the treating clinician or the PHU.

##### 6.2.4 Collate information onto a line list

A line list is a simple and useful record of both staff and residents who meet the ILI case definition, or are confirmed influenza cases. The line list provides important information for the RCF outbreak control team about the number of affected residents, their symptoms and vaccination status, and can be used to summarise information each day. Additionally, the PHU may use a copy of the line list to track progress and inform advice provided to the RCF on management of the outbreak.

An outbreak line list is cumulative; that is, it retains past information and is updated daily. It should include individual information such as:

- Demographic details of cases (name, age/date of birth, sex, RCF area, etc.).
- Symptoms – onset date, type, and duration.
- Results of any laboratory tests.
- Both vaccination and clinical status, including antiviral treatment.
- Outcome of infection (recovered, hospitalised, died).

Note. PHU: The local PHU may have a preferred line list format which they supply to an affected RCF. Example templates for line lists for residents and staff are available in Appendix 13.

For large RCFs, ensure each area/unit is included in the line list or keep a separate line listing for each area affected by the outbreak. A separate line listing should be completed for staff with ILI.

When completing a line list, it is important to understand that it is a cumulative document. All previous cases listed remain on the list, and each day new cases are added to the bottom of the list. An RCF will need to update the information recorded for previous cases, for example, with new test results, hospitalisation and deaths.

The use of a line-list continues throughout the outbreak response, but is particularly important in the initial phase to allow proper characterisation of the outbreak.

Note. PHU: From the line-list, as appropriate:

- Develop an epidemic curve, tracking the magnitude of the outbreak.
- Review symptoms and duration of illness; these indicate ‘what to look for’.
- Follow up and review all laboratory tests results.
- Summarise information such as characteristics of cases, attack rate, severity of illness (morbidity), and deaths (mortality).
- Track the vaccination status of those who are ill with confirmed influenza.

##### 6.2.5 Confirm and declare an influenza outbreak

Outbreaks can be caused by various infectious respiratory pathogens. If one or more respiratory samples from ILI cases in a suspected outbreak returns a positive laboratory result for influenza, then an influenza outbreak is confirmed.

Confirming influenza as the pathogen is an important step that will guide decisions regarding control measures such as vaccination, and use of antiviral medications.

##### 6.2.6 Forming an outbreak management team

When a team is formed, it is important to meet regularly, usually daily at the height of the outbreak, to monitor the outbreak, initiate changes to response measures and to discuss outbreak management roles and responsibilities.

Even a relatively small respiratory outbreak in an RCF is disruptive because of the risks of complications in vulnerable residents and transmission to staff, with resulting absenteeism. Early recognition of suspected outbreaks and swift management actions are essential for effective response to control spread.

Ideally, a full outbreak management team (OMT) should be formed by the RCF to coordinate the response. The OMT directs and oversees the management of all aspects of an outbreak, meeting at least daily during the peak of the outbreak. It considers the progress of the response, undertakes ongoing monitoring, deals with unexpected issues, and initiates changes, as required.

Several functions are critical within the OMT, and some roles may be performed by the same person. In reality, a small number of people often perform multiple roles and undertake many tasks. For further details, refer to Appendices 9 - 12.

The outbreak management team should initially meet daily to:

- Direct and oversee the management of the outbreak.
- Monitor the outbreak progress and initiate changes in response, as required.
- Liaise with GPs and PHU, as arranged.
- The Outbreak Management Team may include the following people:

Chairperson (Facility Director, Manager, or Nursing Manager)

The chairperson is responsible for co-ordinating outbreak control meetings, setting meeting times, agenda and delegating tasks. Secretary

The secretary organises OMT meetings, notifies committee members of any changes, and records and distributes minutes of meetings.

Outbreak Coordinator (Nurse / Infection Control Practitioner or delegate)

The coordinator ensures that all infection control decisions of the OMT are carried out, and coordinates activities required to contain and investigate the outbreak. This role is often given to an Infection Control Practitioner(ICP) or delegate.

Media spokesperson/s (Facility Director, or Manager, or delegate)

Significant media interest in outbreaks in RCFs is common, especially if there are adverse outcomes. A media spokesperson from the OMT should be designated as the only individual with responsibility to provide information to the media. This media spokesperson is usually a representative from the RCF, but occasionally, can be from the PHU, if involved. In rare situations, there may be joint spokespeople, one from each organisation.

Visiting General Practitioners

Some GPs may be available to participate in the OMT and their role should be identified during the planning process. It is valuable to identify a clinical lead amongst those GPs who attend a facility. In the management of an outbreak, the role of this person is important in facilitating assessment and management of ill residents, and in working with the RCF and PHU to implement control strategies.

Public health authority staff

An understanding of what assistance can be provided by the public health authority and role/responsibility clarification should be confirmed at the initial OMT meeting, although it is usually not necessary for PHU staff to be part of the OMT.

Refer to Appendix 9 for an outbreak Plan checklist, Appendix 10 for details of OMT tasks and Appendix 11 for an OMT task checklist.

##### 6.2.7 Infection control during an outbreak

During an influenza outbreak it is essential that infection control measures are put in place expeditiously to minimise the spread of disease in the facility. These procedures are detailed in Section 5. A checklist of infection control procedures to be considered in an outbreak can be found at Appendix 12.

##### 6.2.8 Vaccination during an outbreak

During influenza outbreaks, the influenza vaccination records of all residents and staff should be reviewed as a priority.

Influenza vaccination clinics may be arranged for unvaccinated residents and staff, and recommended for unvaccinated visitors. Alternatively, the RCF may recommend directing the person to consult their primary health provider for vaccination; remembering it takes around two weeks to develop a protective immune response following vaccination.

##### 6.2.9 Antiviral medication during an outbreak

Residents’ GPs are responsible for prescribing antiviral medications.

Antiviral use for case management - treatment

- Early initiation of antiviral treatment (within 48 hours of symptom onset) in adults with confirmed influenza reduces the risk of secondary complications requiring antibiotic therapy, and hospitalisation. Provision of medication after this time to cases will decrease shedding time and reduce transmission but not affect the course of illness.
- To facilitate early case treatment in the context of an identified influenza outbreak, treatment on syndromic grounds may be warranted,29 particularly for individuals with underlying chronic conditions that place them at increased risk of a severe clinical course with influenza, and for whom observational studies suggest the greatest clinical benefit.

Antiviral use for prophylaxis

The widespread use of antivirals in institutions that house residents at high risk of severe disease and death from influenza is supported by observational cohort studies and one randomised controlled trial. During an outbreak, other facility residents will have been, or may become, exposed to infectious residents. The provision of antivirals works as early treatment for those incubating disease and reduces shedding in those infected. Data are available to support the premise that this treatment/shedding reduction process prevents continued spread in the facility. (Appendix 16)

Antiviral prophylaxis should only be used in addition to other outbreak control measures. The decision to administer antivirals as prophylaxis should be made by the OMT in collaboration with local public health authorities and residents’ GPs, depending on local arrangements.

If recommended, to optimise the chances of reducing transmission and bring the outbreak under control, antiviral prophylaxis should be given to ALL asymptomatic residents (regardless of vaccination status) and ALL unvaccinated staff.

Prompt administration of medication and estimated levels of compliance should be considered in the decision to use antivirals for prophylaxis. Hence, outbreak plans should incorporate the expediency of acquiring and administrating medication (for residents), an acceptable compliance level and a response option for poor compliance.

If antivirals are used for influenza prophylaxis in an RCF:

- Ideally, antivirals should be commenced by all targeted residents and staff within 24 hours, AND
- Medication safety issues, including renal function/renal insufficiency, must be appropriately considered during the prescribing phase.
- RCF staff need to be aware of the most common side effects, e.g. for oseltamivir, nausea and vomiting.

Further information is available in the appendices including:

Appendix 16. Table 16.1 Antiviral prophylaxis in RCF decision tool;

Appendix 17. Letter regarding antivirals for treating doctors;

Appendix 18. Antiviral dosage recommendations;

Appendix 19. Patient information on Tamiflu (oseltamivir).

#### 6.3 Monitoring the outbreak

An RCF member of the OMT, or delegate, should update the line listing with new information daily, by midday (or another agreed time), or more frequently if major changes occur, and communicate this to the PHU each day (as arranged, by email (preferred), fax or telephone). Updates to information should occur through daily meetings of the OMT.

Note. PHU: Updated information should be reviewed by the PHU for evidence of ongoing transmission and effectiveness of control measures and prophylaxis. The PHU should discuss this with the RCF OMT and advise of any indicated changes for current outbreak control measures.

Ongoing resident surveillance should include the following:

- Monitoring residents for ILI symptoms.
- Addition of all new cases to the resident line list.
- Updating the status of ill residents: hospitalised, recovered, deceased.
- Recording the use of antiviral prophylactic medication and any adverse reactions to or cessation of any prescribed antiviral medication.

Ongoing staff surveillance should include all the following:

- Addition of all new staff cases to the staff line list.
- Identification of staff who have recovered, and confirmation with the PHU of their return to work date.

The OMT should review all control measures and consider seeking further advice from PHU if:

- The outbreak comprises more than 10 cases.
- The rate of new cases is not decreasing.
- Three (3) or more residents are hospitalised related to influenza, OR
- An influenza-related death has occurred: telephone to notify the PHU of this.

Specialised advice is available from the following sources:

- A local state, territory or regional PHU.
- Infection control practitioners may be available for advice in local hospitals, state and territory health departments, or as private consultants.
- Geriatricians or Infectious Disease physicians may be approached for specialist management of complex infections.

#### 6.4 Concluding the outbreak

##### 6.4.1 Declaring the outbreak over

The OMT, with approval from the local public health authority, has responsibility for declaring the outbreak is over.

The time from the onset of symptoms of the last case until the outbreak is declared over can vary, and depends on whether the last case was a resident or staff member. Generally, an influenza outbreak can be declared over if no new cases occur within 8 days following the onset of symptoms in the last resident case (8 days is the sum of the usual infectious period [5 days] plus maximum incubation period [3days]).

A decision to declare the outbreak over should be made by the OMT, in consultation with the PHU.

The OMT may take decisions about ongoing RCF surveillance after declaring the outbreak over, considering the following needs:

- To maintain general infection control measures.
- To monitor the status of ill residents, communicating with the public health authority if their status changes.
- To notify any late, influenza-related deaths to the PHU.
- To alert the PHU to any new cases, signaling either re-introduction of infection or previously undetected ongoing transmission.
- To advise relevant state/territory/national agencies of the outbreak, including the Commonwealth Department of Health and Ageing state/territory office for outbreaks in Aged Care facilities, if applicable.

##### 6.4.2 Communication

Once the outbreak has been declared over, the OMT should notify all individuals and agencies involved in the investigation of the declaration.

The Outbreak Management Team is responsible for notifying the following people:

- All staff and residents.
- Visiting GPs of residents.
- Visiting service providers, families of residents and other regular visitors.
- The ambulance service and any relevant receiving hospitals or other RCFs.

##### 6.4.3 Organising a debrief after the outbreak

Following a declaration that an outbreak is over, it is important for all parties to reflect on what worked well during the outbreak and which policies, practices or procedures need to be modified to improve responses for future outbreaks.

Although a debrief may seem unnecessary for outbreaks of short duration involving a small number of cases, the OMT in collaboration with the local PHU should consider a debrief for any outbreak with many cases, a prolonged outbreak, or one with unusual features in relation to outbreak management. Outbreak investigation is a core function of PHUs, and after respiratory outbreaks in RCFs, a debrief whether formal or informal, is recommended in partnership with the RCF outbreak management team.

Audits are commonly used in clinical medical and nursing practice as part of continuous quality improvement, and may be an appropriate method by which to conduct a debrief. Australian public health practitioners and researchers have proposed a guide for an outbreak audit process, with a framework for deciding which outbreak investigations to audit, an approach for conducting a successful audit, and a template for trigger questions. This tool enables agencies such as RCFs to assess their outbreak response against best practice.

If a formal debrief or an audit is undertaken, the OMT should arrange a meeting between key RCF staff, PHU staff, and other relevant agencies, to review the course and management of the outbreak. The purpose of this meeting is to identify what worked well, and what could be improved for future outbreaks. For preference, this meeting should occur within two weeks of the outbreak being declared over.

A debrief provides the opportunity to identify strengths and weaknesses in outbreak response and investigation processes, and provide information to help improve the management of similar outbreaks in the future. It should involve all members of the OMT and any others who participated in the response to the outbreak.

Preparation, training and experience for RCF personnel in outbreak management, response, and infection control for influenza is directly applicable to outbreaks in RCFs caused by any respiratory pathogen; much applies to gastrointestinal illness.

California Dept of Public Health, 2019; Country: USA, Sponsor: California Department of Public Health

Scope: provides and clarifies recommendations to prevent and manage influenza outbreaks in skilled nursing facilities (SNF). The recommendations may also apply to other long-term care facilities (LTCF), for example, congregate living health facilities and intermediate care facilities.

Implementing Enhanced Standard Precautions by using gown, gloves, and performing frequent hand hygiene while caring for residents at increased risk of transmitting infectious agents is necessary year-long and especially during influenza season.

6. SNF must develop plans to be able to accept new admissions during influenza season while maintaining capacity to care safely for other residents. This requires planning for implementing Transmission-Based Precautions and other infection control measures.

7. Respiratory hygiene/cough etiquette is necessary for all individuals year-long. Influenza virus is transmissible to others for 24 hours before an individual has typical signs and symptoms of influenza. Additionally, older individuals and those who are immunocompromised may not present with classical signs of influenza (2, 4, 5). Containing all respiratory secretions (source containment) at all times is therefore necessary.

8. When an influenza outbreak in a SNF is suspected (2, 13), prompt and simultaneous implementation of interventions (https://academic.oup.com/cid/article/68/6/e1/5251935) can minimize the size and scope of the outbreak and adverse impact on resident health. Outbreak management requires a collaborative effort among all HCP with specific task assignments and tracking their completion.

a. Prompt administration of antiviral agents for treatment and prophylaxis will shorten an outbreak (6, 7).
b. Communicating with residents, HCP, and families, during an outbreak provides needed reassurance.
c. Communicating with the local health department will facilitate additional guidance during an outbreak.

Table 1 includes action to take for planning management of an influenza illness and outbreaks including towards residents, health care providers and visitors/families

Table 2 outlines actions for identifying and managing influenza outbreaks in SNF

CDC, 2020; Country: USA, Sponsor: Centres for Disease Control

Scope: Preventing COVID-19 in nursing homes

Visitor Restrictions

III visitors and healthcare personnel (HCP) are the most likely sources of introduction of COVID-19 into a facility. CDC recommends aggressive visitor restrictions and enforcing sick leave policies for ill HCP, even before COVID-19 is identified in a community or facility.

Educate Residents, Healthcare Personnel, and Visitors

Share the latest information about COVID-2019.

Review CDC’s Interim Infection Prevention and Control Recommendations for Patients with Confirmed Coronavirus Disease 2019 (COVID-19) or Persons Under Investigation for COVID-19 in Healthcare Settings.

Educate and train HCP.

Reinforce sick leave policies. Remind HCP not to report to work when ill.

Reinforce adherence to infection prevention and control measures, including hand hygiene and selection and use of personal protective equipment (PPE).

Have HCP demonstrate competency with putting on and removing PPE.

Educate both facility-based and consultant personnel (e.g., wound care, podiatry, barber) and volunteers. Including consultants is important because they often provide care in multiple facilities and can be exposed to or serve as a source of pathogen transmission.

Educate residents and families including:

information about COVID-19

actions the facility is taking to protect them and their loved ones, including visitor restrictions

actions residents and families can take to protect themselves in the facility

Provide Supplies for Recommended Infection Prevention and Control Practices

Hand hygiene supplies:

Put alcohol-based hand sanitizer with 60–95% alcohol in every resident room (ideally both inside and outside of the room) and other resident care and common areas (e.g., outside dining hall, in therapy gym).

Make sure that sinks are well-stocked with soap and paper towels for handwashing.

Respiratory hygiene and cough etiquette:

Make tissues and facemasks available for coughing people.

Consider designating staff to steward those supplies and encourage appropriate use by residents, visitors, and staff.

Make necessary Personal Protective Equipment (PPE) available in areas where resident care is provided. Put a trash can near the exit inside the resident room to make it easy for staff to discard PPE prior to exiting the room, or before providing care for another resident in the same room.

Facilities should have supplies of:

facemasks

respirators (if available and the facility has a respiratory protection program with trained, medically cleared, and fit-tested HCP)

gowns

gloves

eye protection (i.e., face shield or goggles).

Environmental cleaning and disinfection:

Make sure that EPA-registered, hospital-grade disinfectants are available to allow for frequent cleaning of high-touch surfaces and shared resident care equipment.

Refer to List Nexternal icon on the EPA website for EPA-registered disinfectants that have qualified under EPA’s emerging viral pathogens program for use against SARS-CoV-2.

Evaluate and Manage HCP with Symptoms of Respiratory Illness

Implement sick leave policies that are non-punitive, flexible, and consistent with public health policies that allow ill HCP to stay home.

As part of routine practice, ask HCP (including consultant personnel) to regularly monitor themselves for fever and symptoms of respiratory infection.

Remind HCP to stay home when they are ill.

If HCP develop fever or symptoms of respiratory infection while at work, they should immediately put on a facemask, inform their supervisor, and leave the workplace.

Consult occupational health on decisions about further evaluation and return to work.

When transmission in the community is identified, nursing homes and assisted living facilities may face staffing shortages. Facilities should develop (or review existing) plans to mitigate staffing shortages.

Consider New Policies and Procedures for Visitors

Because of the ease of spread in a long-term care setting and the severity of illness that occurs in residents with COVID-19, facilities should discourage visitation and begin screening visitors even before COVID-19 is identified in their community.

Facilities should:

Send letters or emails to families advising them to consider postponing or using alternative methods for visitation (e.g., video conferencing) during the next several months.

Post signs at the entrances to the facility instructing visitors to not enter if they have fever or symptoms of a respiratory infection.

Consider having visitors sign visitor logs in case contact tracing becomes necessary.

Ask all visitors about fever or symptoms of respiratory infection. Restrict anyone with:

Fever or symptoms of respiratory infection (e.g., cough, sore throat, or shortness of breath).

International travel within the last 14 days to affected countries. Information on high-risk countries is available on CDC’s COVID-19 travel website.

Contact with an individual with COVID-19.

When allowed, visitors should be encouraged to frequently perform hand hygiene and limit their movement and interactions with others in the facility (e.g., confine themselves to the resident’s room).

When visitor restrictions are implemented, the facility should facilitate remote communication between the resident and visitors (e.g., video-call applications on cell phones or tablets), and have policies addressing when and how visitors might still be allowed to enter the facility (e.g., end of life situations).

Evaluate and Manage Residents with Symptoms of Respiratory Infection

Ask residents to report if they feel feverish or have symptoms of respiratory infection.

Promptly assess residents for fever and symptoms and signs of respiratory infection upon admission and throughout their stay in the facility.

Implement appropriate infection prevention practices for symptomatic residents:

If a resident has severe respiratory infection, or a cluster (e.g., ≥ 3 residents or HCP with new-onset respiratory symptoms over 72 hours) of residents has symptoms of respiratory infection, or there is an increase in cases reported in the community, begin active monitoring of all residents and HCP in the facility for signs and symptoms.

Notify the health department about residents with severe respiratory infection and clusters of respiratory infection. See State-Based Prevention Activities for contact information for the healthcare-associated infections program in each state health department.

CDC has resources for performing respiratory infection surveillance in long-term care facilities during an outbreak.

In general, when caring for residents with undiagnosed respiratory infection use Standard, Contact, and Droplet Precautions with eye protection unless the suspected diagnosis requires Airborne Precautions (e.g., tuberculosis). This includes restricting residents with respiratory infection to their rooms. If they leave the room, residents should wear a facemask (if tolerated) or use tissues to cover their mouth and nose.

Continue to assess the need for Transmission-Based Precautions as more information about the resident’s suspected diagnosis becomes available

If COVID-19 is suspected, based on evaluation of the resident or prevalence of COVID-19 in the community, Residents with known or suspected COVID-19 do not need to be placed into an airborne infection isolation room (AIIR) but should ideally be placed in a private room with their own bathroom.

Room sharing might be necessary if there are multiple residents with known or suspected COVID-19 in the facility. As roommates of symptomatic residents might already be exposed, it is generally not recommended to separate them in this scenario. Public health authorities can assist with decisions about resident placement.

Facilities should notify the health department immediately and follow the Interim Infection Prevention and Control Recommendations for Patients with COVID-19 or Persons Under Investigation for COVID-19 in Healthcare Settings, which includes detailed information regarding recommended PPE.

If a resident requires a higher level of care or the facility cannot fully implement all recommended precautions, the resident should be transferred to another facility that is capable of implementation. Transport personnel and the receiving facility should be notified about the suspected diagnosis prior to transfer.

While awaiting transfer, symptomatic residents should wear a facemask (if tolerated) and be separated from others (e.g., kept in their room with the door closed). Appropriate PPE should be used by healthcare personnel when coming in contact with the resident.

Additional Measures

Minimize group activities inside the facility or field trips outside of the facility.

Develop criteria for halting group activities and communal dining, closing units or the entire facility to new admissions, and restricting visitation.

Create a plan for cohorting residents with symptoms of respiratory infection, including dedicating HCP to work only on affected units. In addition to the actions described above, these are things facilities should do when there are cases in their community but none in their facility.

Policies and Procedures for Visitors

Visitation should be limited further to only those who are essential for the resident’s emotional well-being and care. The facility should send communications to families advising the COVID-19 has been identified in the community and re-emphasizing the importance of postponing visitation. Ideally, visits should be scheduled in advance during a limited number of hours. Any visitors (that are permitted after screening) should wear a facemask while in the building and restrict their visit to the resident’s room.

Healthcare Personnel Monitoring and Restrictions

Restrict non-essential personnel including volunteers and non-essential consultant personnel (e.g., barbers) from entering the building.

Screen all HCP at the beginning of their shift for fever and respiratory symptoms.

Actively take their temperature and document absence of shortness of breath, new or change in cough, and sore throat. If they are ill, have them put on a facemask and self-isolate at home.

HCP who work in multiple locations may pose higher risk and should be asked about exposure to facilities with recognized COVID-19 cases.

Consider implementing universal use of facemasks for HCP while in the facility.

Resident Monitoring and Restrictions

Actively monitor all residents (at least daily) for fever and respiratory symptoms (shortness of breath, new or change in cough, and sore throat).

If positive for fever or symptoms, implement recommended IPC practices

Cancel group field trips and activities and consider cancelling communal dining.

In addition to the actions described above, these are things facilities should do when there are cases in their facility or sustained transmission in the community.

Policies and Procedures for Visitors:

Restrict all visitors to the facility. Exceptions might be considered in limited circumstances (e.g., end of life situations). In those circumstances the visitor should wear a facemask and restrict their visit to the resident’s room.

Healthcare Personnel Monitoring and Restrictions:

Implement universal use of facemask for HCP while in the facility.

Consider having HCP wear all recommended PPE (gown, gloves, eye protection, N95 respirator (or facemask if not available)) for the care of all residents, regardless of presence of symptoms. Implement protocols for extended use of eye protection and facemasks.

Resident Monitoring and Restrictions:

Encourage residents to remain in their room. If there are cases in the facility, restrict residents (to the extent possible) to their rooms except for medically necessary purposes.

If they leave their room, residents should wear a facemask, perform hand hygiene, limit their movement in the facility, and perform social distancing (stay at least 6 feet away from others).

In addition to cancelling group field trips and activities, cancel communal dining.

Implement protocols for cohorting ill residents with dedicated HCP

ECRI, 2020; Country: USA, Sponsor: ECRI

Scope: 2003 outbreak of severe acute respiratory syndrome

ECRI has established the following levels of concern related to the risks of infection associated with the circumstances described:

No infection concern. There is no concern about infection from external surfaces that have been cleaned and disinfected.

Minimal infection concern. If the interior of a device is exposed to SARS-CoV from room air—most likely air drawn into the device by a cooling fan—there may be some concern about contamination. Until more is known about the transmission of SARS, we suggest that hospitals err on the side of caution and use simple protective measures for even such minimal-risk situations. These measures are readily implemented (see Specific Protective Steps, below). Note, however, that the warm air that is generally circulating inside a device with a cooling fan promotes drying and dilution of contaminants, which may reduce the viability of some viruses.

Higher infection concern.

Surfaces that have been in contact with the patient’s oral secretions or other excretions and cannot be readily disinfected pose a bigger concern. This is because, although the infection risk is still likely to be extremely low on these surfaces, there is a greater probability that the virus has remained viable. Such surfaces include the following:

Breathing circuits (including ventilator accessories and any portions of the breathing circuit inside ventilators), suction devices and systems, or any other devices that are exposed to the patient’s oral secretions (including contaminated condensate), urine, feces, and other excretions

HEPA filtration systems, which are installed systems or portable systems (i.e., mobile high-efficiency-filter air cleaners [MHEFACs]) used to control room air contamination levels and ensure negative pressure in isolation rooms

Any handheld items or other items that can be found in beds, including nurse call buttons, remote controls for televisions, pillow speakers, blood pressure cuffs, and telemetry transmitters

Highest infection concern.

The highest level of concern is posed by entering the room of a SARS patient without appropriate personal protective equipment (PPE). This is because of the close proximity to the patient, the potential risk of exposure to droplet secretions from coughs and sneezes, exposure to contaminated surfaces, and possible exposure associated with aerosol-generating procedures.

Keep in mind that even the highest level of risk is not an insurmountable concern as long as servicing personnel take the protective steps listed in the next section.

Specific Protective Steps

The following practices should be implemented to protect personnel while they are maintaining and repairing equipment that has been used on—or that has been in the same room as—patients who have or who are suspected of having SARS. For the most part, these represent good infection control practices that should be followed when servicing any device, independent of SARS concerns.

1. Do not enter the room of a SARS patient

Access to such rooms should be restricted to essential personnel only. This is to ensure the safety of personnel and to minimize disease transmission. However, if you must enter the room, follow relevant hospital procedures to minimize the exposure risks.

2. Minimize exposure of medical equipment to SARS

Before a patient with SARS is brought into a room, remove any unessential equipment. Use breathing-circuit filters to protect exhalation valves and other ventilation components from contamination (for more on this topic, see the Guidance Article Mechanical Ventilation of SARS Patients: Lessons from the 2003 SARS Outbreak). Use disposable devices or accessories for SARS patients whenever possible.

3. Observe proper hand hygiene

Frequent and thorough handwashing with soap and water is essential. Alcohol-based handrubs can be used when hands are not visibly soiled and handwashing facilities are not immediately available. Personnel should not rub their eyes or touch their mouth, nose, or other mucous membranes while working on exposed equipment. While wearing gloves, personnel should also avoid touching other surfaces in the room that are not involved in the equipment repair (e.g., doorknobs, telephones, test equipment, computer terminals, keyboards, manuals). In addition, personnel should not eat, drink, chew gum, smoke, or apply cosmetics until they have removed all protective wear and washed their hands.

4. Use proper decontamination and transport procedures

Equipment should not be transported until it has been cleaned and disinfected and disposables have been removed by housekeeping, central processing, or other appropriate personnel. (Note that commonly used disinfectants are effective against the SARS virus.) If equipment from the room of a SARS patient must be removed before the exterior can be cleaned and disinfected, follow any hospital policies on transporting contaminated devices.

5. Choose an appropriate work area

Equipment that poses particular infection concerns should be worked on in designated areas where servicing can be performed without the risk of infecting patients or other employees. These areas should not be near any patient care areas, food preparation or storage areas, medication areas, or other clean areas.

6. Wear protective equipment when appropriate

For personnel working on minimal-risk surfaces, we recommend using the following PPE:

Gloves

Clean, nonsterile gown, apron, or laboratory coat

Eye protection (e.g., goggles)

A face shield is an alternative form of eye protection. Although a face shield should provide adequate protection against an occasional minor splatter that may occur during servicing, the U.S. Occupational Safety and Health Administration (OSHA) requires the use of goggles (or special protective eyeglasses) when eye protection is used. CDC, on the other hand, recommends goggles or a face shield for protection against a splash or spray of body fluids.

Respiratory protection

Update: As of February 3, 2020, CDC recommends, in the case of the COVID-19 outbreak, that staff “use respiratory protection (i.e., a respirator) that is at least as protective as a fit-tested NIOSH-certified disposable N95 filtering facepiece respirator before entry into the patient room or care area.” That recommendation matches the guidance ECRI Institute initially issued (in June 2003) in the case of the 2003 SARS outbreak.

Note, however, that respiratory protection recommendations can evolve over time. For example, as new information about airborne transmission of SARS became available and as concern lessened, we updated our recommendations for that circumstance: In February 2004, we specified that it would be prudent for personnel working with the equipment to wear a surgical mask, but that there was no significant benefit to using an N95 respirator for that application, particularly in light of the considerable time and effort that respirator use entails. (For additional information about respirator types and implementation challenges, see Selecting Respiratory Protection for Equipment Servicers and Other Hospital Personnel: Lessons from the 2003 SARS Outbreak.) It is unknown at this time if similar changes will develop in the case of the COVID-19 outbreak.

7. Before starting work . . .

If there is any question about whether the exterior surfaces of the equipment were adequately disinfected, including the bottom and back, disinfect those surfaces immediately. Also, if disposable components have not already been discarded, do that right away as well. If the equipment is not needed immediately, ECRI suggests allowing time—several hours to overnight—for viruses to die before servicing is carried out. (Note, however, that this waiting period should not be seen as a substitute for other infection control procedures.)

8. If the interior of the equipment is dusty . . .

Use a vacuum cleaner with a HEPA filter to remove dust as soon as adequate access is gained during disassembly and before working on the interior. Never blow on the equipment or use compressed air to remove dust or other particulates.

9. Clean up when done.

Clean and disinfect the work area after servicing is complete.

10. If an exposure occurs . . .

If you believe you have been exposed to SARS-CoV while unprotected, consult with the hospital’s infection control practitioner, epidemiologist, or employee health staff for the procedures to follow.

Manitoba Health Seniors and Active Living, 2019; Country: Canada, Sponsor: Not reported Scope: these guidelines support infection prevention and control (IP&C) professionals, health care organizations and health care providers in developing, implementing and evaluating IP&C policies, procedures and programs to improve the quality and safety of health care and patient outcomes

13. Modifications for Contact Precautions for Long Term Care, Ambulatory Care, Home Care, Prehospital. Routine Practices (as per Part B, Section III) and Contact Precautions should be followed for all health care settings (as per Part B, Section IV) sub-section i) and modified as noted below:

For Long Term Care

a. Patient Placement, Accommodation and Activities
  - Perform a point of care risk assessment to determine patient placement, removal from a shared room or participation in group activities on a case-by case basis, balancing infection risks to other patients in the room, the presence of risk factors that increase the likelihood of transmission, and the potential adverse psychological impact on the infected patient.
  - Participation in group activities should be restricted only when wound drainage or diarrhea cannot be contained.
  - Ensure patient performs hand hygiene or is assisted as necessary before participation with group activities.
b. Use of Personal Protective Equipment
  - Wear gloves if direct personal care contact with the patient is required or if direct contact with frequently touched environmental surfaces is anticipated.
  - Wear gowns for direct hands-on care.
c. Cleaning of Patient Environment
  - In outbreaks, consider more frequent cleaning and cleaning with disinfectants. This includes bathing and toileting facilities, recreational equipment and horizontal surfaces in the patient room, and in particular, areas and items that are frequently touched (e.g., hand and bedrails, and light cords).

Modifications of Droplet Precautions in Long Term Care

a. In long term care and other patient settings, perform a point of care risk assessment to determine patient placement, considering infection risks to other patient(s) in the room and available alternatives.
b. If a two-metre spatial separation is not possible, manage the patient in their bed space, with privacy curtains drawn.
c. Participation in group activities may need to be restricted while the patient is symptomatic.
d. During an outbreak in a facility, restrict social activities to units and areas.
e. Restrictions in the number of visitors may be advisable during community or facility outbreaks of respiratory infections.

Modifications of Airborne Precautions for Long Term Care

a. Tuberculosis (infectious, respiratory (pleural or laryngeal))
  i. Determine the tuberculosis infection status of patients in LTC facilities at the time of admission.
  ii. If an airborne infection isolation room is not available in the long term care setting, arrange for transfer to a facility with airborne infection isolation rooms. When transfer is delayed, reduce the risk of transmission of tuberculosis by the following:
    - Place the patient in a single room with the door closed, preferably without recirculation of air from the room and as far away from rooms of other patients as possible.
    - Limit the number of people entering the room (e.g., no non-essential visitors).
b. Varicella, disseminated herpes zoster or localized herpes zoster that cannot be kept covered, or measles:
  i. Determine the immune status (measles or varicella) of patients in LTC facilities at the time of admission and offer immunization, if appropriate.
  ii. If an airborne infection isolation room is not available in the long term care setting, arrange for transfer to a facility with airborne infection isolation rooms. If transfer is delayed, reduce the likelihood of transmission by the following:
    - Place the patient in a single room with the door closed, preferably without recirculation of air from the room and as far away from rooms of other patients as possible.
    - Limit the number of people entering the room (e.g., no non-essential visitors).
    - If all personnel and all other patients in the facility are immune and if non-immune visitors can be excluded, transfer to a facility with an AIIR may not be essential.
  iii. Do not place infectious patients on units where there are susceptible immunocompromised patients.

Ofelia, 2003; Country: USA, Sponsor: Centres for Disease Control

Scope: Reduce the incidence of nosocomial pneumonia

PREVENTION AND CONTROL OF HEALTH-CARE ASSOCIATED RSV, PARAINFLUENZA VIRUS, AND ADENOVIRUS INFECTIONS

I. Staff Education And Monitoring And Infection Surveillance

A. Staff Education and Monitoring

1. Staff education

a. Educate personnel in accordance with their level of responsibility in the health-care setting about the epidemiology, modes of transmission, and means of preventing the transmission of RSV within health-care facilities (817).

CATEGORY IB

b. Educate personnel in accordance with their level of responsibility in the health-care setting, about the epidemiology, modes of transmission, and means of preventing the spread of parainfluenza virus and adenovirus within health-care facilities.

CATEGORY II

2. In acute-care facilities, establish mechanisms by which the infection-control staff can monitor personnel compliance with the facility’s infection-control policies about these viruses (817).

CATEGORY II

B. Surveillance

1. Establish mechanisms by which the appropriate health-care personnel are promptly alerted to any increase in the activity of RSV, parainfluenza virus, adenovirus, or other respiratory viruses in the local community. Establish mechanisms by which the appropriate health-care personnel can promptly inform the local and state health departments of any increase in the activity of the above-named viruses or of influenza-like illness in their facility.

CATEGORY IB

2. In acute-care facilities during periods of increased prevalence of symptoms of viral respiratory illness in the community or health-care facility, and/or during the RSV and influenza season (i.e., December-March), attempt prompt diagnosis of respiratory infections caused by RSV, influenza, parainfluenza, or other respiratory viruses. Use rapid diagnostic techniques as clinically indicated in patients who are admitted to the health-care facility with respiratory illness and are at high risk for serious complications from viral respiratory infections (e.g., pediatric patients, especially infants, and those with compromised cardiac, pulmonary, or immune function) (749;764;815;817;897).

CATEGORY IA

3. No recommendation can be made for routinely conducting surveillance cultures for RSV or other respiratory viruses in respiratory secretions of patients (including immunocompromised patients, such as recipients of HSCT) (507).

UNRESOLVED ISSUE

4. In LTCFs, establish mechanism(s) for continuing surveillance to allow rapid identification of a potential outbreak in the facility. CATEGORY II

II. Prevention Of Transmission Of RSV, Parainfluenza Virus, Or Adenovirus

A. Prevention of Person-to-Person Transmission

1. Standard and contact precautions for RSV and parainfluenza virus; and standard, contact, and droplet precautions for adenovirus

a. Hand hygiene

Decontaminate hands after contact with a patient or after touching respiratory secretions or fomites potentially contaminated with respiratory secretions, whether or not gloves are worn. Use soap and water when hands are visibly dirty or contaminated with proteinaceous material or are visibly soiled with blood or other body fluids, and use an alcohol-based hand rub if hands are not visibly soiled (271;279;284;782;802-804;813;817).

CATEGORY IA

b. Gloving

(1) Wear gloves when entering the room of patients with confirmed or suspected RSV, parainfluenza, or adenovirus infection, and/or before handling the patients, their respiratory secretions, or fomites potentially contaminated with the patients’ secretions (279;280;782;802;803;810;815;817;819).

CATEGORY IA

(2) Change gloves between patients or after handling respiratory secretions or fomites contaminated with secretions from one patient before contact with another patient (279;280;282;817). Decontanimate hands after removing gloves (see II-A-1-a).

CATEGORY IA

(3) After glove removal and hand decontamination, do not touch potentially contaminated environmental surfaces or items in the patient’s room (279).

CATEGORY IB

c. Gowning

Wear a gown when entering the room of a patient suspected or proven to have RSV, parainfluenza virus, or adenovirus infection, and when soiling with respiratory secretions from a patient is anticipated (e.g., when handling infants with suspected or proven RSV, parainfluenza, or adenovirus infection). Change the gown after touch contact and before giving care to another patient or when leaving the patient’s room. After gown removal, ensure that clothing does not come into contact with potentially contaminated environmental surfaces (279;280).

CATEGORY IB

d. Masking and wearing eye protection

(1) Wear a surgical mask and eye protection or a face shield when performing procedures or patient-care activities that might generate sprays of respiratory secretions from any patient whether or not the patient has confirmed or suspected viral respiratory tract infection (279).

CATEGORY IB

(2) Wear a surgical mask and eye protection or a face shield when within 3 feet of a patient with suspected or confirmed adenovirus infection (279).

CATEGORY IB

e. Patient placement in acute-care facilities

(1) Place a patient with diagnosed RSV, parainfluenza, adenovirus, or other viral respiratory tract infection in a private room when possible or in a room with other patients with the same infection and no other infection (279;749;810;813;815;819).

CATEGORY IB

(2) Place a patient with suspected RSV, parainfluenza, adenovirus, or other viral respiratory tract infection in a private room.

CATEGORY II

(a) Promptly perform rapid diagnostic laboratory tests on patients who are admitted with or who have symptoms of RSV infection after admission to the health-care facility to facilitate early downgrading of infection-control precautions to the minimum required for each patient’s specific viral infection (817;819).

CATEGORY IB

(b) Promptly perform rapid diagnostic laboratory tests on patients who are admitted with or who have symptoms of parainfluenza or adenovirus infection after admission to the health-care facility to facilitate early downgrading of infection-control precautions to the minimum required for each patient’s specific viral infection and early initiation of treatment when indicated.

CATEGORY II

f. Limiting patient movement or transport in acute-care facilities (1) Limit to essential purposes only the movement or transport of patients from their rooms when they are diagnosed or suspected to be infected with RSV, parainfluenza virus, or adenovirus (279).

CATEGORY IB

(2) If transport or movement from the room is necessary (a) For a patient with diagnosed or suspected RSV or parainfluenza virus infection, ensure that precautions are maintained to minimize the risk for transmission of the virus to other patients and contamination of environmental surfaces or equipment by ensuring that the patient does not touch other persons’ hands or environmental surfaces with hands that have been contaminated with his/her respiratory secretions (279).

CATEGORY IB

(b) For a patient with diagnosed or suspected adenovirus infection, minimize patient dispersal of droplets by having the patient wear a surgical mask, and ensure that contact precautions are maintained to minimize the risk of transmission of the virus to other patients and contamination of environmental surfaces or equipment (279).

CATEGORY IB

2. Other measures in acute-care facilities

a. Staffing

(1) Restrict health-care personnel in the acute stages of an upper respiratory tract infection from caring for infants and other patients at high risk for complications from viral respiratory tract infections (e.g., children with severe underlying cardiopulmonary conditions, children receiving chemotherapy for malignancy, premature infants, and patients who are otherwise immunocompromised) (279;507;813;815;817).

CATEGORY II

(2) When feasible, perform rapid diagnostic testing on healthcare personnel with symptoms of respiratory tract infection, especially those who provide care to patients at high-risk for acquiring and/or developing severe complications from RSV, parainfluenza, or adenovirus infection, so that their work status can be determined promptly.

CATEGORY II

b. Limiting visitors

Do not allow persons who have symptoms of respiratory infection to visit pediatric, immunocompromised, or cardiac patients (279;507;810;817;819).

CATEGORY IB

c. Use of monoclonal antibody (palivizumab) for attenuation of RSV infection

Follow the recommendations of the American Academy of Pediatrics to consider monthly administration of palivizumab, an RSV monoclonal-antibody preparation, to the following infants and children aged <24 months: 1) those born prematurely at <32 weeks of gestational age and have bronchopulmonary dysplasia, and those born prematurely at <32 weeks gestation without chronic lung disease who will be aged <6 months at the beginning of the RSV season, and (2) those born at 32-35 weeks gestation, if two or more of the following risk factors are present: child-care attendance, school-aged siblings, exposure to environmental pollutants, congenital abnormalities of the airways, or severe neuromuscular disease (820;822-824).

CATEGORY II

3. Control of outbreaks in acute-care facilities

a. Perform rapid screening diagnostic tests for the particular virus(es) known or suspected to be causing the outbreak on patients who are admitted with symptoms of viral respiratory illness. Promptly cohort the patients (according to their specific infections) as soon as the results of the screening tests are available (749;764;810;813;815;817;819). In the interim, when possible, admit patients with symptoms of viral respiratory infections to private rooms

CATEGORY IB

b. Personnel cohorting

(1) During an outbreak of health-care-associated RSV infection, cohort personnel as much as practical (e.g., restrict personnel who give care to infected patients from giving care to uninfected patients) (810;813;815).

CATEGORY II

(2) No recommendation can be made for routinely cohorting personnel during an outbreak of health-care-associated adenovirus or parainfluenza virus infection.

UNRESOLVED ISSUE

c. Use of RSV immune globulin or monoclonal antibody No recommendation can be made for the use of RSV immune globulin or monoclonal antibody to control outbreaks of RSV infection in the health-care setting (820-825;827-829).

UNRESOLVED ISSUE.

PREVENTION AND CONTROL OF HEALTH-CARE-ASSOCIATED INFLUENZA

I. Staff Education

Provide health-care personnel continuing education or access to continuing education about the epidemiology, modes of transmission, diagnosis, and means of preventing the spread of influenza, in accordance with their level of responsibility in preventing healthcare-associated influenza (1029;1044-1046).

CATEGORY II

II. Surveillance

A. Establish mechanisms by which facility personnel are promptly alerted about increased influenza activity in the community. CATEGORY II

B. Establish protocols for intensifying efforts to promptly diagnose cases of facility-acquired influenza.

1. Determine the threshold incidence or prevalence of influenza or influenza-like illness in the facility at which laboratory testing of patients with influenza-like illness is to be undertaken and outbreak control measures are to be initiated (942).

CATEGORY II

2. Arrange for laboratory tests to be available to clinicians for prompt diagnosis of influenza, especially during November-April (936-939).

CATEGORY II

III. Modifying Host Risk For Infection

B. Use of Antiviral Agents (See Section V-C)

IV. PREVENTION OF PERSON-TO-PERSON TRANSMISSION

A. Droplet Precautions

1. Place a patient who is diagnosed to have influenza in a private room or in a room with other patients with confirmed influenza, unless there are medical contraindications to doing so (279).

CATEGORY IB

2 Place a patient suspected to have influenza in a private room and promptly perform rapid diagnostic laboratory tests to facilitate early downgrading of infection-control precautions to the minimum required for the patient’s infection (279).

CATEGORY II

3. Wear a surgical mask upon entering the patient’s room or when working within 3 feet of the patient (279).

CATEGORY IB

4. Limit the movement and transport of the patient from the room to those for essential purposes only. If patient movement or transport is necessary, have the patient wear a surgical mask, if possible, to minimize droplet dispersal by the patient (279).

CATEGORY II

B. Eye Protection

No recommendation can be made for wearing eye-protective device upon entering the room of a patient with confirmed or suspected influenza or when working within 3 feet of the patient.

UNRESOLVED ISSUE

C. Contact Precautions

No recommendation can be made for observance of contact precautions (in addition to droplet precautions) for patients with confirmed or suspected influenza (279;921).

UNRESOLVED ISSUE

D. Standard Precautions

1. Decontaminate hands before and after giving care to and/or touching a patient or after touching a patient’s respiratory secretions, whether or not gloves are worn: if hands are visibly dirty or contaminated with proteinaceous material or are visibly soiled with blood or body fluids, wash them with either a nonantimicrobial soap and water or an antimicrobial soap and water; and if hands are not visibly soiled, use an alcohol-based hand rub for their decontamination (278).

CATEGORY IA

2. Wear gloves if hand contact with patient’s respiratory secretions is expected(279;921).

CATEGORY II

3. Wear a gown if soiling of clothes with patient’s respiratory secretions is expected (279).

CATEGORY II

E. Air Handling

No recommendation can be made for maintaining negative air pressure in rooms of patients in whom influenza is suspected or diagnosed, or in placing together persons with influenza-like illness in a hospital area with an independent air-supply and exhaust system (909;922;924).

UNRESOLVED ISSUE

F. Personnel Restrictions

In acute-care facilities, utilize the facility’s employee health service or its equivalent to evaluate personnel with influenza-like illness and determine whether they should be removed from duties that involve direct patient contact. Use more stringent criteria for personnel who work in certain patient-care areas (e.g., ICUs, nurseries, and organ-transplant [especially HSCT] units) where patients who are most susceptible to influenza-related complications are located (920;993;1051).

CATEGORY IB

V. Control Of Influenza Outbreaks

A. Determining the Outbreak Strain

Early in the outbreak, perform rapid influenza virus testing on nasopharyngeal swab or nasal-wash specimens from patients with recent onset of symptoms suggestive of influenza. In addition, obtain viral cultures from a subset of patients to determine the infecting virus type and subtype (936-939).

CATEGORY IB

C. Antiviral Agent Administration

1. When a facility outbreak of influenza is suspected or recognized:

a. Administer amantadine, rimantadine, or oseltamivir as prophylaxis to all patients without influenza illness in the involved unit for whom the antiviral agent is not contraindicated (regardless of whether they received influenza vaccinations during the previous fall) for a minimum of 2 weeks or until approximately 1 week after the end of the outbreak. Do not delay administration of the antiviral agent(s) for prophylaxis unless the results of diagnostic tests to identify the infecting strain(s) can be obtained within 12-24 hours after specimen collection (931;941;964;1050).

CATEGORY IA

b. Administer amantadine, rimantadine, oseltamivir, or zanamivir to patients acutely ill with influenza, within 48 hours of illness onset. Choose the agent appropriate for the type of influenza virus circulating in the community (931;941;964;967;1050;1052).

CATEGORY IA

c. Offer antiviral agent(s) (amantadine, rimantadine, or oseltamivir) for prophylaxis to unvaccinated personnel for whom the antiviral agent is not contraindicated and who are in the involved unit or taking care of patients at high-risk (931;941;964;975;1050). CATEGORY IB

d. Consider prophylaxis for all healthcare personnel, regardless of their vaccination status, if the outbreak is caused by a variant of influenza that is not well matched by the vaccine (941).

CATEGORY IB

e. No recommendation can be made about the prophylactic administration of zanamivir to patients or personnel (915;941 ;974;977).

UNRESOLVED ISSUE

f. Discontinue the administration of influenza antiviral agents to patients or personnel if laboratory tests confirm or strongly suggest that influenza is not the cause of the facility outbreak (962).

CATEGORY IA

g. If the cause of the outbreak is confirmed or believed to be influenza and vaccine has been administered only recently to susceptible patients and personnel, continue prophylaxis with an antiviral agent until 2 weeks after the vaccination (941;1053). CATEGORY IB

2. To reduce the potential for transmission of drug-resistant virus, do not allow contact between persons at high risk for complications from influenza and patients or personnel who are taking an antiviral agent for treatment of confirmed or suspected influenza during and for 2 days after the latter discontinue treatment (963;984;985;987;988).

CATEGORY IB

D. Other Measures in Acute-Care Facilities:

When influenza outbreaks, especially those characterized by high attack rates and severe illness, occur in the community and/or facility: 1. Curtail or eliminate elective medical and surgical admissions (993).

CATEGORY II

2. Restrict cardiovascular and pulmonary surgery to emergency cases only (993).

CATEGORY II

3. Restrict persons with influenza or influenza-like illness from visiting patients in the health-care facility (993).

CATEGORY II

Restrict personnel with influenza or influenza-like illness from patient care (993).

Ricardo, 2013; Country: USA,

Sponsor: Americas Health Foundation (Washington, D.C., United States) and Fighting Infectious Diseases in Emerging Countries

(Miami, Florida, United States)

Scope: Influenza among the elderly in Latin America

Patients with flu-like symptoms, who are frail, immunosuppressed, have severe chronic comorbidities, are respiratory compromised, or have been admitted to a hospital should have laboratory confirmation of influenza. Patients experiencing severe influenza- like symptoms out of an epidemic season should also be given a diagnostic test. Immunofluorescense assays or PCR are the tests of choice. /Evidence for use of antiviral agents to treat influenza in the elderly, to prevent its occurrence, or to mitigate its complications, is unclear. Until this issue is resolved, physicians should use antiviral agents on a case-by-case basis.

Traditional infection control measures are essential when there is any case of influenza, especially when there is an outbreak in a long-term care facility.

Measures to avoid transmission include non-pharmacological interventions, i.e., frequent hand washing, respiratory hygiene, and cough etiquette. Traditional infection control measures should obviously be instituted when there is any case of influenza (32, 34). In long-term care facilities, outbreaks of influenza should lead to the initiation of a comprehensive approach to contain virus transmission. Increased hand hygiene practices, as well as cleaning and disinfecting surfaces with an approved antiseptic product, use of droplet precautions (surgical masks), cohorting of residents, vaccination of those previously not immunized against influenza, and possibly prophylaxis with antiviral drugs. The utilization of these interventions should not replace vaccine administration (32).

Smith, 2019; Country: USA,

Sponsor: The Society for Healthcare Epidemiology of America and the Association for Professionals in Infection Control Scope: Recommendations are developed for long-term care (LTC) infection control programs based on interpretation of currently available evidence. The recommendations cover the structure and function of the infection control program

A. Infection Control Program

1. An active, effective, facility-wide infection control program should be established in the LTCF. The purpose of the program is to help prevent the development and spread of infectious diseases (Category IC).

Comment: The elements of a program generally include the following:

a. Surveillance—Systematic data collection to identify infections in residents
b. Outbreak control—A system for detection, investigation, and control of epidemic infectious diseases in the LTCF
c. Isolation—An isolation and precautions system to reduce the risk of transmission of infectious agents
d. Policies and procedures—Relevant to infection control (see Table 2)
e. Education—Continuing education in infection prevention and control
f. Resident health program
g. Employee health program
h. Antibiotic stewardship—A system for antibiotic review and control
i. Disease reporting to public health authorities
j. Facility management, including environmental control, waste management, product evaluation and disinfection, sterilization and asepsis
k. Performance improvement/resident safety

1. Preparedness planning

2. The infection control program must be in compliance with federal, state, and local regulations (Category IC).

B. Infection Control Administrative Structure

1. Oversight of the infection control program should be defined and should include participation of the ICP, administration, nursing staff, and physician staff (Category II).

Comment: A committee, traditionally the ICC (infection control committee), may oversee the infection control program for the facility. ICC members often include the ICP; the medical director; and representatives from nursing, administration, and pharmacy. Participation of other departments, such as dietary, housekeeping, and physical therapy, should be considered on an ad hoc basis. Administrative structures other than an ICC may provide oversight to the infection control program. One example is an infection control oversight committee, a small group consisting of the LTCF administrator, the ICP, and the medical director. Alternatively, the performance improvement committee or patient safety committee and the ICC may be combined, but it is important to maintain the identity of the infection control program. The duties of the ICC should be delegated appropriately if no formal ICC exists.

2. Formal delegation of infection control oversight should be made in writing (Category II).

3. The infection control oversight committee should meet on a regular basis and have a mechanism for emergent meetings as needed (Category II).

4. This committee should maintain written minutes with identification of problems and plans for action (Category II).

5. The effectiveness of the infection control program should be evaluated by the administration on at least an annual basis (Category II).

6. Policies and procedures for investigating, controlling, and preventing infection transmission in the facility should be established (Category IC).

Comment: Other functions include (a) review of infection control data, (b) approval of policies and procedures, (c) monitoring program activities, and (d) recommending policy to the facility administration.

7. Consultation should be available as needed including with an infectious disease physician or other professional with expertise in infection control (Category II).

C. ICP

1. One person, the ICP, should be assigned the responsibility of directing infection control activities in the LTCF. The ICP should be someone familiar with LTCF resident care problems (Category IC).

2. The ICP should have a written job description of infection control duties (Category II).

3. The ICP is responsible for implementing, monitoring, and evaluating the infection control program for the LTCF (Category II).

4. The ICP should be guaranteed sufficient time and the support of the administration to effectively direct the infection control program (Category II).

5. The ICP (or another appropriate individual, such as the medical director) should have written authority to institute infection control measures in emergency situations (Category IB).

6. The ICP should have a sufficient infection control knowledge base to carry out responsibilities appropriately (Category II). Comment: A background in infectious diseases, microbiology, geriatrics, and educational methods is advisable. Management and teaching skills also are helpful. Continuing education is essential for the ICP (eg, meetings, courses, journals).

7. The ICP should know the federal, state, and local regulations dealing with infection control in the LTCF (Category II).

8. The ICP should communicate with relevant facility committees and personnel within the facility, ICPs from transferring facilities, and public health authorities to ensure appropriate isolation and collection of surveillance information (Category II).

9. No recommendation on number of ICPs per 100 LTCF beds.

D. Surveillance

1. The LTCF should have a system for ongoing collection of data on infections in the institution (Category IC).

2. A documented surveillance procedure should be used, including written definitions of infections (Category IB).

Comment: Concurrent surveillance is preferable to retrospective surveillance. The frequency of surveillance for HAIs in the LTCF should be based on factors such as acuity level of the resident population. Surveillance at least once a week generally is needed to collect timely data. Surveillance data should be collected from communication with staff; this may be during walking rounds in the LTCF. Medical progress notes in the chart, laboratory or radiology reports, nursing notes, treatment records, medication records, physical assessments, environmental observations, and follow-up information from transfers to acute care hospitals provide clues to the presence of infections.

3. The ICP should review surveillance data frequently and recommend infection control measures, as appropriate, in response to identified problems (Category IB).

Comment: Analysis of surveillance data should include at least the following elements on each infection to detect clusters and trends: resident identifier, type of infection, date of onset, location in the facility, and appropriate laboratory information.

4. Infection rates should be calculated periodically, recorded, analyzed, and reported to the administration and the infection control oversight committee (Category IB). Comment: Infection rates usually are calculated monthly, quarterly, and annually. HAI rates are calculated preferably as infections per 1000 resident- days. A standard infection report form facilitates reporting of surveillance information. Tables, graphs, and charts may be used and facilitate education of personnel.

5. Surveillance data should be used for planning infection control efforts, detecting epidemics, directing continuing education, and identifying individual resident problems for intervention (Category IB).

Comment: In addition to collection of baseline infection rates, the ICP should perform problem-focused studies. Examples of special studies are evaluation of UTIs in catheterized residents, a study of the occurrence of influenza in vaccinated versus unvaccinated residents, or the prevalence of pressure ulcers in bed-bound residents.

6. In addition to the above outcome measures, surveillance should also include analysis of process measures relevant to infection control (Category II).

Comment: Examples include monitoring hand hygiene compliance, observation of aseptic technique, and measuring HCW influenza vaccination rates.

E. Outbreak Control

1. Surveillance data should be used to detect and prevent outbreaks in the LTCF (Category IB/IC).

Comment: The occurrence of even a single verified case of a highly transmissible disease (such as infectious TB, influenza, scabies, Salmonella, and norovirus) in the LTCF should prompt notification of appropriate individuals (such as the medical director or administrator), consideration of an outbreak, and institution of control measures. After the institution of isolation precautions, assessment of exposed residents and personnel should be made in a timely fashion to detect other cases.

2. The facility should define authority for intervention during an outbreak (Category IB).

Comment: The LTCF should have a preexisting protocol for dealing with infectious disease epidemics, including the authority to relocate residents, confine residents to their rooms, restrict visitors, obtain cultures, isolate, and administer relevant prophylaxis or treatment (such as antivirals during an influenza outbreak).

3. In order to facilitate response to an outbreak, consent for appropriate diagnostic or therapeutic measures should be obtained from the resident or medical decision maker and the resident’s primary physician on admission to the facility (Category II).

4. Obtaining cultures of the environment or from asymptomatic personnel is not recommended except as targeted by an epidemiologic investigation (Category II).

5. A TB control program should focus on detection of active cases in residents and staff and isolation or transfer of residents with known or suspected pulmonary TB disease (Category IC).

Comment: TB control programs are mandated by OSHA. A case of TB in residents or staff that was or may have been acquired in the facility should lead to clinical evaluation and TB testing of residents and employees.

F. The Facility

1. Hand hygiene facilities and supplies should be available and conveniently located for residents and staff (Category IA).

2. Clean and soiled utility areas should be functionally separate and clearly designated (Category IC).

3. Appropriate ventilation and air filtration should be addressed by the LTCF (Category IC).

Comment: If the LTCF provides care for residents or accepts residents with a diagnosis of active TB, the airborne infection isolation (AII) requirement should be met. If these requirements cannot be met, a system for transfer of cases to an appropriate institution that provides AII should be part of the overall infection control plan.

4. Housekeeping in the facility should be performed on a routine and consistent basis to provide for a safe and sanitary environment (Category IC).

Comment: Cleaning schedules should be kept for all areas in the LTCF. Cleaning products should be approved and labeled appropriately; manufacturer’s (or other authoritative) recommendations for use and dilution should be followed.

5. Measures should be instituted to correct unsafe and unsanitary practices (Category II).

Comment: Environmental cleanliness may be monitored by walking rounds with a checklist for each area of the LTCF. Nursing interventions may be monitored by direct observation during such rounds.

6. Areas in the LTCF with unique infection control concerns (eg, laundry, kitchen, rehabilitation) should have the appropriate policies and procedures developed (Category II).

Comment: Laundry policies and procedures should address the following: proper bagging of linen at the site of use, transporting linen in appropriate carts, cleaning of the carts on a regular basis, separation of clean and soiled linen, washing temperatures or use of an appropriate chemical mix for low-temperature washing, covering of clean linen, protection of personnel handling soiled laundry, and hand hygiene after contact with soiled linen. Adequate supplies of clean linen should be available. Laundry regulations should be addressed if the facility does its own laundry. Dietetic service area policies and procedures should address the following: handling of uncooked foods, cooking of food, cleaning of food preparation areas, food storage, cooking and refrigeration temperatures, cleaning of ice machines, hand hygiene indications, and employee health. Food and drink should be limited to specific areas. Policies and procedures covering infection control aspects of physical therapy (including cleaning of hydrotherapy tanks) should be developed. It should include cleaning and disinfection of hydrotherapy equipment, hand hygiene indications, and cleaning of exercise equipment. If pets are allowed, the LTCF should have a policy defining access, containment, cleanliness, and vaccination of pets.

7. Policies and procedures for disposal of infectious medical waste (including waste categorization, packaging, storage, collection, transport, and disposal) should be developed in accordance with federal, state, and local regulations (Category IC).

Comment: Examples of specific issues include types of waste disposal bags, cleaning of waste transportation carts, and types of waste storage containers. Policies for sharps disposal should be developed.

G. Isolation and Precautions

1. Isolation and precautions policies and procedures should be developed, evaluated, and updated in accordance with most recent CDC/HICPAC guidance (Category IC).

2. Regular education programs should be developed to reinforce understanding and compliance (Category IC).

3. Compliance with these infection control practices (eg, hand hygiene, isolation) should be monitored (Category IC).

4. Any isolation and precautions system used should include implementation of Standard Precautions for all residents (eg, wearing of gloves, masks, eye protection, and gowns when contamination or splashing with blood or body fluids is likely) (Category IC).

5. Any isolation and precautions system should include the implementation of transmission-based precautions (Contact Precautions, Droplet Precautions, or Airborne Precautions) in accordance with current CDC/HICPAC guidance (Category IB/IC).

6. The LTCF should have a policy dealing with MDROs (such as MRSA or VRE) that is compatible with current national standards (such as the HICPAC isolation and MDRO guidelines) and appropriate to the LTCF setting (Category IB).

Comment: This policy should deal with issues such as acceptance of colonized or infected patients into the facility, inquiring about colonization of admissions with MDROs, and isolation of residents with MDROs. Denial of admission to the LTCF solely on the basis of colonization or infection with a resistant organism is not appropriate. HICPAC recommends intensification of containment measures for MDROs if ongoing transmission is occurring.

7. The individual resident’s clinical situation should be considered when deciding whether to implement or modify the use of Contact Precautions in addition to Standard Precautions if colonized or infected with a MDRO (Category IB/IC).

Comment: Routine glove use is an example of a form of modified Contact Precautions, but it has not been validated in the LTCF setting.

8. A program of safe work practices to prevent HCW exposure should be developed in accordance with CDC/HICPAC and OSHA guidance. Used needles and syringes should not be manually recapped, broken, or bent. Self-capping needles should be used. They should be disposed of, with all sharps, in a puncture-resistant, leakproof container (Category IC).

9. Gloves are indicated for contact with blood or body fluids, contaminated items, mucous membranes, or nonintact skin (Category IC).

10. Policies should be developed to deal with spills and personnel exposure to blood or body fluids. Employees should know how to respond to an exposure (eg, immediately washing the skin in the event of a blood exposure). Postexposure prophylaxis should be readily available (Category IC).

11. Residents with suspected TB should be placed in a negative-pressure room or transferred to a facility with such a room (Category IC).

H. Asepsis and Hand Hygiene

1. Routine hand hygiene should be encouraged. Hands should be washed after any patient contact but especially after contact with body fluids, after removing gloves, when soiled, and when otherwise indicated (Category IA). Unless hands are visibly soiled, use of alcohol-based hand gels is encouraged (Category IA/IC).

2. A hand hygiene policy and procedure should be developed by the LTCF in accordance with current CDC/HICPAC guidance with a program of ongoing hand hygiene education (Category IB/IC).

3. Hand hygiene compliance should be monitored (Category IC).

4. Policies and procedures for disinfection and sterilization should be developed (Category IB).

Comment: These policies and procedures should address issues such as sterile supplies, reuse of disposable items, disinfection of equipment (such as thermometers), and cleaning of noncritical items. All items, other than disposables, should be cleaned, disinfected, or sterilized, following published guidelines and manufacturers’ recommendations. The ICP should identify those resident care procedures that require aseptic technique.

I. Resident Care

1. Resident rooms should have an accessible sink, with soap, water, towels, and toilet facilities (Category II).

Comment: Provision should be made for maintaining adequate resident personal hygiene and for instructing residents in hygiene and hand hygiene as appropriate to their functional status.

2. A resident skin care program should be developed to maintain the skin as a barrier to infection (Category II).

Comment: Resident skin care should include the following: routine frequent turning for those unable to do so themselves, keeping the residents clean and dry, inspecting all residents’ skin on a routine basis, ensuring appropriate nutrition, treating pressure ulcers, and providing prompt care for any other breaks in skin integrity. Turning schedules and pressure ulcer assessment forms may be useful.

3. A program to prevent UTIs should be developed, including the following:

- Routine urinalysis or urine culture to screen for bacteriuria or pyuria is not recommended (Category IA).
- Residents with impaired bladder emptying managed with intermittent catheterization should be managed with a clean technique (Category IA).
  – Policies for catheter use should address catheter insertion, closed drainage systems, maintenance of urinary flow, and indications for changing the catheter (Category IB).
  – Irrigation of indwelling catheters with saline or antiseptics is not routinely recommended (Category IB).
- If leg bags are used, the LTCF should develop policies and procedures for aseptic connection, cleaning, and storage of leg bags (Category II).
- Adequate hydration should be maintained (Category II).

Comment: Men with incontinence should have voiding managed by a condom catheter rather than indwelling catheter, where possible. Residents with chronic indwelling catheters should have the catheter replaced and a specimen collected immediately prior to initiating antimicrobial therapy for symptomatic infection.

4. A program to minimize the risk of pneumonia in the LTCF should address the following: reducing the potential for aspiration, minimizing atelectasis, and caring for respiratory therapy equipment (Category II).

Comment: Pneumonia prevention guidelines are available, and many of the suggested measures are applicable to the LTCF.

5. Policies and procedures should be developed for prevention of infections associated with nasogastric and gastrostomy feeding tubes, including the following: preparation, storage, refrigeration, and administration of feeding solutions and care of percutaneous feeding tube skin sites (Category II).

6. Policies and procedures should be developed for prevention of IV infections, including central lines, if these devices are used (Category IB).

Comment: Policies should address indications for IV therapy, the type of dressing used to cover the IV exit site, cannula insertion, site maintenance, and changing fluids or tubing.

J. Resident Health Program

1. A resident health program should be implemented (Category II).

- There should be explicit and accessible documentation of program components in the resident record (Category II).

2. At admission, each resident should have a complete history (including important past and present infectious diseases), immunization status evaluation, and recent physical examination (Category II).

3. All newly admitted residents should receive TB screening unless a physician’s statement is obtained that the resident had a past positive TST (Category IA/IC).

Comment: A 2-step booster TST is often recommended in this setting.

4. When new or active TB is suggested by a positive skin-test result, or symptoms are consistent with active TB, a chest radiograph and medical evaluation should be obtained (Category II).

5. Follow-up TST for TB should be performed periodically or after discovery of a new case of TB in a resident or staff member (Category IB). No recommendation on frequency of routine follow-up TSTs for residents.

6. Each resident should receive current vaccinations for tetanus, diphtheria, influenza, pertussis, pneumococcal pneumonia, and any other vaccines recommended by the ACIP (Category IB/IC). ‘

7. Each resident should receive the influenza vaccine annually in the fall, unless medically contraindicated (Category IC).

Comment: Facilities should obtain resident consent at admission for yearly influenza vaccination and use standing orders for yearly influenza vaccination.

8. Policies and procedures addressing visitors should be developed to limit introduction of community infections (such as influenza) into the LTCF (Category II).

K. Employee Health Program

1. All new employees should have a baseline health assessment, including immunization status and history of relevant past or present infectious diseases (Category 1B/IC).

Comment: The past history of infectious diseases should address contagious diseases such as chickenpox, measles, hepatitis, furunculosis, and bacterial diarrhea. Screening cultures of new employees are rarely indicated.

2. All new employees should receive TST unless there is written documentation that the employee had a positive reaction to a tuberculin test. When new or active TB is suggested by a positive TST result or by symptoms, a chest radiograph and medical evaluation should be obtained (Category 1A/IC).

Comment: A 2-step booster TST technique is recommended when indicated. Only employees who have active pulmonary TB should be restricted from work.

3. Follow-up skin testing of staff who are TST negative should be performed periodically based on the facility’s annual risk assessment or after discovery of a new case of TB in a resident or staff member (Category 1A/IC).

Comment: The intradermal Mantoux method or licensed blood test should be used. The frequency of testing depends on the regional prevalence of TB; the facility’s annual risk assessment; and federal, state, or local regulations.

4. All employees should have current immunizations as recommended for HCWs by the Advisory Committee on Immunization Practices (ACIP), with documentation in the employee record (Category 1A/IC).

5. Employees with blood or body fluid contact should be offered HBV immunization within 10 working days of hire and after training has been completed (Category 1C).

Comment: Refusal of this vaccine should be documented, using the OSHA-required Declination Statement for Hepatitis B vaccine.

6. Employees should be offered the influenza vaccine annually (Category 1A/1C).

Comment: A vaccine declination statement may be signed by each employee who declines influenza vaccination.

7. Each employee should be taught basic use of personal protective equipment and hand hygiene and to consider blood and all body fluids as potentially infectious (Category 1C).

8. Employees with signs or symptoms of communicable diseases (eg, cough, rash, diarrhea) should not have contact with the residents or their food (Category 1B).

9. All employees should be educated to report any significant infectious illnesses to their supervisor and the staff member responsible for employee health (Category 1B).

Comment: Each employee record should include factors affecting immune status (such as steroid therapy, diabetes, HIV infection), history of communicable diseases, illnesses, and incidents such as exposures to contagious diseases, needlesticks, injuries, and accidents.

10. The LTCF should develop protocols for managing employee illnesses and exposures (such as bloodborne pathogens like HIV and hepatitis B and C, as well as TB, scabies, or gastroenteritis) (Category 1B/IC).

Comment: An employee absentee policy that discourages the employee from working while ill should be developed.

L. Education

1. Infection control education should be provided at the initiation of employment and regularly thereafter. Training should include all staff, especially those providing direct resident care (Category IC).

2. All programs should be documented with the date, topic, names of attendees, and evaluations (Category IC).

Comment: Program topics should be timely and relevant to infection prevention and control. Basic hygiene, hand hygiene, respiratory etiquette, transmission of infectious diseases, occupational health, prevention of TB and bloodborne pathogens, Standard and Transmission-based Precautions, infection control standards, and the susceptibility of residents to infectious diseases are topics that should be included. The ICP may recommend topics. Surveillance data are of interest to staff and may be included as appropriate. The educators should evaluate the educational program and outcomes and use that information to modify future programs.

M. Policies and Procedures

1. Infection control policies and procedures dealing with relevant aspects of infection control such as hand hygiene, disinfection, and isolation precautions should be in place and compatible with current regulations and infection control knowledge (Category IC).

2. Infection control policies and procedures should be approved, reviewed, and revised on a regular basis (Category IC).

Comment: The ICP should assist in the development and updating of infectionrelated policies and procedures.

3. Employees should be made aware of infection control policies and procedures (Category IC).

Comment: The ICP should develop a system for monitoring staff compliance with infection control policies and procedures.

N. Antibiotic Stewardship

1. Infection control programs in LTCFs should be encouraged to include a component of antimicrobial stewardship (Category IB). Comment: The LTCF should encourage judicious use of antimicrobials with guidelines based in part on local susceptibility patterns. Antibiotic utilization and appropriateness may be monitored, and these data used for interventions (eg, education, antibiotic restrictions).

2. The ICP should monitor antibiotic susceptibility results from cultures to detect clinically significant antibiotic-resistant bacteria (such as MRSA or VRE) in the institution. Changes in antibiotic-susceptibility trends should be communicated to appropriate individuals and committees (Category IB).

O. Miscellaneous Aspects

1. There should be a system for reporting notifiable diseases to proper public health officials (Category 1C).

2. The infection control program should collaborate with the performance improvement (PI) program, if a formal program exists (Category II).

Comment: Infection control is an important component of PI, and the epidemiological techniques used in infection control will assist the PI program.

3. The ICP should be involved with the review and selection of new products that have infection control implications (Category II).

4. The ICP should be involved with LTCF influenza pandemic preparedness planning (Category II).

5. Infection control activities should address relevant resident safety issues (Category II).

P. Regulations

1. The infection control program must be in compliance with federal, state, and local regulations (Category IC).

2. The infection control program should reflect national, evidence-based standards of practice for infection prevention and control (Category IC).

Uyeki, 2019; Country: USA,

Sponsor: Support for these guidelines was provided by the Infectious Diseases Society of America Scope: diagnostic testing and treatment of illness caused by infection with influenza A and B viruses circulating among humans during seasonal epidemics and does not address asymptomatic infections. The guidelines also address diagnostic testing and use of antivirals for management of institutional influenza outbreaks

What Test(s) Should Be Used to Diagnose Influenza?

Recommendations

Clinicians should not use viral culture for initial or primary diagnosis of influenza because results will not be available in a timely manner to inform clinical management (A-III), but viral culture can be considered to confirm negative test results from RIDTs and immunofluorescence assays, such as during an institutional outbreak, and to provide isolates for further characterization (C-II).

Which Patients With Suspected or Confirmed Influenza Should Be Treated With Antivirals?

Recommendations

18. Clinicians should start antiviral treatment as soon as possible for adults and children with documented or suspected influenza, irrespective of influenza vaccination history, who meet the following criteria:

- Persons of any age who are hospitalized with influenza, regardless of illness duration prior to hospitalization (A-II).

Clinicians can consider antiviral treatment for adults and children who are not at high risk of influenza complications, with documented or suspected influenza, irrespective of influenza vaccination history, who are either:

- Symptomatic healthcare providers who care for patients who are at high risk of developing complications from influenza, particularly those who are severely immunocompromised (C-III).

Patients Who Are Recommended to Receive Antiviral Treatment for Suspected or Confirmed Influenza, Which Antiviral Should Be Prescribed, at What Dosing, and for What Duration?

Recommendations

20. Clinicians should start antiviral treatment as soon as possible with a single neuraminidase inhibitor (NAI) (either oral oseltamivir, inhaled zanamivir, or intravenous peramivir) and not use a combination of NAIs (A-1).

21. Clinicians should not routinely use higher doses of US Food and Drug Administration-approved NAI drugs for the treatment of seasonal influenza (A-II).

22. Clinicians should treat uncomplicated influenza in otherwise healthy ambulatory patients for 5 days with oral oseltamivir or inhaled zanamivir, or a single dose of intravenous peramivir (A-1).

23. Clinicians can consider longer duration of antiviral treatment for patients with a documented or suspected immunocompromising condition or patients requiring hospitalization for severe lower respiratory tract disease (especially pneumonia or acute respiratory distress syndrome [ARDS]), as influenza viral replication is often protracted (C-III).

INSTITUTIONAL OUTBREAK CONTROL

When Is There Sufficient Evidence of an Influenza Outbreak in a Longterm Care Facility or Hospital to Trigger Implementation of Control Measures Among Exposed Residents or Patients and Healthcare Personnel to Prevent Additional Cases of Influenza? Recommendations

48. Active surveillance for additional cases should be implemented as soon as possible when one healthcare-associated laboratory-confirmed influenza case is identified in a hospital or one case of laboratory-confirmed influenza is identified in a long-term care facility (A-III).

49. Outbreak control measures should be implemented as soon as possible, including antiviral chemoprophylaxis of residents/ patients, and active surveillance for new cases, when 2 cases of healthcare-associated laboratory-confirmed influenza are identified within 72 hours of each other in residents or patients of the same ward or unit (A-III).

50. Implementation of outbreak control measures can be considered as soon as possible if one or more residents or patients has suspected healthcare-associated influenza and results of influenza molecular testing are not available on the day of specimen collection (B-III).

Which Residents/Patients Should Be Considered to Have Influenza and Be Treated With Antivirals During an Influenza Outbreak in a Long-term Care Facility or Hospital?

Recommendations

51. When an influenza outbreak has been identified in a longterm care facility or hospital, influenza testing should be done for any resident/patient with one or more acute respiratory symptoms, with or without fever, or any of the following without respiratory symptoms: temperature elevation or reduction, or behavioral change (A-III).

52. Empiric antiviral treatment should be administered as soon as possible to any resident or patient with suspected influenza during an influenza outbreak without waiting for the results of influenza diagnostic testing (A-III).

To Control an Influenza Outbreak in a Long-term Care Facility or Hospital, Should Antiviral Chemoprophylaxis Be Administered to Exposed Residents/Patients?

Recommendation

53. Antiviral chemoprophylaxis should be administered as soon as possible to all exposed residents or patients who do not have suspected or laboratory-confirmed influenza regardless of influenza vaccination history, in addition to implementation of all other recommended influenza outbreak control measures, when an influenza outbreak has been identified in a long-term care facility or hospital (A-III).

During an Influenza Outbreak at a Long-term Care Facility, Should Antiviral Chemoprophylaxis Be Administered to Residents Only on Affected Units or to All Residents in the Facility?

Recommendation

54. Antiviral chemoprophylaxis should be administered to residents on outbreak-affected units, in addition to implementing active daily surveillance for new influenza cases throughout the facility (A-II).

Which Healthcare Personnel Should Receive Antiviral Chemoprophylaxis During an Institutional Outbreak?

Recommendations

55. Clinicians can consider antiviral chemoprophylaxis for unvaccinated staff, including those for whom chemoprophylaxis may be indicated based upon underlying conditions of the staff or their household members (see recommendations 40-44) for the duration of the outbreak (C-III).

56. Clinicians can consider antiviral chemoprophylaxis for staff who receive inactivated influenza vaccine during an institutional influenza outbreak for 14 days postvaccination (C-III).

57. Clinicians can consider antiviral chemoprophylaxis for staff regardless of influenza vaccination status to reduce the risk of short staffing in facilities and wards where clinical staff are limited and to reduce staff reluctance to care for patients with suspected influenza (C-III).

How Long Should Antiviral Chemoprophylaxis Be Given to Residents During an Influenza Outbreak in a Long-term Care Facility? Recommendation

58. Clinicians should administer antiviral chemoprophylaxis for 14 days and continue for at least 7 days after the onset of symptoms in the last case identified during an institutional influenza outbreak (A-III).

Victoria State Government, 2018; Country: Australia, Sponsor: Victoria State Government Scope: guidelines apply to all residential care facilities (RCFs) in Victoria. This refers to any public or private aged care, disability services or other congruent accommodation setting in Victoria where residents are provided with personal care or health care by facility staff.

Prevention and preparedness

Facilities must ensure they are prepared for respiratory outbreaks prior to the start of the influenza season (March / April).

#### 3.7 Antiviral medication

There is good evidence that the timely administration of antiviral medication, as treatment and/or prophylaxis (prevention), can reduce the duration of an influenza outbreak and the number of residents who are affected. As per the Communicable Disease Network Australia (CDNA) Guidelines for the Prevention, Control and Public Health Management of Influenza Outbreaks in Residential Care Facilities in Australia (2017) <http://www.health.gov.au/internet/main/publishing.nsf/Content/cdna-flu-guidelines.htm>, the department recommends the use of antiviral medications in symptomatic residents during laboratory confirmed influenza outbreaks, and the consideration of prophylaxis for asymptomatic residents (regardless of vaccination status) and unvaccinated staff3.

The department recommends that facilities have clear internal policies in place regarding the use of antiviral medication as prophylaxis and/or treatment of influenza during outbreaks. It is strongly recommended that clinical staff liaise with residents’ families and GPs / clinicians regarding this aspect of the outbreak management plan prior to an outbreak occurring. Where possible, a decision should be made prior to the influenza season regarding the intention to use antiviral medication in an outbreak setting for each resident which should be documented in the resident’s file and/or the facility’s outbreak management plan.

Identifying respiratory outbreaks

#### 4.1 Identifying an outbreak

This section includes how to identify and whom to report outbreaks to

#### 5 Outbreak management

##### 5.1 Establishing an outbreak management team

It is the facility’s responsibility to self-manage the outbreak. If possible, an internal outbreak management team (OMT) should be established to direct, monitor and oversee the outbreak, confirm roles and responsibilities and liaise with CDPC.

##### 5.2 Implementing infection prevention and control measures

###### 5.2.1 Standard precautions

Standard precautions refer to the work practices required to achieve a basic level of infection prevention and control. They apply to all residents, regardless of suspected or confirmed infection status, in all health care and long-term residential care settings.

Standard precautions include (but are not limited to):

- hand hygiene
- use of personal protective equipment
- respiratory hygiene/cough etiquette
- routine environment cleaning
- cleaning of shared equipment.

Consistent application of the precautions noted above will reduce the risk of transmission of respiratory infections.

###### 5.2.1.1 Hand hygiene

Hand hygiene is one of the most effective infection control measures for preventing the spread of infectious pathogens. Emphasis should be placed on the importance of hand hygiene for all staff, residents and visitors.

###### 5.2.1.2 Use of Personal Protective Equipment (PPE)

Staff must wear appropriate PPE when it is anticipated that there may be contact with a resident’s blood or body fluids, mucous membranes, non-intact skin or other potentially infectious material or equipment. Gloves, mask and eye protection should be used whenever any aerosol producing procedures are undertaken such as when obtaining a respiratory specimen from a resident.

PPE should be removed in a manner that prevents contamination of the HCW’s clothing, hands and the environment. PPE should be immediately discarded into appropriate waste bins.

Always perform hand hygiene before putting on PPE and immediately after removal of PPE.

Staff should be trained and deemed proficient in donning and doffing PPE before an outbreak occurs. The following resources are highly recommended showing the correct technique for donning and doffing PPE. Tasmanian Infection Prevention and Control Unit’s videos <http://www.dhhs.tas.gov.au/publichealth/tasmanian_infection_prevention_and_control

###### 5.2.1.2.1 Eye protection

When there is the risk of splash or splattering of blood or body fluids, secretions or excretions, eye protection should be worn. Personal eye glasses are not adequate eye protection. Eye protection includes safety glasses, goggles or face shields.

###### 5.2.1.2.2 Single use face masks

Single use face masks (also called surgical face masks) should be worn when exposure to respiratory droplets is likely

###### 5.2.1.3 Respiratory hygiene / cough etiquette

Respiratory hygiene and cough etiquette should be applied as a standard infection control precaution at all times. Covering coughs and sneezes prevents respiratory secretions from dispersing into the air and contaminating environmental surfaces.

Respiratory hygiene / cough etiquette includes the following actions.

- Cover the nose/mouth with disposable single-use tissues when coughing, sneezing, wiping and blowing noses.
- Use tissues to contain respiratory secretions.
- Dispose of tissues immediately into the nearest waste container.
- If no tissues are available, cough or sneeze into the inner elbow rather than the hand.
- Attend to hand hygiene immediately after contact with respiratory secretions and contaminated objects/materials.
- Keep contaminated hands away from mucous membranes of the eyes and nose.

Residents may require assistance with attending to their respiratory hygiene. Hand hygiene facilities should be provided for residents, particularly when they are immobile.

###### 5.2.1.4 Cleaning shared equipment

Ensure that shared equipment (for example, lifting machine, commode, thermometer) is not used for another resident until it has been appropriately cleaned (and disinfected or reprocessed if required).

Items such as slings should be dedicated to one resident’s use and must be laundered before use on another resident.

Anything labelled as single-use must be discarded after use and not reprocessed or used on another resident.

###### 5.2.1.5 Routine environmental cleaning

Regular scheduled cleaning of the environment is essential to prevent the spread of infectious organisms. The influenza virus can survive on surfaces for up to 24 hours, thereby leading to contamination of hands. Frequently touched surfaces, such as door handles, over bed tables, mobility aids and light switches require more frequent cleaning compared to other surfaces. Respiratory Illness in Residential and Aged Care Facilities Guidelines 2018 Page 19

###### 5.2.2 Transmission-based precautions (droplet precautions)

Transmission based precautions are infection control precautions used in addition to standard precautions to prevent the spread of certain infectious pathogens. Droplet precautions are the additional infection control precautions required when caring for residents suspected or confirmed as having influenza or other respiratory illness.

Droplet precautions (in addition to the standard precautions listed above) include the procedures described below.

###### 5.2.2.1 Resident placement

It is preferable that all residents with a respiratory illness be cared for in a single room with their own ensuite facilities. The resident should be restricted to their room for five days after symptom onset.

If single rooms are not available, use the following principles to guide resident placement.

- Give highest priority to single room placement to residents with excessive cough and sputum production.
- Place residents together in the same room (cohort) with similar signs and symptoms or infected with the same pathogen (if known) and assessed as being suitable roommates.
- When a single room is not available, and cohorting of ill residents is not possible, a resident with a respiratory illness may be cared for in a room with a roommate(s) who does not have a respiratory illness. This is the least favourable option. In this case
  – residents’ beds should be separated by at least one metre
  – the curtain should be kept drawn between residents’ beds
  – the roommate must be vaccinated against influenza with the current season’s vaccine prior to that year’s season starting or at least two weeks prior to being in the same room as the ill resident.
- In shared rooms (both cohorted with like illness and residents with and without illness), staff must ensure they change their PPE and perform hand hygiene when moving between residents.

###### 5.2.2.2 Resident movement

- Where possible, residents in droplet precautions should be restricted to their room for five days after symptom onset. Residents may attend necessary medical or procedural appointments.
- Residents with a respiratory illness who must leave their room should wear a mask if tolerated.
- Consider placing all residents at least one metre apart in communal areas such as dining, lounge and other areas, where possible, during a respiratory outbreak.
- Communicate the respiratory illness status of the resident to other healthcare facilities and services before transfer so that appropriate infection prevention and control precautions can be implemented at the accepting facility.

###### 5.2.2.3 Signage

A droplet precaution sign must be placed outside symptomatic residents’ rooms to alert staff and visitors to the requirement for transmission-based precautions.

###### 5.2.2.4 Personal protective equipment

All staff and visitors entering the room of a person with a respiratory illness should wear a single use face mask for close contact, generally within one metre. For further information regarding mask use see section 5.2.1.2.2 Single use face masks.

Gowns, gloves and protective eyewear need only be worn as per standard precautions, that is, if contact or splash with blood or body fluids is anticipated.

###### 5.2.2.5 Equipment and instruments/devices

Use disposable equipment where possible (for example, blood pressure cuffs) or dedicate use of non-disposable equipment to any residents with a respiratory illness. If equipment must be shared (for example, lifting machine) for multiple residents, ensure the equipment has been cleaned and disinfected before use on another resident.

Consider cohorting equipment to use for patients in droplet precautions, or to a wing or unit under isolation. If equipment is cohorted, it must still be cleaned and disinfected between each patient use.

###### 5.2.2.6 Linen and laundry items

Handle, transport, and process used linen or items requiring laundering (for example, clothing) in a manner that avoids contamination of air, surfaces and persons. If linen or resident clothing is laundered onsite compliance with the Australian Standard AS/NZS 4146:2000 Laundry Practice is required.

No additional precautions are required for the management of linen for residents with influenza or other respiratory infections. Linen and clothing items from residents with a respiratory illness do not need to be segregated or laundered separately.

###### 5.2.2.7 Eating utensils

Crockery and cutlery should be washed in a dishwasher or if not available by hand using hot water and detergent, rinsed in hot water and dried. The use of disposable cutlery or separation of cutlery and crockery during an outbreak is not required.

###### 5.2.2.8 Waste management

Ensure waste is appropriately segregated into the different waste streams, for example, general, recyclable, or clinical and related waste. Storage and handling of all waste must meet the Environment Protection Authority (EPA) Victoria legislative requirements. For more information refer to EPA Victoria’s Clinical and Related Waste - Operational Guidance <www.epa.vic.gov.au/business-and-industry/guidelines/waste-guidance/clinical-waste-guidance>.

All gloves, masks, protective eyewear or gowns used whilst caring for a resident with influenza should be disposed of into clinical waste (yellow bin or bag).

###### 5.2.2.9 Ceasing droplet precautions

Generally, people with influenza are considered infectious for five days. Droplet precautions should therefore continue for at least five days after symptom onset.

###### 5.2.3 Environmental cleaning and disinfection

Influenza viruses are able to survive on environmental surfaces, particularly hard surfaces, for periods of one to two days. Infection can occur through contact with contaminated surfaces then infecting oneself or others by touching eyes, nose or mouth with contaminated hands. As such, regular cleaning and disinfection of surfaces should be undertaken during a suspected or confirmed outbreak to minimise the spread of influenza and other respiratory viruses.

###### 5.2.5 Visitors and communal activities

During an outbreak, where possible, minimise the movement of visitors into and within the facility. Facilities should consider implementing the following strategies.

- Suspend all group activities, particularly those that involve visitors (for example, musicians).
- Postpone visits from non-essential external providers (for example, hairdressers).
- Inform regular visitors and families of residents of the respiratory outbreak and request that they only undertake essential visits.
- Ask visitors who do visit an ill resident to
  – visit only the ill resident
  – wear PPE as directed by staff
  – enter and leave the facility directly without spending time in communal areas
  – perform hand hygiene before entering and after leaving the resident’s room.

###### 5.2.6 Staff

For suspected or confirmed cases of respiratory illness in residents caused by influenza it is preferable that only staff who have been vaccinated for influenza provide care for these residents.

During a confirmed influenza outbreak unvaccinated staff are recommended to attend work only if they are asymptomatic and wear a single use face mask, or are asymptomatic and taking appropriate antiviral prophylaxis.

All staff members (vaccinated and unvaccinated) should self-monitor for signs and symptoms of respiratory illness and self-exclude if unwell. Staff with respiratory illnesses should be excluded from work for the period during which they are infectious (generally five days after the onset of the acute illness or until symptoms have ceased).

During an outbreak, wherever possible, HCWs should not move between wings or units of the facility to provide care for other residents. This is particularly important if not all wings/units are affected by the outbreak. It is preferable to cohort staff to areas (in isolation or not in isolation) for the duration of the outbreak.

Unvaccinated staff who have been working in the outbreak affected area should not be moved as they may be incubating infection. They should be offered immediate influenza vaccination. Please note that vaccination may not prevent illness if already incubating. A protective immune response takes approximately two weeks to develop.

###### 5.2.7.1 New admissions

An ongoing outbreak does not mean the facility has to go into complete “lock down”. It is preferable that admission of new residents to an affected unit during an outbreak does not take place. Where new admissions are unavoidable, new residents and their families must be informed about the current outbreak and adequate outbreak control measures must be in place for these new residents. Families may wish to make alternative arrangements until the outbreak is over.

###### 5.2.7.2 Re-admission of cases

The re-admission of residents who met the case definition and have been hospitalised for their illness is permitted, provided appropriate accommodation and infection prevention and control requirements can be met.

###### 5.2.7.3 Re-admission of non-cases

The re-admission of residents that have not been on the respiratory outbreak case lists (that is, are not a known case) should be avoided during the outbreak period if possible. If non-cases are re-admitted, the resident and their family must be informed about the current outbreak and adequate outbreak control measures must be in place. Families may wish to make alternative arrangements until the outbreak is over.

###### 5.2.7.4 Transfers

If transfer to hospital is required, the ambulance service and receiving hospital must be notified of the outbreak/suspected outbreak verbally and through using a resident transfer advice form. An example form <https://www2.health.vic.gov.au/public-health/infectious-diseases/infection-control-guidelines/respiratory-illness-management-in-aged-care-facilities> is provided on the department’s webpage.

###### 5.2.7.5 Non-infected residents

In some circumstances, it may be feasible to transfer residents who are not symptomatic, to other settings (for example, family care) for the duration of the outbreak. The family or receiving facility should be made aware that the resident may have been exposed and is at risk of developing disease.

Woodhead, 2011; Country: N/A (international guideline),

Sponsor: European Respiratory Society (ERS), in collaboration with The European Society for Clinical Microbiology and Infectious

Diseases (ESCMID)

Scope: common management questions occurring in routine clinical practice in the management of adult patients with LRTI Who should be admitted to hospital?

Recommendation: The decision to hospitalize remains a clinical decision. However, this decision should be validated against an objective tool of risk assessment In patients residing in nursing homes, a predefined clinical pathway can help to reduce hospitalization by about 50%, with comparable clinical outcomes [260].

Are amplification tests useful for the diagnosis of LRTI?

Recommendation: Application of molecular tests for the detection of influenza and RSV should be considered during the winter season and for the detection of atypical pathogens, provided the tests are validated and the results can be obtained sufficiently rapidly to be therapeutically relevant [A3]. Previously unknown viruses have been discovered: several coronaviruses, human metapneumovirus and bocavirus. They are detected in CAP by NAATs. Reports on infection by a mixture of several viruses or infection by a mixture of viruses and bacteria exist. Systematic comprehensive studies are awaited to define the clinical importance of these viral and mixed infections.

What classification should be used for treatment?

Recommendation: Antimicrobial treatment has to be empirical and should follow an approach according to the individual risk of mortality. The assessment of severity according to mild, moderate and severe pneumonia implies a decision about the most appropriate treatment setting (ambulatory, hospital ward or ICU) [A4]. Antimicrobial treatment should be initiated as soon as possible [A3].

Which additional therapies are recommended?

Recommendation: All patients should be subject to early mobilization [A3]. Low molecular heparin should be given in patients with acute respiratory failure [A3]. The use of non-invasive ventilation is not yet standard care but can be considered, particularly in patients with COPD [B3] and ARDS [A3]. The treatment of severe sepsis and septic shock is confined to supportive measures [A3]. Steroids are not recommended in the treatment of pneumonia [A3]. Early mobilization has been shown to be associated with better outcome. For the purpose of the study, early mobilization was defined as movement out of bed with change from horizontal to upright position for at least 20 min during the first 24 h of hospitalization, with progressive movement each subsequent day during hospitalization [430].

Do antiviral substances prevent influenza virus infection?

Recommendation: Prevention of influenza by antiviral substances should only be considered in special situations (for example in outbreaks in closed communities during influenza seasons) [A1]. In the case of seasonal influenza outbreaks or a pandemic situation, the national recommendations should be followed.

PHAC, 2010; Country: Canada Sponsor: Public Health Agency of Canada Scope: infection prevention and control guidance to healthcare workers in the management of patients with suspected or confirmed seasonal influenza

Source Control, achieved through administrative and engineering measures, is the most effective way to prevent the transmission of infectious agents in all healthcare settings. The most effective way to prevent and control seasonal influenza is through immunization of both healthcare workers and patients.

In addition to Routine Practices, patients with suspected or confirmed seasonal influenza in acute care and LTC settings should be placed on Droplet and Contact Precautions.

1. Source Control

a. Respiratory hygiene Respiratory hygiene should be encouraged for patients and accompanying individuals who have signs and symptoms of an influenza-like-illness (ILI) 5,(see 3.b.), beginning at the point of initial encounter in any healthcare setting (e.g., inpatient, triage, reception and waiting areas in emergency departments, outpatient clinics, etc.). Respiratory hygiene includes coughing into one’s sleeve and using tissues and, masks when coughing, sneezing, or for controlling nasal secretions. Healthcare facilities should provide tissues and masks for respiratory hygiene, as well as instructions on how and where to dispose of them, and on the importance of performing hand hygiene (see 6. Hand Hygiene) after handling this material. Patients should be taught to perform hand hygiene and how to perform respiratory hygiene. Patients with suspected/confirmed influenza should wear a mask (if tolerated) when HCWs, other staff, or visitors are present. Patients may remove their masks once accommodated in their rooms (see 7. Accommodation).
b. Spatial separation There should be at least a 2-metre separation between patients who have signs and symptoms of an ILI 5 and those who do not.

2. Screening a. Patients symptomatic with an ILI5 should be assessed in a timely manner and potential causes of acute respiratory infection other than influenza should be considered (e.g., tuberculosis, respiratory syncytial virus, etc.). b. The following criteria for ILI5 can be used to determine the need for applying the infection prevention and control measures found in this guidance.

- Acute onset of respiratory illness with fever and cough, and with one or more of the following: sore throat, arthralgia, myalgia, or prostration which is likely due to influenza.
- In patients under 5 years or 65 years and older, fever may not be prominent.

3. Surveillance Prospective surveillance for ILI 5 should be established (see 2.b.).

4. Laboratory Testing/Reporting

a. Provisions for influenza diagnostic testing should be in place before the onset of influenza season each year.
b. A protocol for testing patients with ILI5 to confirm the presence of influenza by direct testing or viral culture should be established.
c. Diagnostic samples (nasopharyngeal swab or aspiration) should be taken from symptomatic patients as soon as influenza has been recognized in the community.
d. Rapid diagnostic tests with a high degree of sensitivity should be available to facilitate the earlier detection of influenza and thereby enable appropriate medical management, earlier initiation or discontinuation of additional precautions, and reduction in transmission. Negative results of tests with low sensitivity should not be used to make decisions regarding stopping precautions.
e. Prompt notification to infection prevention and control professionals and attending physicians as well as regional, provincial/territorial public health authorities as required should be ensured.

6. Hand Hygiene HCWs should perform hand hygiene frequently (as recommended in the PHAC “Hand Hygiene Practices in Health Care” guideline 9 and the healthcare organization’s policy) preferably using an alcohol-based hand rub (60-90%) or soap and water if hands are visibly soiled. Other types of waterless products may contain either no alcohol or alcohol in concentrations of less than 60%. There is no efficacy data on these products, and they should not be used for hand hygiene in healthcare settings.

Accommodation

Patients suspected or confirmed to have influenza should be cared for in single rooms if possible. Perform a risk assessment to determine patient placement and/or suitability for cohorting when single rooms are limited or if in a LTC setting. Patients who are known to have influenza should be cohorted with suitable roommates. If cohorting is not possible and a cubicle or designated bedspace is used in a shared room, privacy curtains should be drawn between beds. Infection control signage should be placed at the room entrance indicating Droplet and Contact Precautions required upon entry to the room.

8. Patient Flow/Activities Patients with ILI 5 should be restricted to their room until symptoms have resolved. Participation in group activities should be restricted while the patient is symptomatic. Movement/transport of patients with suspected or confirmed influenza should be restricted to essential diagnostic and therapeutic tests only. Transfer within facilities should be avoided unless medically indicated. If transport is necessary, transport services and personnel in the receiving area should be advised of the required precautions for the patient being transported. Patients with influenza who leave their room for medical reasons (i.e., essential diagnostic and therapeutic tests) should wear a mask and adhere to respiratory hygiene.

9. Personal Protective Equipment (PPE)

a) PPE in addition to Routine Practices

- Facial protection (masks and eye protection, face shields, mask with visor attachment) should be worn when within 2 metres of a patient with suspected or confirmed influenza.
- In acute care settings, gloves should be worn when entering the room of a patient with suspected or confirmed influenza.
- In LTC settings, gloves should be worn for direct personal care with the patient or if direct contact with frequently touched environmental surfaces is anticipated.
- A long-sleeved gown should be worn if it is anticipated that clothing or forearms will be in direct contact with the patient or with environmental surfaces or objects in the patient care environment.
- Remove all PPE just before leaving the patient’s room and discard in a hands-free waste or linen receptacle within the room.
- Hand hygiene should be performed after removing gloves and gown, before removing facial protection, and after leaving the room. Note: In a shared room/cohort setting, facial protection may be worn for the care of successive patients.

b) Aerosol generating medical procedures (AGMPs) • AGMPs should only be performed on patients with confirmed or suspected influenza if medically necessary.

- The number of HCWs present should be limited to only those essential for patient care and support.
- Wherever possible, the HCWs present during the procedure should be limited to those who have received influenza vaccine.
- Droplet and Contact Precautions in addition to Routine Practices as outlined in 9.

a) should be used when performing AGMPs on patients with seasonal influenza.

10. Patient Care Equipment All patient care equipment (e.g., thermometers, blood pressure cuff, pulse oximeter, etc.) should be dedicated to the use of one patient. All patient care equipment should be cleaned and disinfected as per Routine Practices before reuse with another patient or a single use device should be used and discarded in a waste receptacle after use. Toys, electronic games or personal effects should not be shared by patients.

11. Environmental Cleaning Hospital-grade cleaning and disinfecting agents are sufficient for environmental cleaning in the context of influenza. All horizontal and frequently touched surfaces should be cleaned at least twice daily and when soiled. The healthcare organization’s terminal cleaning protocol for cleaning of the patient’s room following discharge, transfer or discontinuation of Droplet and Contact Precautions should be followed.

12. Laundry/Waste Management Routine Practices are sufficient.

13. Discontinuing Droplet and Contact Precautions Droplet and Contact Precautions for seasonal influenza should be discontinued when the patient is no longer symptomatic or according to the organization’s policy. Medical procedures that can generate aerosols as a result of artificial manipulation of a person’s airway. Procedures include: intubation and related procedures (e.g. manual ventilation, open endotracheal suctioning), cardiopulmonary resuscitation, sputum induction, nebulized therapy, surgery and autopsy, noninvasive positive pressure ventilation (CPAP, BiPap)

14. Management of Visitors Individuals who have ILI5 symptoms should be restricted from visiting except for compassionate reasons. Visitors with ILI5 should be instructed in respiratory hygiene, wear a mask, perform hand hygiene and visit the patient directly and exit directly after the visit. Visitors for patients with influenza should be limited to persons who have been identified by the patient or next-of-kin as necessary for the patient’s emotional well-being and care. They should be instructed in hand hygiene and asked to limit their movement within the facility.

15. Outbreak Management

a. All healthcare organizations should have a written plan for managing an influenza outbreak in their facilities.
b. Consider deferring admissions to an affected unit/ward.
c. Consider cohorting patients with confirmed influenza in a single geographic area of the unit/ward or hospital.
d. Consider restricting social activities or education programs to the affected ward/unit. e. Consider influenza vaccination for all unvaccinated patients unless contraindications exist.
e. Consider chemoprophylaxis to all patients, whether vaccinated or not, who are in the outbreak area, are not already ill with influenza, and have no contraindications.
f. Consider chemoprophylaxis for all unvaccinated HCWs, unless contraindications exist.
g. Consider excluding unvaccinated HCWs who are not taking chemoprophylaxis from direct patient care.
h. Consider influenza vaccination (unless contraindications exist) for unvaccinated HCWs who receive chemoprophylaxis; these individuals may continue to work without restrictions.

MOHLTC, 2018; Country: Canada

Sponsor: Ontario Ministry of Health and Long-term Care

Scope: assist long-term care facilities and public health units with prevention, detection and management of respiratory infection outbreaks

#### 3.1 Outbreak Management

Step 1 - Assess the Suspect or Confirmed Outbreak, Establish a Preliminary Outbreak Case Definition, and Begin a Line-list.

Step 2 - Implement General IPAC Measures.

Step 3 - Declare an Outbreak in your LTCH and Notify the Local Medical Officer of Health or Designate at your PHU of the Suspect or Confirmed Outbreak.

Step 4 - Notify Appropriate Individuals Associated with the LTCH of the Outbreak and Establish OMT Membership.

Step 5 - Call an initial OMT Meeting.

Step 6 - Communicate the Results of Laboratory Tests Step 7 - Monitor the Outbreak on an Ongoing Basis.

Step 8 - Declare the Outbreak Over.

#### 4.1 General IPAC Measures

##### 4.1.2 Hand Hygiene

Four Moments for HH:

1. Before initial resident/resident environment contact.
2. Before invasive/aseptic procedures.
3. After body fluid exposure risk and contact with blood, body fluids, secretions and excretions.
4. After resident/ resident environment contact.

##### 4.1.3 Personal Protective Equipment

PPE is used alone or in combination to prevent exposure, by placing a barrier between the infectious source and one’s own mucous membranes, airways, skin and clothing.

Control of Respiratory Infection Outbreaks in Long-Term Care Homes

The selection of PPE is based on the nature of the interaction with the resident and/or the likely mode(s) of transmission of infectious agents. Selection of the appropriate PPE is based on the risk assessment (e.g., interaction, status of resident) that dictates what is worn to break the chain of transmission.

The selection of controls should be based on a hierarchy of controls approach. PPE ranks lowest in the hierarchy of controls. PPE is a last line of defense for workers against hazards related to infectious agents that cannot otherwise be eliminated or controlled.

LTCHs must ensure that staff has sufficient supplies of, and quick, easy access to, the PPE required. As well, LTCHs should provide education in the proper selection and use of PPE to all health care providers and other staff who have the potential to be exposed to blood and body fluids.

LTCHs should also consider the requirement of the HCRF Regulation 67/93 section 10 in this regard, which states that:

1. A worker who is required by his or her employer or by this Regulation to wear or use any protective clothing, equipment or device shall be instructed and trained in its care, use and limitations before wearing or using it for the first time and at regular intervals thereafter and the worker shall participate in such instruction and training.
2. Personal protective equipment that is to be provided, worn or used shall,
  a. be properly used and maintained;
  b. be a proper fit;
  c. be inspected for damage or deterioration; and
  d. be stored in a convenient, clean and sanitary location when not in use.

Gloves

When a resident is placed on Contact or Droplet-Contact precautions, gloves are used when direct care will be provided. In addition, gloves must be worn when it is anticipated that the hands will be in contact with mucous membranes, non-intact skin, tissue, blood, body fluids, secretions, excretions, or equipment and environmental surfaces contaminated with the above.

Indiscriminate or improper glove use has been linked to transmission of microorganisms. Gloves are task specific and single use for the task

Masks

A mask is used by a health care provider (in addition to eye protection) to protect the mucous membranes of the nose and mouth when it is anticipated that procedure or care activity is likely to generate splashes or sprays of blood, body fluids, secretions or excretions, or when within two metres of a coughing resident

Eye Protection

Eye protection is used by health care providers (in addition to a mask) to protect the mucous membranes of the eyes when it is anticipated that a procedure or care activity is likely to generate splashes or sprays of blood, body fluids, secretions or excretions, or within two meters of a coughing resident

Gowns

A gown is recommended when it is anticipated that a procedure or care activity is likely to generate splashes or sprays of blood, body fluids, secretions, or excretions, or a resident is on contact or droplet/contact precautions and direct care will be provided. Long-sleeved gowns protect the forearms and clothing of the health care provider from splashing and soiling with body substances during procedures and resident care activities which are likely to generate splashes or sprays of blood, body fluids, secretions, or excretions.

#### 4.2 IPAC Measures for Residents

Generally, LTCHs and PHUs discuss with OMT members the respiratory infection outbreak IPAC measures and decide jointly on appropriate measures to implement. The extent to which outbreak IPAC measures can be implemented and what is considered reasonable throughout the course of each outbreak will vary. Examples of reasonable and appropriate measures during the course of an outbreak include:

- limiting visiting hours;
- limiting the number of residents with whom the visitor has contact;
- requiring anyone (including visitors, other residents, etc.) providing direct care toa resident on Additional Precautions to wear the necessary PPE;
- requiring visitors to not visit when they have an ARI;
- requiring visitors to wear the required PPE when visiting a resident on Additional Precautions and to carefully remove and discard PPE and perform HH upon leaving the room;
- requiring residents to wear a gown, masks or other PPE, if they have an ARI and are leaving their room or are within 2 metres of others who are not wearing PPE;
- posting signs at entrances of LTCHs and/or affected unit/area, discouraging visitors during the outbreak period; and
- notifying persons of the outbreak.

However, under outbreak conditions that present a greater risk to the resident population of the LTCH, more restrictive control measures may be required and occasionally there may be a conflict relating to PHU recommended outbreak IPAC measures. If the PHU assesses the risk of not complying with outbreak control recommendations to be high - that is, the probability of adverse health events to other residents, such as disease transmission, is high - the PHU may have to consider a written order from the Medical Officer of Health or designate under the HPPA to the licensee of the LTCH to ensure compliance with outbreak IPAC measures. Under these circumstances, it is reasonable and necessary for the Medical Officer of Health or designate, if he or she is satisfied that the statutory test in the HPPA is met, to issue a section 22 order under the HPPA to the licensee of the LTCH, to:

- stop new admissions to the LTCH
- restrict resident movement to and from the home, or
- restrict visitors from the home

These, and other possible actions which the Medical Officer of Health or designate may list in the order, are fairly significant measures, and presumably lesser measures would be discussed with the LTCH and implemented before admissions would be stopped or visitors restricted completely from the LTCH.

##### 4.2.1 Admissions and Returns from Absences

When experiencing an outbreak, LTCHs, with support from the PHU as necessary, need to be vigilant regarding admissions and residents returning after absences to ensure that due diligence has been exercised in order to protect these residents and/or the residents with whom they may come into contact. Admissions and return from absence decisions may be made in consultation with the PHU if necessary. See below for factors to consider.

A comprehensive approach to new admissions or return of residents from hospitals back to the LTCH requires consideration of a number of factors, including the risk of remaining in hospital (and being exposed to other nosocomial risks) and the risk to individual residents, as well as patients in the larger context of health care. Restricting admissions to a LTCH in outbreak may create a backlog in emergency departments or acute care, with a risk to patients in that system. On the other hand, admission of an unexposed resident into a LTCH that is experiencing an outbreak may put them at risk and may lengthen the duration of the outbreak, with an impact to the larger resident population. Therefore a measured and considered approach is required, in consultation with the Medical Officer of Health or designate if necessary.

##### 4.2.3 Restriction of Symptomatic Residents to Their Room

Cases (ill residents) should be encouraged to stay in their room, and should be on Droplet and Contact Precautions until 5 days after the onset of acute illness or until symptoms have resolved (whichever is shorter). For some pathogens, the period of communicability may be longer than 5 days, but for practical reasons, this could still be applied to outbreaks caused by respiratory viruses other than influenza or in the case of outbreaks when the pathogen is not known. There may be some respiratory outbreaks for which longer isolation periods are required. This would occur in consultation with the OMT. For influenza, the recommendation to isolate residents to their rooms is due to the increased viral shedding that occurs when patients are symptomatic; restriction of ill residents to their room is recommended as long as it does not cause the resident undue stress or agitation. If, however, restriction causes undue stress or agitation, alternative IPAC measures can be considered, including the use of a surgical mask and compliance with HH, at the discretion of the LTCH in consultation with the PHU.

Residents with an ARI who are not in single room accommodation can be managed in their bed space using Droplet and Contact Precautions with privacy curtains drawn, where these accommodations are available.25 However, residents may leave their room if they are able to comply with HH requirements and with the use of a surgical mask. This strategy may not work with all populations and its application is left to the discretion of the LTCH in consultation with the PHU.

##### 4.2.4 Restriction of Residents to Their Unit

In some LTCHs, if ill residents cannot be contained in one geographical area of the LTCH, then the outbreak must be considered facility-wide. If cases are confined to one unit, all residents and staff from that unit should avoid contact with residents and staff in the remainder of the LTCH.25 Additional recreational activities/resources should be made available. In some LTCHs, the outbreak may be confined to one or more units without declaring a facility-wide outbreak, however, this will depend on a number of factors (e.g. design of the facility, causative organism, speed of spread, proximity of cases, staffing resources).

##### 4.2.5 Communal Meetings and Other Activities

As much as possible, encourage symptomatic residents to stay within their own units within the home. It is always important to balance the rights of residents with the need to manage the outbreak. Previously scheduled events, (e.g. holiday events) may have to be rescheduled. The OMT should discuss restriction of activities, revisiting the issue as the outbreak progresses.

If possible, consideration should be given to planning events in such a way as to permit well residents to participate, according to geographical areas.

The following should be considered for implementation during an outbreak, based on OMT and local decision making:

- Reschedule communal meetings on the affected unit/floor. However, other meetings or activities may proceed in non-affected areas;
- Discontinue group outings from the affected unit/floor;
- The OMT should discuss restricting meetings or activities in the entire LTCH if the outbreak spreads to two or more units/floors;
- Restrict visits by outside groups, such as entertainers, volunteer organizations and community groups, as deemed necessary by the OMT;
- Conduct on-site programs such as physiotherapy and foot care for residents in their rooms, if possible. Proper precautions should be taken for ill residents; and
- Ensure there is no interaction between the affected floor/unit and participants in on-site child-care or other day programs.

##### 4.2.6 Medical Appointments

At the discretion of and after consultation with the treating physician, non-urgent appointments may be rescheduled, with the consent of the resident/SDM.

##### 4.2.7 Transfer to Hospital

It is the policy of the MOHLTC that no inter-facility resident transfer takes place without the sending facility first obtaining a Medical Transfer (MT) authorization number from the Provincial Transfer Authorization Centre (PTAC). This policy also applies to residents being transported from a healthcare facility to and from a private doctors’ or dentists’ office for treatment. Of course, the policy does not apply to life threatening emergencies which DO NOT require authorization to transfer.

Before sending an ill resident to acute care, the facility should notify the receiving healthcare facility and the PTAC that the home is experiencing an outbreak.

If approved, an authorization number will be issued immediately and either sent on-line or by fax depending on the method used to obtain the MT authorization number from PTAC.

The goal is to protect sending and receiving facilities, paramedic and private transfer companies and the public by ensuring appropriate personal protective measures are taken thus containing any risk of spreading.

The hospital ICP must be provided with the details of the outbreak to ensure control measures are in place when the resident arrives at the hospital. The hospital ICP shall be informed of whether or not the resident to be transferred has been identified as a case. The outbreak transfer letter attached in Appendix 9 can be used to provide the required information. In addition, notifying the receiving hospital whether the transferred resident was or was not on the line list, allows the hospital to start discharge planning.

##### 4.2.8 Transfer to another Long-Term Care Home

Symptomatic resident transfers (from anywhere in the home) to another LTCH are not recommended during an outbreak. The OMT should discuss exceptions to this recommendation and make decisions on a case by case basis. All transfers must go through PTAC. Refer to the PTAC approval process above.

#### 4.3 IPAC Measures for Staff, Students and Volunteers

##### 4.3.1 Reporting of Respiratory Illness

Staff, volunteers or contracted service workers with an ARI should not enter the LTCH; they should report any respiratory illness to their supervisor who shall report to the employee health nurse or the ICP.

##### 4.3.2 Exclusion of Staff, Students, and Volunteers, with an Acute Respiratory Infection

Staff, students, or volunteers with any respiratory infection symptoms should not return to work/placement for 5 days from the onset of symptoms of a respiratory illness or until symptoms have resolved whichever is shorter. This includes staff, students and volunteers on antiviral medication.

##### 4.3.3 Working at Other Facilities

During non-influenza outbreaks, well staff, students, and volunteers may be able to work/provide services at other facilities based on OMT and local decision making.

During an influenza outbreak, staff protected by either immunization or antiviral have no restrictions on their ability to work at other facilities. However, unimmunized staff not receiving prophylactic therapy must wait one incubation period (3 days) from the last day that they worked at the outbreak facility/unit prior to working in a non-outbreak facility, to ensure they are not incubating influenza. However, unimmunized staff on antiviral prophylactic therapy that wishes to work at another facility may do so, assuming the following considerations:

- They do not have a fever and/or other symptoms of ARI.
- This does not conflict with the policies of the receiving facility, as these would supersede the general direction provided here.
- This does not conflict with direction provided by the Medical Officer of Health or designate based on information available to them about the epidemiology of the outbreak or other local considerations.

Staff, students, and volunteers experiencing respiratory symptoms or fever should not work/provide services in any health care setting.5

##### 4.3.4 Cohort Staffing

During non-influenza outbreaks, consider cohorting staff and staff assignment between the affected residents/outbreak units and unaffected units. Alternatively, consider the possibility of keeping staff members working on only one unit if possible. Attempts should be made to minimize movement of staff, students, or volunteers between floors/resident home areas, especially if some units are unaffected.25

##### 4.3.5 Exclusion of Unimmunized Staff

During a laboratory-confirmed influenza outbreak, immunized staff who has been immunized at least two weeks prior to outbreak declaration may work in the outbreak home. Unimmunized staff may resume work at the affected home as soon as they are taking antiviral prophylaxis. If issues arise regarding compliance with work exclusions, options should be reviewed with the OMT.5

#### 4.4 IPAC Measures for Visitors and Private Pay Caregivers

##### 4.4.1 Notification of Visitors and Private Pay Caregivers

During a vaccine-preventable disease outbreak, such as influenza or pneumonia, all visitors/private pay caregivers should be encouraged to be immunized if applicable (e.g. pneumonia vaccine only if >65 years of age), as some residents may not be immunized, or may have waning protection from immunization.10 LTCHs shall post outbreak notification signs at all entrances to the home indicating the institution is in outbreak.

Visitors/private pay caregivers shall be advised of the potential risk of acquiring illness within the home, and the re-introduction of illness into the home, and of the visiting restrictions as indicated below. LTCHs may choose to notify families of the outbreak and the impact on visitation.

##### 4.4.2 Visitor and Private Pay Caregiver Restrictions

Ill visitors/private pay caregivers shall not be permitted in the home, unless under extenuating circumstances. Under these circumstances, they should wear the appropriate PPE, perform HH upon arrival, as needed during their stay and when leaving both the room of the resident and the LTCH, and finally, they should restrict their visit to the resident.

Well visitors/private pay caregivers who choose to visit during an outbreak and who are not going to be providing direct care to an ill resident should be asked to:

- Consider wearing PPE if they plan on visiting again in the next week; will also help them avoid getting sick themselves;
- Perform hand hygiene when entering the LTCH, before entering and upon leaving the resident’s room;
- Visit residents only in their rooms and avoid communal areas;
- If possible, visit only one resident and leave the LTCH immediately after the visit; if multiple residents are in the home but in different locations, it is recommended that the healthy resident(s) (non-outbreak case) be visited first; and
- Not mingle with other residents.

In addition to these recommendations, well visitors/private pay caregivers who choose to visit during an outbreak and are going to be providing direct care to an ill resident should be asked to wear the appropriate PPE. Moreover, the following recommendations apply regarding visitor restrictions:

- Notices shall be placed on the door of the rooms of ill residents or in other visible locations advising all visitors to check at the nursing station before entering the room. Visitors are to be advised of the above visitor restrictions.
- Ill residents should be visited in their room only.

Complete closure of a LTCH to visitation is not permitted unless there is an order issued by the Medical Officer of Health or designate as it may cause residents and visitors emotional hardship. Under exceptional circumstances, the Medical Officer of Health or designate may assess the risk to be significant such that it requires complete closure to visitors. In these circumstances, an order from the MOH to the licensee is required to ensure compliance. It is important to note however that even under these circumstances, that there are exceptional personal circumstances under which barring visitors is neither ethical nor permitted. In these situations, the LTCH must ensure full compliance with infection control requirements. Furthermore, decisions to restrict visitors with or without an order of the MOH may be challenged and therefore need to be carefully considered and implemented. Visitation restrictions should be discussed by the OMT.

#### 4.5 Environmental Cleaning and Disinfection

The principles of Routine Practices are based on the premise that all residents, their secretions, excretions and body fluids and their environment might potentially be contaminated with harmful microorganisms. By following simple preventive practices at all times regardless of whether or not an illness is ‘known’, staff will be protecting residents/visitors and themselves from an unknown, undiagnosed infectious risk.

During an outbreak there may be a requirement for additional or enhanced environmental cleaning of a health care setting, in order to contain the spread of the microorganism causing the outbreak. Policies and procedures regarding staffing in Environmental Services (ES) departments should allow for surge capacity (e.g., additional staff, supervision, supplies, equipment) during outbreaks as determined by the outbreak management committee. The outbreak management committee should include, among other departments, representation from ES who will lead the coordination of the department’s activities.

PHUs and LTCHs should become familiar with PIDAC’s Best Practices for Environmental Cleaning for Prevention and Control of Infections in All Health Care Settings, April 2018. In addition, procedures for assigning responsibility and accountability of routine cleaning of all environmental surfaces and non-critical resident care items should be established.

#### 5 Use of Influenza Antivirals for Residents and Staff

Antiviral prophylaxis should not replace annual influenza immunization.

Antiviral medication is recommended for the management of institutional outbreaks of influenza A and/or influenza B. Antivirals play a key role in outbreak management and control. Research has shown that antiviral drugs are effective for both the prevention (prophylaxis) and early treatment of influenza infection. The use of antiviral medication, in conjunction with other outbreak IPAC measures, can quickly bring influenza outbreaks in health care facilities under control.28 Influenza vaccination provides incomplete protection to the elderly and the immunocompromised.9 Antiviral medications offer protection that is additive to that of annual influenza immunization in these populations. Antiviral medications are also effective in the prevention of influenza in unvaccinated healthy adults.

Two antiviral drugs are currently used in Canada for the treatment and prophylaxis of influenza. Oseltamivir (oral) and zanamavir (inhaled) are both neuraminidase inhibitors that help to prevent further replication of the influenza virus.28 Zanamivir is generally not recommended for residents, as they may have difficulty using the inhaler.

Decisions regarding influenza antiviral prophylaxis or treatment should be made based on current data of circulating influenza strains, including antiviral resistance. Testing of influenza isolates for antiviral resistance is performed as part of routine laboratory surveillance at the National Microbiology Laboratory (NML), and is reported by PHO’s Ontario Respiratory Pathogen Bulletin, the Laboratory-Based Respiratory Pathogen Surveillance Report and the national FluWatch report.

Health care providers are advised to refer to updates on influenza activity and antiviral resistance patterns in ongoing surveillance reports mentioned above.

If antiviral drug resistance is detected or suspected in an institutional outbreak (e.g. if an outbreak appears poorly controlled despite proper antiviral use), or resistance has been reported in local community, local and provincial health authorities should be contacted for up-to-date advice on antiviral use.

##### 5.1 Antiviral Medication Recommendations

PHUs should be aware that clinical recommendations for the use of antiviral medications may change from season to season, as additional evidence becomes available.

To ensure guidance related to the use of antivirals reflects the most up-to-date seasonal antiviral medication use recommendations, PHUs should be aware of the Association of Medical Microbiology and Infectious Disease (AMMI) Canada’s current guidelines for the use of antiviral drugs for influenza.

##### 5.2 Antivirals as Part of an Influenza Outbreak Preparedness Plan

An outbreak plan should include measures that will expedite the administration of antiviral medication for staff and residents. A plan is required to begin antivirals quickly not only because treatment is most effective when started within 48 hours of symptom onset, but also because prophylaxis should begin as soon as possible to stop the progression of the outbreak. This plan should include measures to ensure rapid access to antiviral medications from local pharmacies.

The following recommendations, excerpted from the Influenza Prevention and Surveillance Protocol for Long-Term Care Homes, September 2014, should be addressed in appropriate LTCH policies in preparation for an outbreak to ensure that there are no delays in providing influenza immunization and/or antiviral medication.

- Consent for antiviral medication use during the entire influenza season should be obtained from residents or their substitute decision-makers in advance of each influenza season. For LTCHs, this consent may be obtained at the same time that consent is obtained for influenza immunization.
- In LTCHs, advance medical orders for influenza antiviral medication for residents should be obtained from medical staff at the beginning of each influenza season, or a plan should be in place to obtain physician’s orders quickly in the event of an outbreak. Advance medical orders can substantially expedite administration of antiviral medications.
- Staff who are unimmunized against influenza for any reason should be informed that in the event of an outbreak, they may be given the option of taking antiviral medication for the duration of the outbreak in order to continue their duties, or if unable or refuse to take antiviral medication for the duration of the outbreak, they may be excluded from working in the LTCH depending on the policy of theLTCH.5
- To facilitate antiviral treatment during outbreaks, staff who are unable to receive the influenza vaccine should be assessed for eligibility for antiviral drugs such asoseltamivir or zanamivir prior to the influenza season. A record of this information should be kept on-hand at the LTCH to expedite timely implementation of antiviral prophylaxis. In addition, staff who are not immunized, who conduct activities in the LTCH, and who are assessed as being able to take antiviral medication, may wish to obtain and keep prescriptions on hand to assist with timely commencement of antivirals, in the event of an influenza outbreak.5
- During the influenza season, LTCH administration must keep a current list of staff working in the LTCH who are not immunized, in order to promptly implement control measures such as antiviral prophylaxis and cohorting staff.5 Other control measures such as non-patient care work arrangements or staff exclusions may also be considered.

##### 5.3 Antiviral Medication for Prevention (Prophylaxis)

During a public health-confirmed influenza outbreak, antiviral medication for prevention shall be offered to all residents/patients in the outbreak-affected area who are not already ill with influenza, whether previously vaccinated or not, until the outbreak is declared over.

In addition, all unvaccinated asymptomatic staff who work in the area of the LTCH where the influenza outbreak is occurring should be advised to take prophylactic antiviral medication until the outbreak is declared over.

During a confirmed influenza outbreak, when the circulating strain is not well-matched by the vaccine, antiviral prophylaxis may be offered to all staff, regardless of vaccination status, as determined by the OMT or in consultation with the Medical Officer of Health or designate until the outbreak is declared over. PHUs may consult with PHO regarding scientific and technical support regarding evidence of a mismatch.

Antiviral prophylaxis should be initiated as soon as an influenza outbreak is declared. In almost all situations, it is prudent to wait for laboratory confirmation of influenza before initiating prophylaxis and treatment. Once the specimen reaches the appropriate laboratory, rapid test results are usually available within one business day. In some circumstances, the Medical Officer of Health may provide recommendations for prophylaxis prior to laboratory confirmation.

Institutions should consult with PHU representatives on the outbreak management team when starting antiviral prophylaxis and treatment.

Recommendations regarding influenza antiviral prophylaxis:

- It is reasonable to allow unvaccinated staff to work with residents or patients on an outbreak unit as soon as they start antiviral prophylaxis. Unless there is a contraindication, consenting staff should also immediately be offered immunization against influenza with the current seasonal influenza vaccine.
- In healthy adults, it takes two weeks to develop antibodies to the influenza virus after receiving the influenza vaccine. Staff who have been vaccinated for less than two weeks at the time the influenza outbreak is declared should take antiviral prophylaxis for two weeks after vaccination or until the outbreak is declared over (whichever comes first). Note: Antiviral medications do not interfere with the immune response to vaccine.
- Staff should be alerted to the symptoms and signs of influenza, particularly within the first 4 days after starting antiviral prophylaxis. Staff illness should immediately be reported to the supervisor, ICP and/or occupational health. Staff reporting signs and symptoms of influenza should be excluded from working in any healthcare setting if symptoms develop. This information should be shared with the local public health representative.
- Prophylaxis may be discontinued once the influenza outbreak is declared over.
- Prophylaxis may also be given during influenza season in institutional settings not experiencing an influenza outbreak to unvaccinated individuals at high-risk of influenza-related complications, at the discretion of the treating physician.
- If a person taking a neuraminidase inhibitor (i.e. oseltamivir or zanamivir) for prophylaxis of influenza develops symptoms of an influenza-like illness, the neuraminidase inhibitor can be continued, however, the neuraminidase inhibitor should be increased to the recommended treatment dose. Consideration should be given to obtaining a nasopharyngeal specimen if the individual has been on antiviral prophylaxis for more than four days to determine the presence of a resistant strain or another respiratory virus.

###### 5.3.1 Antiviral prophylaxis of the outbreak unit(s) only versus the whole facility

The advantages and disadvantages of providing antiviral prophylaxis to the outbreak unit(s) only or to the whole facility may be evaluated based on the specific characteristics of the outbreak and the design of the facility.

Advantages of a whole-facility approach include: preventing the spread of the outbreak to other units, preventing the introduction from another outside source when influenza is circulating in the community, not needing to be as vigilant to detect the spread on another unit as would be needed if surveillance is being used as a trigger for prophylaxis, and preventing the need to manage an outbreak unit by unit as new units are added.

Disadvantages to the whole-facility approach include: logistics of using antiviral medication for a large number of residents, the theoretical potential for resistance if the drug is widely used for prevention, antiviral availability, and the potential for side effects occurring in a larger number of residents.

##### 5.4 Antiviral Medication for Treatment

Treatment decisions for the residents/patients are the responsibility of the attending physicians. However, treatment decisions for health care staff that work in the LTCH rest with their health care provider and as such, obtaining prescriptions for antiviral treatment is the responsibility of the staff.5 See section 5.6 for payment information.

Treatment should be started within 48 hours (or less) of onset of symptoms for maximum effectiveness. This may also decrease complications of influenza infection.

Recommendations regarding antiviral treatment:

- Antiviral treatment should be started for ill residents/patients (who meet the outbreak case definition), as soon as possible and preferably within 48 hours of symptom onset. As much as possible symptomatic residents should be encouraged to remain in their rooms for the duration of antiviral treatment.
- Once an outbreak has been laboratory-confirmed as influenza, additional laboratory confirmation of each new case is not required in order to initiate antiviral treatment in individuals who meet the outbreak case definition.

MOH, 2020; Country: Canada

Sponsor: Ministry of Health

Scope: screening, infection control for COVID-19 in long-term care

Long-term care homes (LTCHs) should take enhanced precautions to prevent visitors who screen positive for COVID-19 from visiting the home.

- Staff, other caregivers, and volunteers with symptoms of an acute respiratory infection must not come to work and must report their symptoms to the LTCH.
- All testing for COVID-19 will take place in hospitals or as otherwise arranged in consultation with local public health, unless the LTCH has the capacity to safely conduct a clinical examination and collect specimens for a patient at risk of having COVID-19 as detailed in the Novel Coronavirus (2019-nCoV) Guidance for Primary Care Providers in a Community Setting.
- LTCH staff should follow routine precautions as well as contact and droplet precautions when providing health care services to any person under investigation for COVID-19 . LTCHs that can safely conduct a clinical examination and collect specimens should also follow airborne precautions as detailed in the Novel Coronavirus (COVID-19) Guidance for Primary Care Providers in a Community Setting.

LTCHs are being requested to conduct passive screening of visitors, staff, and volunteers, and active screening of residents. The LTCH should also ensure that an employee health policy is in place to send employees home if symptoms begin to develop at work.

1. Passive screening for staff, volunteers and visitors

- Signage should be posted on entry to the building and at reception areas for anyone entering the LTCH (e.g., visitors, staff, volunteers) to self-identify if they have relevant symptoms and travel history/exposure, including:
  - Fever, and/or
  - Acute respiratory illness, and
  - Travel history to mainland, China in the last 14 days since onset of illness OR have had contact with a person who has the above travel history and is ill.
- If experiencing respiratory symptoms, visitors must not visit the LTCH until symptoms completely resolve. For more information, please see: Control of Respiratory Infection Outbreaks in Long-Term Care Homes, 2018.
- As part of routine measures for the respiratory season, existing signage should be visible that reminds residents and visitors to perform hand hygiene, sneeze/cough into their elbow, wear a procedure mask if needed, put used tissues in a waste receptacle and to wash hands immediately after using tissues.
- LTCHs must instruct all staff and volunteers to self-screen at home. Staff, other caregivers, and volunteers with symptoms of an acute respiratory infection must not come to work and must report their symptoms to the LTCH. All staff should be aware of early signs and symptoms of acute respiratory infection. For more information, please see: Control of Respiratory Infection Outbreaks in Long-Term Care Homes, 2018.

2. What to do if a staff member, volunteer or visitor screens positive?

- LTCHs should provide further guidance (e.g., over the phone or at the reception desk) to volunteers and visitors who are experiencing symptoms of COVID-19 and have a recent travel history (within 14 days) to mainland China (e.g., they should call Telehealth Ontario or their local public health unit).
- More information on what to do if a staff member screens positive can be found below under occupational health & safety and infection, prevention &control advice.

3. Active screening for resident admissions and re-admissions or returning residents Sample Screening

Is the resident presenting with:

1. Fever, and/or new onset of cough or difficulty breathing,

AND any of the following:

2. Travel to mainland China in the 14 days before the onset of illness OR Close contact with a confirmed or probable case of COVID- 19ORClose contact with a person with acute respiratory illness who has been to mainland China in the 14 days before their symptom onset.

- LTCHs should conduct screening (when possible, over-the-phone screening) for new admissions, re-admissions or returning residents.
- LTCHs must consult with the local public health unit if exposure to, or transmission of, COVID-19 has been confirmed in order to determine any additional public health actions, including implementing active staff screening.

4. What to do if a resident screens positive?

- If a resident of a LTCH answers yes to both questions (1) and (2), and they are onsite, they should:
- Be instructed to wear a procedure mask (if tolerated) and placed in a single room to wait for further assessment.

If necessary, a health care provider in the LTCH can conduct a clinical history and visual assessment using routine, droplet, and contact precautions and maintaining a 2 metre distance from the resident, but should not conduct a physical examination unless the

LTCH has the capacity to safely conduct a clinical examination and collect specimens for a patient at risk of having COVID-19 as detailed in the Novel Coronavirus (COVID-19 Guidance for Primary Care Providers in a Community Setting.

The LTCH staff do not need to use enhanced airborne precautions (i.e., are not required to have an airborne infection isolation room with negative pressure) unless they determine they are able to safely conduct a clinical examination and collect specimens as detailed in the Novel Coronavirus (COVID-19) Guidance for Primary Care Providers in a Community Setting.

- The LTCH should contact their local public health unit to report the suspect case and discuss the most appropriate setting for the resident to be clinically assessed and tested, if warranted. All referrals to hospital should be made to a triage nurse.
- If a resident is referred to a hospital, the LTCH should coordinate with the hospital, local public health unit, paramedic services and the resident to make safe arrangements for travel to the hospital that maintains isolation of the patient.

Patient transfer services should not be used to transfer a resident from the LTCH who screens positive to the questions above.

Occupational health & safety and infection prevention & control advice

Within LTCHs, the ministry recommends the use of routine practices and additional precautions (contact and droplet) in the event the resident receives a health assessment by a health care provider. These precautions include:

- hand hygiene
- environmental cleaning
- wearing appropriate personal protective equipment (PPE) for droplet precautions (e.g. procedural masks) if a resident screens positive
- proper training in donning (putting on) and doffing (taking off) of PPE in order to prevent cross-contamination and the potential spread of infection

Testing

- At this time, health care providers in LTCHs are not expected to conduct testing for COVID-19 unless the LTCH has determined they have the capacity to safely conduct a clinical examination and collect specimens as detailed in the Novel Coronavirus (COVID-19) Guidance for Primary Care Providers in a Community Setting.
- All health care providers have a duty to report a patient who has or may have COVID-19 to the local public health unit.
- The LTCH should contact their local public health unit to report the suspect case and discuss the most appropriate setting for the resident to be clinically assessed and tested.

Reporting

The LTCH should use routine reporting procedures to contact their local public health unit. COVID-19 is a designated disease of public health significance (O. Reg. 135/18) and thus reportable under the Health Protection and Promotion Act.

Burdsall, 2017; Country: United States

Sponsor: Agency for Healthcare Research and Quality

Scope: Infection Prevention and Control in Long-Term Care

Infection Prevention and Control in Long-Term Care

KEY MESSAGES

- Break the chain of infection.
- Detect, diagnose, and treat infections quickly and effectively.
- Do not rush to use antibiotics.
- Carefully follow facility policies and procedures to prevent infections.

Health care workers can reduce the risk of infection by—

- Cleaning hands with an alcohol-based hand rub or soap and water, also known as practicing hand hygiene
- Wearing gloves and other personal protective equipment per facility policy
- Keeping the environment clean and properly disinfecting surfaces and medical equipment
- Handling waste safely
- Avoiding touching your face
- Covering mouths and noses when sneezing or coughing
- Not coming to work when sick
- Staying up to date on all recommended vaccinations
- Practicing standard precautions for all residents

STANDARD PRECAUTIONS: INFECTION PREVENTION BASICS

KEY MESSAGES

- Practice standard precautions for the care of all residents all the time.4
- Observe the standard precautions of not touching blood, body fluids, mucous membranes, cuts, wounds, or rashes with bare hands—and not letting these touch your skin, face, or clothes.
- Use personal protective equipment (PPE) when contact is possible with blood, body fluids, mucous membranes, or nonintact skin.
- Practice hand hygiene.
- Use safety needles and sharps.
- Practice respiratory etiquette by covering coughs in sleeves and wearing masks when recovering from coughs or colds.

Specific Standard Precaution: HAND HYGIENE

KEY MESSAGES

- Practice hand hygiene when moving among residents and from soiled to clean spaces.
- Wash hands with soap and water for at least 20 seconds, or use an alcohol-based hand rub or alcohol hand wipe, covering all surfaces of the hands.

Specific Standard Precaution: ENVIRONMENTAL CLEANING AND DISINFECTION

KEY MESSAGES

- All staff have a role in keeping the facility and equipment clean and disinfected.
- The best cleaning products—
- Clean and disinfect at the same time
- Are safe on surfaces
- Hospital-approved cleaners and disinfectants are adequate for most situations in LTC facilities.
- All staff at the LTC facility should receive training before using any cleaning products.

Specific Standard Precaution: PERSONAL PROTECTIVE EQUIPMENT

KEY MESSAGES

- The PPE must prevent contact between skin, mucous membranes, and clothes from blood, body fluids, and other potentially infectious materials.
- All staff should wear PPE any time there is a chance of contact with blood and body fluids.
- All staff, family members, and visitors should wear PPE when isolation precautions are in place.

Specific Standard Precaution: RESIDENT PLACEMENT

KEY MESSAGES

- Good communication among all staff is critical so that everyone knows how to best care for residents’ individual needs, including their placement.
- Private rooms are the best way to prevent the spread of germs and infections.
- When private rooms are not available—
- Residents infected or colonized with the same germ can be placed together.
- If that is not possible, place infected residents with low-risk residents.

Specific Standard Precaution: RESPIRATORY HYGIENE AND ETIQUETTE

KEY MESSAGES

- Everyone needs to watch for and report respiratory illness.
- Vaccinations are an important tool for preventing respiratory illnesses such as influenza and pneumococcal pneumonia.
- Staff should stay home if they are sick.
- Staff should go home if they develop respiratory symptoms while working.
- A virus can cause a cold for a staff member but may develop into a serious illness for an older adult.
- Visitors, families, and staff can be a source of respiratory illness outbreaks.
- Cover coughs, and wear a mask if recovering from an illness.
- Educate residents and visitors to cover their mouths and noses with a tissue (or if not available, upper sleeve) when coughing or sneezing.
- Residents should stay in their rooms if they develop a new cough with fever or other symptoms of a respiratory infection.

Specific Standard Precaution: SAFE INJECTION PRACTICES

KEY MESSAGES

- All sharps (needles/lancets/syringes) used for injections or obtaining blood must be designed to reduce the risk of needle sticks. This can be done with needle guards and automatic retraction devices, or with safety engineered sharps, such as nonremovable needles and syringes with fixed doses.
- While residents may have their own equipment for their own personal use, health care workers cannot use residents’ equipment.

Specific Standard Precaution: SOILED LINEN

KEY MESSAGES

- Treat all soiled linen as potentially infectious.
- Linen must be processed in a way that not only kills germs but also does not spread germs from dirty to clean linens.
- Heat and chemical disinfection are two methods used to kill germs in laundry.
- Use one color of bag for soiled linen and a different-colored bag for trash.
- Don’t put linens in red biohazard bags unless they are soaked with blood and are being discarded in a biohazard bin.

III. TRANSMISSION-BASED PRECAUTIONS AND OUTBREAK MANAGEMENT Transmission-Based Precautions (General)

KEY MESSAGES

There are three mechanisms of infection transmission:

- Contact
- Indirect
- Direct ▪ Droplet
- Large respiratory particles that travel short distances (up to 6 feet)
- Airborne
- Small respiratory particles that stay suspended in the air
- Contact, droplet, and airborne precautions are used in addition to standard precautions.
- Adopt a person-centered approach to practicing transmission-based precautions: “Only when necessary for only as long as necessary.”

Specific Transmission-Based Precaution: CONTACT PRECAUTIONS

KEY MESSAGES

- Use contact precautions to prevent the spread of germs by direct or indirect contact with residents or their environments.
- Contact precautions are special safeguards that must be put in place when dealing with residents who are infected with certain germs.
- Adopt a person-centered approach: “Only when necessary for only as long as necessary.”

Specific Transmission-Based Precaution: DROPLET PRECAUTIONS KEY MESSAGES

- Droplet precautions are used against influenza (also known as the flu).
- Wear a mask in addition to using standard precautions.
- Residents on droplet precautions should stay in their rooms.
- If a resident on droplet precautions has to leave his or her room, the resident must wear a mask.
- Consider using both droplet and contact precautions if the respiratory virus causing the illness is unknown or if the resident has nausea, vomiting, or diarrhea.

Specific Transmission-Based Precaution: AIRBORNE PRECAUTIONS

KEY MESSAGES

- Airborne precautions are used for diseases such as tuberculosis and chicken pox.
- Airborne precautions are rarely used in LTC facilities.
- An LTC facility must have negative pressure rooms and a respiratory fit-test program in order to safely maintain airborne precautions

Outbreak Management

KEY MESSAGES

- Quick identification of clusters of infections is critical.
- Keep the environment and equipment clean and disinfected.
- Make sure there are disinfectants at the point of care.

IV. ENGAGING EVERYONE IN INFECTION PREVENTION AND CONTROL

KEY MESSAGES

- Good infection prevention practices, including hand hygiene, respiratory hygiene, safe injection practices, and appropriate antibiotic use, contribute to a safe facility for residents and a safe workplace for staff.
- Everybody who works in the facility needs to work together to practice infection prevention to prevent harm and increase resident safety.
- Residents and family members play a role in increasing resident safety by practicing infection prevention themselves, and in supporting the health care team in prevention practices.

